# Optimal Duration of Antibiotic Treatment for Group A Streptococcal Pharyngitis in Children: A Systematic Review and Dose-Response Meta-Analysis

**DOI:** 10.64898/2026.06.25.26356472

**Authors:** João Pedro Lima, Mahya Dorri, Michael Ling, Brandon Lee, Sarah Kirsh, Saveen Dhanoya, Alicia Walch, Tanvir Jassal, Mahsa Raji Lahiji, Ashlyn Chou, Haotao Li, Andy Cui, Oswin Chang, Madison Bigler, Jeffrey M. Pernica, Mohamed Eltorki, Deborah Yamamura, Bradley J. Langford, Mark Loeb, Alena Tse-Chang, Nicole Le Saux, Dena Zeraatkar

## Abstract

**Importance:** Group A streptococcal (GAS) pharyngitis drives substantial morbidity and antibiotic prescribing in children. Ten days of antibiotics remains standard, although shorter courses could reduce treatment burdens, cumulative antibiotic exposure, and selective pressure for resistance. Whether shorter courses achieve comparable outcomes remains uncertain.

**Objective:** To assess the effects of oral antibiotic treatment duration on clinical outcomes in children and adolescents with GAS pharyngitis.

**Data Source:** MEDLINE, Embase, CENTRAL, Web of Science, and CINAHL from inception to July 2025, without language restrictions.

**Study Selection:** **R**andomized trials of participants ≤18 years with suspected or confirmed GAS pharyngitis comparing different durations of oral antibiotics, or oral antibiotics against placebo or no treatment.

**Data Extraction and Synthesis:** Pairs of reviewers selected studies and extracted data. Random-effects dose-response meta-analysis summarized the effect of duration on outcomes. The GRADE approach guided judgments about the certainty of evidence.

**Main Outcomes and Measures:** Clinical cure, relapse, adverse events, mortality, serious adverse events, acute rheumatic fever, purulent complications, and poststreptococcal glomerulonephritis.

**Results:** Forty-five trials enrolling 22,636 participants met eligibility criteria. Moderate certainty evidence suggests little to no clinically important difference between 5 vs 10 days in clinical cure (14 fewer cures per 1000 [95% CI 27 fewer to 2 fewer]), relapse (5 fewer per 1000 [11 fewer to 3 more]), or adverse events (10 more per 1000 [6 fewer to 30 more]). Sensitivity analyses excluding macrolides produced similar results. Low certainty evidence also suggests little to no clinically important difference in clinical cure, relapse, or adverse events between 3 vs 10 days of treatment, but nearly all evidence came from trials of azithromycin, and these findings do not establish the effects of 3-day regimens of penicillins or other antibiotics. Serious adverse events were rare: no deaths and 4 cases of acute rheumatic fever among 776 and 8,818 participants, respectively, making clinically important differences across treatment durations unlikely.

**Conclusions and Relevance:** For children at low risk of acute rheumatic fever, 5 days of oral antibiotics likely provides similar benefits and harms as 10 days while reducing treatment burden and antibiotic exposure. The effects of 3-day regimens remains uncertain.

**Funding:** Canadian Institutes of Health Research (CIHR). Grant No. 540708

**Key Points:** *Question:* Do shorter courses of oral antibiotics provide clinical outcomes comparable with 10 days of treatment in children and adolescents with group A streptococcal pharyngitis?

*Findings:* In this systematic review and dose-response meta-analysis of 45 randomized trials enrolling 22 636 participants, 5 days compared with 10 days of antibiotic treatment produced no clinically important differences in clinical cure, relapse, or adverse events. Evidence for 3-day regimens was less certain and came almost entirely from azithromycin trials.

*Meaning:* Five-day antibiotic courses likely provide an effective alternative to 10-day courses for children in populations resembling those enrolled in trials, but evidence remains insufficient for 3-day regimens of penicillins or other antibiotics. These findings may not extrapolate to populations with a higher risk of complications.

## Introduction

Pharyngitis is a major driver of outpatient medical visits and antibiotic prescribing in children and adolescents.^1–6^ Viruses cause most pediatric cases of pharyngitis, while Group A β-hemolytic *Streptococcus* (GAS) accounts for only 5% to 36% of presentations.^7,8^ Clinicians prescribe antibiotics for GAS mainly to prevent purulent complications and acute rheumatic fever, and benefits for symptomatic relief appear modest.^9–12^

Inappropriate antibiotic use promotes antimicrobial resistance.^6,13^ Prolonged treatment increases selective pressure, disrupts the microbiota, and causes adverse effects; shorter courses may reduce these harms and treatment burden.^14–19^

Undertreatment of GAS pharyngitis can cause acute rheumatic fever—an immune reaction that may damage the heart and lead to lifelong disability or death.^20,21^ Whether shorter durations of antibiotic treatment achieve comparable clinical benefits, including similar protection against acute rheumatic fever, remains uncertain. Most guidelines recommend 10 days of oral antibiotics,^9,22,23^ although the World Health Organization recommends 5 days for children at low risk of rheumatic heart disease and the National Institute for Health and Care Excellence recommends 5 to 10 days.^24,25^

The 10-day penicillin standard arose from mid-century studies showing that intramuscular penicillin prevented rheumatic fever during an epidemic and later trials reporting lower bacteriologic eradication with shorter oral courses.^26–30^ Existing reviews compare antibiotics with no treatment, alternative antibiotics, or broad short- vs long-course categories, but do not model duration-response relationships, separate duration from antibiotic choice, consistently assess certainty of evidence, or synthesize serious complications.^11,12,31–33^

We present a systematic review and dose-response meta-analysis of trials examining how the duration of antibiotic treatment affects clinical outcomes in children with suspected or confirmed GAS pharyngitis. This approach models outcomes across durations.^34^ To distinguish between the effect of duration from the choice of antibiotic, we performed additional analyses restricted to trials comparing different durations of the same antibiotic, the same antibiotic class, penicillins, and regimens excluding azithromycin or macrolides. These findings informed the Canadian Antibiotic Treatment Guidance, Canada’s national guidelines on empiric antibiotic prescribing.^35^

## Methods

We registered our protocol on Open Science Framework (osf.io/gfxhz/files/s4qe9) in October 2024 and report our findings following the PRISMA checklist.^36^

### Eligibility Criteria

We included randomized trials of participants aged 18 years or younger with suspected or confirmed GAS pharyngitis. Mixed-age trials were eligible if they reported separate pediatric results or more than 80% of participants were aged 18 years or younger. We defined confirmed GAS as a positive culture or rapid test, and suspected GAS as sore throat accompanied by two or more signs suggesting bacterial infection, such as fever ≥38·0°C, tender cervical lymph nodes, pharyngeal exudates, and absence of cough. We planned sensitivity analyses restricted to confirmed GAS, but all trials limited eligibility to confirmed GAS.

We excluded nonrandomized and crossover trials and trials with fewer than 25 participants per arm to limit potential prognostic imbalance, unrepresentative samples, and publication bias.^37^ Eligible comparisons involved different durations of the same or different oral antibiotics or oral antibiotics vs placebo or no treatment. We excluded nonoral antibiotics because of pharmacokinetic and dosing differences. Abstracts, preprints, and grey literature were eligible.

### Search Strategy

With support from a health research librarian, we searched MEDLINE, Embase, CENTRAL, Web of Science, and CINAHL, from inception to July 14, 2025, without language restrictions (Supplement 1). We supplemented our search with references of eligible records and relevant reviews.^11,12,31,33,38^

### Screening

Following training and calibration to promote accurate application of the eligibility criteria, pairs of reviewers worked independently to screen the titles and abstracts of search records and, subsequently, the full-texts of records deemed potentially eligible at the title and abstract screening stage. Reviewers resolved discrepancies by discussion and, if necessary, by adjudication with a third reviewer. Reviewers screened non-English records either directly, when fluent in the language, or with machine translation using DeepL Pro (deepl.com).

### Data Extraction

Following training and calibration, paired reviewers independently extracted data using a standardized form and resolved disagreements by discussion or adjudication with a third reviewer. For non-English publications, reviewers used machine translation, and a native speaker verified extractions.

We extracted trial design; participant age, setting, symptoms, and diagnostic test results; antibiotic, dose, frequency, and duration; and outcome data. The Canadian Antibiotic Treatment Guidance panel prioritized clinical cure, relapse, adverse events, mortality, serious adverse events, purulent complications, acute rheumatic fever, and poststreptococcal glomerulonephritis. We excluded bacteriologic outcomes and included only clinically defined cure and relapse. We extracted number of participants and events per arm, prioritizing unimputed intention-to-treat data, followed by modified intention-to-treat, per-protocol, and as-treated data.^42,43^

We extracted clinical cure nearest day 7 but not before day 2 and other outcomes at the latest follow-up preserving randomization. When overall cure was unavailable, we used resolution of fever, sore throat, or headache. For adverse events, we prioritized all-cause over treatment-related events. We calculated relapse using the number cured at the preceding follow-up as the denominator; or, if unavailable, all analyzed participants.

### Risk of Bias

Reviewers independently and in duplicate assessed the risk of bias of each outcome from each trial using a modified Cochrane RoB 2 tool,^44,45^ and resolved disagreements by consensus or by a third reviewer. The RoB 2 assesses risk of bias across five domains: bias from randomization, deviations from intended interventions, missing outcome data, outcome measurement, and selective reporting. The modified version of the RoB 2 domains replaced ‘some concerns’ with probably low or high risk to encourage reviewers to make definitive judgments instead of defaulting to an intermediate category.

We assessed the effect of assignment rather than adherence.^46^ We rated trials without blinding of participants, caregivers, or treating clinicians as probably high because we could not exclude co-interventions arising from the trial context—rescue antibiotics, analgesics, or other treatments—that awareness of assignment might prompt. For outcome measurement, lack of blinding indicated high risk for all outcomes except all-cause mortality. Clinical cure, relapse, and adverse events depended on participant- or caregiver-reported symptoms; knowledge of treatment could also influence investigation and classification of serious complications.

### Data Analysis

We used random-effects dose-response meta-analysis with restricted maximum likelihood, treating treatment duration as ‘dose’.^34,47^ We pooled arms of equal duration, applied a 0.5 continuity correction to studies with zero events in one or more arms, and reconstructed covariance using the Greenland-Longnecker method.

Based on the hypothesis that treatment duration follows a non-linear pattern of diminishing returns, we prioritized non-linear models for our primary analyses.^48^ To model non-linear relationships, we used 1-stage restricted cubic splines with knots at the 25th, 50th, and 75th percentiles.^48–51^

When sparse data precluded nonlinear models, we used 2-stage linear models, which also served as secondary analyses.^34,47^ To assess heterogeneity, we considered I^2^ statistics, forest plots for 2-stage linear models, and bubble plots for 1-stage nonlinear models.^52^ Bubble plots overlaid observed study-arm data on fitted curves relative to 10 days. For trials without a 10-day arm, the model estimated the shortest duration vs 10 days and combined this with observed within-trial contrasts for longer durations. For analyses with at least 10 trials, funnel plots and Egger tests assessed small-study effects using study-specific 5- vs 10-day contrasts from linear models.^53^ This approach assumes a linear dose-response relationship, but it yields the independent study-specific estimates.

We report risk ratios (RRs) with 95% CIs and calculated risk differences per 1000 patients using the median risk in 10-day arms.^54^ We present comparisons for 5 and 3 days vs 10 days. We performed all analyses using the meta and dosresmeta packages in R (version 4.4.0).^48,55^

#### Effect modification and sensitivity analyses

We prespecified subgroup analyses by risk of bias and confirmed vs suspected GAS,^56^ but all trials confirmed GAS and only 2 provided data at low or probably low risk of bias, precluding these analyses. Sensitivity analyses restricted comparisons to trials that compared different durations of the same antibiotic at the same dose, the same antibiotic at any dose, antibiotics of the same class, and penicillin-class antibiotics and excluded azithromycin (due to its prolonged tissue half-life) and macrolides. An additional sensitivity analysis included trials that compared antibiotics with placebo or no antibiotics, with the latter reflecting 0 days of treatment. We grouped β-lactams as penicillins, cephalosporins by generation, or β-lactam/β-lactamase inhibitor combinations.

### Certainty of Evidence

We used GRADE to rate certainty separately for 5 vs 10 days and 3 vs 10 days, considering risk of bias, inconsistency, indirectness, publication bias, and imprecision.^57^

Review authors defined decision thresholds representing the smallest differences patients might find important: 2% for mortality; 10% for clinical cure, relapse, and adverse events; and 5% for serious adverse events, acute rheumatic fever, and poststreptococcal glomerulonephritis.^58^ We rated certainty in an important effect when the point estimate exceeded the threshold and otherwise in little or no effect. Confidence intervals and optimal information size informed imprecision judgments. The optimal information size represented the sample size needed for a single trial to detect the decision threshold with 80–90% power.

High certainty indicates that the estimate and true effect likely fall on the same side of the decision threshold; low or very low certainty reflects uncertainty, with the effect possibly falling on either side. We describe our results using GRADE plain language: declarative statements for high, ‘likely’ for moderate, ‘may’ for low, and ‘very uncertain’ for very low certainty.^59^

### Deviations from the Protocol

The protocol also planned network meta-analyses, which will be reported in a separate publication. After librarian consultation, we added Web of Science and CINAHL. We also extended dose-response analyses beyond clinical cure and adverse events.

## Results

Our search yielded 12,683 records, of which 48 records reporting on 45 trials with 22,636 participants proved eligible (Supplements 2 and 3).

### Trial and Participant Characteristics

All 45 trials used parallel designs (Table 1; Supplement 4). The median publication year was 1998 (range, 1956-2024), and most trials occurred in Europe. More than half did not report funding; among those that did, industry support predominated. One trial was published in French and one in German. Participants were predominantly young children (<7 years); All trials confirmed GAS by culture, rapid test, or both. Most participants had fever, tonsillar swelling or exudates, and cervical lymphadenopathy.

**Table 1:**
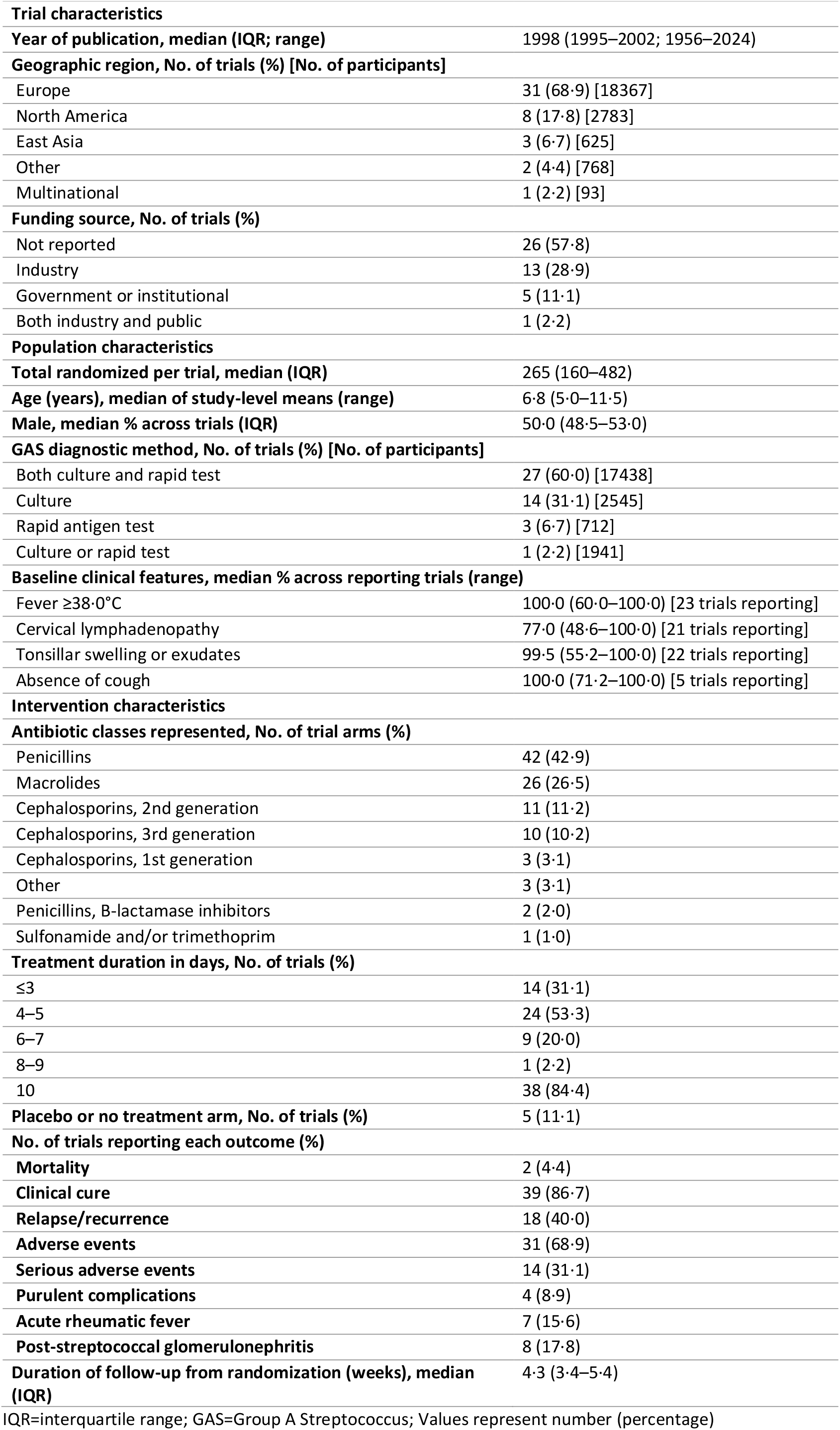
Trial and participant characteristics.

|  |  |
| --- | --- |
| **Trial characteristics** |  |
| **Year of publication, median (IQR; range)** | 1998 (1995–2002; 1956–2024) |
| **Geographic region, No. of trials (%) [No. of participants]** |  |
| Europe | 31 (68.9) [18367] |
| North America | 8 (17.8) [2783] |
| East Asia | 3 (6.7) [625] |
| Other | 2 (4.4) [768] |
| Multinational | 1 (2.2) [93] |
| **Funding source, No. of trials (%)** |  |
| Not reported | 26 (57.8) |
| Industry | 13 (28.9) |
| Government or institutional | 5 (11.1) |
| Both industry and public | 1 (2.2) |
| **Population characteristics** |  |
| **Total randomized per trial, median (IQR)** | 265 (160–482) |
| **Age (years), median of study-level means (range)** | 6.8 (5.0–11.5) |
| **Male, median % across trials (IQR)** | 50.0 (48.5–53.0) |
| **GAS diagnostic method, No. of trials (%) [No. of participants]** |  |
| Both culture and rapid test | 27 (60.0) [17438] |
| Culture | 14 (31.1) [2545] |
| Rapid antigen test | 3 (6.7) [712] |
| Culture or rapid test | 1 (2.2) [1941] |
| **Baseline clinical features, median % across reporting trials (range)** |  |
| Fever $\geq 38.0^{\circ}\text{C}$ | 100.0 (60.0–100.0) [23 trials reporting] |
| Cervical lymphadenopathy | 77.0 (48.6–100.0) [21 trials reporting] |
| Tonsillar swelling or exudates | 99.5 (55.2–100.0) [22 trials reporting] |
| Absence of cough | 100.0 (71.2–100.0) [5 trials reporting] |
| **Intervention characteristics** |  |
| **Antibiotic classes represented, No. of trial arms (%)** |  |
| Penicillins | 42 (42.9) |
| Macrolides | 26 (26.5) |
| Cephalosporins, 2nd generation | 11 (11.2) |
| Cephalosporins, 3rd generation | 10 (10.2) |
| Cephalosporins, 1st generation | 3 (3.1) |
| Other | 3 (3.1) |
| Penicillins, B-lactamase inhibitors | 2 (2.0) |
| Sulfonamide and/or trimethoprim | 1 (1.0) |
| **Treatment duration in days, No. of trials (%)** |  |
| $\leq 3$ | 14 (31.1) |
| 4–5 | 24 (53.3) |
| 6–7 | 9 (20.0) |
| 8–9 | 1 (2.2) |
| 10 | 38 (84.4) |
| **Placebo or no treatment arm, No. of trials (%)** | 5 (11.1) |
| **No. of trials reporting each outcome (%)** |  |
| **Mortality** | 2 (4.4) |
| **Clinical cure** | 39 (86.7) |
| **Relapse/recurrence** | 18 (40.0) |
| **Adverse events** | 31 (68.9) |
| **Serious adverse events** | 14 (31.1) |
| **Purulent complications** | 4 (8.9) |
| **Acute rheumatic fever** | 7 (15.6) |
| **Post-streptococcal glomerulonephritis** | 8 (17.8) |
| **Duration of follow-up from randomization (weeks), median (IQR)** | 4.3 (3.4–5.4) |

IQR=interquartile range; GAS=Group A Streptococcus; Values represent number (percentage)

Trials tested 22 antibiotics in 7 classes; 5 trials included placebo arms (Supplements 5-8). Penicillins and macrolides predominated. Most trials tested 10 days of treatment, more than half tested 4 to 5 days, and all classes had 5- and 10-day data. Of 14 trials testing 3 days or less, 12 evaluated azithromycin; the others tested 3-day amoxicillin-clavulanate and 2-day penicillin V. Among 11 trials comparing durations of the same antibiotic, shorter regimens maintained the same daily dose in 8 and increased it in 3. Median follow-up was 2 weeks for cure, 4 weeks for relapse, and 1 year for acute rheumatic fever.

### Risk of Bias

All but 2 trials had high or probably high risk of bias, mainly because lack of blinding (Figure 1; Supplements 9-15). Approximately 3 quarters also had randomization concerns, often because methods were poorly reported. Many trials predated modern reporting guidance or its widespread adoption, which may explain omissions in reporting. Nearly all predated routine registration and were therefore rated probably low rather than low for selective reporting.

**Figure 1:**
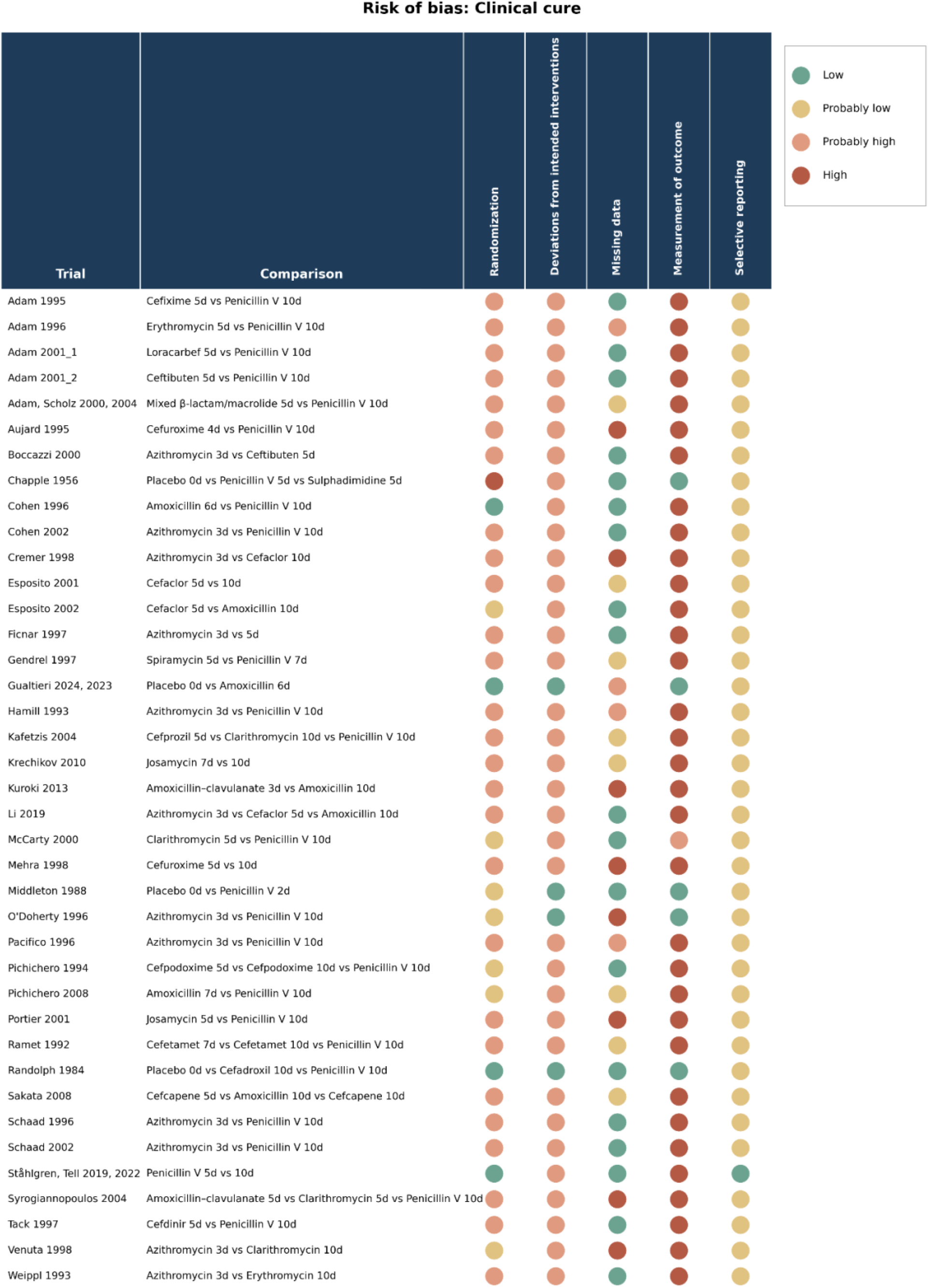
Risk of bias of trials reporting on clinical cure.

**Figure 2.**
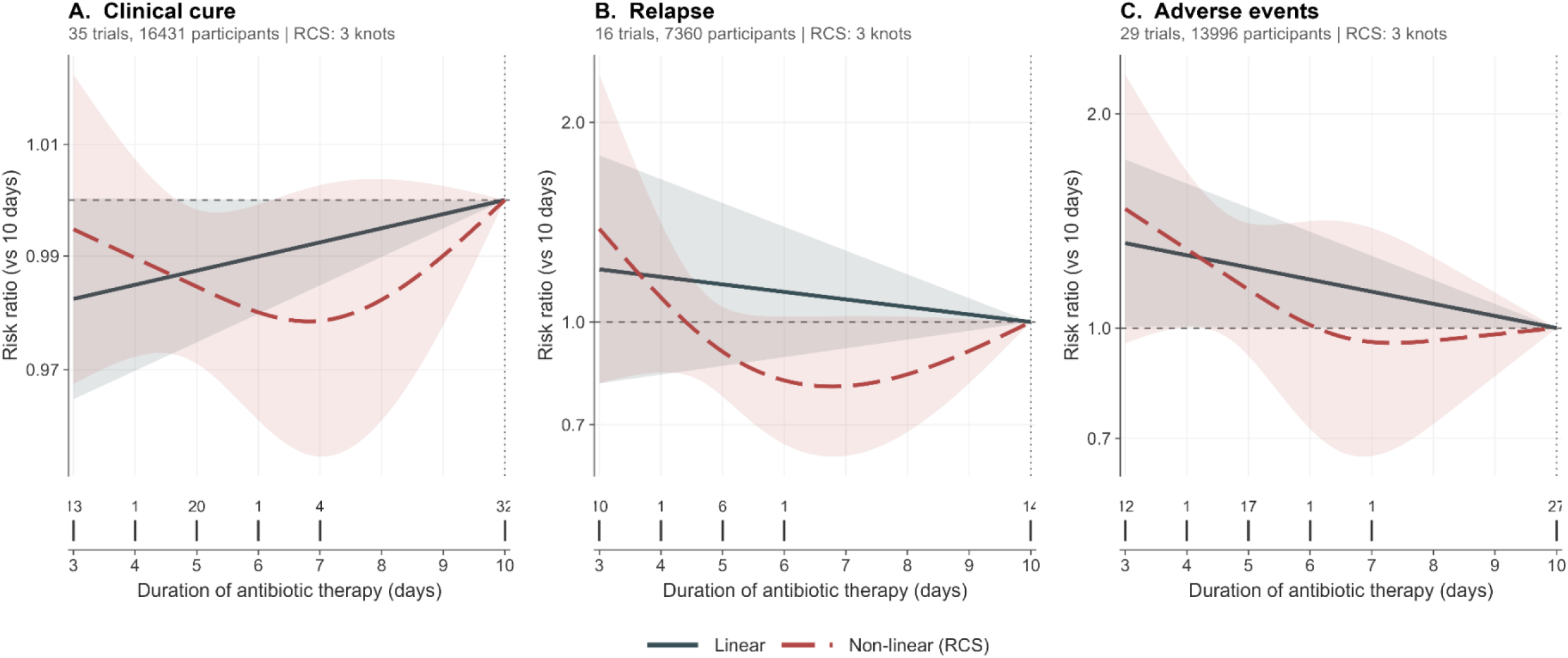
Dose-response relationships between oral antibiotic treatment duration and clinical cure, relapse, and adverse events in children with group A streptococcal pharyngitis. Each panel plots the pooled risk ratio relative to 10 days of therapy. The slate solid curve represents the results of a two-stage linear dose-response model; the red long-dashed curve represents a non-linear a one-stage restricted cubic spline. The shaded bands represent 95% confidence intervals. The horizontal dashed line marks the null (RR=1); the vertical dotted line marks the 10-day reference. A curve above RR=1 indicates a higher event rate at that duration than at 10 days, and a curve below indicates a lower rate. The strip beneath each panel displays the durations observed in the primary analysis: each tick marks a contributing duration, and the number above the tick is the count of trials reporting at that duration.

### Summary of Results

Table 2 summarizes dose-response meta-analysis results and certainty (Supplements 16-18).

**Table 2:**
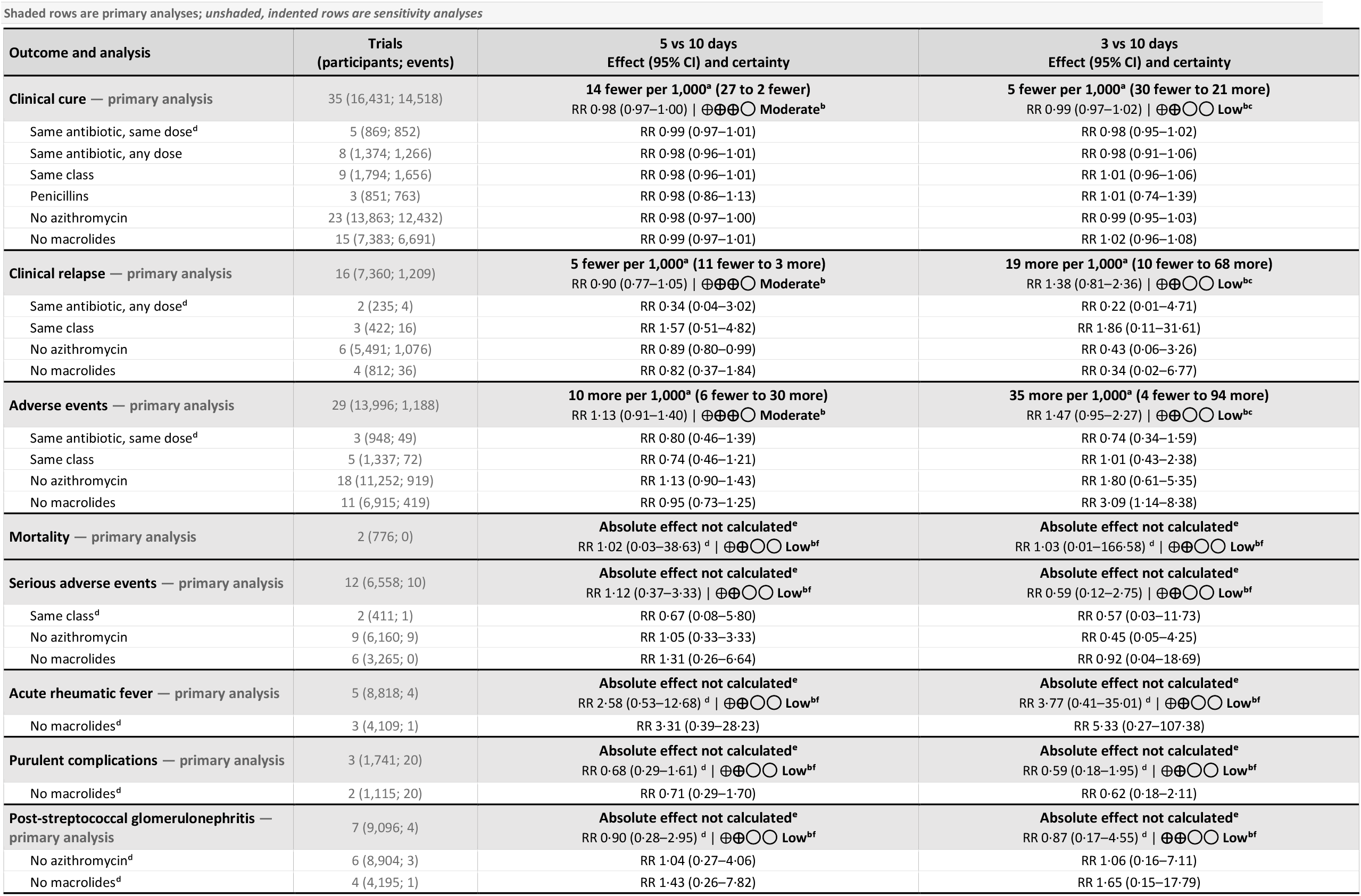
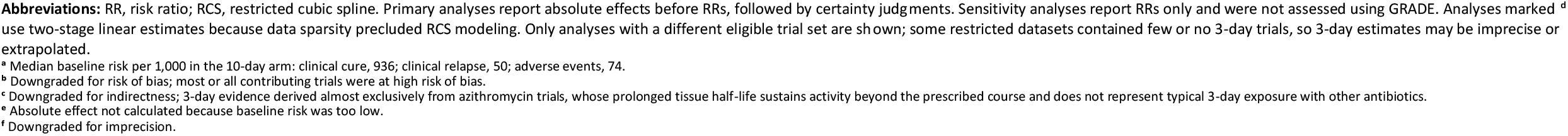
Summary of findings.

| Outcome and analysis | Trials<br>(participants; events) | 5 vs 10 days<br>Effect (95% CI) and certainty | 3 vs 10 days<br>Effect (95% CI) and certainty |
| --- | --- | --- | --- |
| **Clinical cure — primary analysis** | 35 (16,431; 14,518) | **14 fewer per 1,000<sup>a</sup> (27 to 2 fewer)**<br>RR 0.98 (0.97–1.00) \| ⊕⊕⊕○ Moderate<sup>b</sup> | **5 fewer per 1,000<sup>a</sup> (30 fewer to 21 more)**<br>RR 0.99 (0.97–1.02) \| ⊕⊕○○ Low<sup>bc</sup> |
| Same antibiotic, same dose<sup>d</sup> | 5 (869; 852) | RR 0.99 (0.97–1.01) | RR 0.98 (0.95–1.02) |
| Same antibiotic, any dose | 8 (1,374; 1,266) | RR 0.98 (0.96–1.01) | RR 0.98 (0.91–1.06) |
| Same class | 9 (1,794; 1,656) | RR 0.98 (0.96–1.01) | RR 1.01 (0.96–1.06) |
| Penicillins | 3 (851; 763) | RR 0.98 (0.86–1.13) | RR 1.01 (0.74–1.39) |
| No azithromycin | 23 (13,863; 12,432) | RR 0.98 (0.97–1.00) | RR 0.99 (0.95–1.03) |
| No macrolides | 15 (7,383; 6,691) | RR 0.99 (0.97–1.01) | RR 1.02 (0.96–1.08) |
| **Clinical relapse — primary analysis** | 16 (7,360; 1,209) | **5 fewer per 1,000<sup>a</sup> (11 fewer to 3 more)**<br>RR 0.90 (0.77–1.05) \| ⊕⊕⊕○ Moderate<sup>b</sup> | **19 more per 1,000<sup>a</sup> (10 fewer to 68 more)**<br>RR 1.38 (0.81–2.36) \| ⊕⊕○○ Low<sup>bc</sup> |
| Same antibiotic, any dose<sup>d</sup> | 2 (235; 4) | RR 0.34 (0.04–3.02) | RR 0.22 (0.01–4.71) |
| Same class | 3 (422; 16) | RR 1.57 (0.51–4.82) | RR 1.86 (0.11–31.61) |
| No azithromycin | 6 (5,491; 1,076) | RR 0.89 (0.80–0.99) | RR 0.43 (0.06–3.26) |
| No macrolides | 4 (812; 36) | RR 0.82 (0.37–1.84) | RR 0.34 (0.02–6.77) |
| **Adverse events — primary analysis** | 29 (13,996; 1,188) | **10 more per 1,000<sup>a</sup> (6 fewer to 30 more)**<br>RR 1.13 (0.91–1.40) \| ⊕⊕⊕○ Moderate<sup>b</sup> | **35 more per 1,000<sup>a</sup> (4 fewer to 94 more)**<br>RR 1.47 (0.95–2.27) \| ⊕⊕○○ Low<sup>bc</sup> |
| Same antibiotic, same dose<sup>d</sup> | 3 (948; 49) | RR 0.80 (0.46–1.39) | RR 0.74 (0.34–1.59) |
| Same class | 5 (1,337; 72) | RR 0.74 (0.46–1.21) | RR 1.01 (0.43–2.38) |
| No azithromycin | 18 (11,252; 919) | RR 1.13 (0.90–1.43) | RR 1.80 (0.61–5.35) |
| No macrolides | 11 (6,915; 419) | RR 0.95 (0.73–1.25) | RR 3.09 (1.14–8.38) |
| **Mortality — primary analysis** | 2 (776; 0) | **Absolute effect not calculated<sup>e</sup>**<br>RR 1.02 (0.03–38.63) <sup>d</sup> \| ⊕⊕○○ Low<sup>bf</sup> | **Absolute effect not calculated<sup>e</sup>**<br>RR 1.03 (0.01–166.58) <sup>d</sup> \| ⊕⊕○○ Low<sup>bf</sup> |
| **Serious adverse events — primary analysis** | 12 (6,558; 10) | **Absolute effect not calculated<sup>e</sup>**<br>RR 1.12 (0.37–3.33) \| ⊕⊕○○ Low<sup>bf</sup> | **Absolute effect not calculated<sup>e</sup>**<br>RR 0.59 (0.12–2.75) \| ⊕⊕○○ Low<sup>bf</sup> |
| Same class<sup>d</sup> | 2 (411; 1) | RR 0.67 (0.08–5.80) | RR 0.57 (0.03–11.73) |
| No azithromycin | 9 (6,160; 9) | RR 1.05 (0.33–3.33) | RR 0.45 (0.05–4.25) |
| No macrolides | 6 (3,265; 0) | RR 1.31 (0.26–6.64) | RR 0.92 (0.04–18.69) |
| **Acute rheumatic fever — primary analysis** | 5 (8,818; 4) | **Absolute effect not calculated<sup>e</sup>**<br>RR 2.58 (0.53–12.68) <sup>d</sup> \| ⊕⊕○○ Low<sup>bf</sup> | **Absolute effect not calculated<sup>e</sup>**<br>RR 3.77 (0.41–35.01) <sup>d</sup> \| ⊕⊕○○ Low<sup>bf</sup> |
| No macrolides<sup>d</sup> | 3 (4,109; 1) | RR 3.31 (0.39–28.23) | RR 5.33 (0.27–107.38) |
| **Purulent complications — primary analysis** | 3 (1,741; 20) | **Absolute effect not calculated<sup>e</sup>**<br>RR 0.68 (0.29–1.61) <sup>d</sup> \| ⊕⊕○○ Low<sup>bf</sup> | **Absolute effect not calculated<sup>e</sup>**<br>RR 0.59 (0.18–1.95) <sup>d</sup> \| ⊕⊕○○ Low<sup>bf</sup> |
| No macrolides<sup>d</sup> | 2 (1,115; 20) | RR 0.71 (0.29–1.70) | RR 0.62 (0.18–2.11) |
| **Post-streptococcal glomerulonephritis — primary analysis** | 7 (9,096; 4) | **Absolute effect not calculated<sup>e</sup>**<br>RR 0.90 (0.28–2.95) <sup>d</sup> \| ⊕⊕○○ Low<sup>bf</sup> | **Absolute effect not calculated<sup>e</sup>**<br>RR 0.87 (0.17–4.55) <sup>d</sup> \| ⊕⊕○○ Low<sup>bf</sup> |
| No azithromycin<sup>d</sup> | 6 (8,904; 3) | RR 1.04 (0.27–4.06) | RR 1.06 (0.16–7.11) |
| No macrolides<sup>d</sup> | 4 (4,195; 1) | RR 1.43 (0.26–7.82) | RR 1.65 (0.15–17.79) |

**Abbreviations:** RR, risk ratio; RCS, restricted cubic spline. Primary analyses report absolute effects before RRs, followed by certainty judgments. Sensitivity analyses report RRs only and were not assessed using GRADE. Analyses marked <sup>d</sup> use two-stage linear estimates because data sparsity precluded RCS modeling. Only analyses with a different eligible trial set are shown; some restricted datasets contained few or no 3-day trials, so 3-day estimates may be imprecise or extrapolated.
<sup>a</sup> Median baseline risk per 1,000 in the 10-day arm: clinical cure, 936; clinical relapse, 50; adverse events, 74.
<sup>b</sup> Downgraded for risk of bias; most or all contributing trials were at high risk of bias.
<sup>c</sup> Downgraded for indirectness; 3-day evidence derived almost exclusively from azithromycin trials, whose prolonged tissue half-life sustains activity beyond the prescribed course and does not represent typical 3-day exposure with other antibiotics.
<sup>e</sup> Absolute effect not calculated because baseline risk was too low.
<sup>f</sup> Downgraded for imprecision.

### Clinical cure

Thirty-five trials (16,431 participants), testing 3 to 10 days of treatment, informed the primary analysis. Moderate certainty evidence, downgraded for risk of bias, suggests that 5 vs 10 days yields 14 fewer cures per 1000 (95% CI, 27 to 2 fewer), a clinically trivial difference. A 3- vs. 10-day course may result in little or no difference (5 fewer cures per 1000 [30 fewer to 21 more]), but the evidence was low certainty, additionally downgraded for indirectness because it came almost entirely from trials of azithromycin, limiting extrapolation to penicillin V or amoxicillin.

Results were similar after excluding azithromycin or all macrolides and when restricting analyses to trials comparing different durations of the same antibiotic or antibiotic class. Penicillin-only estimates aligned in direction but were imprecise; only 1 trial informed 3-day β-lactam treatment. Linear models and an additional analysis with placebo-controlled trials yielded similar results to the primary analysis. The funnel plot and Egger test showed no evidence of small-study effects (Supplements 22-41).

### Clinical relapse

Sixteen trials (7,360 participants), testing 3 to 10 days of treatment, informed the primary analysis. Moderate certainty evidence, downgraded for risk of bias, suggests little or no difference with 5 vs 10 days (5 fewer relapses per 1000; 95% CI, 11 fewer to 3 more). A 3- vs. 10-day course may result in little or no difference (19 more per 1000; 95% CI, 10 fewer to 68 more), but the evidence was low certainty, additionally downgraded for indirectness because it came almost entirely from trials of azithromycin (Supplements 42-44).

Excluding azithromycin supported the 5-day results with greater precision (RR, 0.89; 95% CI, 0.80-0.99); excluding all macrolides was consistent but less precise. The shortest included course in these analyses was 4 days, so 3-day estimates were extrapolated and imprecise. Same-antibiotic and same-class analyses were also imprecise, and data were insufficient to perform same-dose or penicillin-only analyses. Linear models and an additional analysis with placebo-controlled trials yielded similar results to the primary analysis. We did not find evidence of small-study effects (Supplements 45-59).

### Adverse events

Twenty-nine trials (13,996 participants), testing 3 to 10 days of treatment, informed the primary analysis. Moderate certainty evidence, downgraded for risk of bias, suggests little or no clinically important difference with 5 vs 10 days (10 more events per 1000; 95% CI, 6 fewer to 30 more). A 3- vs. 10-day course may result in little or no difference (35 more per 1000; 95% CI, 4 fewer to 94 more) but the evidence was low certainty, additionally downgraded for indirectness because it came almost entirely from trials of azithromycin (Supplements 60-62).

Sensitivity analyses excluding azithromycin or all macrolides supported the 5-day result but reduced 3-day evidence to 1 amoxicillin-clavulanate trial. The nonlinear model excluding macrolides suggested more adverse events at 3 days, but the estimate was imprecise and the linear model did not reproduce it. Same-antibiotic and same-class analyses also supported the primary result but were imprecise; data were insufficient to allow a penicillin-only analysis. Linear models and an additional analysis with placebo-controlled trials were aligned with the primary analysis. We did not find evidence of small-study effects (Supplements 63-78).

### Mortality, acute rheumatic fever, and other complications

Mortality and serious complications were rare. Sparse data precluded nonlinear modeling except for serious adverse events and so we relied on linear models. Although relative effects were imprecise, the observed event counts made clinically important absolute differences across durations highly improbable. Across evaluated treatment durations (ranging from 0 to 10 days), low certainty evidence, downgraded for risk of bias and imprecision, suggests little to no clinically important difference.

No *deaths* occurred among 776 participants in 2 trials that compared 3 days of azithromycin, 5 days of ceftibuten and clarithromycin, and 10 days of penicillin V (Supplements 79–82).

Fourteen trials reported 15 *serious adverse events* among 7,035 participants. Most were gastrointestinal (severe vomiting, nausea, diarrhea, and abdominal pain) or suggestive of hypersensitivity (pruritus and urticaria), with isolated cases of asthma, transient proteinuria, and appendectomy without appendicitis (Supplements 83–98).

Seven trials reported *acute rheumatic fever*; four events occurred among 9,189 participants. One event occurred among 481 participants assigned to 5 days of loracarbef, and three events occurred among 3,052 participants assigned to 5 days of either amoxicillin-clavulanate, ceftibuten, cefuroxime, loracarbef, clarithromycin, or erythromycin (Supplements 99–107).

Four trials reported 24 purulent *complications* among 1806 participants (Supplements 108–116). Eight trials reported 4 cases of poststreptococcal *glomerulonephritis* among 9184 participants (Supplements 117–125).

## Discussion

### Summary of Findings

This systematic review of 45 randomized trials with more than 20,000 children with confirmed GAS pharyngitis shows moderate certainty evidence that 5 vs 10 days of antibiotic treatment likely produces no clinically important differences in clinical cure (14 fewer per 1000), relapse (5 fewer per 1000), or adverse events (10 more per 1000). Clinical cure results persisted after excluding azithromycin or all macrolides and restricting analyses to trials comparing different durations of the same antibiotic or class, reducing concern that antibiotic choice explains the findings. Evidence for 3 days remains uncertain because nearly all trials tested azithromycin, whose prolonged activity does not establish effects of 3-day penicillins. Notably, severe complications, including acute rheumatic fever, were rare, making important absolute differences across durations highly unlikely in populations resembling those studied, despite imprecise relative estimates.

### Interpretation of Findings

The 10-day standard for GAS pharyngitis traces to mid-century studies, where penicillin prevented acute rheumatic fever during a streptococcal epidemic, and later studies showing residual streptococci after shorter oral courses.^26,60,61^ This rationale assumes incomplete eradication increases rheumatic fever risk, but strain rheumatogenicity, host susceptibility, repeated exposure, and overcrowding may also influence risk.^62^ Acute rheumatic fever can also follow GAS impetigo, which may be especially relevant where rheumatic fever is endemic.^63^ The historical rationale may therefore have limited applicability to most contemporary low-risk populations.^20^ Although we did not assess bacteriologic clearance, the findings suggest that reduced treatment duration may not translate into important differences in patient-important outcomes.

Trials comparing different durations of the same antibiotic usually maintained or increased the daily dose with shorter regimens; the findings support shorter duration, but not lower daily exposure. Most trials lacked blinding due to which we downgraded the certainty of evidence for risk of bias. Participants and providers, however, likely favored longer treatment, which would bias results against shorter courses.

The results for 5- vs 10-day courses appear robust. Results generally persisted after excluding azithromycin or all macrolides and, for clinical cure, restricting analyses to the same antibiotic or class. Three-day findings require caution because data came largely from azithromycin, which remains active beyond the prescribed course.^64,65^

Mortality and serious complications occurred rarely, making important absolute differences across durations unlikely in populations resembling those studied. Only a subset of trials reported these outcomes, but trials omitting them likely observed no events, potentially causing this review to overestimate their incidence. Short follow-up may have missed delayed events, although acute rheumatic fever typically begins 1 to 5 weeks after infection and trials reporting this outcome followed participants for a median of 52 weeks.^62^

Absolute effects depend on baseline risk, which we derived from the median risk in 10-day trial arms. Rheumatic fever is now rare in most high-income settings but persists in the Global South and among some Indigenous and structurally disadvantaged populations.^66–68^ In these higher-risk settings, the same relative effect could translate into larger absolute effects.

Finally, while longer courses should increase adverse events,^69^ we found little difference. We analyzed adverse events as a binary outcome—any versus none—which may obscure harms that accumulate with longer exposure.

### Strengths and Limitations

Strengths include comprehensive search without language restrictions, duplicate screening and extraction, and a focus on patient-important outcomes. Dose-response meta-analysis characterized the full range of oral antibiotic durations, while sensitivity analyses of trials comparing different durations of the same antibiotic or class and analyses excluding azithromycin or all macrolides helped isolate duration from antibiotic choice. We also assessed certainty using GRADE.

Several limitations qualify the findings. Trials of the shortest courses tested almost exclusively azithromycin, leaving too few penicillin or other β-lactam trials to estimate 3-day effects reliably. Sensitivity analyses restricted to the same antibiotic or class were imprecise and sometimes could not exclude clinically important effects. Most macrolide trials also predated contemporary susceptibility patterns, so applicability depends on local resistance.

Continuity corrections for zero-event trials can artificially narrow variance, but likely introduced little bias because nearly all trials used 1:1 allocation, shifting risks proportionally across arms. Retaining these trials preserves informative evidence that mortality and acute rheumatic fever were rare. At the observed baseline risks, even substantial relative effects would yield small absolute differences.

Author-defined decision thresholds remain subjective.^58,70^ We therefore report estimates and CIs to support alternative calculation of absolute effects.

### Implications

Although most guidelines recommend 10 days of penicillin or amoxicillin, some permit 5 to 10 days or recommend 5 days for children at low rheumatic fever risk.^9,22–25^ In populations resembling those studied, 5 days may offer a safe, effective alternative that reduces adherence burden, antibiotic exposure, and selective pressure. Surveillance should monitor rheumatic fever as practice changes.

In high-income countries, acute rheumatic fever incidence is approximately 0.5 to 2 per 100,000, so preventing 1 case may require treating hundreds of thousands of episodes of GAS pharyngitis.^62,67,71,72^ At this scale, extending treatment to prevent few cases exposes children to serious harms, including anaphylaxis and severe cutaneous reactions, and many more to diarrhea, rashes, and yeast infections. Future trials should test 3-day penicillin regimens and whether antibiotics remain necessary in contemporary low-risk settings.

## Conclusion

Moderate certainty evidence suggests no clinically important differences in cure, relapse, or adverse events between 5 and 10 days of antibiotics for pediatric GAS pharyngitis. Findings were similar after excluding macrolides, although evidence on penicillins was sparse. Evidence for 3-day regimens came almost entirely from azithromycin trials and thus remains uncertain. Severe complications, including acute rheumatic fever, were rare. The low observed incidence of these outcomes makes clinically important differences across durations unlikely. These findings support 5-day courses in children at low risk of acute rheumatic fever.

## Data Availability

All data produced in the present study are available upon reasonable request to the authors.

## Contributors

JPL and DZ were responsible for conceptualization, formal analysis, resources, writing the original draft, visualization, and project administration. JPL, ML, BJL, and DZ were responsible for methodology. JPL, MD, ML, BJL, SK, SD, AW, TJ, MRL, AC, HL, ACH, OC, and MB were responsible for screening and data collection. JPL, MD, and DZ were responsible for data curation. All authors contributed to writing, review, and editing. JMP, ME, DY, BJL, ML, and DZ were responsible for supervision.

## AI Disclosures

We used Claude (Anthropic) and ChatGPT (OpenAI) to refine and format text, tables, and figures. All AI-generated outputs were reviewed, edited, and confirmed by the authors, who take full responsibility for the content of this manuscript.

## Data sharing

The study protocol is available at https://osf.io/gfxhz/files/s4qe9. All requests can be made to the corresponding author.

## Acknowledgments

None

