## Supplementary Material for "Optimal Duration of Antibiotic Treatment for Group A Streptococcal Pharyngitis in Children: A Systematic Review and Dose-Response Meta-Analysis"

**Contents**


| Supplement | Title | Page |
|---|---|---|
| 1 | Database search strategies | 5 |
| 2 | Search and screening of eligible records | 14 |
| 3 | Included studies | 15 |
| 4 | Trial and participant characteristics | 19 |
| 5 | Treatment durations by antibiotic | 23 |
| 6 | Treatment durations by antibiotic class | 24 |
| 7 | Antibiotics represented across outcomes | 25 |
| 8 | Antibiotic classes represented across outcomes | 28 |
| 9 | Risk of bias assessment for clinical relapse | 30 |
| 10 | Risk of bias assessment for adverse events | 31 |
| 11 | Risk of bias assessment for mortality | 32 |
| 12 | Risk of bias assessment for serious adverse events | 33 |
| 13 | Risk of bias assessment for acute rheumatic fever | 34 |
| 14 | Risk of bias assessment for purulent complications | 35 |
| 15 | Risk of bias assessment for poststreptococcal glomerulonephritis | 36 |
| 16 | Dose-response meta-analysis results: study and model characteristics | 37 |
| 17 | Dose-response meta-analysis results: linear models | 39 |
| 18 | Dose-response meta-analysis results: restricted cubic spline models | 42 |
| 19 | Clinical cure: dose-response analysis for the primary analysis | 44 |
| 20 | Clinical cure: linear estimates by risk of bias for the primary analysis | 45 |
| 21 | Clinical cure: restricted cubic spline estimates by risk of bias for the primary analysis | 46 |
| 22 | Clinical cure: funnel plot for the primary analysis | 47 |
| 23 | Clinical cure: dose-response analysis restricted to the same antibiotic and dose | 48 |
| 24 | Clinical cure: dose-response analysis restricted to the same antibiotic | 49 |
| 25 | Clinical cure: restricted cubic spline estimates by risk of bias restricted to the same antibiotic | 50 |
| 26 | Clinical cure: dose-response analysis restricted to the same antibiotic class | 51 |
| 27 | Clinical cure: linear estimates by risk of bias restricted to the same antibiotic class | 52 |
| 28 | Clinical cure: restricted cubic spline estimates by risk of bias restricted to the same antibiotic class | 53 |
| 29 | Clinical cure: dose-response analysis restricted to the penicillins | 54 |
| 30 | Clinical cure: linear estimates by risk of bias restricted to the penicillins | 55 |
| 31 | Clinical cure: restricted cubic spline estimates by risk of bias restricted to the penicillins | 56 |
| 32 | Clinical cure: dose-response analysis excluding azithromycin | 57 |
| 33 | Clinical cure: linear estimates by risk of bias excluding azithromycin | 58 |

| Supplement | Title | Page |
|---|---|---|
| 34 | Clinical cure: restricted cubic spline estimates by risk of bias excluding azithromycin | 59 |
| 35 | Clinical cure: dose-response analysis excluding macrolides | 60 |
| 36 | Clinical cure: linear estimates by risk of bias excluding macrolides | 61 |
| 37 | Clinical cure: restricted cubic spline estimates by risk of bias excluding macrolides | 62 |
| 38 | Clinical cure: dose-response analysis including placebo or no treatment as 0 days | 63 |
| 39 | Clinical cure: linear estimates by risk of bias including placebo or no treatment as 0 days | 64 |
| 40 | Clinical cure: restricted cubic spline estimates by risk of bias including placebo or no treatment as 0 days | 65 |
| 41 | Clinical cure: funnel plot including placebo or no treatment as 0 days | 66 |
| 42 | Clinical relapse: dose-response analysis for the primary analysis | 67 |
| 43 | Clinical relapse: linear estimates by risk of bias for the primary analysis | 68 |
| 44 | Clinical relapse: restricted cubic spline estimates by risk of bias for the primary analysis | 69 |
| 45 | Clinical relapse: funnel plot for the primary analysis | 70 |
| 46 | Clinical relapse: dose-response analysis restricted to the same antibiotic | 71 |
| 47 | Clinical relapse: dose-response analysis restricted to the same antibiotic class | 72 |
| 48 | Clinical relapse: linear estimates by risk of bias restricted to the same antibiotic class | 73 |
| 49 | Clinical relapse: restricted cubic spline estimates by risk of bias restricted to the same antibiotic class | 74 |
| 50 | Clinical relapse: dose-response analysis excluding azithromycin | 75 |
| 51 | Clinical relapse: linear estimates by risk of bias excluding azithromycin | 76 |
| 52 | Clinical relapse: restricted cubic spline estimates by risk of bias excluding azithromycin | 77 |
| 53 | Clinical relapse: dose-response analysis excluding macrolides | 78 |
| 54 | Clinical relapse: linear estimates by risk of bias excluding macrolides | 79 |
| 55 | Clinical relapse: restricted cubic spline estimates by risk of bias excluding macrolides | 80 |
| 56 | Clinical relapse: dose-response analysis including placebo or no treatment as 0 days | 81 |
| 57 | Clinical relapse: linear estimates by risk of bias including placebo or no treatment as 0 days | 82 |
| 58 | Clinical relapse: restricted cubic spline estimates by risk of bias including placebo or no treatment as 0 days | 83 |
| 59 | Clinical relapse: funnel plot including placebo or no treatment as 0 days | 84 |
| 60 | Adverse events: dose-response analysis for the primary analysis | 85 |
| 61 | Adverse events: linear estimates by risk of bias for the primary analysis | 86 |
| 62 | Adverse events: restricted cubic spline estimates by risk of bias for the primary analysis | 87 |
| 63 | Adverse events: funnel plot for the primary analysis | 88 |
| 64 | Adverse events: dose-response analysis restricted to the same antibiotic and dose | 89 |
| 65 | Adverse events: dose-response analysis restricted to the same antibiotic | 90 |
| 66 | Adverse events: dose-response analysis restricted to the same antibiotic class | 91 |
| 67 | Adverse events: linear estimates by risk of bias restricted to the same antibiotic class | 92 |
| 68 | Adverse events: restricted cubic spline estimates by risk of bias restricted to the same antibiotic class | 93 |
| 69 | Adverse events: dose-response analysis excluding azithromycin | 94 |
| 70 | Adverse events: linear estimates by risk of bias excluding azithromycin | 95 |

| Supplement | Title | Page |
|---|---|---|
| 71 | Adverse events: restricted cubic spline estimates by risk of bias excluding azithromycin | 96 |
| 72 | Adverse events: dose-response analysis excluding macrolides | 97 |
| 73 | Adverse events: linear estimates by risk of bias excluding macrolides | 98 |
| 74 | Adverse events: restricted cubic spline estimates by risk of bias excluding macrolides | 99 |
| 75 | Adverse events: dose-response analysis including placebo or no treatment as 0 days | 100 |
| 76 | Adverse events: linear estimates by risk of bias including placebo or no treatment as 0 days | 101 |
| 77 | Adverse events: restricted cubic spline estimates by risk of bias including placebo or no treatment as 0 days | 102 |
| 78 | Adverse events: funnel plot including placebo or no treatment as 0 days | 103 |
| 79 | Mortality: dose-response analysis for the primary analysis | 104 |
| 80 | Mortality: linear estimates by risk of bias for the primary analysis | 105 |
| 81 | Mortality: dose-response analysis including placebo or no treatment as 0 days | 106 |
| 82 | Mortality: linear estimates by risk of bias including placebo or no treatment as 0 days | 107 |
| 83 | Serious adverse events: dose-response analysis for the primary analysis | 108 |
| 84 | Serious adverse events: linear estimates by risk of bias for the primary analysis | 109 |
| 85 | Serious adverse events: restricted cubic spline estimates by risk of bias for the primary analysis | 110 |
| 86 | Serious adverse events: funnel plot for the primary analysis | 111 |
| 87 | Serious adverse events: dose-response analysis restricted to the same antibiotic class | 112 |
| 88 | Serious adverse events: linear estimates by risk of bias restricted to the same antibiotic class | 113 |
| 89 | Serious adverse events: dose-response analysis excluding azithromycin | 114 |
| 90 | Serious adverse events: linear estimates by risk of bias excluding azithromycin | 115 |
| 91 | Serious adverse events: restricted cubic spline estimates by risk of bias excluding azithromycin | 116 |
| 92 | Serious adverse events: dose-response analysis excluding macrolides | 117 |
| 93 | Serious adverse events: linear estimates by risk of bias excluding macrolides | 118 |
| 94 | Serious adverse events: restricted cubic spline estimates by risk of bias excluding macrolides | 119 |
| 95 | Serious adverse events: dose-response analysis including placebo or no treatment as 0 days | 120 |
| 96 | Serious adverse events: linear estimates by risk of bias including placebo or no treatment as 0 days | 121 |
| 97 | Serious adverse events: restricted cubic spline estimates by risk of bias including placebo or no treatment as 0 days | 122 |
| 98 | Serious adverse events: funnel plot including placebo or no treatment as 0 days | 123 |
| 99 | Acute rheumatic fever: dose-response analysis for the primary analysis | 124 |
| 100 | Acute rheumatic fever: linear estimates by risk of bias for the primary analysis | 125 |
| 101 | Acute rheumatic fever: dose-response analysis excluding azithromycin | 126 |
| 102 | Acute rheumatic fever: linear estimates by risk of bias excluding azithromycin | 127 |
| 103 | Acute rheumatic fever: dose-response analysis excluding macrolides | 128 |
| 104 | Acute rheumatic fever: linear estimates by risk of bias excluding macrolides | 129 |

| Supplement | Title | Page |
|---|---|---|
| 105 | Acute rheumatic fever: dose-response analysis including placebo or no treatment as 0 days | 130 |
| 106 | Acute rheumatic fever: linear estimates by risk of bias including placebo or no treatment as 0 days | 131 |
| 107 | Acute rheumatic fever: restricted cubic spline estimates by risk of bias including placebo or no treatment as 0 days | 132 |
| 108 | Purulent complications: dose-response analysis for the primary analysis | 133 |
| 109 | Purulent complications: linear estimates by risk of bias for the primary analysis | 134 |
| 110 | Purulent complications: dose-response analysis excluding azithromycin | 135 |
| 111 | Purulent complications: linear estimates by risk of bias excluding azithromycin | 136 |
| 112 | Purulent complications: dose-response analysis excluding macrolides | 137 |
| 113 | Purulent complications: linear estimates by risk of bias excluding macrolides | 138 |
| 114 | Purulent complications: dose-response analysis including placebo or no treatment as 0 days | 139 |
| 115 | Purulent complications: linear estimates by risk of bias including placebo or no treatment as 0 days | 140 |
| 116 | Purulent complications: restricted cubic spline estimates by risk of bias including placebo or no treatment as 0 days | 141 |
| 117 | Poststreptococcal glomerulonephritis: dose-response analysis for the primary analysis | 142 |
| 118 | Poststreptococcal glomerulonephritis: linear estimates by risk of bias for the primary analysis | 143 |
| 119 | Poststreptococcal glomerulonephritis: dose-response analysis excluding azithromycin | 144 |
| 120 | Poststreptococcal glomerulonephritis: linear estimates by risk of bias excluding azithromycin | 145 |
| 121 | Poststreptococcal glomerulonephritis: dose-response analysis excluding macrolides | 146 |
| 122 | Poststreptococcal glomerulonephritis: linear estimates by risk of bias excluding macrolides | 147 |
| 123 | Poststreptococcal glomerulonephritis: dose-response analysis including placebo or no treatment as 0 days | 148 |
| 124 | Poststreptococcal glomerulonephritis: linear estimates by risk of bias including placebo or no treatment as 0 days | 149 |
| 125 | Poststreptococcal glomerulonephritis: restricted cubic spline estimates by risk of bias including placebo or no treatment as 0 days | 150 |

**Supplement 1. Database search strategies**

**Ovid MEDLINE search strategy (1946 to July 11, 2025)**

| Line | Search strategy |
|---|---|
| 1. | exp Pharyngitis/ |
| 2. | (pharyngit* or nasopharyngit* or rhinopharyngit* or tonsillit* or tonsillopharyngit*).mp. |
| 3. | (throat* adj3 (infect* or inflam*)).mp. |
| 4. | (strep* adj3 (throat* or pharyng*)).mp. |
| 5. | (sore adj3 throat*).mp. [mp=title, book title, abstract, original title, name of substance word, subject heading word, floating sub-heading word, keyword heading word, organism supplementary concept word, protocol supplementary concept word, rare disease supplementary concept word, unique identifier, synonyms, population supplementary concept word, anatomy supplementary concept word] |
| 6. | or/1-5 |
| 7. | exp Streptococcal Infections/ |
| 8. | exp Streptococcus/ |
| 9. | gabhs.tw. |
| 10. | streptococc*.mp. |
| 11. | or/7-10 |
| 12. | (throat or phayrng*).mp. |
| 13. | 11 and 12 |
| 14. | 6 or 13 |
| 15. | exp Anti-Bacterial Agents/ |
| 16. | (antibacterial* or anti bacterial*).mp. [mp=title, book title, abstract, original title, name of substance word, subject heading word, floating sub-heading word, keyword heading word, organism supplementary concept word, protocol supplementary concept word, rare disease supplementary concept word, unique identifier, synonyms, population supplementary concept word, anatomy supplementary concept word] |
| 17. | antibiotic*.mp. |
| 18. | exp beta-Lactams/ |
| 19. | exp Aminoglycosides/ |
| 20. | exp Macrolides/ |
| 21. | exp Quinolones/ |
| 22. | exp Sulfonamides/ |
| 23. | exp Tetracyclines/ |
| 24. | (aminoglycoside* or amoxicillin* or amoxycillin* or ampicillin* or azithromycin* or benzylpenicillin* or beta-lactam* or betalactam* or cefaclor* or cefadroxil or cefalexin or cefdinir or cefditoren or cefixime or cefpodoxime or cefprozil or ceLibuten or ceLriaxone or cefuroxime or cephalosporin* or clarithromycin or clavulanic acid* or clindamycin or co-amoxyclav* or doripenem or doxycycline or eratapenem or erythromycin or imipenem or lincomycin or macrolide* or meropenem or moxifloxacin or penicillin* or phenoxymethylpenicillin* or piperacillin* or quinolone* or roxithromycin* or sulfamethoxazole* or sulfonimide* or tetracycline* or ticarcillin or trimethoprim*).tw,nm. |
| 25. | or/15-24 |
| 26. | 14 and 25 |
| 27. | randomized controlled trial.pt. |

| Line | Search strategy |
|---|---|
| 28. | controlled clinical trial.pt. |
| 29. | randomi?ed.ab. |
| 30. | placebo.ab. |
| 31. | drug therapy.fs. |
| 32. | randomly.ab. |
| 33. | trial.ab. |
| 34. | groups.ab. |
| 35. | or/27-34 |
| 36. | exp animals/ not humans.sh. |
| 37. | 35 not 36 |
| 38. | 26 and 37 |

**Embase search strategy (1974 to July 10, 2025)**

| Line | Search strategy |
|---|---|
| 1. | exp pharyngitis/ |
| 2. | (pharyngit* or nasopharyngit* or rhinopharyngit* or tonsillit* or tonsillopharyngit*).mp. [mp=title, abstract, heading word, drug trade name, original title, device manufacturer, drug manufacturer, device trade name, keyword heading word, floating subheading word, candidate term word] |
| 3. | (throat* adj3 (infect* or inflam*)).mp. |
| 4. | (strep* adj3 (throat* or pharyng*)).mp. |
| 5. | (sore adj3 throat*).mp. |
| 6. | or/1-5 |
| 7. | exp Streptococcus infection/ |
| 8. | exp Streptococcus/ |
| 9. | gabhs.tw. |
| 10. | streptococc*.mp. |
| 11. | or/7-10 |
| 12. | (throat or phayrng*).mp. |
| 13. | 11 and 12 |
| 14. | 6 or 13 |
| 15. | exp antibiotic agent/ |
| 16. | (antibacterial* or anti bacterial*).mp. |
| 17. | antibiotic*.mp. |
| 18. | beta lactam/ |
| 19. | aminoglycoside/ |
| 20. | exp macrolide/ |
| 21. | exp quinolone derivative/ |
| 22. | exp sulfonamide/ |
| 23. | exp tetracycline derivative/ |
| 24. | (aminoglycoside* or amoxicillin* or amoxycillin* or ampicillin* or azithromycin* or benzylpenicillin* or beta-lactam* or betalactam* or cefaclor* or cefadroxil or cefalexin or cefdinir or cefditoren or cefixime or cefpodoxime or cefprozil or ceLibuten or ceLriaxone or cefuroxime or cephalosporin* or clarithromycin or clavulanic acid* or clindamycin or co-amoxyclav* or doripenem or doxycycline or eratapenem or erythromycin or imipenem or lincomycin or macrolide* or meropenem or moxifloxacin or penicillin* or phenoxymethylpenicillin* or piperacillin* or quinolone* or roxithromycin* or sulfamethoxazole* or sulfonimide* or tetracycline* or ticarcillin or trimethoprim*).mp. [mp=title, abstract, heading word, drug trade name, original title, device manufacturer, drug manufacturer, device trade name, keyword heading word, floating subheading word, candidate term word] |
| 25. | or/15-24 |
| 26. | 14 and 25 |
| 27. | randomized controlled trial/ |
| 28. | Controlled clinical study/ |
| 29. | random$.ti,ab. |
| 30. | randomization/ |

| Line | Search strategy |
|---|---|
| 31. | intermethod comparison/ |
| 32. | placebo.ti,ab. |
| 33. | (compare or compared or comparison).ti. |
| 34. | ((evaluated or evaluate or evaluating or assessed or assess) and (compare or compared or comparing or comparison)).ab. |
| 35. | (open adj label).ti,ab. |
| 36. | ((double or single or doubly or singly) adj (blind or blinded or blindly)).ti,ab. |
| 37. | double blind procedure/ |
| 38. | parallel group$1.ti,ab. |
| 39. | (crossover or cross over).ti,ab. |
| 40. | ((assign$ or match or matched or allocation) adj5 (alternate or group$1 or intervention$1 or patient$1 or subject$1 or participant$1)).ti,ab. |
| 41. | (assigned or allocated).ti,ab. |
| 42. | (controlled adj7 (study or design or trial)).ti,ab. |
| 43. | (volunteer or volunteers).ti,ab. |
| 44. | human experiment/ |
| 45. | trial.ti. |
| 46. | or/27-45 |
| 47. | (random$ adj sampl$ adj7 ("cross section$" or questionnaire$1 or survey$ or database$1)).ti,ab. not (comparative study/ or controlled study/ or randomi?ed controlled.ti,ab. or randomly assigned.ti,ab.) |
| 48. | Cross-sectional study/ not (randomized controlled trial/ or controlled clinical study/ or controlled study/ or randomi?ed controlled.ti,ab. or control group$1.ti,ab.) |
| 49. | (((case adj control$) and random$) not randomi?ed controlled).ti,ab. |
| 50. | (Systematic review not (trial or study)).ti. |
| 51. | (nonrandom$ not random$).ti,ab. |
| 52. | "Random field$".ti,ab. |
| 53. | (random cluster adj3 sampl$).ti,ab. |
| 54. | (review.ab. and review.pt.) not trial.ti. |
| 55. | "we searched".ab. and (review.ti. or review.pt.) |
| 56. | "update review".ab. |
| 57. | (databases adj4 searched).ab. |
| 58. | (rat or rats or mouse or mice or swine or porcine or murine or sheep or lambs or pigs or piglets or rabbit or rabbits or cat or cats or dog or dogs or cattle or bovine or monkey or monkeys or trout or marmoset$1).ti. and animal experiment/ |
| 59. | Animal experiment/ not (human experiment/ or human/) |
| 60. | or/47-59 |
| 61. | 46 not 60 |
| 62. | 26 and 61 |

**Cochrane Library search strategy (inception to July 14, 2025)**

| Line | Search strategy |
|---|---|
| #1. | MeSH descriptor: [Pharyngitis] explode all trees |
| #2. | pharyngit* or nasopharyngit* or rhinopharyngit* or tonsillit* or tonsillopharyngit* |
| #3. | (throat* NEAR/3 (infect* or inflam*)) |
| #4. | (strep* NEAR/3 (throat* or pharyng*)) |
| #5. | (sore NEAR/3 throat*) |
| #6. | #1 or #2 or #3 or #4 or #5 |
| #7. | MeSH descriptor: [Streptococcal Infections] explode all trees |
| #8. | MeSH descriptor: [Streptococcus] explode all trees |
| #9. | gabhs |
| #10. | streptococc* |
| #11. | #7 or #8 or #9 or #10 |
| #12. | (throat or phayrng*) |
| #13. | #11 and #12 |
| #14. | #6 or #13 |
| #15. | MeSH descriptor: [Anti-Bacterial Agents] explode all trees |
| #16. | antibacterial* or anti bacterial* |
| #17. | antibiotic* |
| #18. | MeSH descriptor: [beta-Lactams] explode all trees |
| #19. | MeSH descriptor: [Aminoglycosides] explode all trees |
| #20. | MeSH descriptor: [Macrolides] explode all trees |
| #21. | MeSH descriptor: [Quinolones] explode all trees |
| #22. | MeSH descriptor: [Sulfonamides] explode all trees |
| #23. | MeSH descriptor: [Tetracyclines] explode all trees |
| #24. | aminoglycoside* or amoxicillin* or amoxycillin* or ampicillin* or azithromycin* or benzylpenicillin* or beta-lactam* or betalactam* or cefaclor* or cefadroxil or cefalexin or cefdinir or cefditoren or cefixime or cefpodoxime or cefprozil or ceLibuten or ceLriaxone or cefuroxime or cephalosporin* or clarithromycin or clavulanic acid* or clindamycin or co-amoxyclav* or doripenem or doxycycline or eratapenem or erythromycin or imipenem or lincomycin or macrolide* or meropenem or moxifloxacin or penicillin* or phenoxymethylpenicillin* or piperacillin* or quinolone* or roxithromycin* or sulfamethoxazole* or sulfonimide* or tetracycline* or ticarcillin or trimethoprim* |
| #25. | #15 or #16 or #17 or #18 or #19 or #20 or #21 or #22 or #23 or #24 |
| #26. | #14 and #25 in Trials |

**Web of Science search strategy (inception to July 14, 2025)**

| Line | Search strategy |
|---|---|
| #1. | pharyngit* or nasopharyngit* or rhinopharyngit* or tonsillit* or tonsillopharyngit* or "sore throat" or "sore throats" (Topic) |
| #2. | TS=(((throat or throats) NEAR/2 (infect* or inflam*))) |
| #3. | TS=((strep or streptococc* or gabhs) NEAR/5 (throat or throats)) |
| #4. | #3 OR #2 OR #1 |
| #5. | TS=(antibiotic* or anti-bacterial* or antibacterial* or aminoglycoside* or amoxicillin* or amoxycillin* or ampicillin* or azithromycin* or benzylpenicillin* or beta-lactam* or betalactam* or cefaclor* or cefadroxil or cefalexin or cefdinir or cefditoren or cefixime or cefpodoxime or cefprozil or ceOibuten or ceOriaxone or cefuroxime or cephalosporin* or clarithromycin or "clavulanic acid*" or clindamycin or co-amoxyclav* or doripenem or doxycycline or eratapenem or erythromycin or imipenem or lincomycin or macrolide* or meropenem or moxifloxacin or penicillin* or phenoxymethylpenicillin* or piperacillin* or quinolone* or roxithromycin* or sulfamethoxazole* or sulfonimide* or tetracycline* or ticarcillin or trimethoprim*) |
| #6. | #4 AND #5 |

| Line | Search strategy |
|---|---|
| #7. | TS=(randomised OR randomized OR randomisation OR randomisation OR placebo* OR (random* AND (allocat* OR assign*)) OR (blind* AND (single OR double OR treble OR triple))) NOT TS=(animal or animals or pisces or fish or fishes or catfish or catfishes or sheatfish or silurus or arius or heteropneustes or clarias or gariepinus or fathead minnow or fathead minnows or pimephales or promelas or cichlidae or trout or trouts or char or chars or salvelinus or salmo or oncorhynchus or guppy or guppies or millionfish or poecilia or goldfish or goldfishes or carassius or auratus or mullet or mullets or mugil or curema or shark or sharks or cod or cods or gadus or morhua or carp or carps or cyprinus or carpio or killifish or eel or eels or anguilla or zander or sander or lucioperca or stizostedion or turbot or turbots or psetta or flatfish or flatfishes or plaice or pleuronectes or platessa or tilapia or tilapias or oreochromis or sarotherodon or common sole or dover sole or solea or zebrafish or zebrafishes or danio or rerio or seabass or dicentrarchus or labrax or morone or lamprey or lampreys or petromyzon or pumpkinseed or pumpkinseeds or lepomis or gibbosus or herring or clupea or harengus or amphibia or amphibian or amphibians or anura or salientia or frog or frogs or rana or toad or toads or bufo or xenopus or laevis or bombina or epidalea or calamita or salamander or salamanders or newt or newts or triturus or reptilia or reptile or reptiles or bearded dragon or pogona or vitticeps or iguana or iguanas or lizard or lizards or anguis fragilis or turtle or turtles or snakes or snake or aves or bird or birds or quail or quails or coturnix or bobwhite or colinus or virginianus or poultry or poultries or fowl or fowls or chicken or chickens or gallus or zebra finch or taeniopygia or guttata or canary or canaries or serinus or canaria or parakeet or parakeets or grasskeet or parrot or parrots or psittacine or psittacines or shelduck or tadorna or goose or geese or branta or leucopsis or woodlark or lullula or flycatcher or ficedula or hypoleuca or dove or doves or geopelia or cuneata or duck or ducks or greylag or graylag or anser or harrier or circus pygargus or red knot or great knot or calidris or canutus or godwit or limosa or lapponica or meleagris or gallopavo or jackdaw or corvus or monedula or ruff or philomachus or pugnax or lapwing or peewit or plover or vanellus or swan or cygnus or columbianus or bewickii or gull or chroicocephalus or ridibundus or albifrons or great tit or parus or aythya or fuligula or streptopelia or risoria or spoonbill or platalea or leucorodia or blackbird or turdus or merula or blue tit or cyanistes or pigeon or pigeons or columba or pintail or anas or starling or sturnus or owl or athene noctua or pochard or ferina or cockatiel or nymphicus or hollandicus or skylark or alauda or tern or sterna or teal or crecca or oystercatcher or haematopus or ostralegus or shrew or shrews or sorex or araneus or crocidura or russula or european mole or talpa or chiroptera or bat or bats or eptesicus or serotinus or myotis or dasycneme or daubentonii or pipistrelle or pipistrellus or cat or cats or felis or catus or feline or dog or dogs or canis or canine or canines or otter or otters or lutra or badger or badgers or meles or fitchew or fitch or foumart or foulmart or ferrets or ferret or polecat or polecats or mustela or putorius or weasel or weasels or fox or foxes or vulpes or common seal or phoca or vitulina or grey seal or halichoerus or horse or horses or equus or equine or equidae or donkey or donkeys or mule or mules or pig or pigs or swine or swines or hog or hogs or boar or boars or porcine or piglet or piglets or sus or scrofa or llama or llamas or lama or glama or deer or deers or cervus or elaphus or cow or cows or bos taurus or bos indicus or bovine or bull or bulls or cattle or bison or bisons or sheep or sheeps or ovis aries or ovine or lamb or lambs or mouflon or mouflons or goat or goats or capra or caprine or chamois or rupicapra or leporidae or lagomorpha or lagomorph or rabbit or rabbits or oryctolagus or cuniculus or laprine or hares or lepus or rodentia or rodent or rodents or murinae or mouse or mice or mus or musculus or murine or woodmouse or apodemus or rat or rats or rattus or norvegicus or guinea pig or guinea pigs or cavia or porcellus or hamster or hamsters or mesocricetus or cricetulus or cricetus or gerbil or gerbils or jird or jirds or meriones or unguiculatus or jerboa or jerboas or jaculus or chinchilla or chinchillas or beaver or beavers or castor fiber or castor canadensis or sciuridae or squirrel or squirrels or sciurus or chipmunk or chipmunks or marmot or marmots or marmota or suslik or susliks or spermophilus or cynomys or cottonrat or cottonrats or sigmodon or vole or voles or microtus or myodes or glareolus or primate or primates or prosimian or prosimians or lemur or lemurs or lemuridae or loris or bush baby or bush babies or bushbaby or bushbabies or galago or galagos or anthropoidea or anthropoids or simian or simians or monkey or monkeys or marmoset or marmosets or callithrix or cebuella or tamarin or tamarins or saguinus or leontopithecus or squirrel monkey or squirrel monkeys or saimiri or night monkey or night monkeys or owl monkey or owl monkeys or douroucoulis or aotus or spider monkey or spider monkeys or ateles or baboon or baboons or papio or rhesus monkey or macaque or macaca or mulatta or cynomolgus or fascicularis or green monkey or green monkeys or chlorocebus or vervet or vervets or pygerythrus or hominoidea or ape or apes or hylobatidae or gibbon or gibbons or siamang or siamangs or nomascus or symphalangus or hominidae or orangutan or orangutans or pongo or chimpanzee or chimpanzees or pan troglodytes or bonobo or bonobos or pan paniscus or gorilla or gorillas or troglodytes) |
| #8. | #6 AND #7 |

**CINAHL search strategy (inception to July 14, 2025)**

| Line | Search strategy |
|---|---|
| S1. | (MH "Pharyngitis+") |
| S2. | TI ( pharyngit* OR nasopharyngit* OR rhinopharyngit* OR tonsillit* OR tonsillopharyngit* ) OR AB ( pharyngit* OR nasopharyngit* OR rhinopharyngit* OR tonsillit* OR tonsillopharyngit* ) |
| S3. | TI ( (throat* N3 (infect* OR inflam*)) ) OR AB ( (throat* N3 (infect* OR inflam*)) ) |
| S4. | TI ( (strep* N3 (throat* OR pharyng*)) ) OR AB ( (strep* N3 (throat* OR pharyng*)) ) |
| S5. | TI ( (sore N3 throat*) ) OR AB ( (sore N3 throat*) ) |
| S6. | S1 OR S2 OR S3 OR S4 OR S5 |
| S7. | (MH "Streptococcal Infections+") |
| S8. | (MH "Streptococcus+") |
| S9. | TI gabhs OR AB gabhs |
| S10. | TI streptococc* OR AB streptococc* |
| S11. | S7 OR S8 OR S9 OR S10 |
| S12. | TI ( throat OR phayrng* ) OR AB ( throat OR phayrng* ) |
| S13. | S11 AND S12 |
| S14. | S6 OR S13 |
| S15. | (MH "Antibiotics+") |
| S16. | TI ( antibacterial* OR "anti bacterial*" ) OR AB ( antibacterial* OR "anti bacterial*" ) |
| S17. | TI antibiotic* OR AB antibiotic* |
| S18. | (MH "Beta-Lactams+") |
| S19. | (MH "Aminoglycosides+") |
| S20. | (MH "Macrolides+") |
| S21. | (MH "Quinolones+") |
| S22. | (MH "Sulfonamides+") |
| S23. | (MH "Tetracyclines+") |
| S24. | TI ( aminoglycoside* OR amoxicillin* OR amoxycillin* OR ampicillin* OR azithromycin* OR benzylpenicillin* OR "beta-lactam*" OR betalactam* OR cefaclor* OR cefadroxil OR cefalexin OR cefdinir OR cefditoren OR cefixime OR cefpodoxime OR cefprozil OR ceLibuten OR ceLriaxone OR cefuroxime OR cephalosporin* OR clarithromycin OR "clavulanic acid*" OR clindamycin OR "co-amoxyclav*" OR doripenem OR doxycycline OR eratapenem OR erythromycin OR imipenem OR lincomycin OR macrolide* OR meropenem OR moxifloxacin OR penicillin* OR phenoxymethylpenicillin* OR piperacillin* OR quinolone* OR roxithromycin* OR sulfamethoxazole* OR sulfonimide* OR tetracycline* OR ticarcillin OR trimethoprim* ) OR AB ( aminoglycoside* OR amoxicillin* OR amoxycillin* OR ampicillin* OR azithromycin* OR benzylpenicillin* OR "beta-lactam*" OR betalactam* OR cefaclor* OR cefadroxil OR cefalexin OR cefdinir OR cefditoren OR cefixime OR cefpodoxime OR cefprozil OR ceLibuten OR ceLriaxone OR cefuroxime OR cephalosporin* OR clarithromycin OR "clavulanic acid*" OR clindamycin OR "co-amoxyclav*" OR doripenem OR doxycycline OR eratapenem OR erythromycin OR imipenem OR lincomycin OR macrolide* OR meropenem OR moxifloxacin OR penicillin* OR phenoxymethylpenicillin* OR piperacillin* OR quinolone* OR roxithromycin* OR sulfamethoxazole* OR sulfonimide* OR tetracycline* OR ticarcillin OR trimethoprim* ) |
| S25. | S15 OR S16 OR S17 OR S18 OR S19 OR S20 OR S21 OR S22 OR S23 OR S24 |
| S26. | S14 AND S25 |
| S27. | PT clinical trial |

| Line | Search strategy |
|---|---|
| S28. | (MH clinical trials+) |
| S29. | TI clinic* trial* |
| S30. | AB clinic* trial* |
| S31. | TI ( (singl* OR doubl* OR trebl* OR tripl*) W1 (blind* OR mask*) ) OR AB ( (singl* OR doubl* OR trebl* OR tripl*) W1 (blind* OR mask*) ) |
| S32. | MH cluster sample |
| S33. | TI (randomised OR randomized) |
| S34. | AB (random*) |
| S35. | TI (trial) |
| S36. | MH (sample size) AND AB (assigned OR allocated OR control) |
| S37. | MH (placebos) |
| S38. | PT (randomized controlled trial) |
| S39. | AB (control W5 group) |
| S40. | MH (crossover design) OR MH (comparative studies) |
| S41. | AB (cluster W3 RCT) |
| S42. | MH animals+ |
| S43. | MH (animal studies) |
| S44. | TI (animal model*) |
| S45. | S42 OR S43 OR S44 |
| S46. | MH (HUMAN) |
| S47. | S45 NOT S46 |
| S48. | S27 OR S28 OR S29 OR S30 OR S31 OR S32 OR S33 OR S34 OR S35 OR S36 OR S37 OR S38 OR S39 OR S40 OR S41 |
| S49. | S48 NOT S47 |
| S50. | S26 AND S49 |
| S51. | S50 [Limiters - Publication Date: 20241001-20250731] |

**Supplement 2. Search and screening of eligible records**

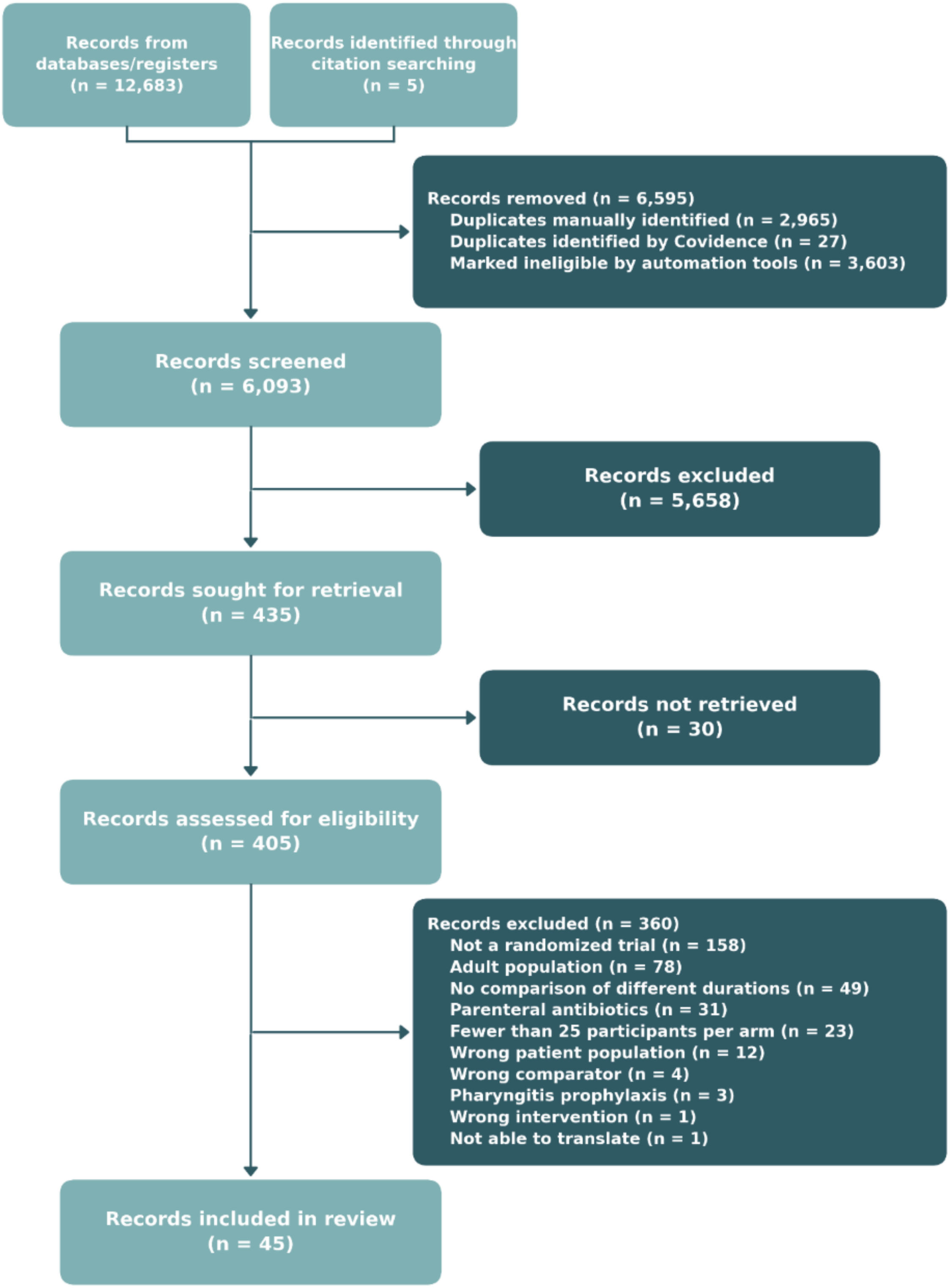


Flow of records through identification, screening, eligibility assessment, and inclusion.

**Supplement 3. Included studies**

**Supplement 4. Trial and participant characteristics**

| Study, Author (Trial name/ Registration | Funding | Country | Male (%) | Age (mean) | Patients tested with rapid test or culture (%) | Patients tested with rapid test (%) | Patients tested with culture (%) | Patients tested GAS positive - rapid test (%) | Patients tested GAS positive - culture (%) |
|---|---|---|---|---|---|---|---|---|---|
| Adam 1995 | NR | Germany | 56.95 | 5.45 | 100 | 100 | NR | 90.07 | NR |
| Adam 1996 | Infectopharm Arzneimittel GmbH, Germany. | Germany | 51.24 | 7.1 | 100 | 100 | 100 | 100 | 100 |
| Adam 2000 | NA | Germany | 49.42 | 6.05 | 100 | 100 | 100 | 100 | 100 |
| Adam 2001_1 | NR | Germany | 50 | 6.2 | 100 | 100 | 100 | 100 | 100 |
| Adam 2001_2 | Schering AG, Berlin, Germany- a pharmaceutical company involved in the development and promotion of ceftibuten at the time | Germany | 48.8 | 6.1 | 100 | NR | NR | NR | 100 |
| Adam, Scholz 2000, 2004 | Cascan, Hamburg; Essex Pharma, Munich; Glaxo Wellcome, Hamburg; Infectopharm Arzneimittel und Consilium, Heppen heim; Lilly Deutschland, Bad Homburg; and SmithKline Beecham Pharma, Munich. | Germany | 50.3 | 6.1 | 100 | 100 | 100 | 100 | 100 |
| Aujard 1995 | NR | France | 46 | 6.95 | 100 | 100 | 100 | 100 | 100 |
| Boccazzi 2000 | NR | Italy | 54.71 | 7.73 | 100 | 100 | 100 | 100 | 91.67 |
| Chapple 1956 | NR | United Kingdom | NR | NR | 100 | NR | 100 | NR | 42.86 |
| Cohen 1996 | SmithKline Beecham laboratory | France | 48.11 | 5.9 | 100 | NR | 100 | NR | 100 |
| Cohen 2002 | Pfizer France, Orsay, France | France | 53.69 | 6.03 | NR | 100 | 100 | 100 | 93.61 |
| Cremer 1998 | NR | Germany | 52.45 | 6.05 | 100 | NR | 100 | NR | 100 |
| Esposito 2001 | NR | Italy | 44.17 | NR | 100 | NR | 100 | NR | 100 |
| Esposito 2002 | Eli Lily Pharmaceuticals | Italy | 51.04 | 5.5 | 100 | 100 | 100 | 100 | 100 |
| Ficnar 1997 | NR | Croatia | 53.1 | 5 | 100 | NR | 100 | NR | 100 |
| Gendrel 1997 | Rhone-Poulenc Rorer. | France | 53.02 | 6.7 | 100 | 100 | 100 | 100 | 100 |

| Study, Author (Trial name/ Registration | Funding | Country | Male (%) | Age (mean) | Patients tested with rapid test or culture (%) | Patients tested with rapid test (%) | Patients tested with culture (%) | Patients tested GAS positive - rapid test (%) | Patients tested GAS positive - culture (%) |
|---|---|---|---|---|---|---|---|---|---|
| Gualtieri 2024, 2023 | University of Geneva. Gertrude Von Meissner Foundation grant, Geneva University Hospitals PRD grant (#1-2015-I) and Société Académique de Genève grant (#2015/36). | Switzerland | 50 | 7.38 | 100 | 100 | 72.72 | 100 | 82.81 |
| Hamill 1993 | NR | Ireland, United Kingdom | 53.13 | 7.45 | 100 | 100 | 100 | 100 | 88.54 |
| Kafetzis 2004 | NR | Greece | 55.85 | 6 | 100 | 100 | 100 | 100 | 100 |
| Krechikov 2010 | NR | Russia | NR | 11.5 | 100 | NR | 100 | NR | 40.17 |
| Kuroki 2013 | The lead author received financial aid from GlaxoSmithKline K.K | Japan | 48.45 | 5.47 | 100 | 100 | NR | 100 | NR |
| Li 2019 | NR | China | 52.34 | 5.77 | 100 | NR | 100 | NR | 100 |
| Lucht 1996 | NR | France | 54.24 | 7.8 | 100 | 100 | 100 | 100 | 88.47 |
| McCarty 2000 | NR | United States | 54.73 | 7.57 | 100 | 100 | 100 | NR | 100 |
| Mehra 1998 | NR | NR | NR | NR | 100 | 100 | 100 | 100 | 100 |
| Middleton 1988 | Supported in part by a grant (AA-3) from the Health Research and Services Foundation, an agency of the Allegheny County United Way. | United States | 50 | 4 to 29 | 100 | NR | 100 | NR | 100 |
| Milatovic 1991 (Study B) | NR | Germany | NR | NR | 100 | NR | 100 | NR | 100 |
| O'Doherty 1996 | NR | Ireland, Sweden, United Kingdom | 48.26 | 7.77 | 100 | 100 | 100 | 100 | 100 |
| Pacifico 1996 | Italian National Research Council as part of targeted project ‘‘Prevention and Control Disease Factors’’ | Italy | 48.7 | 6.8 | 100 | 100 | 100 | 100 | 100 |
| Pichichero 1994 | Upjohn | United States | NR | 7.84 | 100 | 100 | 100 | 100 | 91.32 |
| Pichichero 2008 | NR | United States | 49.29 | 6.84 | 100 | 100 | 100 | 100 | NR |

| Study, Author (Trial name/ Registration | Funding | Country | Male (%) | Age (mean) | Patients tested with rapid test or culture (%) | Patients tested with rapid test (%) | Patients tested with culture (%) | Patients tested GAS positive - rapid test (%) | Patients tested GAS positive - culture (%) |
|---|---|---|---|---|---|---|---|---|---|
| Portier 2001 | NR | France | 44.75 | 6.75 | 100 | 100 | 100 | 100 | 100 |
| Ramet 1992 | NR | Belgium | NR | 5.65 | 100 | 100 | 100 | 100 | 85.47 |
| Randolph 1985 | Mead Johnson and Company | United States | NR | 8.8 | 100 | 0 | 100 | NR | 100 |
| Sakata 2008 | NR | Japan | 55.99 | 5.6 | 100 | NR | 100 | NR | 100 |
| Schaad 1996 | NR | Switzerland | 49.85 | 7 | 100 | 100 | 100 | 100 | 93.88 |
| Schaad 2002 | NR | Switzerland | NR | 2 to 12 | 100 | 100 | 100 | 100 | 97.95 |
| Schwartz 1981 | This study was funded in part by a grant from the American Academy of Pediatrics Memorial and Endowment Fund | United States | 49.21 | 7.85 | 100 | NR | 100 | NR | 100 |
| Ståhlgren, Tell 2019, 2022 | Public Health Agency of Sweden | Sweden | 36.02 | 30.5 (only pediatric subgroup included in analyses) | 100 | 100 | NR | 100 | NR |
| Stillerman 1970 | Supported by a grant from Eli Lilly and Co. | United States | 45.57 | NR | 100 | NR | 100 | NR | 100 |
| Syrogiannopoulos 2004 | Supported in part by a grant from Abbott Laboratories and grant 2491 from the Research Committee of the University of Patras. | Greece | 51.21 | 7.5 | 100 | 100 | 100 | 100 | 100 |
| Tack 1997 | This study was supported by a grant from Parke-Davis Pharmaceutical Research | United States | 51.87 | 7.4 (Cefdinir), 7.7 (Pencillin V) | 100 | 100 | 100 | 100 | 100 |
| Venuta 1998 | NR | Italy | 47.45 | 8.08 (Clarithromycin), 7.92 (Azithromycin) | 100 | 100 | 100 | 100 | 100 |
| Weippl 1993 | NR | Argentina, Austria, Italy | 52.69 | 5.2 | 100 | NR | 100 | NR | 100 |

| Study, Author (Trial name/ Registration | Funding | Country | Male (%) | Age (mean) | Patients tested with rapid test or culture (%) | Patients tested with rapid test (%) | Patients tested with culture (%) | Patients tested GAS positive - rapid test (%) | Patients tested GAS positive - culture (%) |
|---|---|---|---|---|---|---|---|---|---|
| Zwart 2003 | Funded by the Groene Land Achmea Health Insurances and the Stichting Gezondheidszorgonderzoek Ysselmond in Zwolle. | Netherlands | 44.87 | 10.18 | 100 | NR | 100 | NR | 62.18 |

**Supplement 5. Treatment durations by antibiotic**

| Antibiotic class | Antibiotic | 2 days | 3 days | 4 days | 5 days | 6 days | 7 days | 8 days | 10 days | Total trials (participants) |
|---|---|---|---|---|---|---|---|---|---|---|
| Cephalosporins, 1st generation | Cefadroxil | 0 | 0 | 0 | 1 (104) | 0 | 0 | 0 | 1 (70) | 2 (174) |
| Cephalosporins, 1st generation | Cefatrizine | 0 | 0 | 0 | 0 | 0 | 0 | 1 (200) | 0 | 1 (200) |
| Cephalosporins, 1st generation | Cephaloglycin | 0 | 0 | 0 | 0 | 0 | 0 | 0 | 1 (79) | 1 (79) |
| Cephalosporins, 2nd generation | Cefaclor | 0 | 0 | 0 | 3 (350) | 0 | 0 | 0 | 2 (128) | 4 (478) |
| Cephalosporins, 2nd generation | Cefprozil | 0 | 0 | 0 | 1 (88) | 0 | 0 | 0 | 0 | 1 (88) |
| Cephalosporins, 2nd generation | Cefuroxime axetil | 0 | 0 | 1 (152) | 2 (920) | 0 | 0 | 0 | 1 (324) | 3 (1,396) |
| Cephalosporins, 2nd generation | Loracarbef | 0 | 0 | 0 | 1 (544) | 0 | 0 | 0 | 0 | 1 (544) |
| Cephalosporins, 3rd generation | Cefcapene-pivoxil | 0 | 0 | 0 | 1 (82) | 0 | 0 | 0 | 1 (88) | 1 (170) |
| Cephalosporins, 3rd generation | Cefdinir | 0 | 0 | 0 | 1 (240) | 0 | 0 | 0 | 0 | 1 (240) |
| Cephalosporins, 3rd generation | Cefetamet pivoxil | 0 | 0 | 0 | 0 | 0 | 1 (40) | 0 | 1 (37) | 1 (77) |
| Cephalosporins, 3rd generation | Cefixime | 0 | 0 | 0 | 1 (80) | 0 | 0 | 0 | 0 | 1 (80) |
| Cephalosporins, 3rd generation | Cefpodoxime proxetil | 0 | 0 | 0 | 1 (161) | 0 | 0 | 0 | 1 (160) | 1 (321) |
| Cephalosporins, 3rd generation | Ceftibuten | 0 | 0 | 0 | 2 (645) | 0 | 0 | 0 | 0 | 2 (645) |
| Macrolides | Azithromycin | 0 | 12 (1,560) | 0 | 1 (40) | 0 | 0 | 0 | 0 | 12 (1,600) |
| Macrolides | Clarithromycin | 0 | 0 | 0 | 2 (581) | 0 | 0 | 0 | 2 (173) | 4 (754) |
| Macrolides | Erythromycin | 0 | 0 | 0 | 0 | 0 | 0 | 0 | 1 (47) | 1 (47) |
| Macrolides | Erythromycin estolate | 0 | 0 | 0 | 1 (115) | 0 | 0 | 0 | 0 | 1 (115) |
| Macrolides | Josamycin | 0 | 0 | 0 | 1 (168) | 0 | 1 (57) | 0 | 1 (60) | 2 (285) |
| Macrolides | Spiramycin | 0 | 0 | 0 | 1 (149) | 0 | 0 | 0 | 0 | 1 (149) |
| Penicillins | Amoxicillin | 0 | 0 | 0 | 0 | 2 (203) | 1 (290) | 0 | 4 (410) | 7 (903) |
| Penicillins | Penicillin V | 1 (91) | 0 | 0 | 1 (215) | 0 | 2 (195) | 0 | 24 (9,064) | 27 (9,565) |
| Penicillins | Penicillin V potassium | 0 | 0 | 0 | 1 (99) | 0 | 2 (128) | 0 | 4 (583) | 5 (810) |
| Penicillins, β-lactamase inhibitors | Amoxicillin-clavulanate | 0 | 1 (64) | 0 | 1 (155) | 0 | 0 | 0 | 0 | 2 (219) |
| Sulfonamide and/or trimethoprim | Sulfadimidine | 0 | 0 | 0 | 1 (87) | 0 | 0 | 0 | 0 | 1 (87) |
| Other | Amoxicillin-clavulanate, ceftibuten, cefuroxime axetil, loracarbef, clarithromycin or erythromycin | 0 | 0 | 0 | 1 (3,214) | 0 | 0 | 0 | 0 | 1 (3,214) |

### Supplement 6. Treatment durations by antibiotic class

| Antibiotic class | 2 days | 3 days | 4 days | 5 days | 6 days | 7 days | 8 days | 10 days | Total trials (participants) |
|---|---|---|---|---|---|---|---|---|---|
| Cephalosporins, 1st generation | 0 | 0 | 0 | 1 (104) | 0 | 0 | 1 (200) | 2 (149) | 4 (453) |
| Cephalosporins, 2nd generation | 0 | 0 | 1 (152) | 7 (1,902) | 0 | 0 | 0 | 3 (452) | 9 (2,506) |
| Cephalosporins, 3rd generation | 0 | 0 | 0 | 6 (1,208) | 0 | 1 (40) | 0 | 3 (285) | 7 (1,533) |
| Macrolides | 0 | 12 (1,560) | 0 | 6 (1,053) | 0 | 1 (57) | 0 | 4 (280) | 19 (2,950) |
| Penicillins | 1 (91) | 0 | 0 | 2 (314) | 2 (203) | 5 (613) | 0 | 32 (10,057) | 37 (11,278) |
| Penicillins, β-lactamase inhibitors | 0 | 1 (64) | 0 | 1 (155) | 0 | 0 | 0 | 0 | 2 (219) |
| Sulfonamide and/or trimethoprim | 0 | 0 | 0 | 1 (87) | 0 | 0 | 0 | 0 | 1 (87) |
| Other | 0 | 0 | 0 | 1 (3,214) | 0 | 0 | 0 | 0 | 1 (3,214) |

**Supplement 7. Antibiotics represented across outcomes**

| Outcome | Antibiotic | Antibiotic class | Studies | Arms | Durations |
|---|---|---|---|---|---|
| **Clinical cure** | Amoxicillin | Penicillins | 6 | 6 | 6, 7, 10 |
| Clinical cure | Amoxicillin-clavulanate | Penicillins, β-lactamase inhibitors | 2 | 2 | 3, 5 |
| Clinical cure | Amoxicillin-clavulanate, ceftibuten, cefuroxime axetil, loracarbef, clarithromycin or erythromycin | NA | 1 | 1 | 5 |
| Clinical cure | Azithromycin | Macrolide | 12 | 15 | 3, 5 |
| Clinical cure | Cefaclor | Cephalosporins 2nd Gen | 4 | 5 | 5, 10 |
| Clinical cure | Cefcapene-pivoxil | Cephalosporins 3rd Gen | 1 | 2 | 5, 10 |
| Clinical cure | Cefdinir | Cephalosporins 3rd Gen | 1 | 1 | 5 |
| Clinical cure | Cefetamet pivoxil | Cephalosporins 3rd Gen | 1 | 2 | 7, 10 |
| Clinical cure | Cefixime | Cephalosporins 3rd Gen | 1 | 1 | 5 |
| Clinical cure | Cefpodoxime proxetil | Cephalosporins 3rd Gen | 1 | 2 | 5, 10 |
| Clinical cure | Cefprozil | Cephalosporins 2nd Gen | 1 | 1 | 5 |
| Clinical cure | Ceftibuten | Cephalosporins 3rd Gen | 2 | 2 | 5 |
| Clinical cure | Cefuroxime axetil | Cephalosporins 2nd Gen | 2 | 3 | 4, 5, 10 |
| Clinical cure | Clarithromycin | Macrolide | 4 | 5 | 5, 10 |
| Clinical cure | Erythromycin | Macrolide | 1 | 1 | 10 |
| Clinical cure | Erythromycin estolate | Macrolide | 1 | 1 | 5 |
| Clinical cure | Josamycin | Macrolide | 2 | 3 | 5, 7, 10 |
| Clinical cure | Loracarbef | Cephalosporins 2nd Gen | 1 | 1 | 5 |
| Clinical cure | Penicillin V | Penicillins | 21 | 22 | 5, 7, 10 |
| Clinical cure | Penicillin V potassium | Penicillins | 2 | 2 | 10 |
| Clinical cure | Spiramycin | Macrolide | 1 | 1 | 5 |
| **Relapse** | Amoxicillin | Penicillins | 2 | 2 | 6, 10 |
| Relapse | Amoxicillin-clavulanate, ceftibuten, cefuroxime axetil, loracarbef, clarithromycin or erythromycin | NA | 1 | 1 | 5 |
| Relapse | Azithromycin | Macrolide | 10 | 13 | 3, 5 |
| Relapse | Cefaclor | Cephalosporins 2nd Gen | 1 | 1 | 10 |
| Relapse | Cefcapene-pivoxil | Cephalosporins 3rd Gen | 1 | 2 | 5, 10 |
| Relapse | Cefixime | Cephalosporins 3rd Gen | 1 | 1 | 5 |
| Relapse | Ceftibuten | Cephalosporins 3rd Gen | 1 | 1 | 5 |
| Relapse | Cefuroxime axetil | Cephalosporins 2nd Gen | 1 | 1 | 4 |
| Relapse | Erythromycin | Macrolide | 1 | 1 | 10 |
| Relapse | Erythromycin estolate | Macrolide | 1 | 1 | 5 |
| Relapse | Penicillin V | Penicillins | 11 | 11 | 10 |
| **Adverse events** | Amoxicillin | Penicillins | 5 | 5 | 6, 10 |
| Adverse events | Amoxicillin-clavulanate | Penicillins, β-lactamase inhibitors | 2 | 2 | 3, 5 |
| Adverse events | Amoxicillin-clavulanate, ceftibuten, cefuroxime axetil, loracarbef, clarithromycin or erythromycin | NA | 1 | 1 | 5 |
| Adverse events | Azithromycin | Macrolide | 11 | 13 | 3 |
| Adverse events | Cefaclor | Cephalosporins 2nd Gen | 4 | 5 | 5, 10 |
| Adverse events | Cefcapene-pivoxil | Cephalosporins 3rd Gen | 1 | 2 | 5, 10 |
| Adverse events | Cefdinir | Cephalosporins 3rd Gen | 1 | 1 | 5 |
| Adverse events | Cefixime | Cephalosporins 3rd Gen | 1 | 1 | 5 |
| Adverse events | Cefprozil | Cephalosporins 2nd Gen | 1 | 1 | 5 |
| Adverse events | Ceftibuten | Cephalosporins 3rd Gen | 2 | 2 | 5 |
| Adverse events | Cefuroxime axetil | Cephalosporins 2nd Gen | 2 | 3 | 4, 5, 10 |
| Adverse events | Clarithromycin | Macrolide | 4 | 5 | 5, 10 |
| Adverse events | Erythromycin | Macrolide | 1 | 1 | 10 |

| Outcome | Antibiotic | Antibiotic class | Studies | Arms | Durations |
|---|---|---|---|---|---|
| Adverse events | Erythromycin estolate | Macrolide | 1 | 1 | 5 |
| Adverse events | Josamycin | Macrolide | 1 | 1 | 5 |
| Adverse events | Loracarbef | Cephalosporins 2nd Gen | 1 | 1 | 5 |
| Adverse events | Penicillin V | Penicillins | 19 | 19 | 7, 10 |
| Adverse events | Spiramycin | Macrolide | 1 | 1 | 5 |
| **Mortality** | Azithromycin | Macrolide | 1 | 1 | 3 |
| Mortality | Ceftibuten | Cephalosporins 3rd Gen | 1 | 1 | 5 |
| Mortality | Clarithromycin | Macrolide | 1 | 1 | 5 |
| Mortality | Penicillin V | Penicillins | 1 | 1 | 10 |
| **Serious adverse events** | Amoxicillin | Penicillins | 3 | 3 | 6, 10 |
| Serious adverse events | Amoxicillin-clavulanate | Penicillins, β-lactamase inhibitors | 1 | 1 | 3 |
| Serious adverse events | Amoxicillin-clavulanate, ceftibuten, cefuroxime axetil, loracarbef, clarithromycin or erythromycin | NA | 1 | 1 | 5 |
| Serious adverse events | Azithromycin | Macrolide | 3 | 3 | 3 |
| Serious adverse events | Cefaclor | Cephalosporins 2nd Gen | 2 | 2 | 5, 10 |
| Serious adverse events | Cefetamet pivoxil | Cephalosporins 3rd Gen | 1 | 2 | 7, 10 |
| Serious adverse events | Cefuroxime axetil | Cephalosporins 2nd Gen | 2 | 2 | 4, 5 |
| Serious adverse events | Clarithromycin | Macrolide | 1 | 1 | 5 |
| Serious adverse events | Erythromycin | Macrolide | 1 | 1 | 10 |
| Serious adverse events | Penicillin V | Penicillins | 8 | 8 | 7, 10 |
| Serious adverse events | Spiramycin | Macrolide | 1 | 1 | 5 |
| **Acute rheumatic fever** | Amoxicillin | Penicillins | 1 | 1 | 10 |
| Acute rheumatic fever | Amoxicillin-clavulanate, ceftibuten, cefuroxime axetil, loracarbef, clarithromycin or erythromycin | NA | 1 | 1 | 5 |
| Acute rheumatic fever | Cefcapene-pivoxil | Cephalosporins 3rd Gen | 1 | 2 | 5, 10 |
| Acute rheumatic fever | Cefuroxime axetil | Cephalosporins 2nd Gen | 1 | 1 | 5 |
| Acute rheumatic fever | Erythromycin estolate | Macrolide | 1 | 1 | 5 |
| Acute rheumatic fever | Loracarbef | Cephalosporins 2nd Gen | 1 | 1 | 5 |
| Acute rheumatic fever | Penicillin V | Penicillins | 4 | 4 | 10 |
| **Purulent complications** | Amoxicillin-clavulanate | Penicillins, β-lactamase inhibitors | 1 | 1 | 5 |
| Purulent complications | Ceftibuten | Cephalosporins 3rd Gen | 1 | 1 | 5 |
| Purulent complications | Cephaloglycin | Cephalosporins 1st Gen | 1 | 1 | 10 |
| Purulent complications | Clarithromycin | Macrolide | 1 | 2 | 5 |
| Purulent complications | Penicillin V | Penicillins | 2 | 2 | 10 |
| Purulent complications | Penicillin V potassium | Penicillins | 1 | 2 | 7, 10 |
| **Poststreptococcal glomerulonephritis** | Amoxicillin | Penicillins | 2 | 2 | 10 |
| Poststreptococcal glomerulonephritis | Amoxicillin-clavulanate | Penicillins, β-lactamase inhibitors | 1 | 1 | 3 |
| Poststreptococcal glomerulonephritis | Amoxicillin-clavulanate, ceftibuten, cefuroxime axetil, loracarbef, clarithromycin or erythromycin | NA | 1 | 1 | 5 |
| Poststreptococcal glomerulonephritis | Azithromycin | Macrolide | 1 | 1 | 3 |
| Poststreptococcal glomerulonephritis | Cefcapene-pivoxil | Cephalosporins 3rd Gen | 1 | 2 | 5, 10 |
| Poststreptococcal glomerulonephritis | Cefuroxime axetil | Cephalosporins 2nd Gen | 1 | 1 | 5 |
| Poststreptococcal glomerulonephritis | Erythromycin estolate | Macrolide | 1 | 1 | 5 |
| Poststreptococcal glomerulonephritis | Loracarbef | Cephalosporins 2nd Gen | 1 | 1 | 5 |

| Outcome | Antibiotic | Antibiotic class | Studies | Arms | Durations |
|---|---|---|---|---|---|
| Poststreptococcal glomerulonephritis | Penicillin V | Penicillins | 5 | 5 | 10 |

**Supplement 8. Antibiotic classes represented across outcomes**

| Outcome | Antibiotic class | Antibiotics | Studies | Arms | Durations |
|---|---|---|---|---|---|
| **Clinical cure** | Cephalosporins 2nd Gen | Cefaclor; Cefprozil; Cefuroxime axetil; Loracarbef | 8 | 10 | 4, 5, 10 |
| Clinical cure | Cephalosporins 3rd Gen | Cefcapene-pivoxil; Cefdinir; Cefetamet pivoxil; Cefixime; Cefpodoxime proxetil; Ceftibuten | 7 | 10 | 5, 7, 10 |
| Clinical cure | Macrolide | Azithromycin; Clarithromycin; Erythromycin; Erythromycin estolate; Josamycin; Spiramycin | 19 | 26 | 3, 5, 7, 10 |
| Clinical cure | NA | Amoxicillin-clavulanate, ceftibuten, cefuroxime axetil, loracarbef, clarithromycin or erythromycin | 1 | 1 | 5 |
| Clinical cure | Penicillins | Amoxicillin; Penicillin V; Penicillin V potassium | 27 | 30 | 5, 6, 7, 10 |
| Clinical cure | Penicillins, β-lactamase inhibitors | Amoxicillin-clavulanate | 2 | 2 | 3, 5 |
| **Relapse** | Cephalosporins 2nd Gen | Cefaclor; Cefuroxime axetil | 2 | 2 | 4, 10 |
| Relapse | Cephalosporins 3rd Gen | Cefcapene-pivoxil; Cefixime; Ceftibuten | 3 | 4 | 5, 10 |
| Relapse | Macrolide | Azithromycin; Erythromycin; Erythromycin estolate | 11 | 15 | 3, 5, 10 |
| Relapse | NA | Amoxicillin-clavulanate, ceftibuten, cefuroxime axetil, loracarbef, clarithromycin or erythromycin | 1 | 1 | 5 |
| Relapse | Penicillins | Amoxicillin; Penicillin V | 12 | 13 | 6, 10 |
| **Adverse events** | Cephalosporins 2nd Gen | Cefaclor; Cefprozil; Cefuroxime axetil; Loracarbef | 8 | 10 | 4, 5, 10 |
| Adverse events | Cephalosporins 3rd Gen | Cefcapene-pivoxil; Cefdinir; Cefixime; Ceftibuten | 5 | 6 | 5, 10 |
| Adverse events | Macrolide | Azithromycin; Clarithromycin; Erythromycin; Erythromycin estolate; Josamycin; Spiramycin | 17 | 22 | 3, 5, 10 |
| Adverse events | NA | Amoxicillin-clavulanate, ceftibuten, cefuroxime axetil, loracarbef, clarithromycin or erythromycin | 1 | 1 | 5 |
| Adverse events | Penicillins | Amoxicillin; Penicillin V | 23 | 24 | 6, 7, 10 |
| Adverse events | Penicillins, β-lactamase inhibitors | Amoxicillin-clavulanate | 2 | 2 | 3, 5 |
| **Mortality** | Cephalosporins 3rd Gen | Ceftibuten | 1 | 1 | 5 |
| Mortality | Macrolide | Azithromycin; Clarithromycin | 2 | 2 | 3, 5 |
| Mortality | Penicillins | Penicillin V | 1 | 1 | 10 |
| **Serious adverse events** | Cephalosporins 2nd Gen | Cefaclor; Cefuroxime axetil | 4 | 4 | 4, 5, 10 |
| Serious adverse events | Cephalosporins 3rd Gen | Cefetamet pivoxil | 1 | 2 | 7, 10 |
| Serious adverse events | Macrolide | Azithromycin; Clarithromycin; Erythromycin; Spiramycin | 5 | 6 | 3, 5, 10 |
| Serious adverse events | NA | Amoxicillin-clavulanate, ceftibuten, cefuroxime axetil, loracarbef, clarithromycin or erythromycin | 1 | 1 | 5 |
| Serious adverse events | Penicillins | Amoxicillin; Penicillin V | 10 | 11 | 6, 7, 10 |
| Serious adverse events | Penicillins, β-lactamase inhibitors | Amoxicillin-clavulanate | 1 | 1 | 3 |
| **Acute rheumatic fever** | Cephalosporins 2nd Gen | Cefuroxime axetil; Loracarbef | 2 | 2 | 5 |
| Acute rheumatic fever | Cephalosporins 3rd Gen | Cefcapene-pivoxil | 1 | 2 | 5, 10 |
| Acute rheumatic fever | Macrolide | Erythromycin estolate | 1 | 1 | 5 |
| Acute rheumatic fever | NA | Amoxicillin-clavulanate, ceftibuten, cefuroxime axetil, loracarbef, clarithromycin or erythromycin | 1 | 1 | 5 |
| Acute rheumatic fever | Penicillins | Amoxicillin; Penicillin V | 5 | 5 | 10 |
| **Purulent complications** | Cephalosporins 1st Gen | Cephaloglycin | 1 | 1 | 10 |
| Purulent complications | Cephalosporins 3rd Gen | Ceftibuten | 1 | 1 | 5 |
| Purulent complications | Macrolide | Clarithromycin | 1 | 2 | 5 |
| Purulent complications | Penicillins | Penicillin V; Penicillin V potassium | 3 | 4 | 7, 10 |
| Purulent complications | Penicillins, β-lactamase inhibitors | Amoxicillin-clavulanate | 1 | 1 | 5 |
| **Poststreptococcal glomerulonephritis** | Cephalosporins 2nd Gen | Cefuroxime axetil; Loracarbef | 2 | 2 | 5 |
| Poststreptococcal glomerulonephritis | Cephalosporins 3rd Gen | Cefcapene-pivoxil | 1 | 2 | 5, 10 |
| Poststreptococcal glomerulonephritis | Macrolide | Azithromycin; Erythromycin estolate | 2 | 2 | 3, 5 |
| Poststreptococcal glomerulonephritis | NA | Amoxicillin-clavulanate, ceftibuten, cefuroxime axetil, loracarbef, clarithromycin or erythromycin | 1 | 1 | 5 |

| Outcome | Antibiotic class | Antibiotics | Studies | Arms | Durations |
|---|---|---|---|---|---|
| Poststreptococcal glomerulonephritis | Penicillins | Amoxicillin; Penicillin V | 7 | 7 | 10 |
| Poststreptococcal glomerulonephritis | Penicillins, β-lactamase inhibitors | Amoxicillin-clavulanate | 1 | 1 | 3 |

**Supplement 9. Risk of bias assessment for clinical relapse**

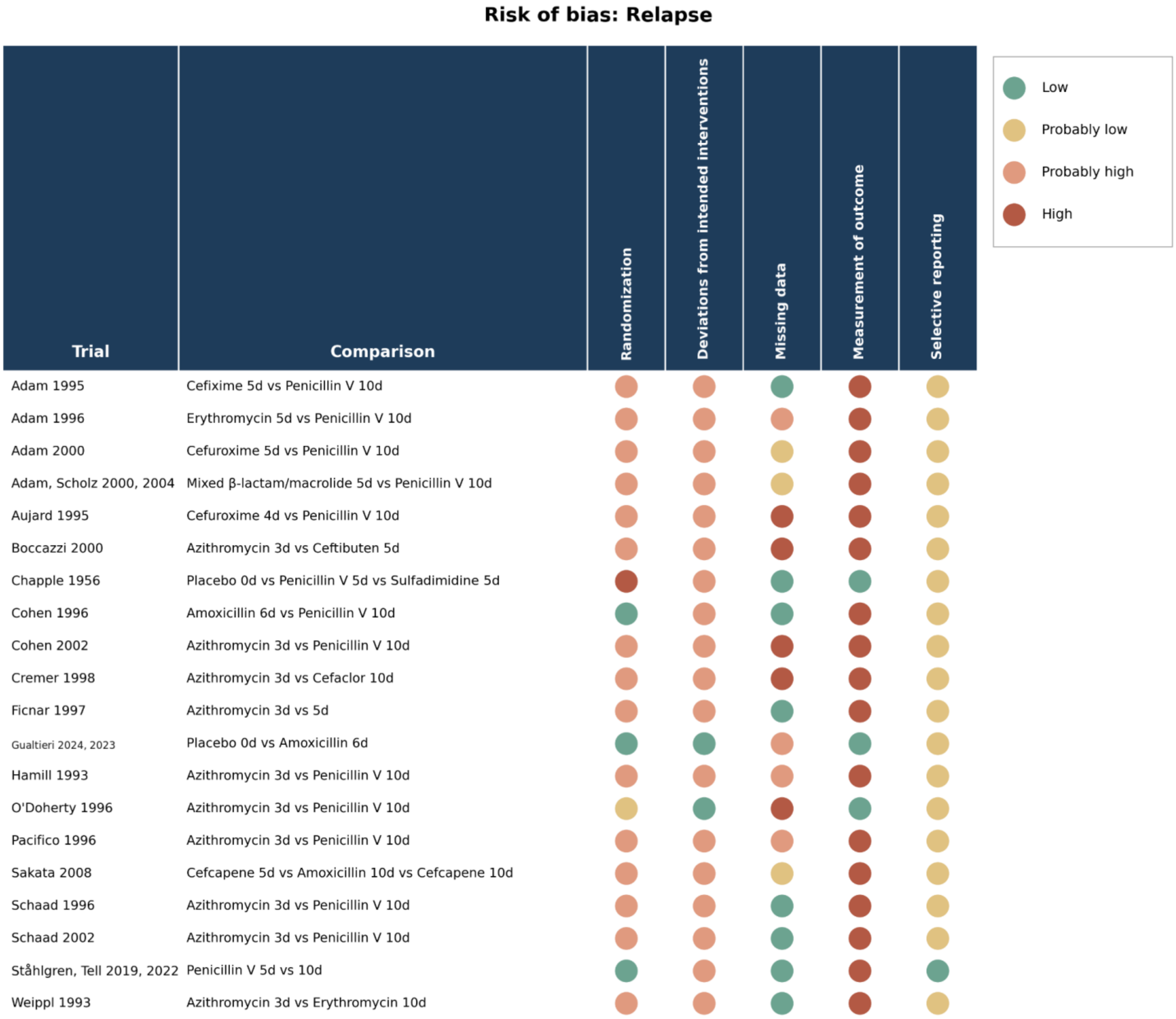


Domain-level judgments are shown for each trial reporting the outcome.

#### Supplement 10. Risk of bias assessment for adverse events

**Risk of bias: Adverse events**

| Trial | Comparison | Randomization | Deviations from intended interventions | Missing data | Measurement of outcome | Selective reporting |
|---|---|---|---|---|---|---|
| Adam 1995 | Cefixime 5d vs Penicillin V 10d | Probably high | Probably high | Low | High | Probably low |
| Adam 1996 | Erythromycin 5d vs Penicillin V 10d | Probably high | Probably high | Probably low | High | Probably low |
| Adam 2001_1 | Loracarbef 5d vs Penicillin V 10d | Probably high | Probably high | Low | High | Probably low |
| Adam 2001_2 | Ceftibuten 5d vs Penicillin V 10d | Probably high | Probably high | Low | High | Probably low |
| Adam, Scholz 2000, 2004 | Mixed β-lactam/macrolide 5d vs Penicillin V 10d | Probably high | Probably high | Low | High | Probably low |
| Aujard 1995 | Cefuroxime 4d vs Penicillin V 10d | Probably high | Probably high | High | High | Probably low |
| Boccazzi 2000 | Azithromycin 3d vs Ceftibuten 5d | Probably high | Probably high | Probably low | High | Probably low |
| Chapple 1956 | Placebo 0d vs Penicillin V 5d vs Sulfadimidine 5d | High | Probably high | Low | Low | Probably low |
| Cohen 1996 | Amoxicillin 6d vs Penicillin V 10d | Low | Probably high | Low | High | Probably low |
| Cohen 2002 | Azithromycin 3d vs Penicillin V 10d | Probably high | Probably high | Low | High | Probably low |
| Cremer 1998 | Azithromycin 3d vs Cefaclor 10d | Probably high | Probably high | Probably high | High | Probably low |
| Esposito 2001 | Cefaclor 5d vs 10d | Probably high | Probably high | Probably low | High | Probably low |
| Esposito 2002 | Cefaclor 5d vs Amoxicillin 10d | Probably low | Probably high | Low | High | Probably low |
| Gendrel 1997 | Spiramycin 5d vs Penicillin V 7d | Probably high | Probably high | Probably low | High | Probably low |
| Hamill 1993 | Azithromycin 3d vs Penicillin V 10d | Probably high | Probably high | Probably low | High | Probably low |
| Kafetzis 2004 | Cefprozil 5d vs Clarithromycin 10d vs Penicillin V 10d | Probably high | Probably high | Probably low | High | Probably low |
| Kuroki 2013 | Amoxicillin-clavulanate 3d vs Amoxicillin 10d | Probably high | Probably high | High | High | Probably low |
| Li 2019 | Azithromycin 3d vs Cefaclor 5d vs Amoxicillin 10d | Probably high | Probably high | Low | High | Probably low |
| McCarty 2000 | Clarithromycin 5d vs Penicillin V 10d | Probably low | Probably high | Low | High | Probably low |
| Mehra 1998 | Cefuroxime 5d vs 10d | Probably high | Probably high | High | High | Probably low |
| O'Doherty 1996 | Azithromycin 3d vs Penicillin V 10d | Probably low | Low | Probably high | Low | Probably low |
| Pacifico 1996 | Azithromycin 3d vs Penicillin V 10d | Probably high | Probably high | Probably high | High | Probably low |
| Portier 2001 | Josamycin 5d vs Penicillin V 10d | Probably high | Probably high | Probably high | High | Probably low |
| Ramet 1992 | Cefetamet 7d vs Cefetamet 10d vs Penicillin V 10d | Probably high | Probably high | Probably low | High | Probably low |
| Sakata 2008 | Cefcapene 5d vs Amoxicillin 10d vs Cefcapene 10d | Probably high | Probably high | Probably low | High | Probably low |
| Schaad 1996 | Azithromycin 3d vs Penicillin V 10d | Probably high | Probably high | Probably high | High | Probably low |
| Schaad 2002 | Azithromycin 3d vs Penicillin V 10d | Probably high | Probably high | Low | High | Probably low |
| Syrogiannopoulos 2004 | Amoxicillin-clavulanate 5d vs Clarithromycin 5d vs Penicillin V 10d | Probably high | Probably high | High | High | Probably low |
| Tack 1997 | Cefdinir 5d vs Penicillin V 10d | Probably high | Probably high | Low | High | Probably low |
| Venuta 1998 | Azithromycin 3d vs Clarithromycin 10d | Probably low | Probably high | High | High | Probably low |
| Weippl 1993 | Azithromycin 3d vs Erythromycin 10d | Probably high | Probably high | Low | High | Probably low |

Legend: Low; Probably low; Probably high; High

Domain-level judgments are shown for each trial reporting the outcome.

**Supplement 11. Risk of bias assessment for mortality**

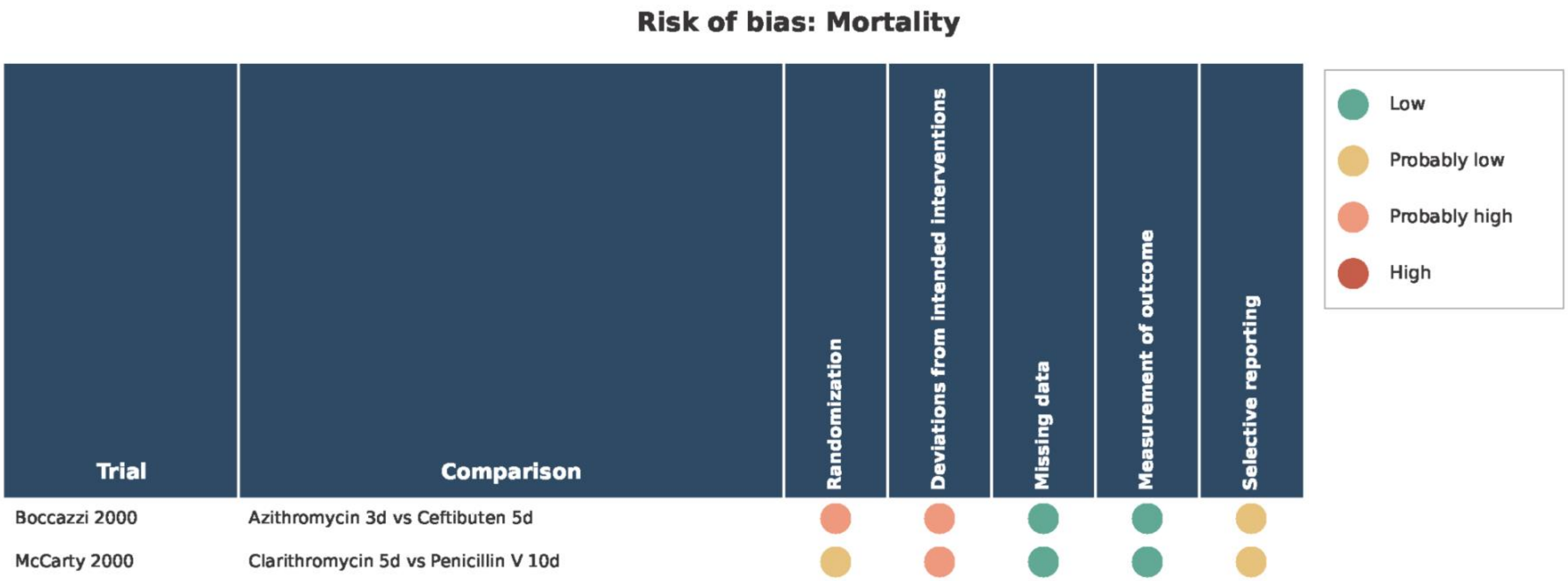


Domain-level judgments are shown for each trial reporting the outcome.

**Supplement 12. Risk of bias assessment for serious adverse events**

#### Risk of bias: Serious adverse events

| Trial | Comparison | Randomization | Deviations from intended interventions | Missing data | Measurement of outcome | Selective reporting |
|---|---|---|---|---|---|---|
| Adam 2000 | Cefuroxime 5d vs Penicillin V 10d | Probably high | Probably high | Probably low | High | Probably low |
| Adam, Scholz 2000, 2004 | Mixed β-lactam/macrolide 5d vs Penicillin V 10d | Probably high | Probably high | Low | High | Probably low |
| Aujard 1995 | Cefuroxime 4d vs Penicillin V 10d | Probably high | Probably high | High | High | Probably low |
| Chapple 1956 | Placebo 0d vs Penicillin V 5d vs Sulfadimidine 5d | High | Probably high | Low | Low | Probably low |
| Cohen 1996 | Amoxicillin 6d vs Penicillin V 10d | Low | Probably high | Low | High | Probably low |
| Cremer 1998 | Azithromycin 3d vs Cefaclor 10d | Probably high | Probably high | Probably high | High | Probably low |
| Esposito 2002 | Cefaclor 5d vs Amoxicillin 10d | Probably low | Probably high | Low | High | Probably low |
| Gendrel 1997 | Spiramycin 5d vs Penicillin V 7d | Probably high | Probably high | Probably low | High | Probably low |
| Kuroki 2013 | Amoxicillin–clavulanate 3d vs Amoxicillin 10d | Probably high | Probably high | High | High | Probably low |
| McCarty 2000 | Clarithromycin 5d vs Penicillin V 10d | Probably low | Probably high | Low | High | Probably low |
| Pacifico 1996 | Azithromycin 3d vs Penicillin V 10d | Probably high | Probably high | Probably high | High | Probably low |
| Ramet 1992 | Cefetamet 7d vs Cefetamet 10d vs Penicillin V 10d | Probably high | Probably high | Probably low | High | Probably low |
| Randolph 1985 | Placebo 0d vs Cefadroxil 10d vs Penicillin V 10d | Low | Low | Low | Low | Probably low |
| Ståhlgren, Tell 2019, 2022 | Penicillin V 5d vs 10d | Low | Probably high | Low | High | Probably low |
| Weippl 1993 | Azithromycin 3d vs Erythromycin 10d | Probably high | Probably high | Low | High | Probably low |

Legend: Low; Probably low; Probably high; High

Domain-level judgments are shown for each trial reporting the outcome.

**Supplement 13. Risk of bias assessment for acute rheumatic fever**

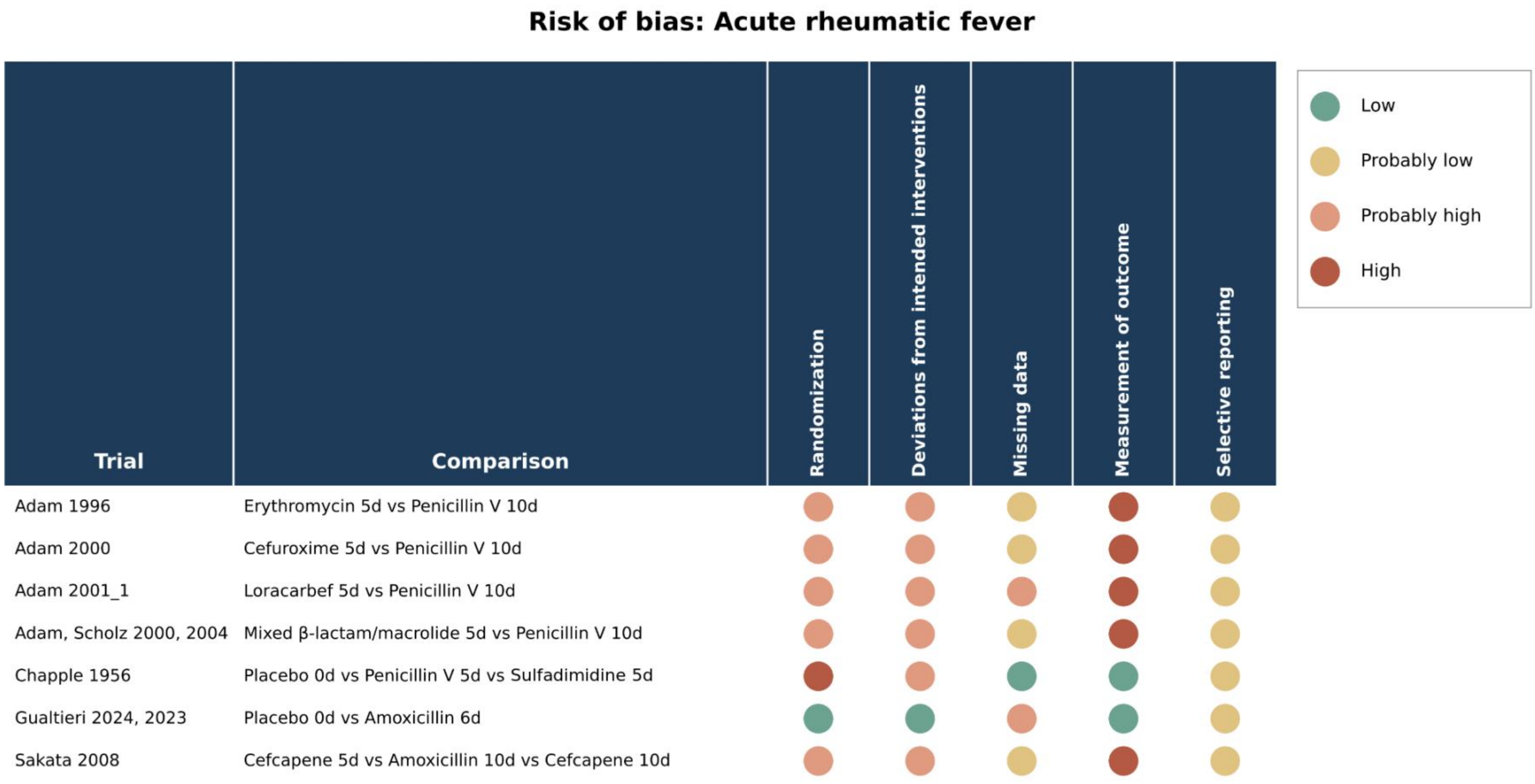


Domain-level judgments are shown for each trial reporting the outcome.

**Supplement 14. Risk of bias assessment for purulent complications**

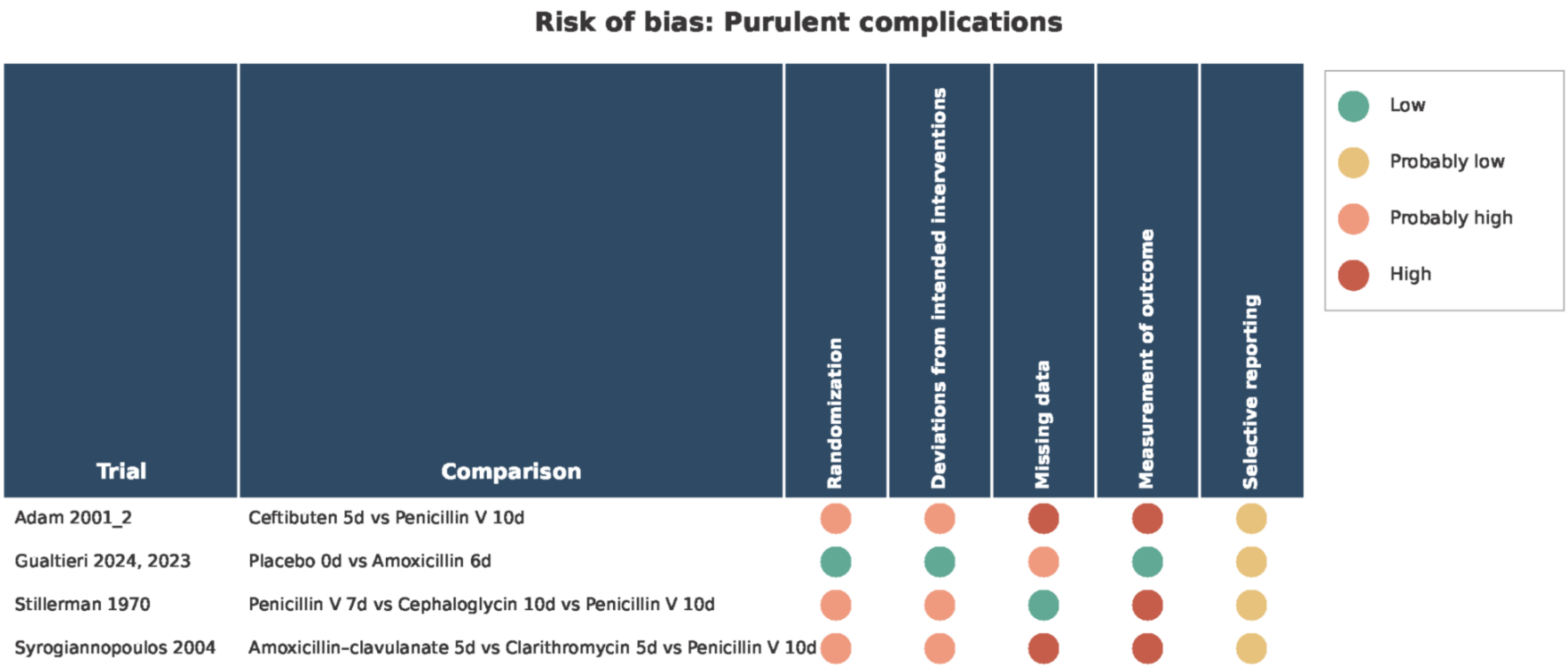


Domain-level judgments are shown for each trial reporting the outcome.

**Supplement 15. Risk of bias assessment for poststreptococcal glomerulonephritis**

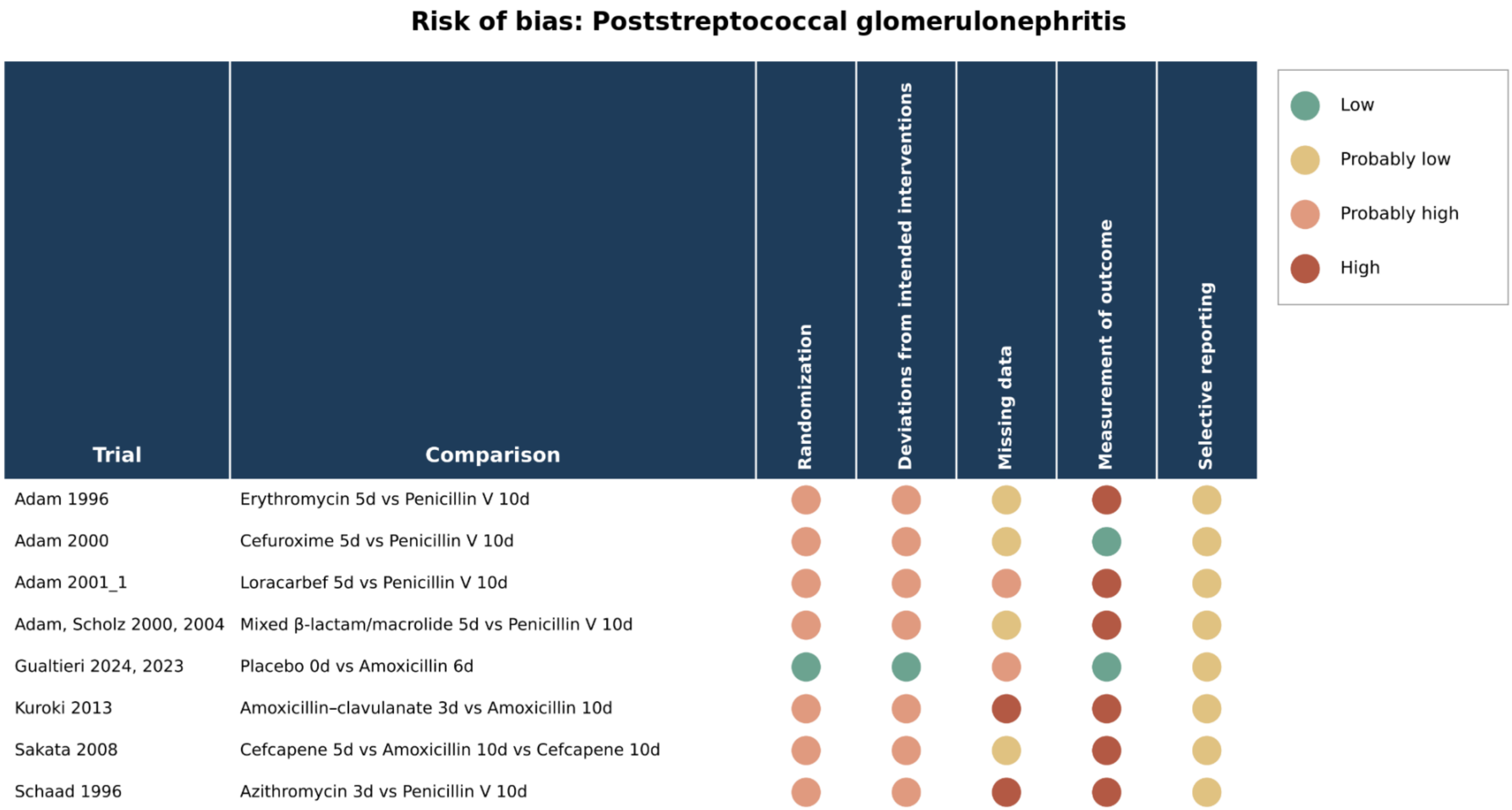


Domain-level judgments are shown for each trial reporting the outcome.

**Supplement 16. Dose-response meta-analysis results: study and model characteristics**

| Analysis | Outcome | Studies | Arms | Participants | Events | Studies per duration | Baseline risk per 1,000 |
|---|---|---|---|---|---|---|---|
| **Primary** | Clinical cure | 35 | 71 | 16,431 | 14,518 | 3d: 13; 4d: 1; 5d: 20; 6d: 1; 7d: 4; 10d: 32 | 936 |
| Primary | Relapse | 16 | 32 | 7,360 | 1,209 | 3d: 10; 4d: 1; 5d: 6; 6d: 1; 10d: 14 | 49.6 |
| Primary | Adverse events | 29 | 59 | 13,996 | 1,188 | 3d: 12; 4d: 1; 5d: 17; 6d: 1; 7d: 1; 10d: 27 | 74 |
| Primary | Mortality | 2 | 4 | 776 | 0 | 3d: 1; 5d: 2; 10d: 1 | 0 |
| Primary | Serious adverse events | 12 | 24 | 6,558 | 10 | 3d: 4; 4d: 1; 5d: 5; 6d: 1; 7d: 2; 10d: 11 | 0 |
| Primary | Acute rheumatic fever | 5 | 10 | 8,818 | 4 | 5d: 5; 10d: 5 | 0 |
| Primary | Purulent complications | 3 | 6 | 1,741 | 20 | 5d: 2; 7d: 1; 10d: 3 | 0 |
| Primary | Poststreptococcal glomerulonephritis | 7 | 14 | 9,096 | 4 | 3d: 2; 5d: 5; 10d: 7 | 0 |
| **Same antibiotic and same dose** | Clinical cure | 5 | 10 | 869 | 852 | 5d: 3; 7d: 2; 10d: 5 | |
| Same antibiotic and same dose | Adverse events | 3 | 6 | 948 | 49 | 5d: 3; 10d: 3 | |
| **Same antibiotic** | Clinical cure | 8 | 16 | 1,374 | 1,266 | 3d: 1; 5d: 6; 7d: 2; 10d: 7 | |
| Same antibiotic | Relapse | 2 | 4 | 235 | 4 | 3d: 1; 5d: 2; 10d: 1 | |
| Same antibiotic | Adverse events | 3 | 6 | 948 | 49 | 5d: 3; 10d: 3 | |
| **Same class** | Clinical cure | 9 | 18 | 1,794 | 1,656 | 3d: 3; 5d: 4; 6d: 1; 7d: 2; 10d: 8 | |
| Same class | Relapse | 3 | 6 | 422 | 16 | 3d: 2; 5d: 1; 6d: 1; 10d: 2 | |
| Same class | Adverse events | 5 | 10 | 1,337 | 72 | 3d: 2; 5d: 2; 6d: 1; 10d: 5 | |
| Same class | Serious adverse events | 2 | 4 | 411 | 1 | 3d: 1; 6d: 1; 10d: 2 | |
| **Penicillins** | Clinical cure | 3 | 6 | 851 | 763 | 5d: 1; 6d: 1; 7d: 1; 10d: 3 | |
| **Excl. azithromycin** | Clinical cure | 23 | 46 | 13,863 | 12,432 | 3d: 1; 4d: 1; 5d: 17; 6d: 1; 7d: 4; 10d: 22 | |
| Excl. azithromycin | Relapse | 6 | 12 | 5,491 | 1,076 | 4d: 1; 5d: 4; 6d: 1; 10d: 6 | |
| Excl. azithromycin | Adverse events | 18 | 36 | 11,252 | 919 | 3d: 1; 4d: 1; 5d: 15; 6d: 1; 7d: 1; 10d: 17 | |
| Excl. azithromycin | Serious adverse events | 9 | 18 | 6,160 | 9 | 3d: 1; 4d: 1; 5d: 5; 6d: 1; 7d: 2; 10d: 8 | |
| Excl. azithromycin | Acute rheumatic fever | 5 | 10 | 8,818 | 4 | 5d: 5; 10d: 5 | |
| Excl. azithromycin | Purulent complications | 3 | 6 | 1,741 | 20 | 5d: 2; 7d: 1; 10d: 3 | |
| Excl. azithromycin | Poststreptococcal glomerulonephritis | 6 | 12 | 8,904 | 3 | 3d: 1; 5d: 5; 10d: 6 | |
| **Excl. macrolides** | Clinical cure | 15 | 30 | 7,383 | 6,691 | 3d: 1; 4d: 1; 5d: 10; 6d: 1; 7d: 2; 10d: 15 | |
| Excl. macrolides | Relapse | 4 | 8 | 812 | 36 | 4d: 1; 5d: 2; 6d: 1; 10d: 4 | |
| Excl. macrolides | Adverse events | 11 | 22 | 6,915 | 419 | 3d: 1; 4d: 1; 5d: 8; 6d: 1; 10d: 11 | |
| Excl. macrolides | Serious adverse events | 6 | 12 | 3,265 | 0 | 3d: 1; 4d: 1; 5d: 2; 6d: 1; 7d: 1; 10d: 6 | |
| Excl. macrolides | Acute rheumatic fever | 3 | 6 | 4,109 | 1 | 5d: 3; 10d: 3 | |
| Excl. macrolides | Purulent complications | 2 | 4 | 1,115 | 20 | 5d: 1; 7d: 1; 10d: 2 | |

| Analysis | Outcome | Studies | Arms | Participants | Events | Studies per duration | Baseline risk per 1,000 |
|---|---|---|---|---|---|---|---|
| Excl. macrolides | Poststreptococcal glomerulonephritis | 4 | 8 | 4,195 | 1 | 3d: 1; 5d: 3; 10d: 4 | |
| **Incl. placebo/no-treatment as 0 days** | Clinical cure | 39 | 79 | 17,213 | 15,126 | 0d: 4; 2d: 1; 3d: 13; 4d: 1; 5d: 21; 6d: 2; 7d: 4; 10d: 33 | |
| Incl. placebo/no-treatment as 0 days | Relapse | 18 | 36 | 7,708 | 1,231 | 0d: 2; 3d: 10; 4d: 1; 5d: 7; 6d: 2; 10d: 14 | |
| Incl. placebo/no-treatment as 0 days | Adverse events | 30 | 61 | 14,279 | 1,194 | 0d: 1; 3d: 12; 4d: 1; 5d: 18; 6d: 1; 7d: 1; 10d: 27 | |
| Incl. placebo/no-treatment as 0 days | Mortality | 2 | 4 | 776 | 0 | 3d: 1; 5d: 2; 10d: 1 | |
| Incl. placebo/no-treatment as 0 days | Serious adverse events | 14 | 28 | 7,035 | 15 | 0d: 2; 3d: 4; 4d: 1; 5d: 6; 6d: 1; 7d: 2; 10d: 12 | |
| Incl. placebo/no-treatment as 0 days | Acute rheumatic fever | 7 | 14 | 9,189 | 4 | 0d: 2; 5d: 6; 6d: 1; 10d: 5 | |
| Incl. placebo/no-treatment as 0 days | Purulent complications | 4 | 8 | 1,806 | 24 | 0d: 1; 5d: 2; 6d: 1; 7d: 1; 10d: 3 | |
| Incl. placebo/no-treatment as 0 days | Poststreptococcal glomerulonephritis | 8 | 16 | 9,184 | 4 | 0d: 1; 3d: 2; 5d: 5; 6d: 1; 10d: 7 | |

AIC = Akaike information criterion; CI = confidence interval; RCS = restricted cubic spline; RD = risk difference; RR = risk ratio. Not estimable indicates that the model did not support the corresponding estimate.

**Supplement 17. Dose-response meta-analysis results: linear models**

| Analysis | Outcome | RR per day (95% CI) | RR, 5 vs 10 days (95% CI) | RD per 1,000, 5 vs 10 days (95% CI) | RR, 3 vs 10 days (95% CI) | RD per 1,000, 3 vs 10 days (95% CI) | Tau-squared | Q test P value | AIC |
|---|---|---|---|---|---|---|---|---|---|
| **Primary** | Clinical cure | 1.003 (1.000 to 1.005) | 0.987 (0.975 to 1.000) | -11.8 (-23.6 to 0.2) | 0.982 (0.965 to 1.000) | -16.5 (-32.9 to 0.2) | 0.00002 | Not estimable | -209.19 |
| Primary | Relapse | 0.974 (0.921 to 1.031) | 1.139 (0.859 to 1.512) | 6.9 (-7.0 to 25.4) | 1.201 (0.808 to 1.784) | 9.9 (-9.5 to 38.9) | 0.003 | Not estimable | -7.72 |
| Primary | Adverse events | 0.962 (0.925 to 0.999) | 1.217 (1.003 to 1.476) | 16.0 (0.2 to 35.2) | 1.316 (1.004 to 1.725) | 23.4 (0.3 to 53.6) | 0.005 | Not estimable | -35.1 |
| Primary | Mortality | 0.996 (0.482 to 2.062) | 1.018 (0.027 to 38.626) | 0.0 (-0.0 to 0.0) | 1.026 (0.006 to 166.581) | 0.0 (-0.0 to 0.0) | 0 | Not estimable | 5.99 |
| Primary | Serious adverse events | 1.027 (0.859 to 1.227) | 0.876 (0.360 to 2.136) | -0.0 (-0.0 to 0.0) | 0.831 (0.239 to 2.893) | -0.0 (-0.0 to 0.0) | 0 | Not estimable | 6.15 |
| Primary | Acute rheumatic fever | 0.827 (0.602 to 1.138) | 2.579 (0.525 to 12.677) | 0.0 (-0.0 to 0.0) | 3.767 (0.405 to 35.011) | 0.0 (-0.0 to 0.0) | 0 | Not estimable | 5.45 |
| Primary | Purulent complications | 1.079 (0.909 to 1.281) | 0.683 (0.290 to 1.609) | -0.0 (-0.0 to 0.0) | 0.587 (0.177 to 1.946) | -0.0 (-0.0 to 0.0) | 0 | Not estimable | 5.93 |
| Primary | Poststreptococcal glomerulonephritis | 1.021 (0.805 to 1.293) | 0.903 (0.276 to 2.950) | -0.0 (-0.0 to 0.0) | 0.867 (0.165 to 4.548) | -0.0 (-0.0 to 0.0) | 0 | Not estimable | 4.66 |
| **Same antibiotic and same dose** | Clinical cure | 1.002 (0.997 to 1.007) | 0.989 (0.966 to 1.013) | -10.0 (-31.6 to 12.2) | 0.985 (0.953 to 1.018) | -13.9 (-44.0 to 17.1) | 0 | Not estimable | -25.17 |
| Same antibiotic and same dose | Adverse events | 1.045 (0.936 to 1.166) | 0.804 (0.464 to 1.392) | -14.5 (-39.6 to 29.0) | 0.737 (0.342 to 1.588) | -19.5 (-48.7 to 43.5) | 0 | Not estimable | 0.32 |
| **Same antibiotic** | Clinical cure | 1.003 (0.999 to 1.008) | 0.984 (0.962 to 1.007) | -14.9 (-35.5 to 6.2) | 0.978 (0.947 to 1.009) | -20.8 (-49.4 to 8.6) | 0 | Not estimable | -41.19 |
| Same antibiotic | Relapse | 1.240 (0.801 to 1.920) | 0.341 (0.038 to 3.024) | -32.7 (-47.7 to 100.3) | 0.221 (0.010 to 4.707) | -38.6 (-49.1 to 183.8) | 0 | Not estimable | 5.91 |
| Same antibiotic | Adverse events | 1.045 (0.936 to 1.166) | 0.804 (0.464 to 1.392) | -14.5 (-39.6 to 29.0) | 0.737 (0.342 to 1.588) | -19.5 (-48.7 to 43.5) | 0 | Not estimable | 0.32 |
| **Same class** | Clinical cure | 1.003 (0.998 to 1.007) | 0.986 (0.964 to 1.008) | -13.1 (-33.6 to 7.8) | 0.980 (0.950 to 1.012) | -18.3 (-46.7 to 11.0) | 0 | Not estimable | -41.48 |
| Same class | Relapse | 0.914 (0.733 to 1.140) | 1.570 (0.520 to 4.737) | 28.3 (-23.8 to 185.3) | 1.880 (0.401 to 8.825) | 43.6 (-29.7 to 388.0) | 0 | Not estimable | 5.13 |
| Same class | Adverse events | 1.038 (0.955 to 1.128) | 0.829 (0.548 to 1.256) | -12.6 (-33.5 to 18.9) | 0.769 (0.430 to 1.375) | -17.1 (-42.1 to 27.8) | 0 | Not estimable | -3.79 |
| Same class | Serious adverse events | 1.084 (0.703 to 1.669) | 0.669 (0.077 to 5.804) | -0.0 (-0.0 to 0.0) | 0.570 (0.028 to 11.728) | -0.0 (-0.0 to 0.0) | 0 | Not estimable | 4.7 |
| **Penicillins** | Clinical cure | 1.009 (0.993 to 1.025) | 0.956 (0.884 to 1.033) | -41.3 (-108.3 to 31.1) | 0.939 (0.842 to 1.047) | -57.3 (-148.1 to 43.9) | 0.00008 | Not estimable | -6.4 |
| **Excl. azithromycin** | Clinical cure | 1.003 (1.000 to 1.006) | 0.985 (0.970 to 1.000) | -14.4 (-28.2 to -0.4) | 0.979 (0.958 to 0.999) | -20.1 (-39.3 to -0.5) | 0.00002 | Not estimable | -142.6 |

| Analysis | Outcome | RR per day (95% CI) | RR, 5 vs 10 days (95% CI) | RD per 1,000, 5 vs 10 days (95% CI) | RR, 3 vs 10 days (95% CI) | RD per 1,000, 3 vs 10 days (95% CI) | Tau-squared | Q test P value | AIC |
|---|---|---|---|---|---|---|---|---|---|
| Excl. azithromycin | Relapse | 1.023 (1.001 to 1.046) | 0.891 (0.798 to 0.995) | -5.4 (-10.0 to -0.2) | 0.851 (0.729 to 0.993) | -7.4 (-13.4 to -0.3) | 0 | Not estimable | -1.83 |
| Excl. azithromycin | Adverse events | 0.970 (0.924 to 1.019) | 1.162 (0.912 to 1.481) | 12.0 (-6.5 to 35.6) | 1.234 (0.879 to 1.733) | 17.4 (-8.9 to 54.3) | 0.005 | Not estimable | -19.09 |
| Excl. azithromycin | Serious adverse events | 1.018 (0.818 to 1.268) | 0.914 (0.305 to 2.737) | -0.0 (-0.0 to 0.0) | 0.881 (0.190 to 4.095) | -0.0 (-0.0 to 0.0) | 0 | Not estimable | 7.94 |
| Excl. azithromycin | Acute rheumatic fever | 0.827 (0.602 to 1.138) | 2.579 (0.525 to 12.677) | 0.0 (-0.0 to 0.0) | 3.767 (0.405 to 35.011) | 0.0 (-0.0 to 0.0) | 0 | Not estimable | 5.45 |
| Excl. azithromycin | Purulent complications | 1.079 (0.909 to 1.281) | 0.683 (0.290 to 1.609) | -0.0 (-0.0 to 0.0) | 0.587 (0.177 to 1.946) | -0.0 (-0.0 to 0.0) | 0 | Not estimable | 5.93 |
| Excl. azithromycin | Poststreptococcal glomerulonephritis | 0.991 (0.756 to 1.301) | 1.044 (0.268 to 4.061) | 0.0 (-0.0 to 0.0) | 1.062 (0.159 to 7.112) | 0.0 (-0.0 to 0.0) | 0 | Not estimable | 5.17 |
| **Excl. macrolides** | Clinical cure | 1.002 (0.999 to 1.006) | 0.989 (0.973 to 1.005) | -10.4 (-25.5 to 4.9) | 0.984 (0.962 to 1.007) | -14.5 (-35.5 to 6.9) | 0.00001 | Not estimable | -91.61 |
| Excl. macrolides | Relapse | 1.013 (0.878 to 1.168) | 0.937 (0.459 to 1.913) | -3.1 (-26.8 to 45.3) | 0.913 (0.336 to 2.480) | -4.3 (-32.9 to 73.4) | 0 | Not estimable | 2.29 |
| Excl. macrolides | Adverse events | 0.995 (0.930 to 1.065) | 1.025 (0.731 to 1.437) | 1.9 (-19.9 to 32.4) | 1.035 (0.645 to 1.662) | 2.6 (-26.3 to 49.0) | 0.006 | Not estimable | -7.85 |
| Excl. macrolides | Serious adverse events | 0.963 (0.707 to 1.312) | 1.208 (0.258 to 5.663) | 0.0 (-0.0 to 0.0) | 1.303 (0.150 to 11.331) | 0.0 (-0.0 to 0.0) | 0 | Not estimable | 6.63 |
| Excl. macrolides | Acute rheumatic fever | 0.787 (0.513 to 1.209) | 3.305 (0.387 to 28.227) | 0.0 (-0.0 to 0.0) | 5.331 (0.265 to 107.379) | 0.0 (-0.0 to 0.0) | 0 | Not estimable | 5.09 |
| Excl. macrolides | Purulent complications | 1.072 (0.899 to 1.277) | 0.708 (0.294 to 1.702) | -0.0 (-0.0 to 0.0) | 0.617 (0.181 to 2.106) | -0.0 (-0.0 to 0.0) | 0 | Not estimable | 5.75 |
| Excl. macrolides | Poststreptococcal glomerulonephritis | 0.931 (0.663 to 1.308) | 1.429 (0.261 to 7.817) | 0.0 (-0.0 to 0.0) | 1.649 (0.153 to 17.793) | 0.0 (-0.0 to 0.0) | 0 | Not estimable | 4.95 |
| **Incl. placebo/no-treatment as 0 days** | Clinical cure | 1.003 (1.001 to 1.006) | 0.983 (0.969 to 0.997) | -16.2 (-29.1 to -3.1) | 0.976 (0.957 to 0.995) | -22.6 (-40.5 to -4.3) | 0.00003 | Not estimable | -193.53 |
| Incl. placebo/no-treatment as 0 days | Relapse | 0.980 (0.930 to 1.032) | 1.107 (0.853 to 1.435) | 5.3 (-7.3 to 21.6) | 1.152 (0.801 to 1.658) | 7.6 (-9.9 to 32.6) | 0.003 | Not estimable | -10.91 |
| Incl. placebo/no-treatment as 0 days | Adverse events | 0.963 (0.927 to 1.001) | 1.207 (0.995 to 1.465) | 15.3 (-0.4 to 34.4) | 1.302 (0.993 to 1.706) | 22.3 (-0.5 to 52.2) | 0.005 | Not estimable | -34.17 |

| Analysis | Outcome | RR per day (95% CI) | RR, 5 vs 10 days (95% CI) | RD per 1,000, 5 vs 10 days (95% CI) | RR, 3 vs 10 days (95% CI) | RD per 1,000, 3 vs 10 days (95% CI) | Tau-squared | Q test P value | AIC |
|---|---|---|---|---|---|---|---|---|---|
| Incl. placebo/no-treatment as 0 days | Mortality | 0.996 (0.482 to 2.062) | 1.018 (0.027 to 38.626) | 0.0 (-0.0 to 0.0) | 1.026 (0.006 to 166.581) | 0.0 (-0.0 to 0.0) | 0 | Not estimable | 5.99 |
| Incl. placebo/no-treatment as 0 days | Serious adverse events | 1.031 (0.882 to 1.205) | 0.860 (0.394 to 1.877) | -0.0 (-0.0 to 0.0) | 0.810 (0.271 to 2.415) | -0.0 (-0.0 to 0.0) | 0 | Not estimable | 5.83 |
| Incl. placebo/no-treatment as 0 days | Acute rheumatic fever | 0.863 (0.660 to 1.129) | 2.090 (0.546 to 8.006) | 0.0 (-0.0 to 0.0) | 2.807 (0.428 to 18.399) | 0.0 (-0.0 to 0.0) | 0 | Not estimable | 5.73 |
| Incl. placebo/no-treatment as 0 days | Purulent complications | 1.026 (0.863 to 1.219) | 0.882 (0.371 to 2.092) | -0.0 (-0.0 to 0.0) | 0.838 (0.250 to 2.811) | -0.0 (-0.0 to 0.0) | 0.003 | Not estimable | 6 |
| Incl. placebo/no-treatment as 0 days | Poststreptococcal glomerulonephritis | 1.020 (0.817 to 1.274) | 0.906 (0.298 to 2.755) | -0.0 (-0.0 to 0.0) | 0.871 (0.183 to 4.133) | -0.0 (-0.0 to 0.0) | 0 | Not estimable | 4.42 |

AIC = Akaike information criterion; CI = confidence interval; RCS = restricted cubic spline; RD = risk difference; RR = risk ratio. Not estimable indicates that the model did not support the corresponding estimate.

**Supplement 18. Dose-response meta-analysis results: restricted cubic spline models**

| Analysis | Outcome | Knots | RR, 5 vs 10 days (95% CI) | RD per 1,000, 5 vs 10 days (95% CI) | RR, 3 vs 10 days (95% CI) | RD per 1,000, 3 vs 10 days (95% CI) | AIC |
|---|---|---|---|---|---|---|---|
| **Primary** | Clinical cure | 3 | 0.985 (0.971 to 0.998) | -14.4 (-27.1 to -1.6) | 0.995 (0.967 to 1.023) | -4.9 (-30.5 to 21.3) | -90.24 |
| Primary | Relapse | 3 | 0.902 (0.772 to 1.054) | -4.9 (-11.3 to 2.7) | 1.382 (0.808 to 2.363) | 18.9 (-9.5 to 67.6) | 50.28 |
| Primary | Adverse events | 3 | 1.133 (0.914 to 1.404) | 9.8 (-6.4 to 29.9) | 1.471 (0.952 to 2.272) | 34.8 (-3.6 to 94.1) | 73.13 |
| Primary | Mortality | Not estimable | Not estimable | Not estimable | Not estimable | Not estimable | Not estimable |
| Primary | Serious adverse events | 3 | 1.117 (0.374 to 3.333) | 0.0 (-0.0 to 0.0) | 0.586 (0.125 to 2.752) | -0.0 (-0.0 to 0.0) | 48.79 |
| Primary | Acute rheumatic fever | Not estimable | Not estimable | Not estimable | Not estimable | Not estimable | Not estimable |
| Primary | Purulent complications | Not estimable | Not estimable | Not estimable | Not estimable | Not estimable | Not estimable |
| Primary | Poststreptococcal glomerulonephritis | Not estimable | Not estimable | Not estimable | Not estimable | Not estimable | Not estimable |
| **Same antibiotic and same dose** | Clinical cure | Not estimable | Not estimable | Not estimable | Not estimable | Not estimable | Not estimable |
| Same antibiotic and same dose | Adverse events | Not estimable | Not estimable | Not estimable | Not estimable | Not estimable | Not estimable |
| **Same antibiotic** | Clinical cure | 3 | 0.984 (0.962 to 1.007) | -14.7 (-35.6 to 6.6) | 0.982 (0.909 to 1.061) | -16.8 (-85.5 to 57.3) | -7.88 |
| Same antibiotic | Relapse | Not estimable | Not estimable | Not estimable | Not estimable | Not estimable | Not estimable |
| Same antibiotic | Adverse events | Not estimable | Not estimable | Not estimable | Not estimable | Not estimable | Not estimable |
| **Same class** | Clinical cure | 3 | 0.984 (0.962 to 1.006) | -15.1 (-35.8 to 6.0) | 1.008 (0.960 to 1.059) | 7.9 (-37.0 to 55.1) | -5.31 |
| Same class | Relapse | 3 | 1.568 (0.511 to 4.816) | 28.2 (-24.3 to 189.2) | 1.861 (0.110 to 31.606) | 42.7 (-44.2 to 1517.6) | 18.13 |
| Same class | Adverse events | 3 | 0.741 (0.455 to 1.205) | -19.2 (-40.3 to 15.1) | 1.015 (0.433 to 2.377) | 1.1 (-41.9 to 101.9) | 19.04 |
| Same class | Serious adverse events | Not estimable | Not estimable | Not estimable | Not estimable | Not estimable | Not estimable |
| **Penicillins** | Clinical cure | 3 | 0.982 (0.856 to 1.127) | -16.6 (-134.5 to 118.7) | 1.012 (0.739 to 1.385) | 11.3 (-244.0 to 360.7) | 11.46 |
| **Excl. azithromycin** | Clinical cure | 3 | 0.985 (0.970 to 1.000) | -14.3 (-28.2 to -0.3) | 0.990 (0.947 to 1.035) | -9.3 (-49.6 to 32.9) | -60.84 |
| Excl. azithromycin | Relapse | 3 | 0.889 (0.796 to 0.993) | -5.5 (-10.1 to -0.4) | 0.427 (0.056 to 3.257) | -28.4 (-46.8 to 111.9) | 22.1 |
| Excl. azithromycin | Adverse events | 3 | 1.131 (0.898 to 1.426) | 9.7 (-7.6 to 31.5) | 1.801 (0.606 to 5.350) | 59.2 (-29.2 to 321.9) | 42.39 |
| Excl. azithromycin | Serious adverse events | 3 | 1.054 (0.333 to 3.333) | 0.0 (-0.0 to 0.0) | 0.451 (0.048 to 4.250) | -0.0 (-0.0 to 0.0) | 38.5 |
| Excl. azithromycin | Acute rheumatic fever | Not estimable | Not estimable | Not estimable | Not estimable | Not estimable | Not estimable |
| Excl. azithromycin | Purulent complications | Not estimable | Not estimable | Not estimable | Not estimable | Not estimable | Not estimable |
| Excl. azithromycin | Poststreptococcal glomerulonephritis | Not estimable | Not estimable | Not estimable | Not estimable | Not estimable | Not estimable |

| Analysis | Outcome | Knots | RR, 5 vs 10 days (95% CI) | RD per 1,000, 5 vs 10 days (95% CI) | RR, 3 vs 10 days (95% CI) | RD per 1,000, 3 vs 10 days (95% CI) | AIC |
|---|---|---|---|---|---|---|---|
| **Excl. macrolides** | Clinical cure | 3 | 0.989 (0.973 to 1.006) | -9.9 (-24.9 to 5.5) | 1.017 (0.961 to 1.076) | 16.1 (-36.0 to 71.4) | -33.94 |
| Excl. macrolides | Relapse | 3 | 0.822 (0.367 to 1.843) | -8.8 (-31.4 to 41.8) | 0.345 (0.018 to 6.773) | -32.5 (-48.7 to 286.3) | 19.52 |
| Excl. macrolides | Adverse events | 3 | 0.953 (0.727 to 1.250) | -3.4 (-20.2 to 18.5) | 3.086 (1.137 to 8.381) | 154.4 (10.1 to 546.1) | 30.11 |
| Excl. macrolides | Serious adverse events | 3 | 1.311 (0.259 to 6.636) | 0.0 (-0.0 to 0.0) | 0.917 (0.045 to 18.689) | -0.0 (-0.0 to 0.0) | 30.06 |
| Excl. macrolides | Acute rheumatic fever | Not estimable | Not estimable | Not estimable | Not estimable | Not estimable | Not estimable |
| Excl. macrolides | Purulent complications | Not estimable | Not estimable | Not estimable | Not estimable | Not estimable | Not estimable |
| Excl. macrolides | Poststreptococcal glomerulonephritis | Not estimable | Not estimable | Not estimable | Not estimable | Not estimable | Not estimable |
| **Incl. placebo/no-treatment as 0 days** | Clinical cure | 3 | 0.987 (0.972 to 1.003) | -12.2 (-26.6 to 2.4) | 0.978 (0.935 to 1.023) | -20.3 (-60.7 to 21.9) | -75.82 |
| Incl. placebo/no-treatment as 0 days | Relapse | 3 | 1.053 (0.763 to 1.453) | 2.6 (-11.7 to 22.4) | 1.165 (0.784 to 1.731) | 8.2 (-10.7 to 36.2) | 56.69 |
| Incl. placebo/no-treatment as 0 days | Adverse events | 3 | 1.133 (0.913 to 1.405) | 9.8 (-6.4 to 30.0) | 1.402 (0.898 to 2.190) | 29.7 (-7.6 to 88.1) | 77.29 |
| Incl. placebo/no-treatment as 0 days | Mortality | Not estimable | Not estimable | Not estimable | Not estimable | Not estimable | Not estimable |
| Incl. placebo/no-treatment as 0 days | Serious adverse events | 3 | 1.181 (0.417 to 3.347) | 0.0 (-0.0 to 0.0) | 0.775 (0.259 to 2.320) | -0.0 (-0.0 to 0.0) | 56.92 |
| Incl. placebo/no-treatment as 0 days | Acute rheumatic fever | 3 | 2.553 (0.520 to 12.534) | 0.0 (-0.0 to 0.0) | 2.882 (0.438 to 18.951) | 0.0 (-0.0 to 0.0) | 35.28 |
| Incl. placebo/no-treatment as 0 days | Purulent complications | 3 | 0.708 (0.303 to 1.653) | -0.0 (-0.0 to 0.0) | 0.948 (0.304 to 2.952) | -0.0 (-0.0 to 0.0) | 23.11 |
| Incl. placebo/no-treatment as 0 days | Poststreptococcal glomerulonephritis | 3 | 0.965 (0.244 to 3.811) | -0.0 (-0.0 to 0.0) | 0.848 (0.172 to 4.172) | -0.0 (-0.0 to 0.0) | 34.66 |

AIC = Akaike information criterion; CI = confidence interval; RCS = restricted cubic spline; RD = risk difference; RR = risk ratio. Not estimable indicates that the model did not support the corresponding estimate.

**Supplement 19. Clinical cure: dose-response analysis for the primary analysis**

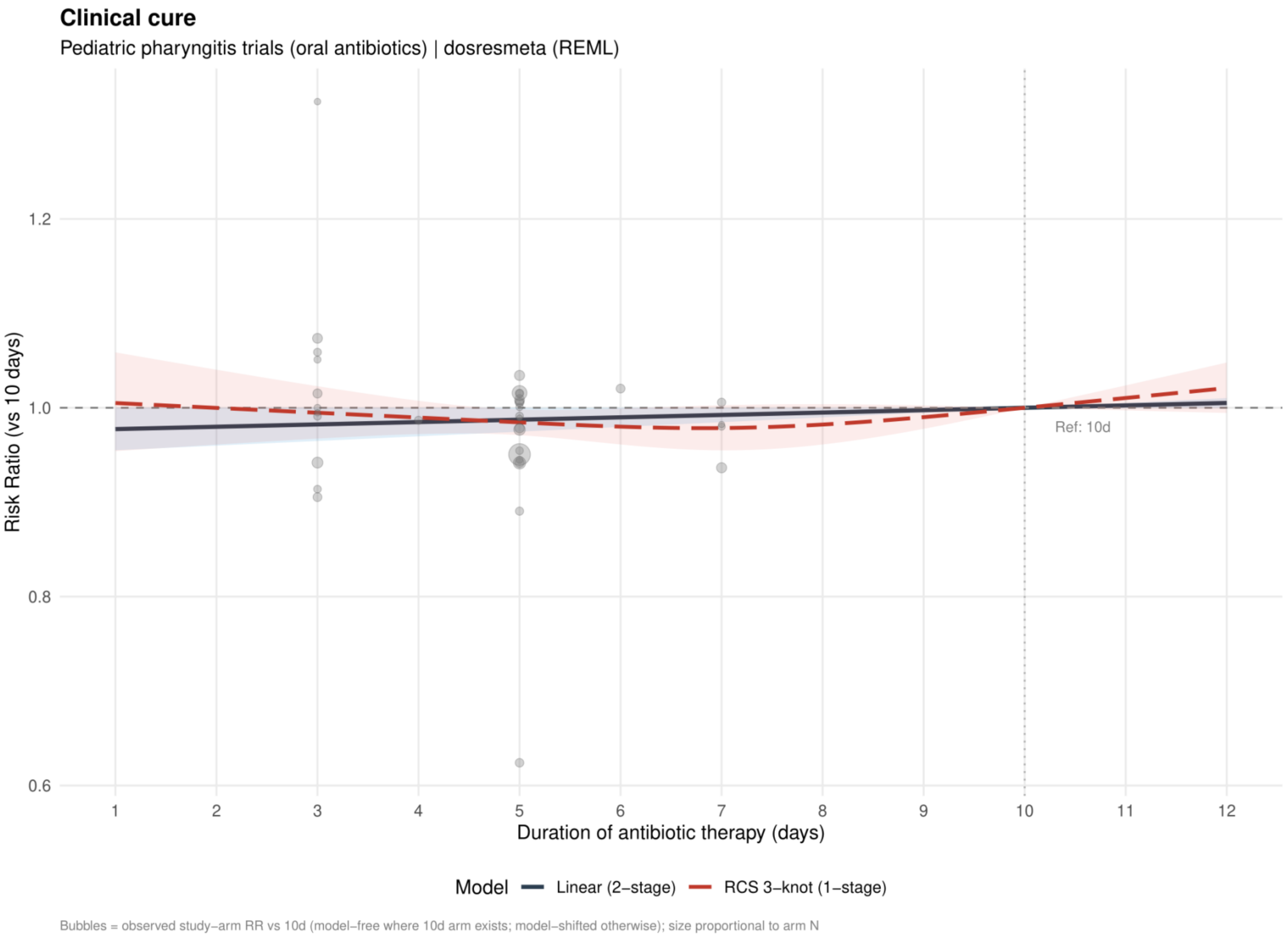


Curves show risk ratios relative to 10 days. The solid slate curve is the two stage linear model and, when present, the dashed red curve is the one stage restricted cubic spline with three knots; shaded bands show 95% confidence intervals. Bubbles show observed study arm effects and are scaled by arm sample size.

**Supplement 20. Clinical cure: linear estimates by risk of bias for the primary analysis**

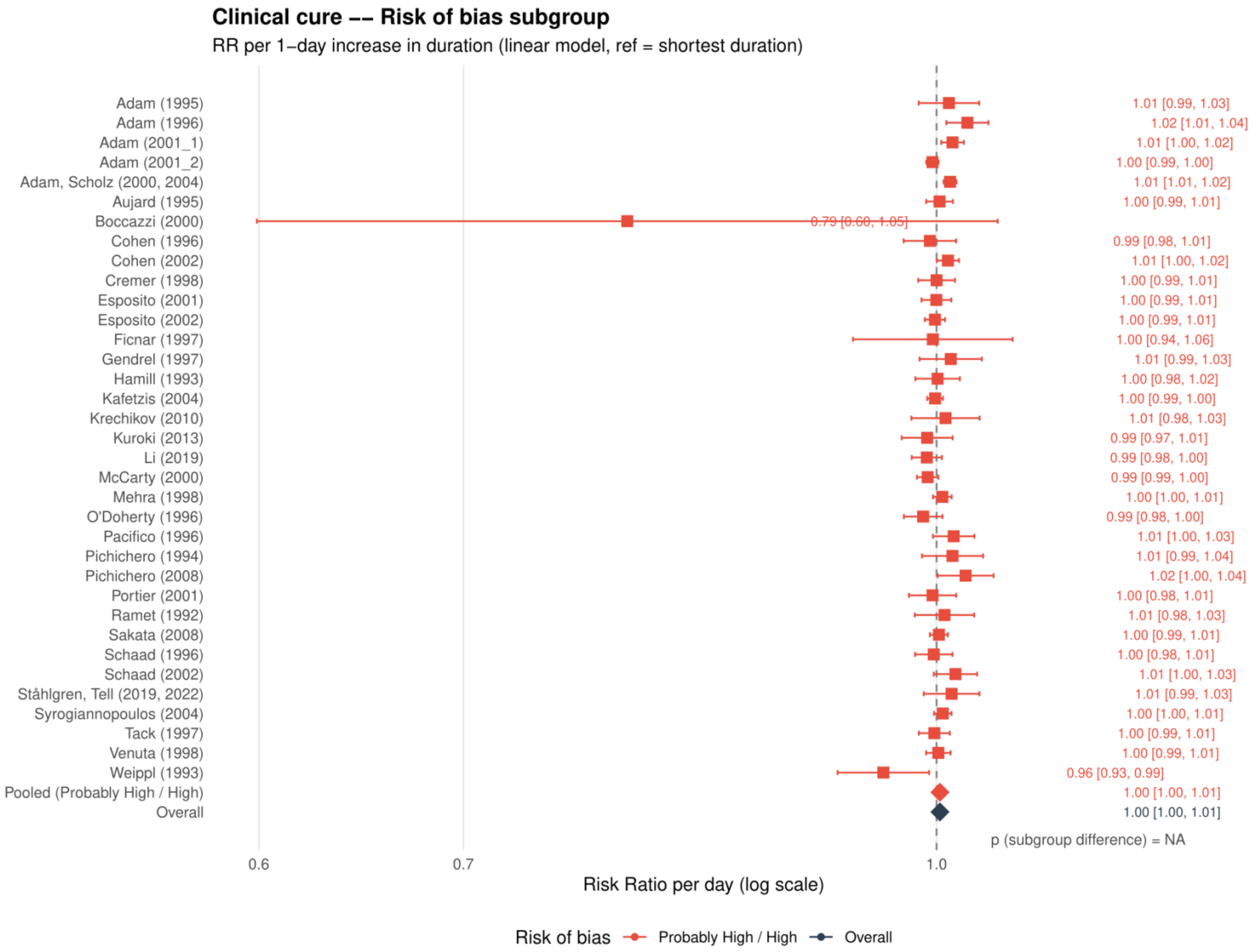


Study specific and pooled estimates are risk ratios per 1 day increase in antibiotic duration, with 95% confidence intervals.
*Only one collapsed risk of bias category contributed; the subgroup difference was not estimable.*

**Supplement 21. Clinical cure: restricted cubic spline estimates by risk of bias for the primary analysis**

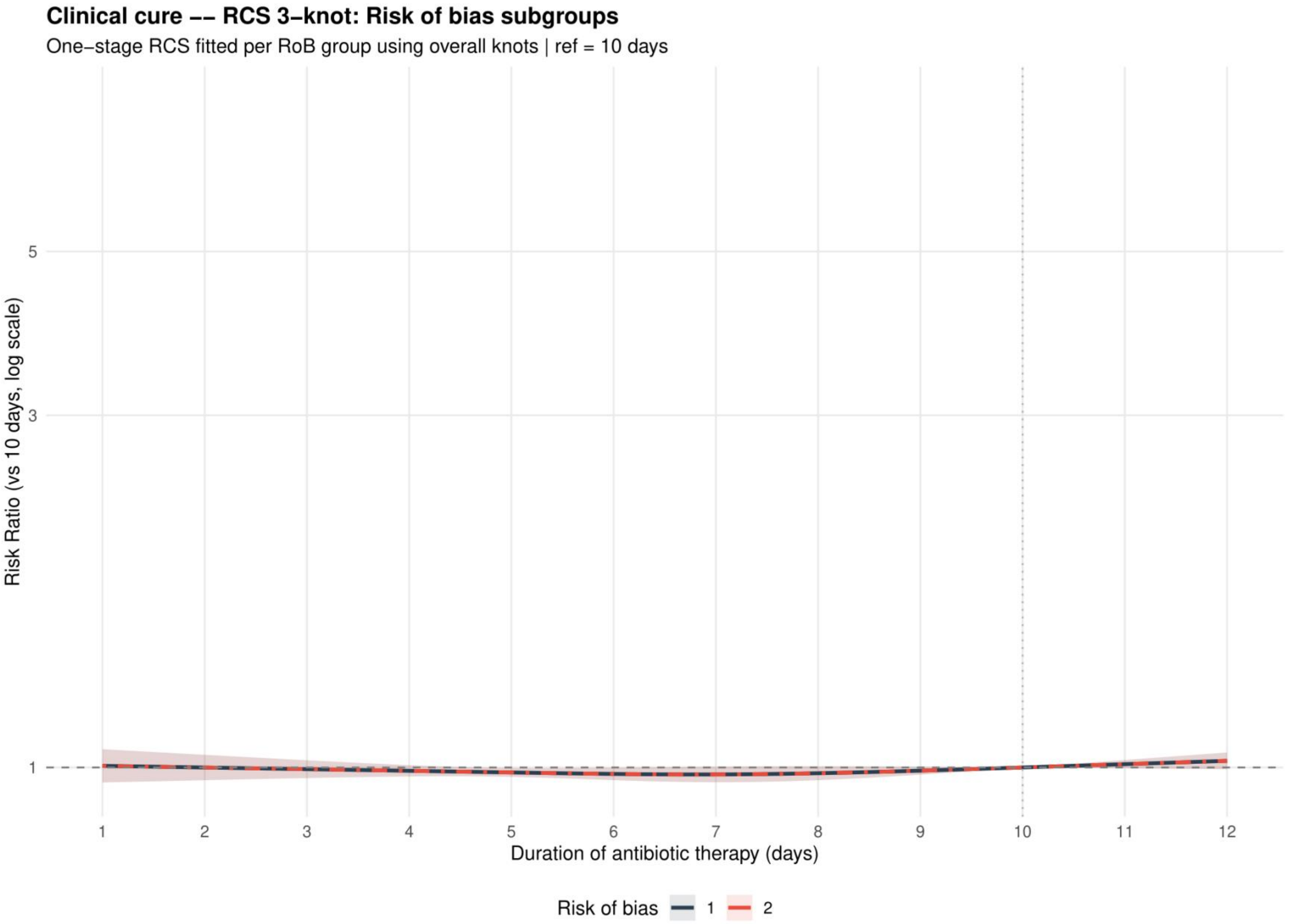


Curves show risk ratios relative to 10 days with 95% confidence intervals. Risk of bias codes: 1 = overall; 2 = low or probably low; 3 = probably high or high.

**Supplement 22. Clinical cure: funnel plot for the primary analysis**

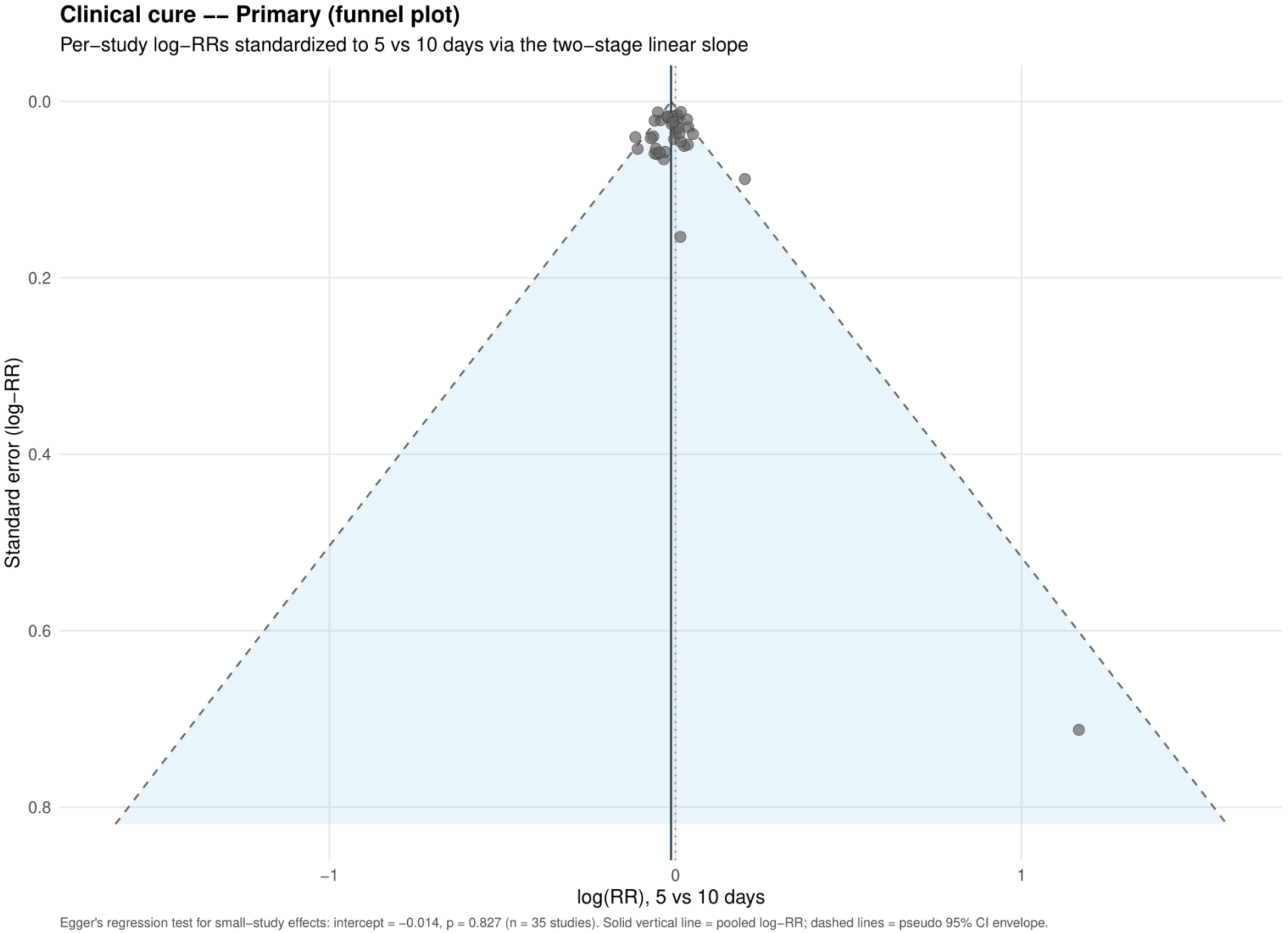


Study specific log risk ratios were standardized to 5 versus 10 days using the two stage linear slope. The solid vertical line is the pooled log risk ratio and dashed lines are pseudo 95% confidence limits; Egger's test is reported below the plot.

**Supplement 23. Clinical cure: dose-response analysis restricted to the same antibiotic and dose**

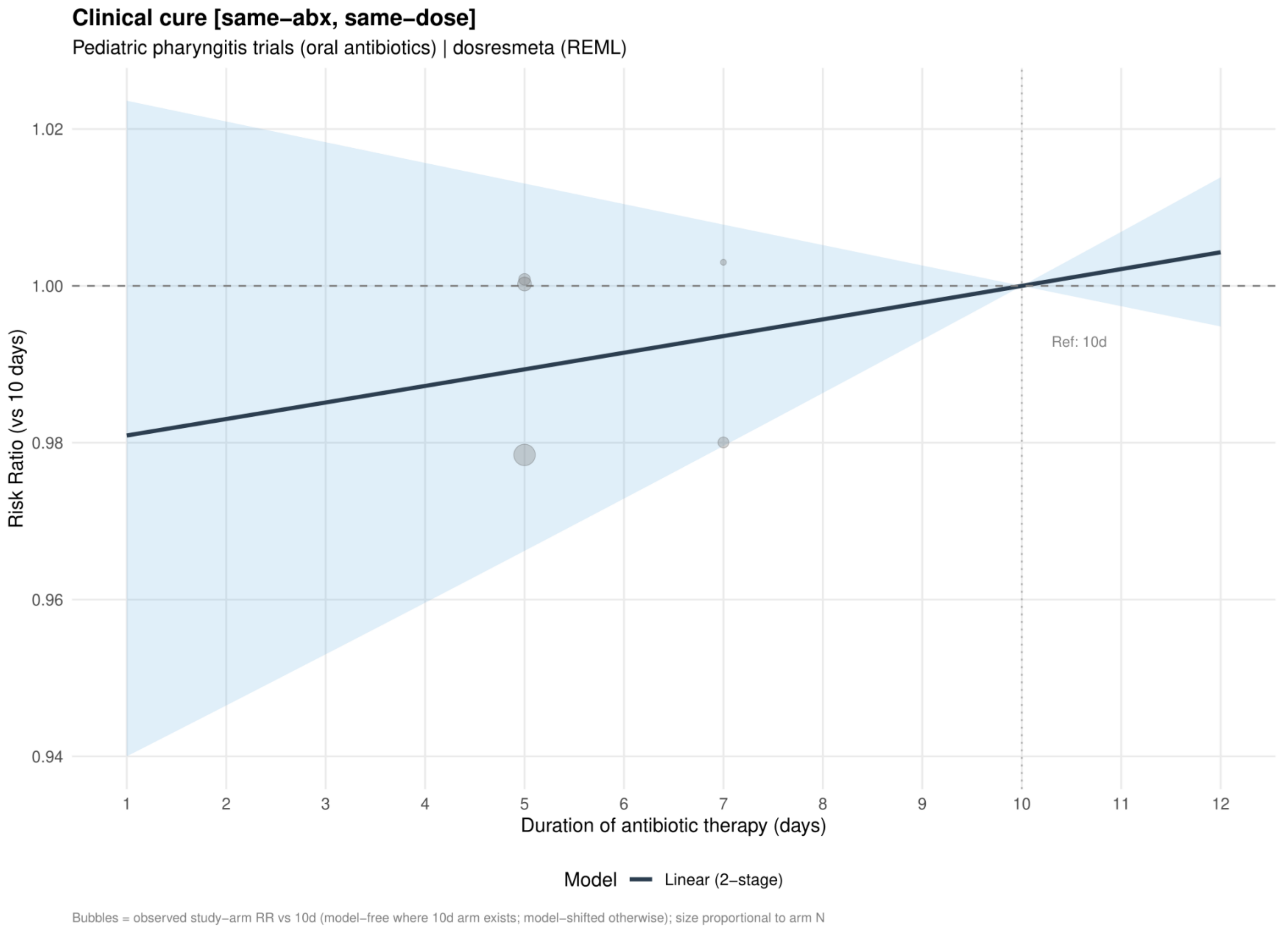


Curves show risk ratios relative to 10 days. The solid slate curve is the two stage linear model and, when present, the dashed red curve is the one stage restricted cubic spline with three knots; shaded bands show 95% confidence intervals. Bubbles show observed study arm effects and are scaled by arm sample size.

**Supplement 24. Clinical cure: dose-response analysis restricted to the same antibiotic**

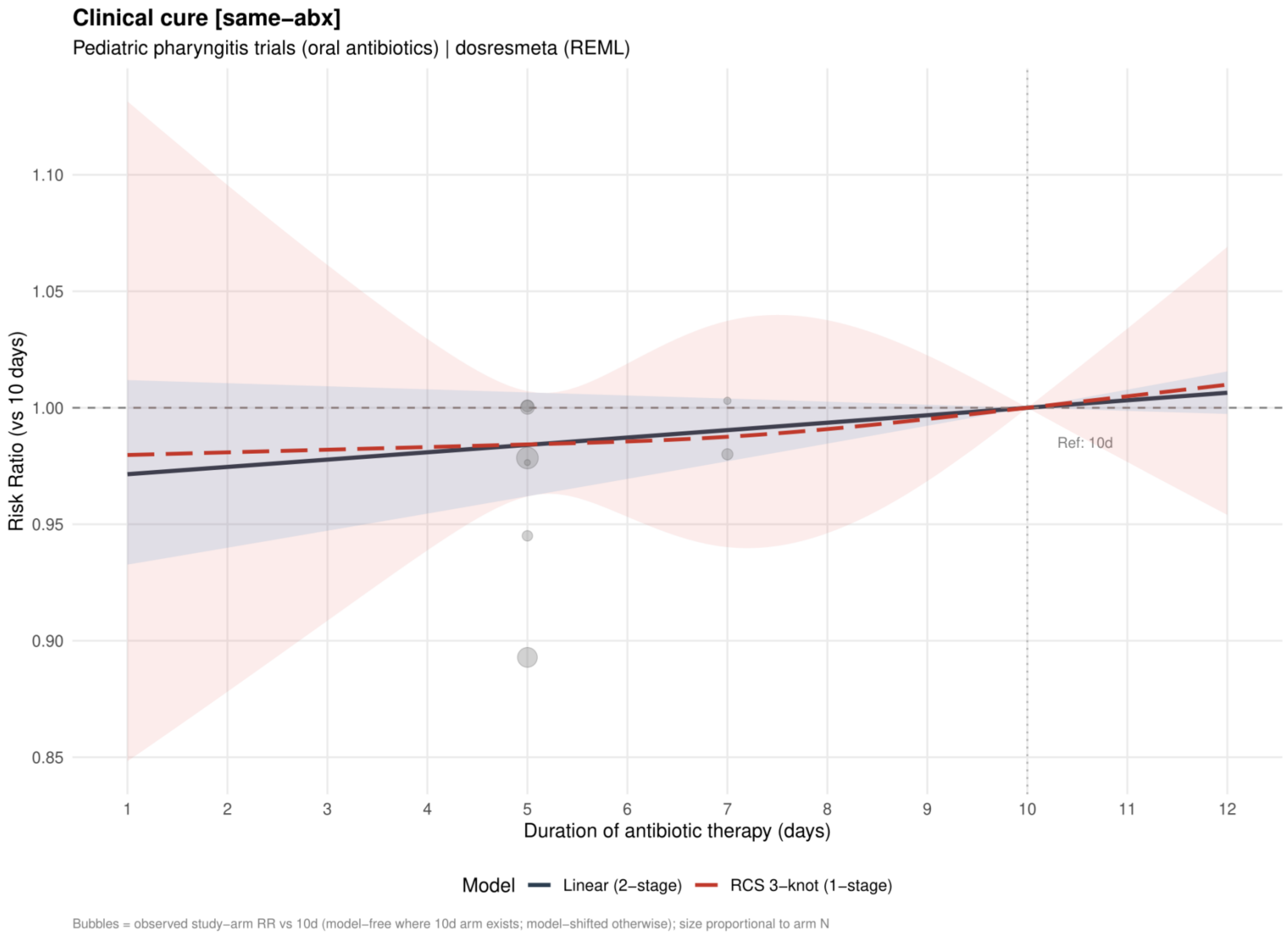


Curves show risk ratios relative to 10 days. The solid slate curve is the two stage linear model and, when present, the dashed red curve is the one stage restricted cubic spline with three knots; shaded bands show 95% confidence intervals. Bubbles show observed study arm effects and are scaled by arm sample size.

**Supplement 25. Clinical cure: restricted cubic spline estimates by risk of bias restricted to the same antibiotic**

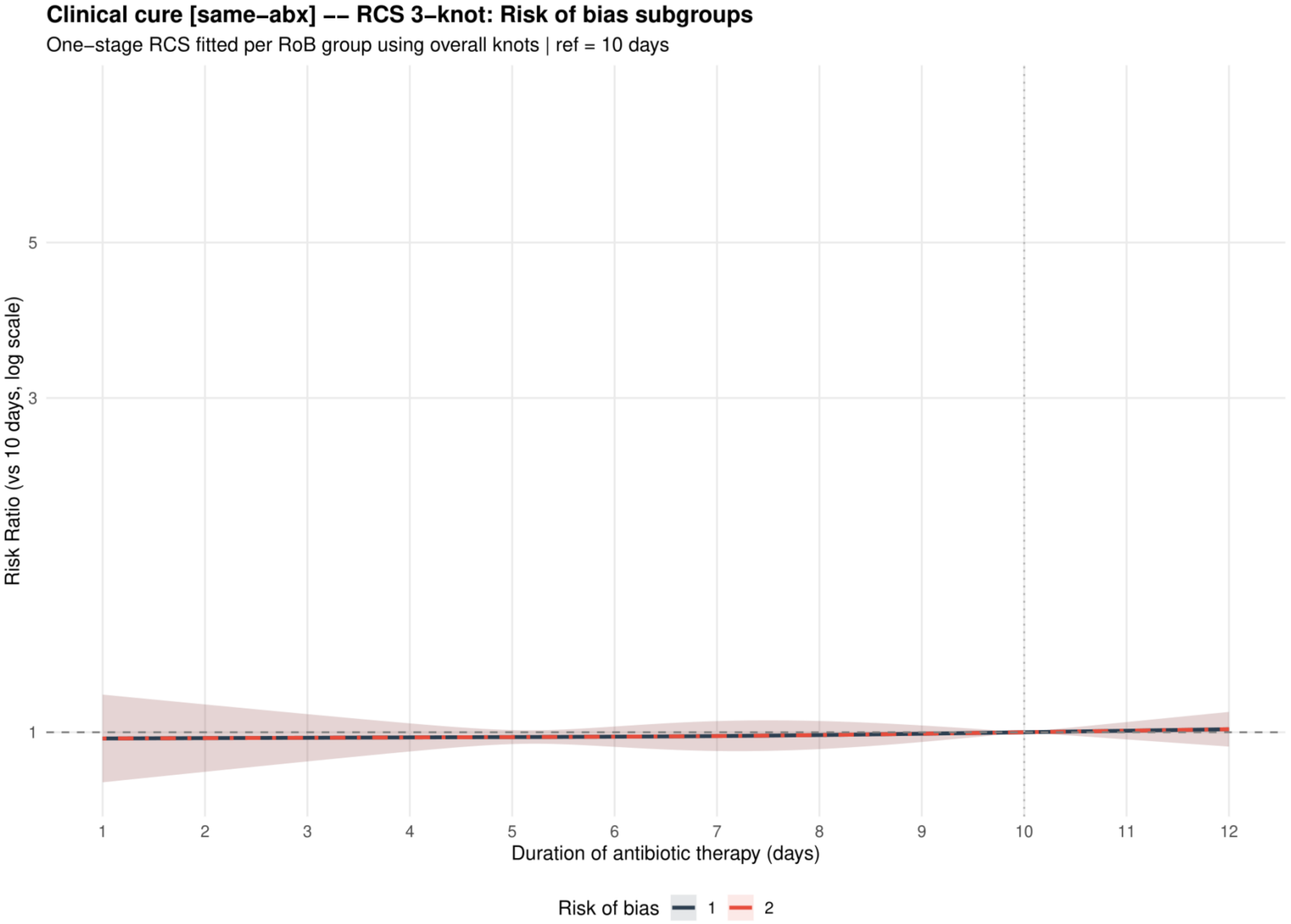


Curves show risk ratios relative to 10 days with 95% confidence intervals. Risk of bias codes: 1 = overall; 2 = low or probably low; 3 = probably high or high.

**Supplement 26. Clinical cure: dose-response analysis restricted to the same antibiotic class**

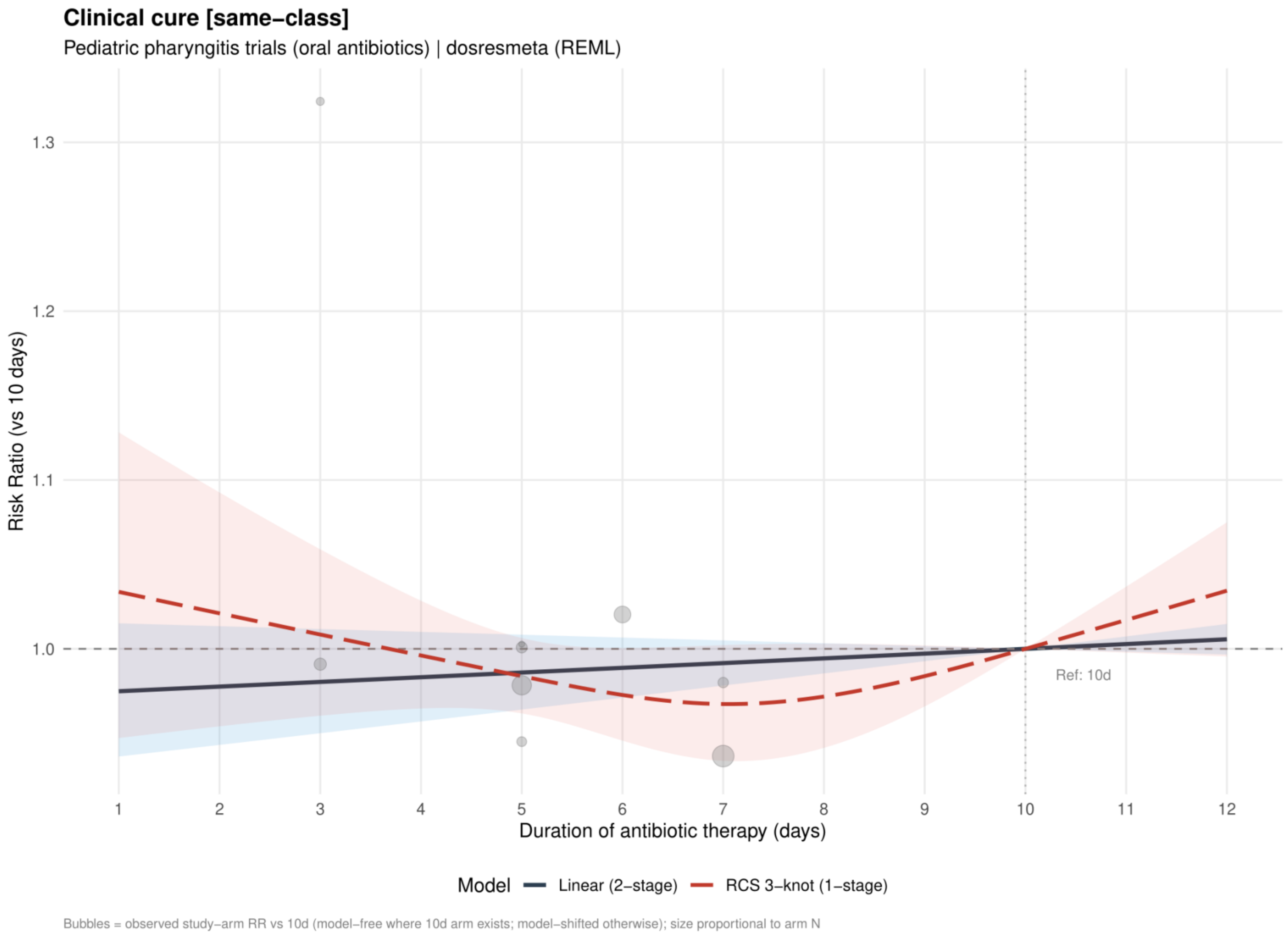


Curves show risk ratios relative to 10 days. The solid slate curve is the two stage linear model and, when present, the dashed red curve is the one stage restricted cubic spline with three knots; shaded bands show 95% confidence intervals. Bubbles show observed study arm effects and are scaled by arm sample size.

**Supplement 27. Clinical cure: linear estimates by risk of bias restricted to the same antibiotic class**

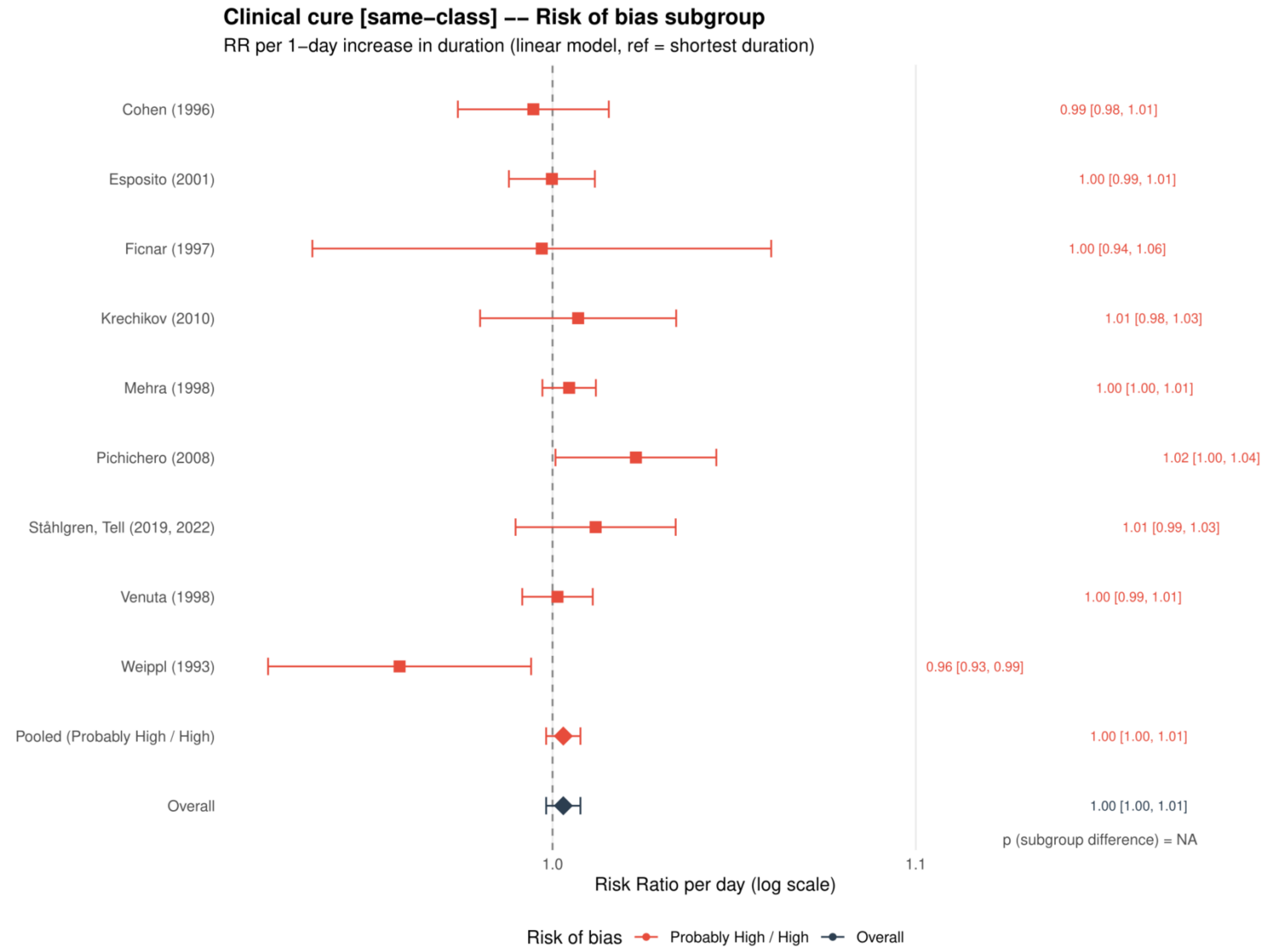


Study specific and pooled estimates are risk ratios per 1 day increase in antibiotic duration, with 95% confidence intervals.
*Only one collapsed risk of bias category contributed; the subgroup difference was not estimable.*

**Supplement 28. Clinical cure: restricted cubic spline estimates by risk of bias restricted to the same antibiotic class**

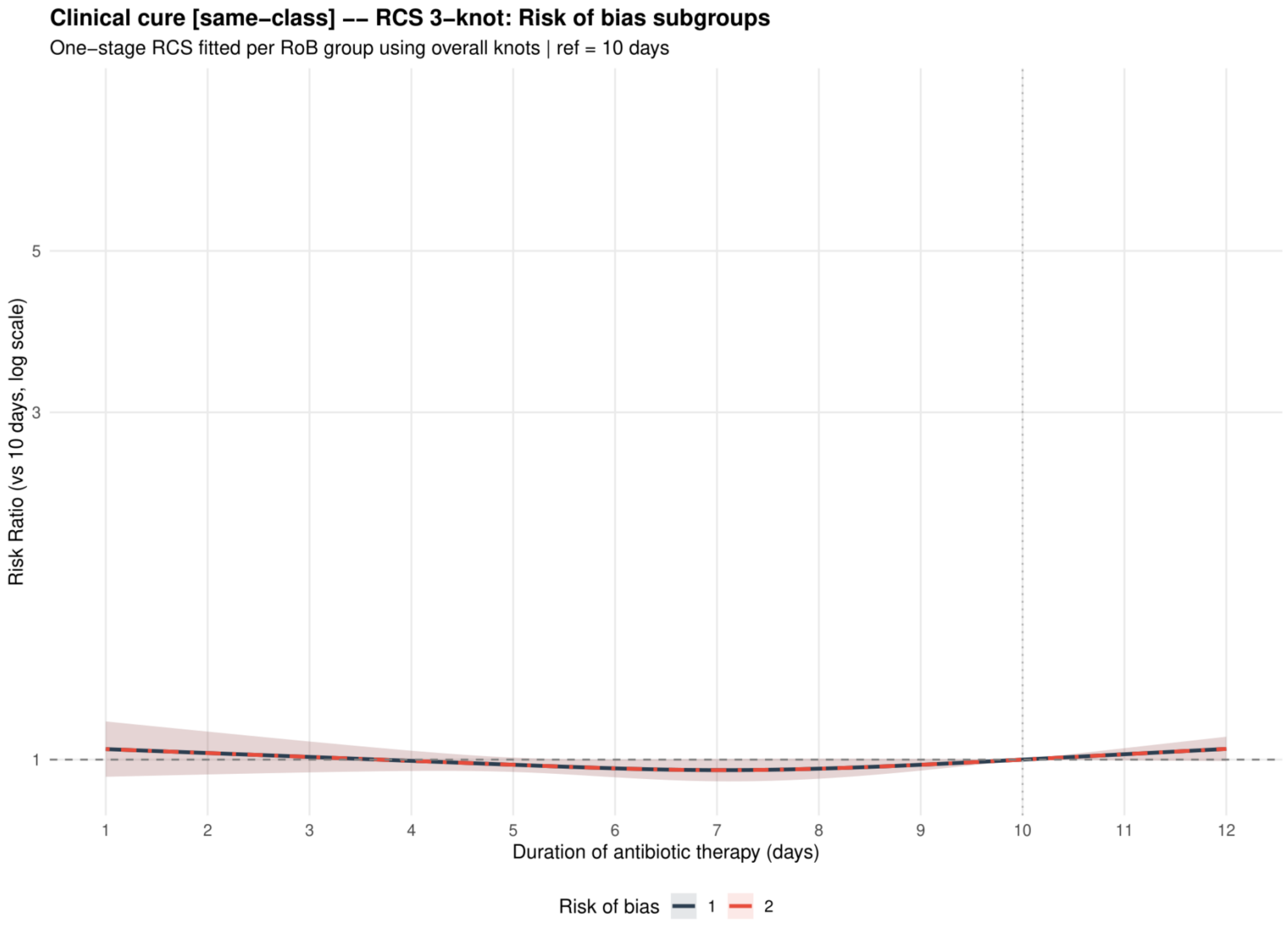


Curves show risk ratios relative to 10 days with 95% confidence intervals. Risk of bias codes: 1 = overall; 2 = low or probably low; 3 = probably high or high.

**Supplement 29. Clinical cure: dose-response analysis restricted to the penicillins**

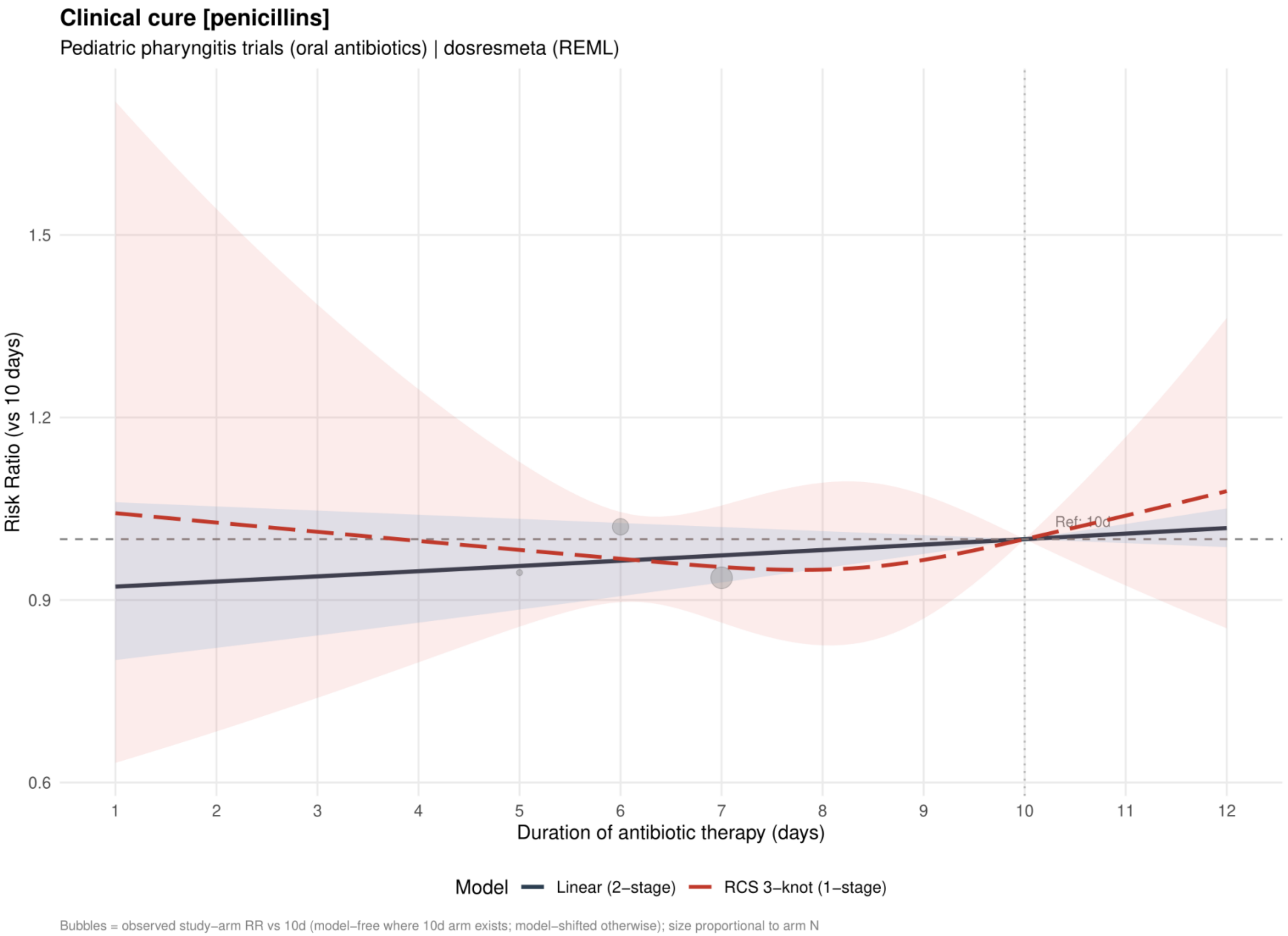


Curves show risk ratios relative to 10 days. The solid slate curve is the two stage linear model and, when present, the dashed red curve is the one stage restricted cubic spline with three knots; shaded bands show 95% confidence intervals. Bubbles show observed study arm effects and are scaled by arm sample size.

**Supplement 30. Clinical cure: linear estimates by risk of bias restricted to the penicillins**

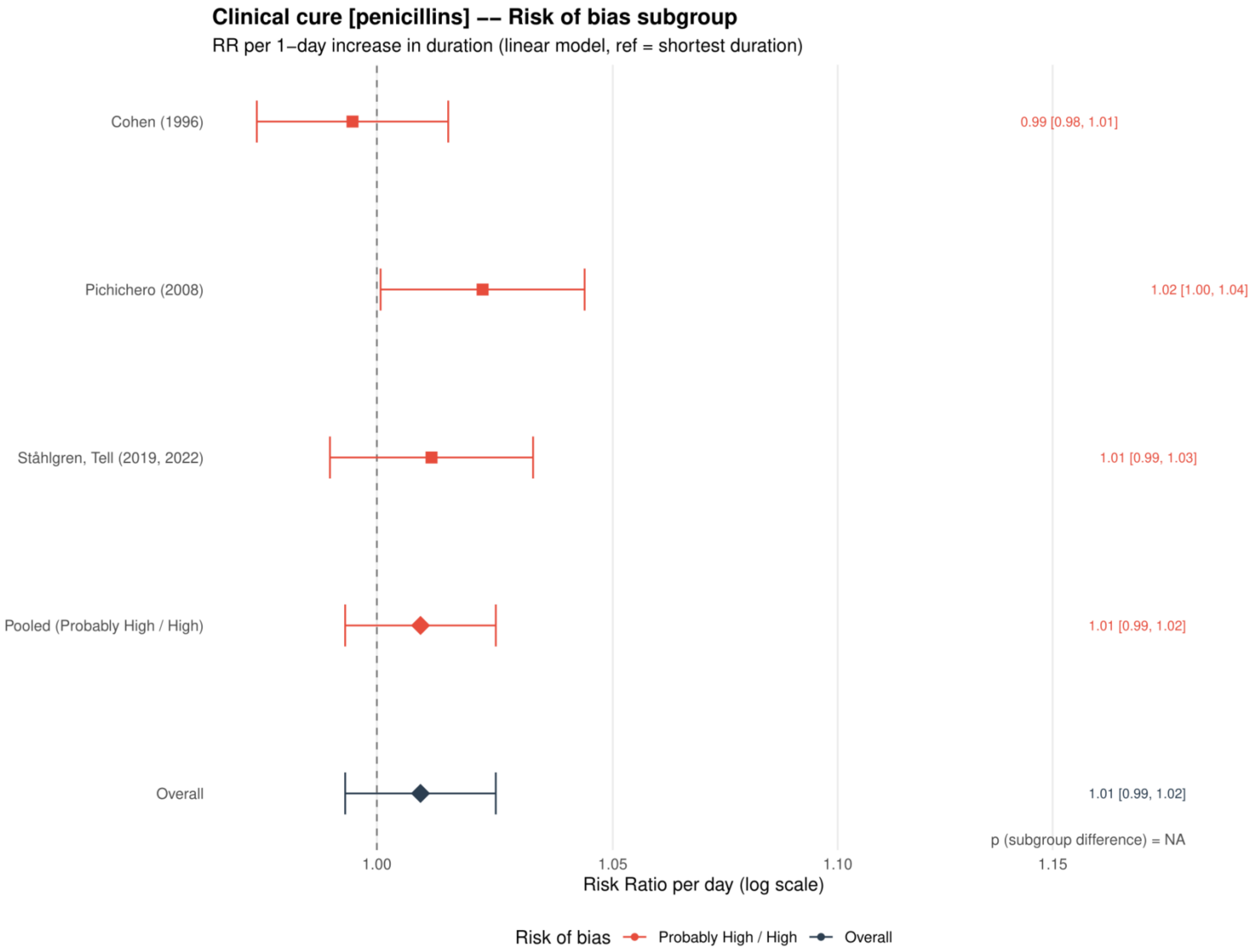


Study specific and pooled estimates are risk ratios per 1 day increase in antibiotic duration, with 95% confidence intervals.
*Only one collapsed risk of bias category contributed; the subgroup difference was not estimable.*

**Supplement 31. Clinical cure: restricted cubic spline estimates by risk of bias restricted to the penicillins**

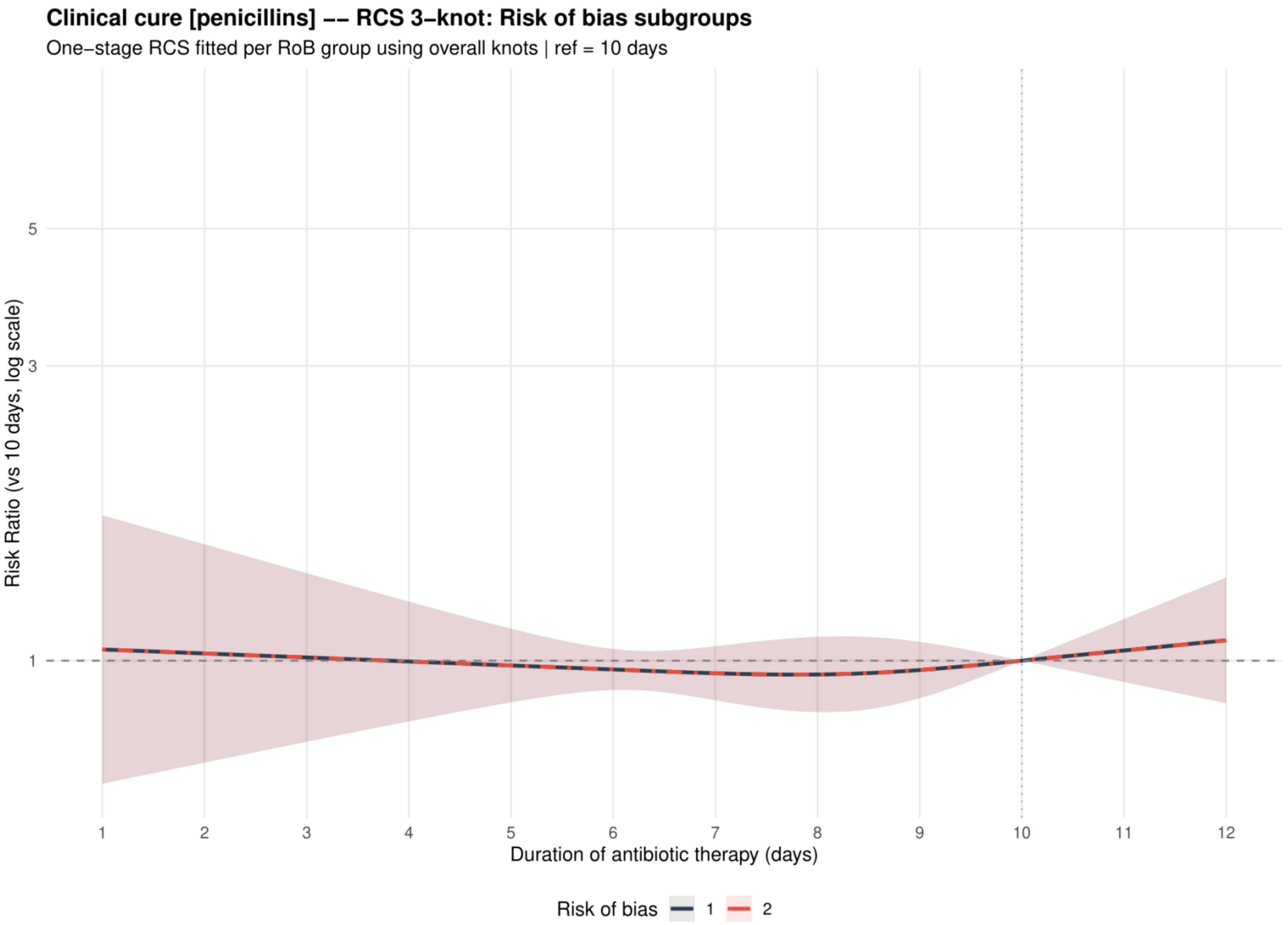


Curves show risk ratios relative to 10 days with 95% confidence intervals. Risk of bias codes: 1 = overall; 2 = low or probably low; 3 = probably high or high.

**Supplement 32. Clinical cure: dose-response analysis excluding azithromycin**

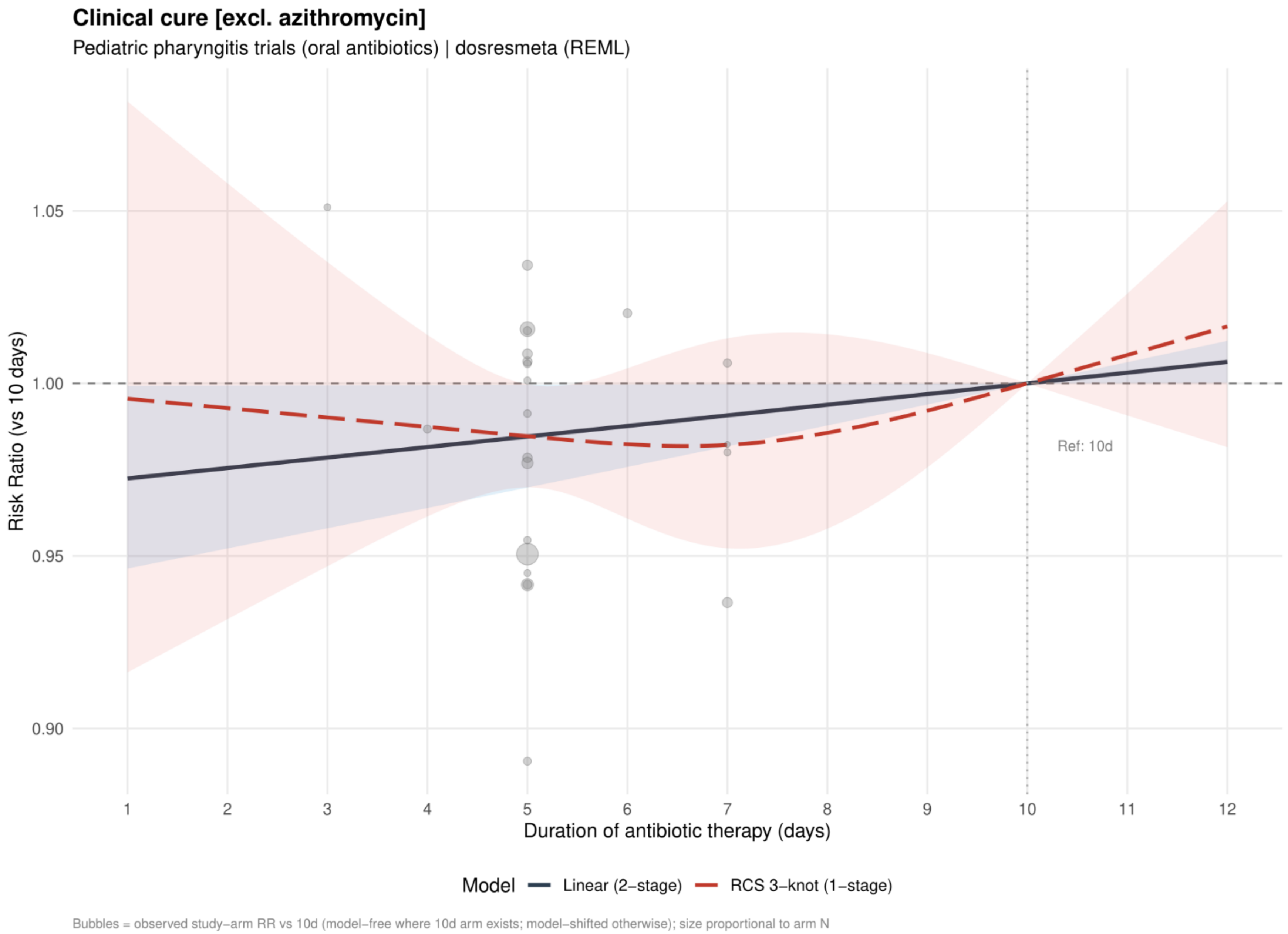


Curves show risk ratios relative to 10 days. The solid slate curve is the two stage linear model and, when present, the dashed red curve is the one stage restricted cubic spline with three knots; shaded bands show 95% confidence intervals. Bubbles show observed study arm effects and are scaled by arm sample size.

**Supplement 33. Clinical cure: linear estimates by risk of bias excluding azithromycin**

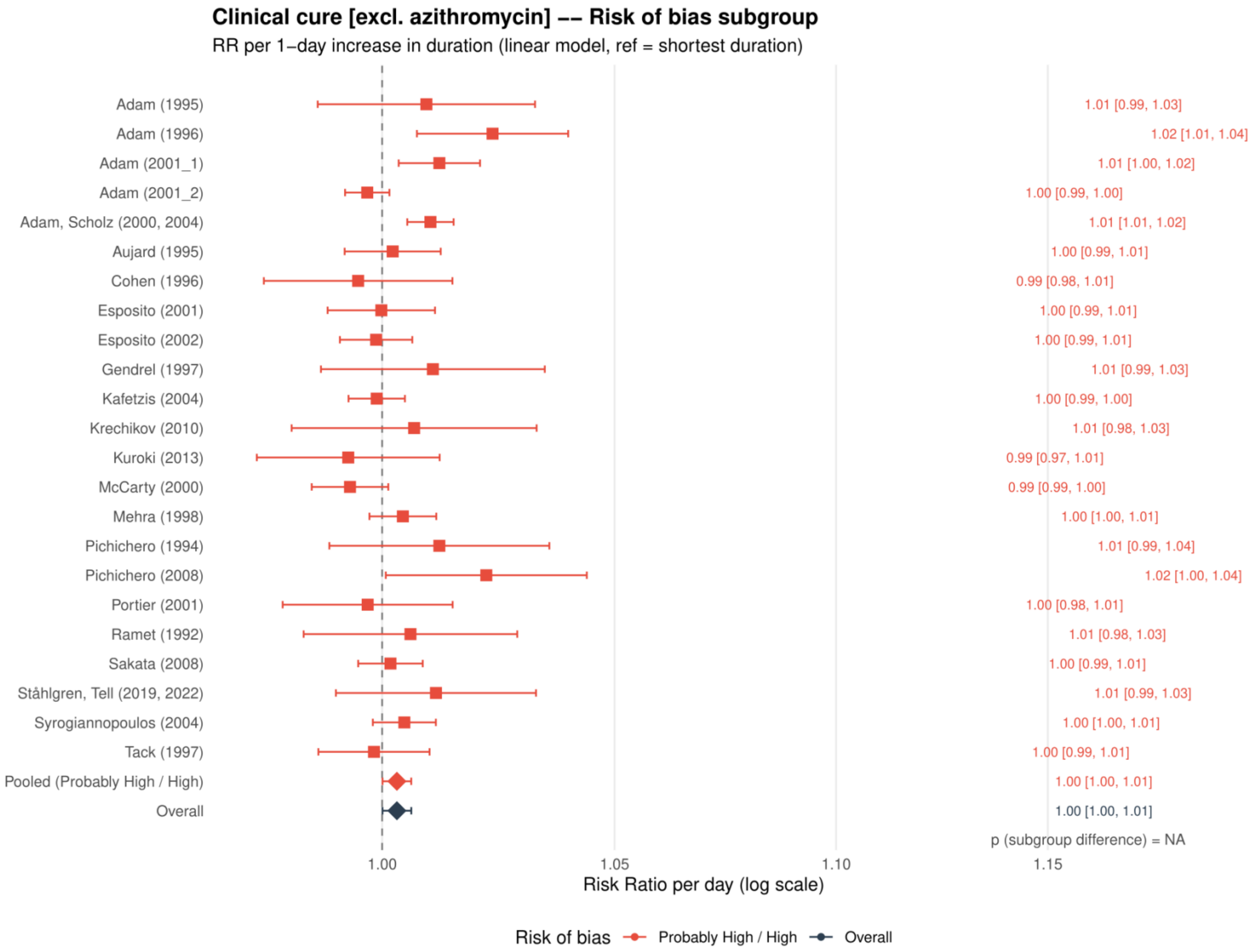


Study specific and pooled estimates are risk ratios per 1 day increase in antibiotic duration, with 95% confidence intervals.
*Only one collapsed risk of bias category contributed; the subgroup difference was not estimable.*

**Supplement 34. Clinical cure: restricted cubic spline estimates by risk of bias excluding azithromycin**

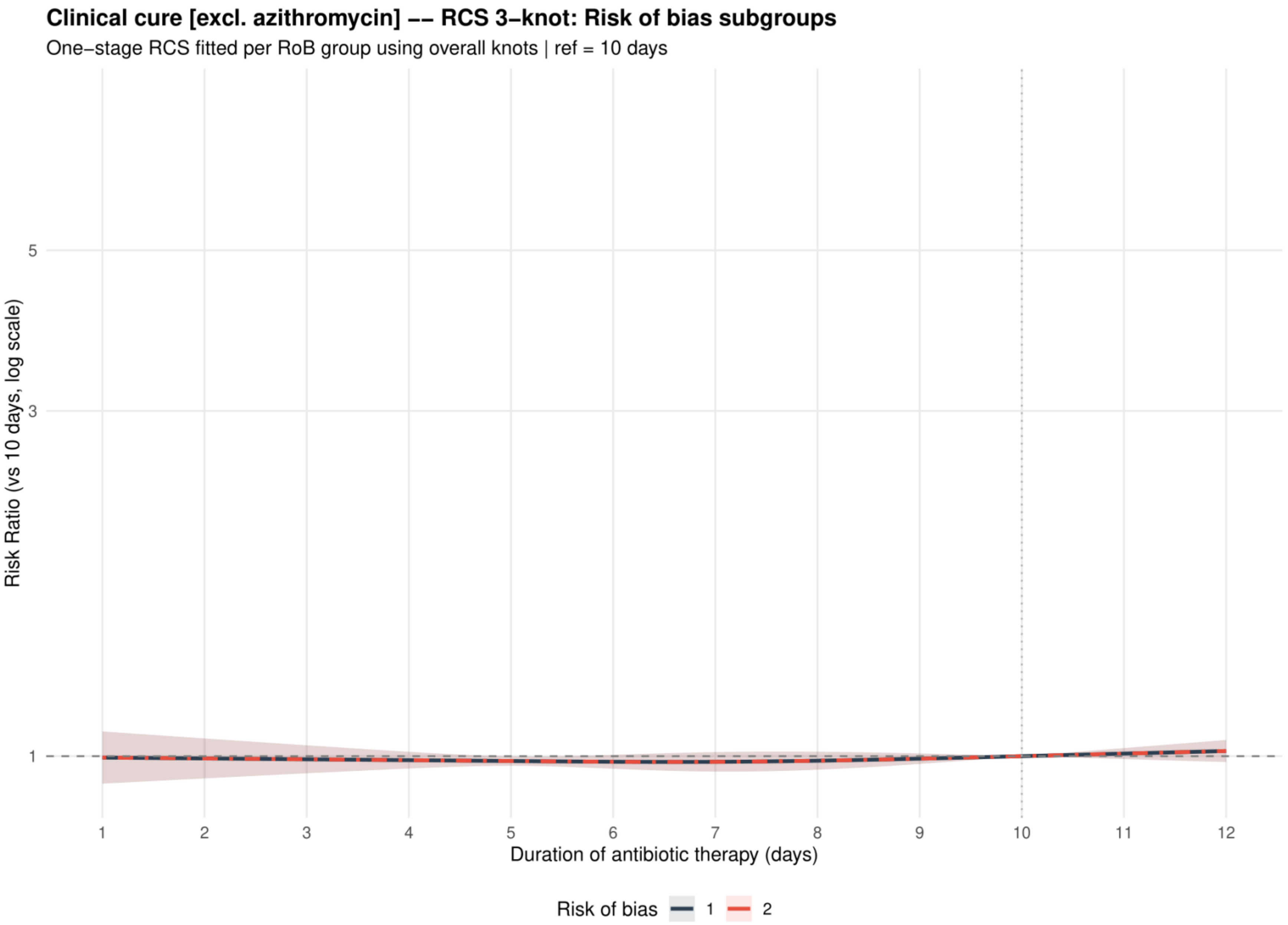


Curves show risk ratios relative to 10 days with 95% confidence intervals. Risk of bias codes: 1 = overall; 2 = low or probably low; 3 = probably high or high.

**Supplement 35. Clinical cure: dose-response analysis excluding macrolides**

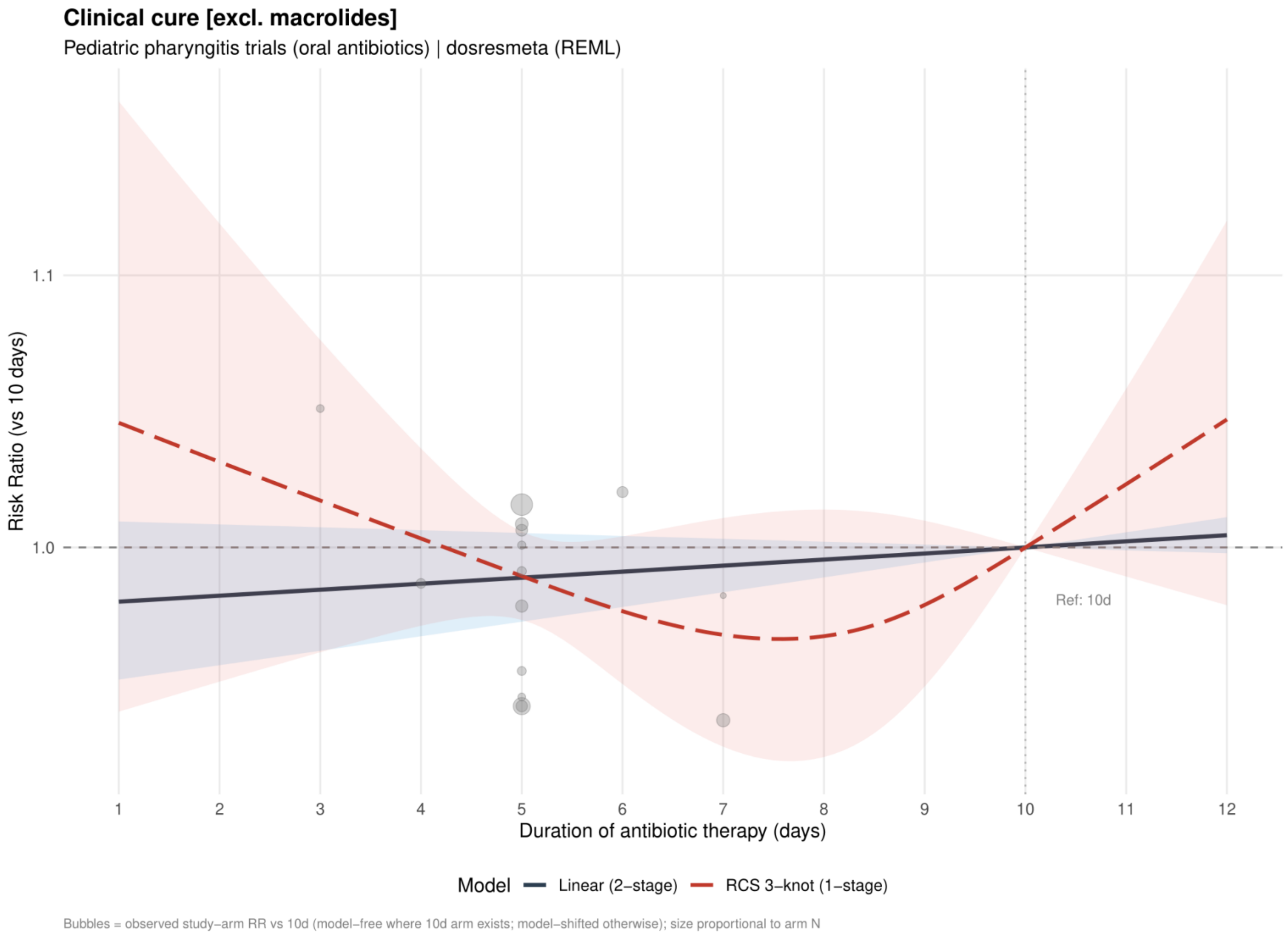


Curves show risk ratios relative to 10 days. The solid slate curve is the two stage linear model and, when present, the dashed red curve is the one stage restricted cubic spline with three knots; shaded bands show 95% confidence intervals. Bubbles show observed study arm effects and are scaled by arm sample size.

**Supplement 36. Clinical cure: linear estimates by risk of bias excluding macrolides**

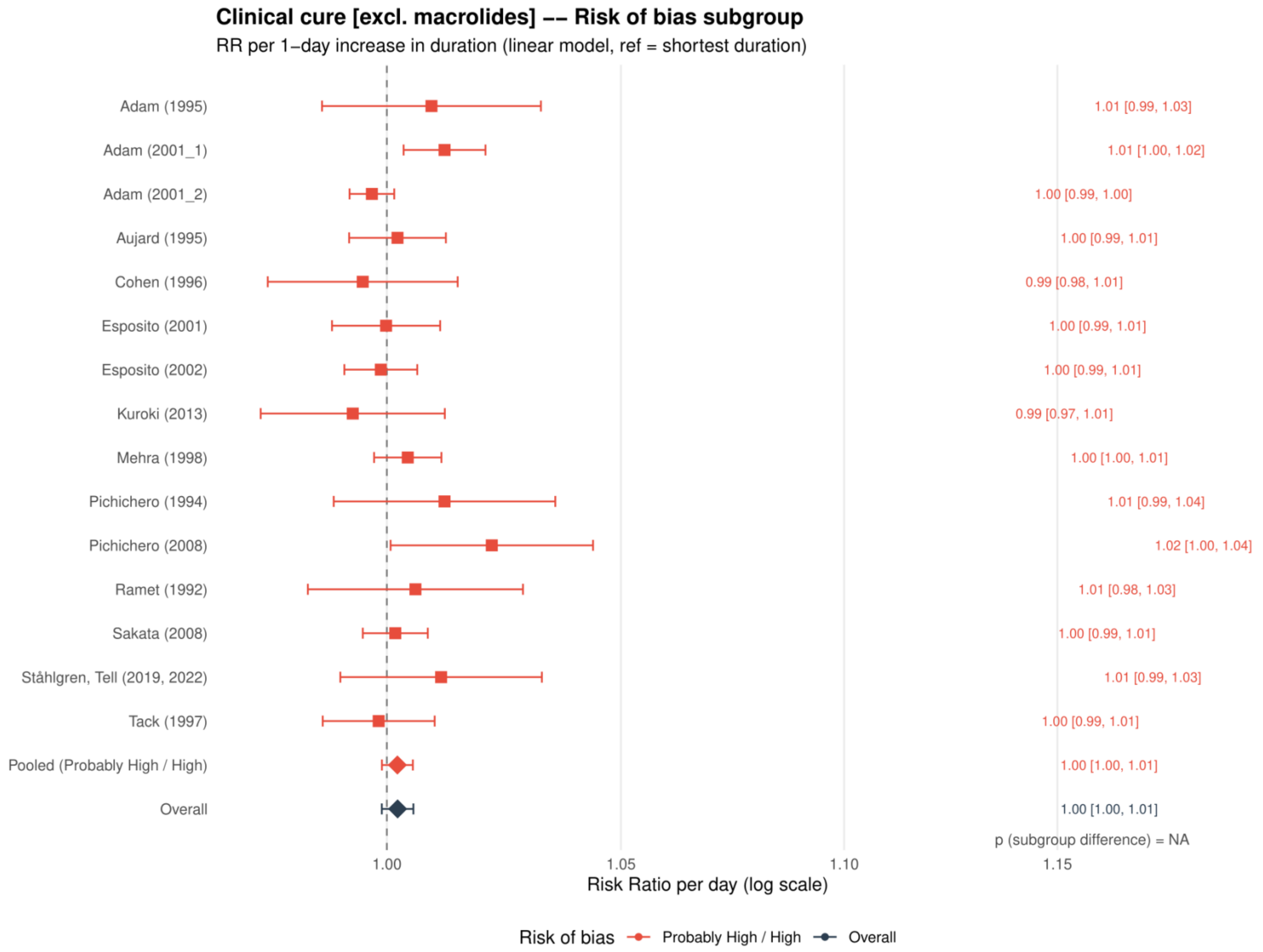


Study specific and pooled estimates are risk ratios per 1 day increase in antibiotic duration, with 95% confidence intervals.
*Only one collapsed risk of bias category contributed; the subgroup difference was not estimable.*

**Supplement 37. Clinical cure: restricted cubic spline estimates by risk of bias excluding macrolides**

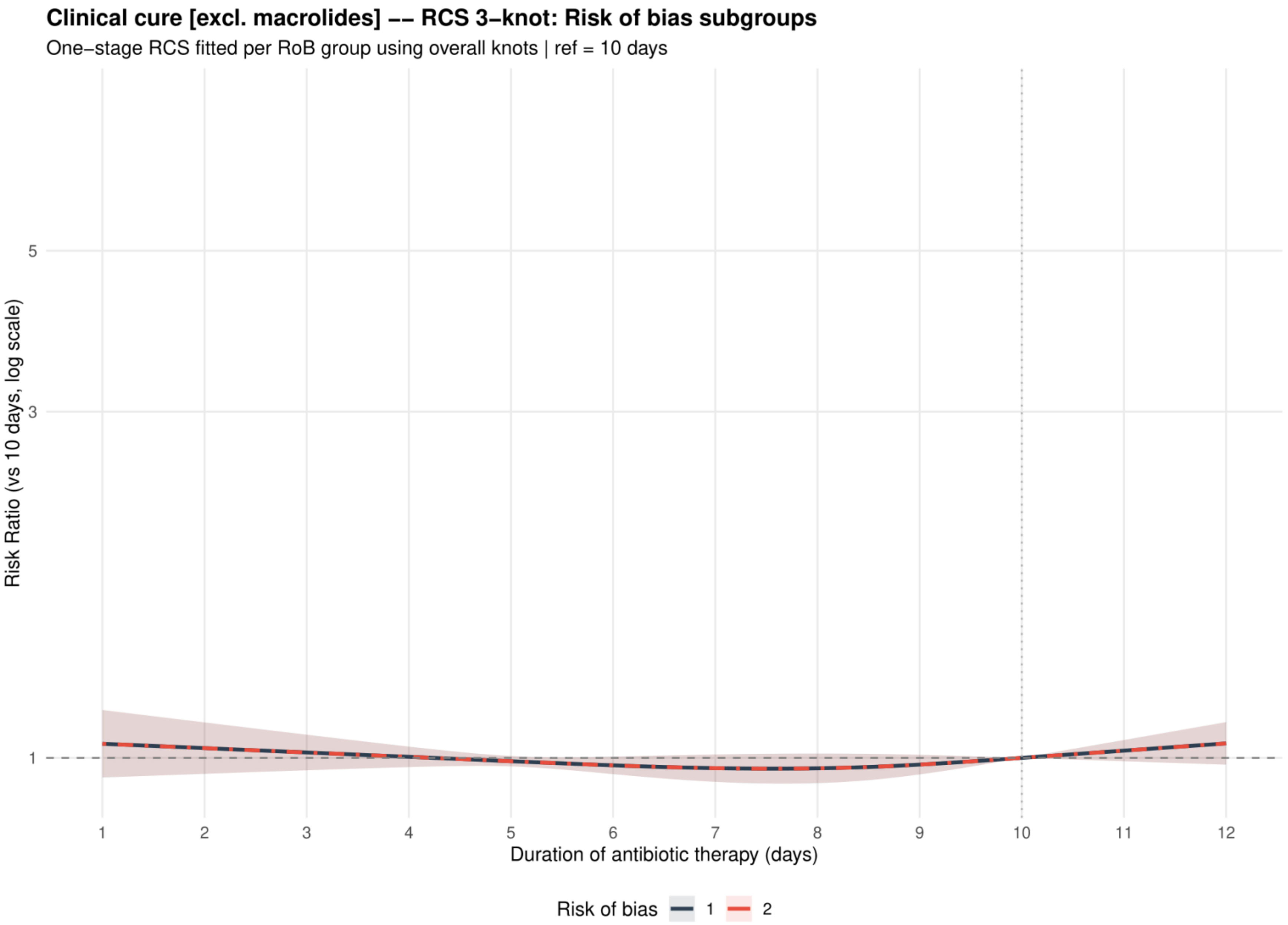


Curves show risk ratios relative to 10 days with 95% confidence intervals. Risk of bias codes: 1 = overall; 2 = low or probably low; 3 = probably high or high.

**Supplement 38. Clinical cure: dose-response analysis including placebo or no treatment as 0 days**

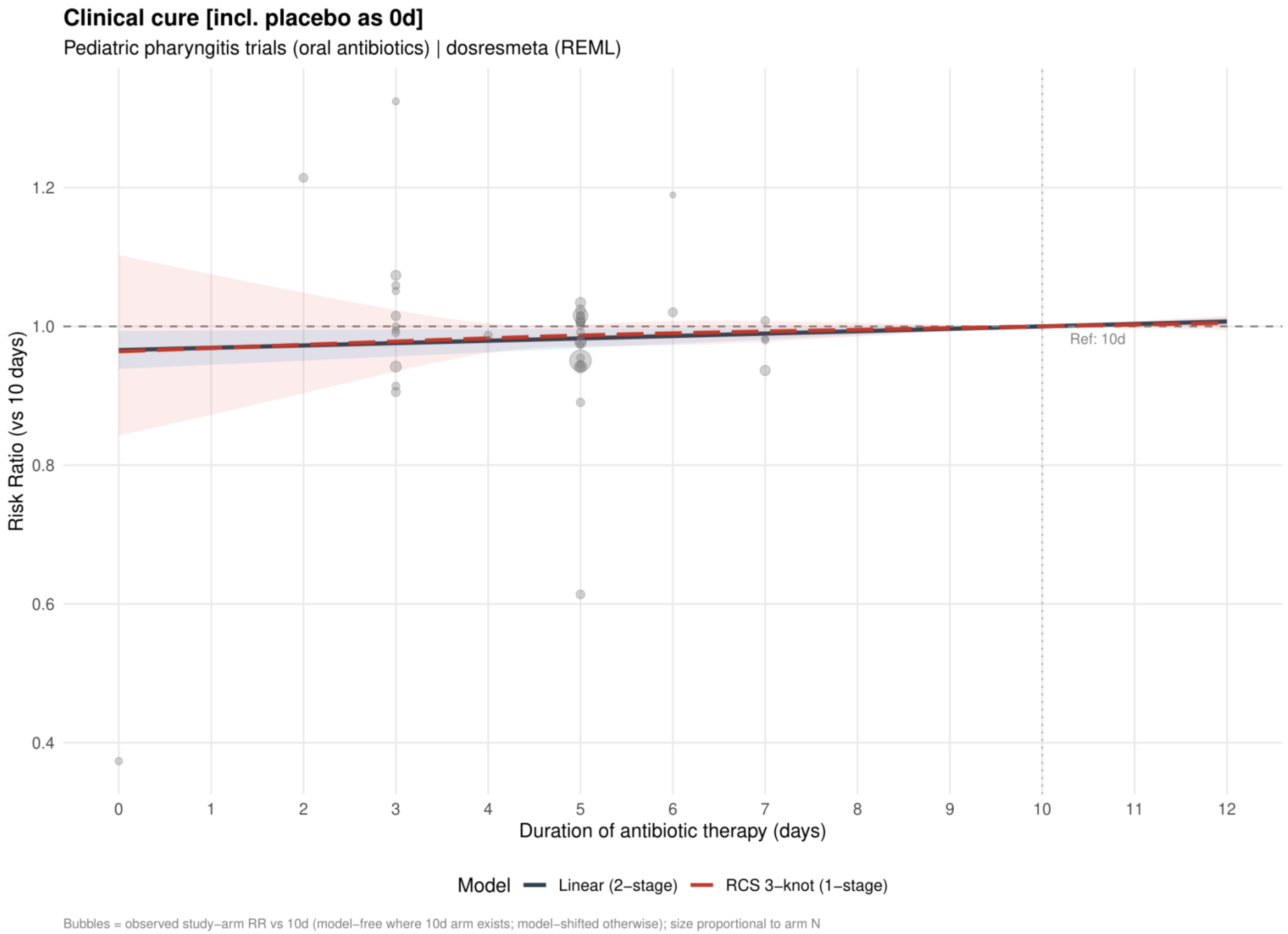


Curves show risk ratios relative to 10 days. The solid slate curve is the two stage linear model and, when present, the dashed red curve is the one stage restricted cubic spline with three knots; shaded bands show 95% confidence intervals. Bubbles show observed study arm effects and are scaled by arm sample size.

**Supplement 39. Clinical cure: linear estimates by risk of bias including placebo or no treatment as 0 days**

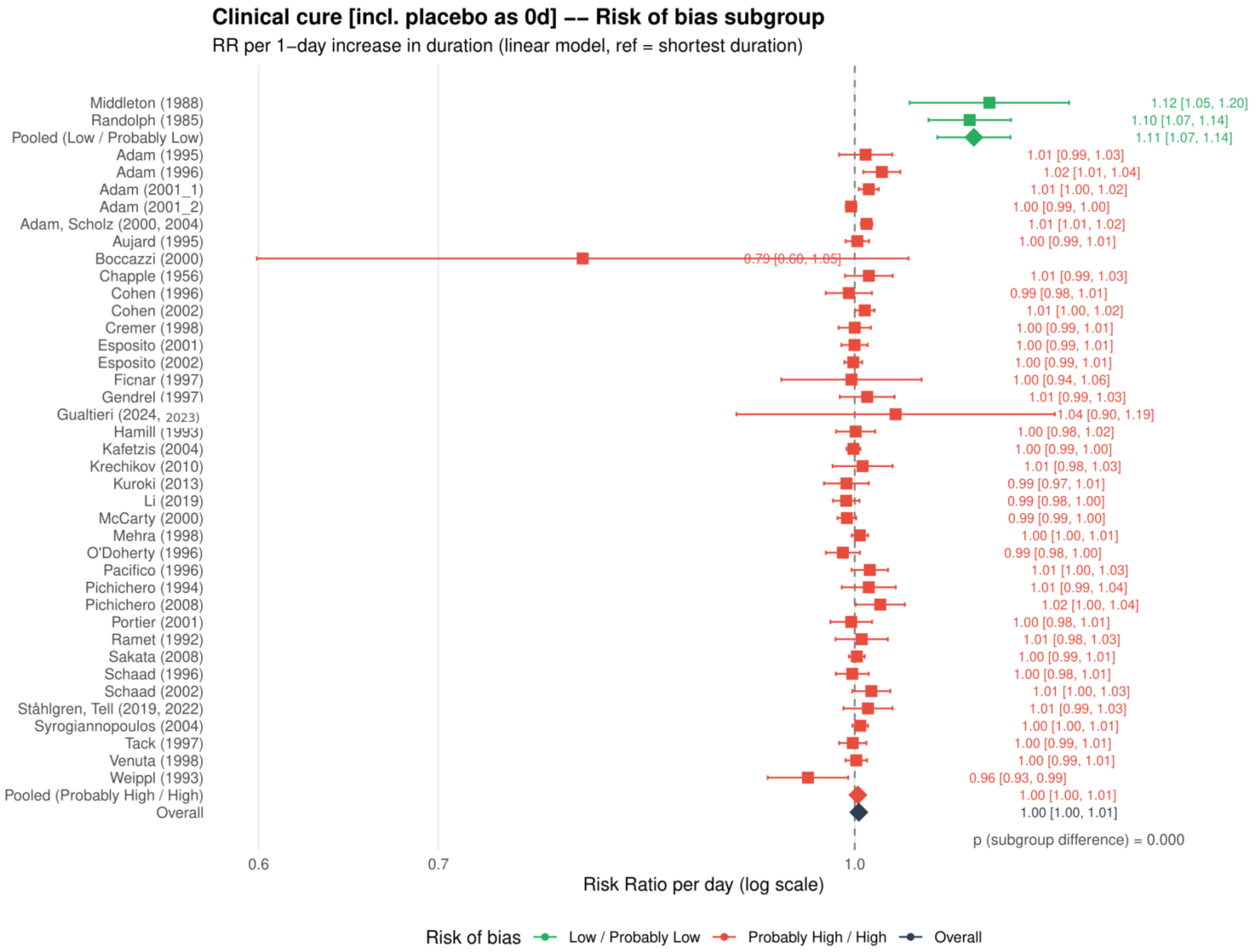


Study specific and pooled estimates are risk ratios per 1 day increase in antibiotic duration, with 95% confidence intervals.

**Supplement 40. Clinical cure: restricted cubic spline estimates by risk of bias including placebo or no treatment as 0 days**

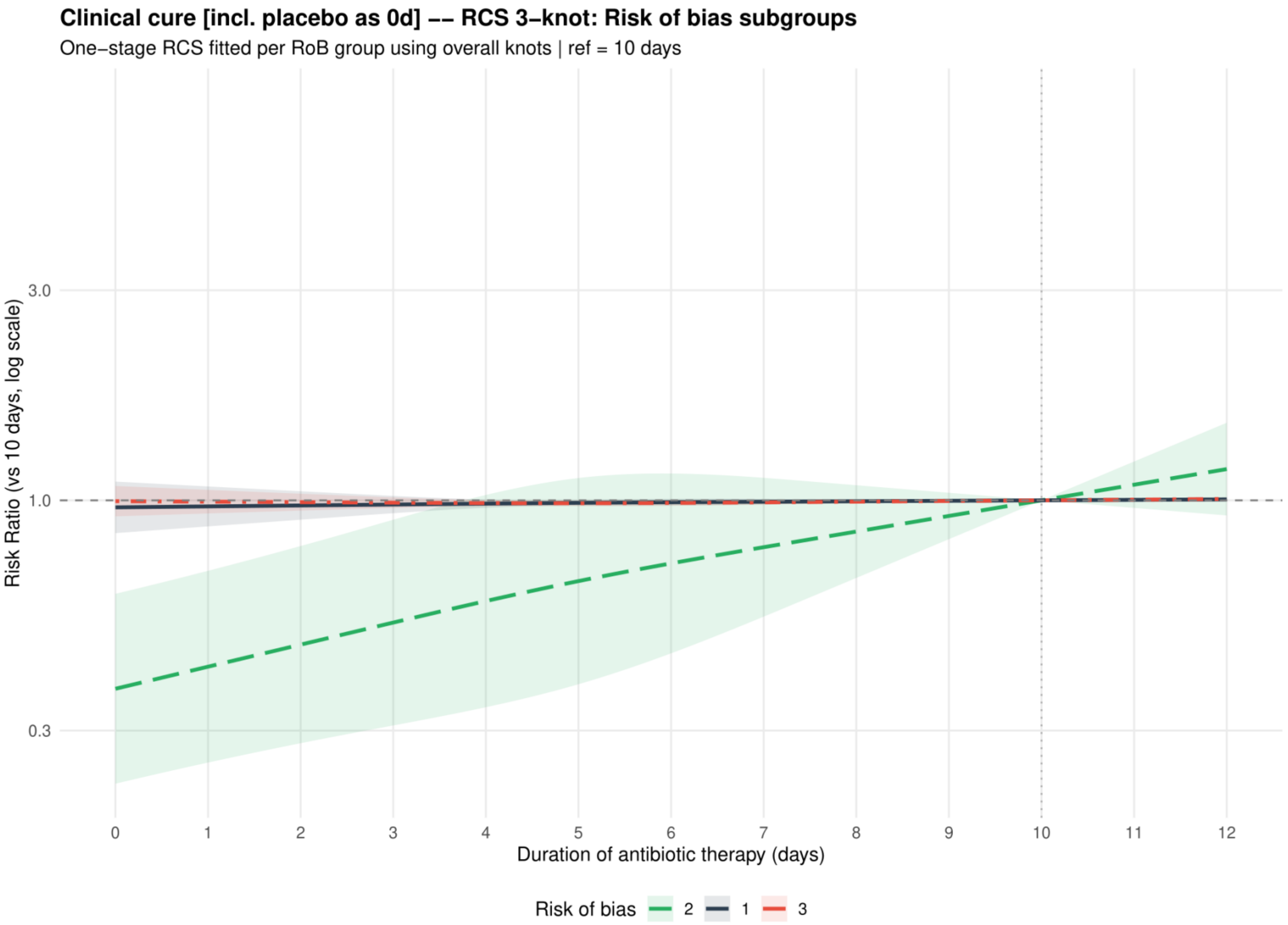


Curves show risk ratios relative to 10 days with 95% confidence intervals. Risk of bias codes: 1 = overall; 2 = low or probably low; 3 = probably high or high.

**Supplement 41. Clinical cure: funnel plot including placebo or no treatment as 0 days**

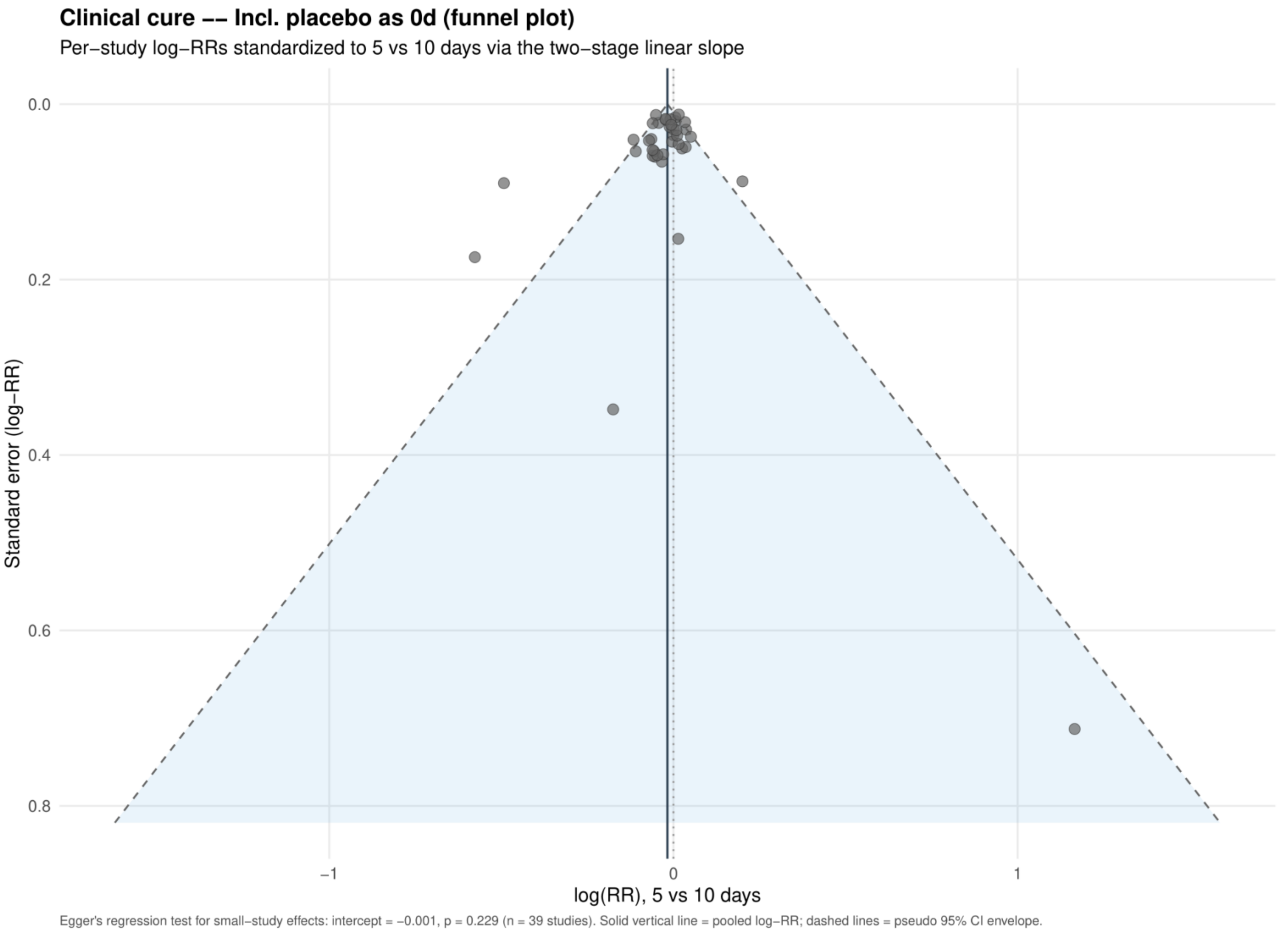


Study specific log risk ratios were standardized to 5 versus 10 days using the two stage linear slope. The solid vertical line is the pooled log risk ratio and dashed lines are pseudo 95% confidence limits; Egger's test is reported below the plot.

**Supplement 42. Clinical relapse: dose-response analysis for the primary analysis**

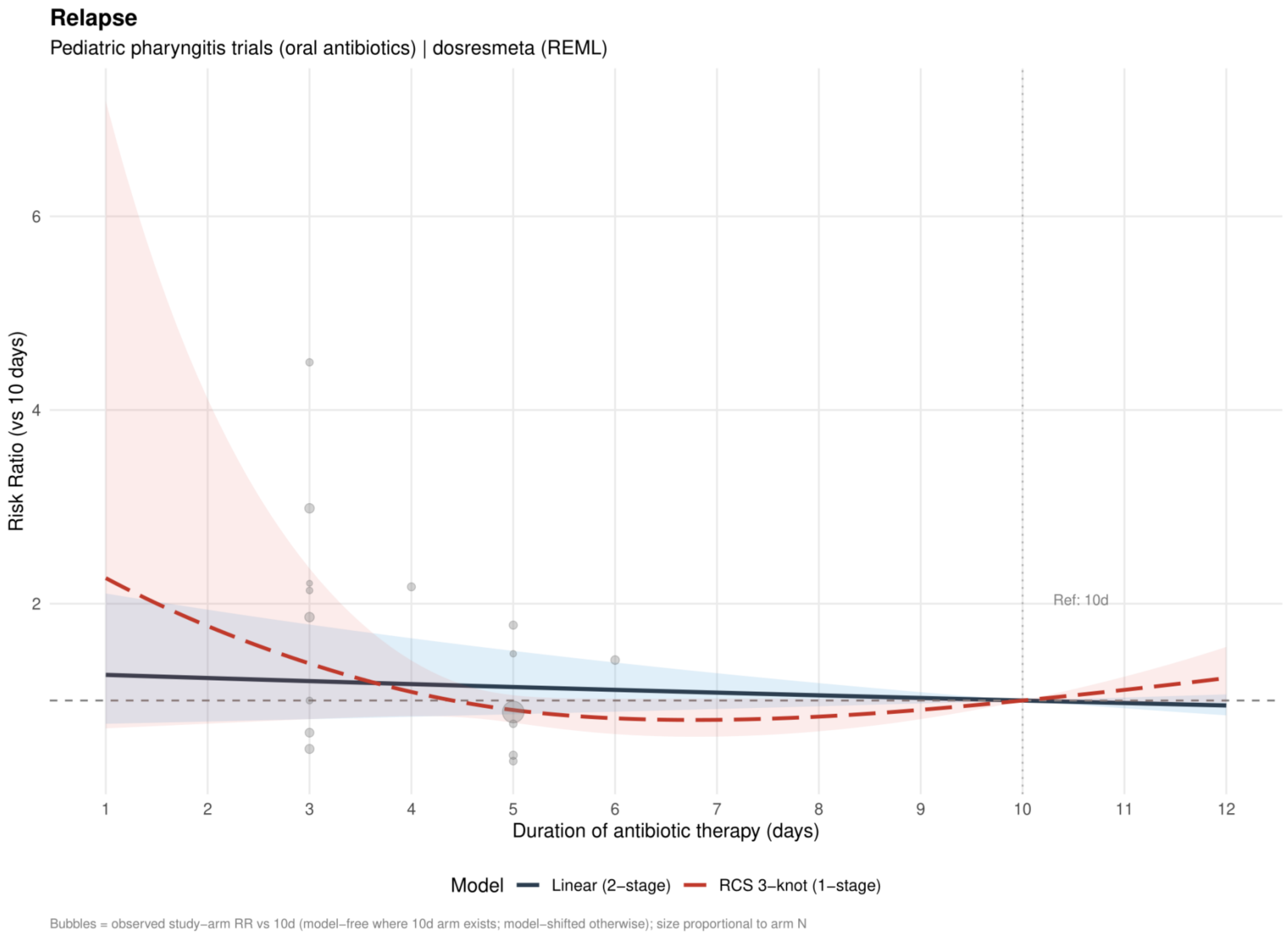


Curves show risk ratios relative to 10 days. The solid slate curve is the two stage linear model and, when present, the dashed red curve is the one stage restricted cubic spline with three knots; shaded bands show 95% confidence intervals. Bubbles show observed study arm effects and are scaled by arm sample size.

**Supplement 43. Clinical relapse: linear estimates by risk of bias for the primary analysis**

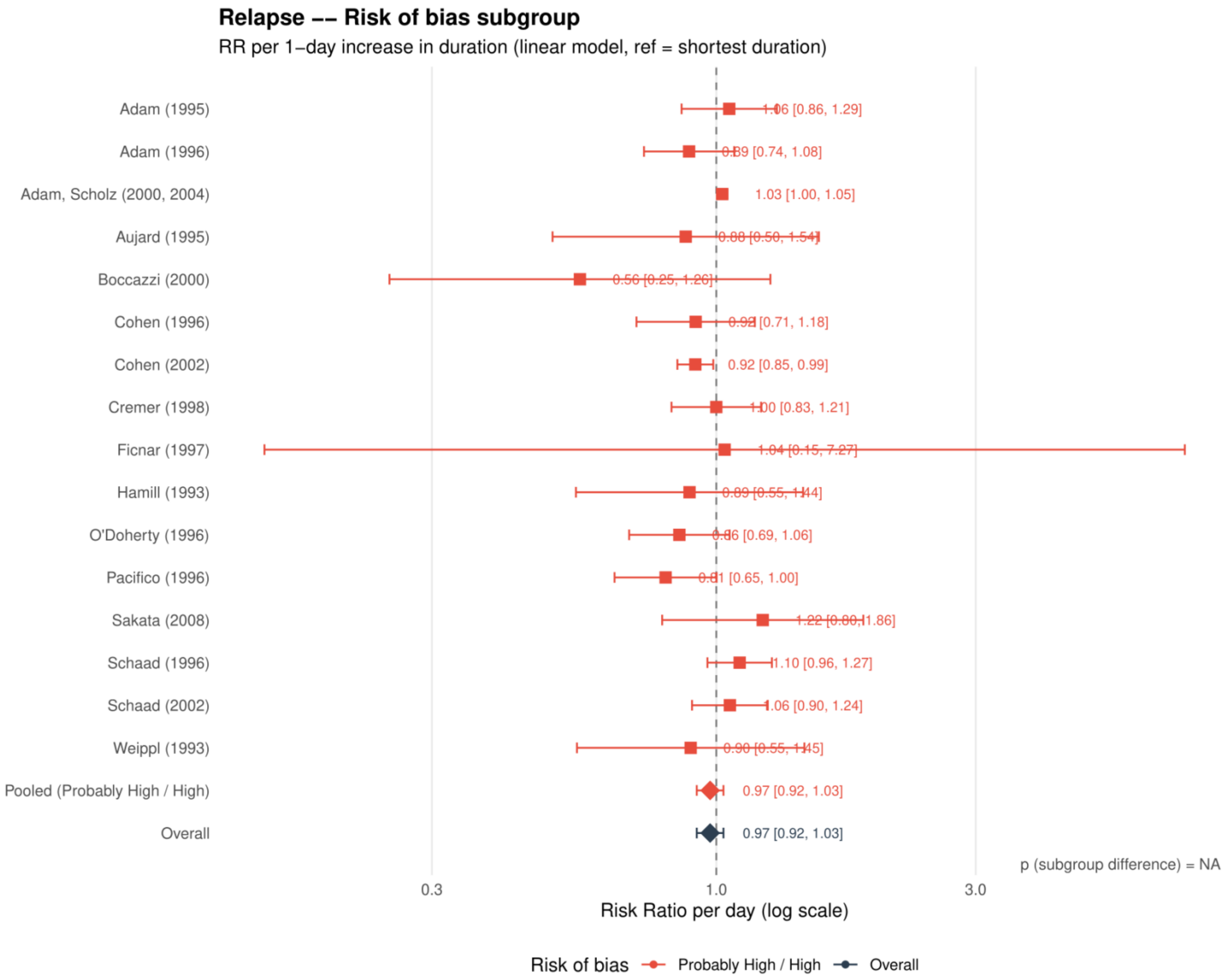


Study specific and pooled estimates are risk ratios per 1 day increase in antibiotic duration, with 95% confidence intervals.
*Only one collapsed risk of bias category contributed; the subgroup difference was not estimable.*

**Supplement 44. Clinical relapse: restricted cubic spline estimates by risk of bias for the primary analysis**

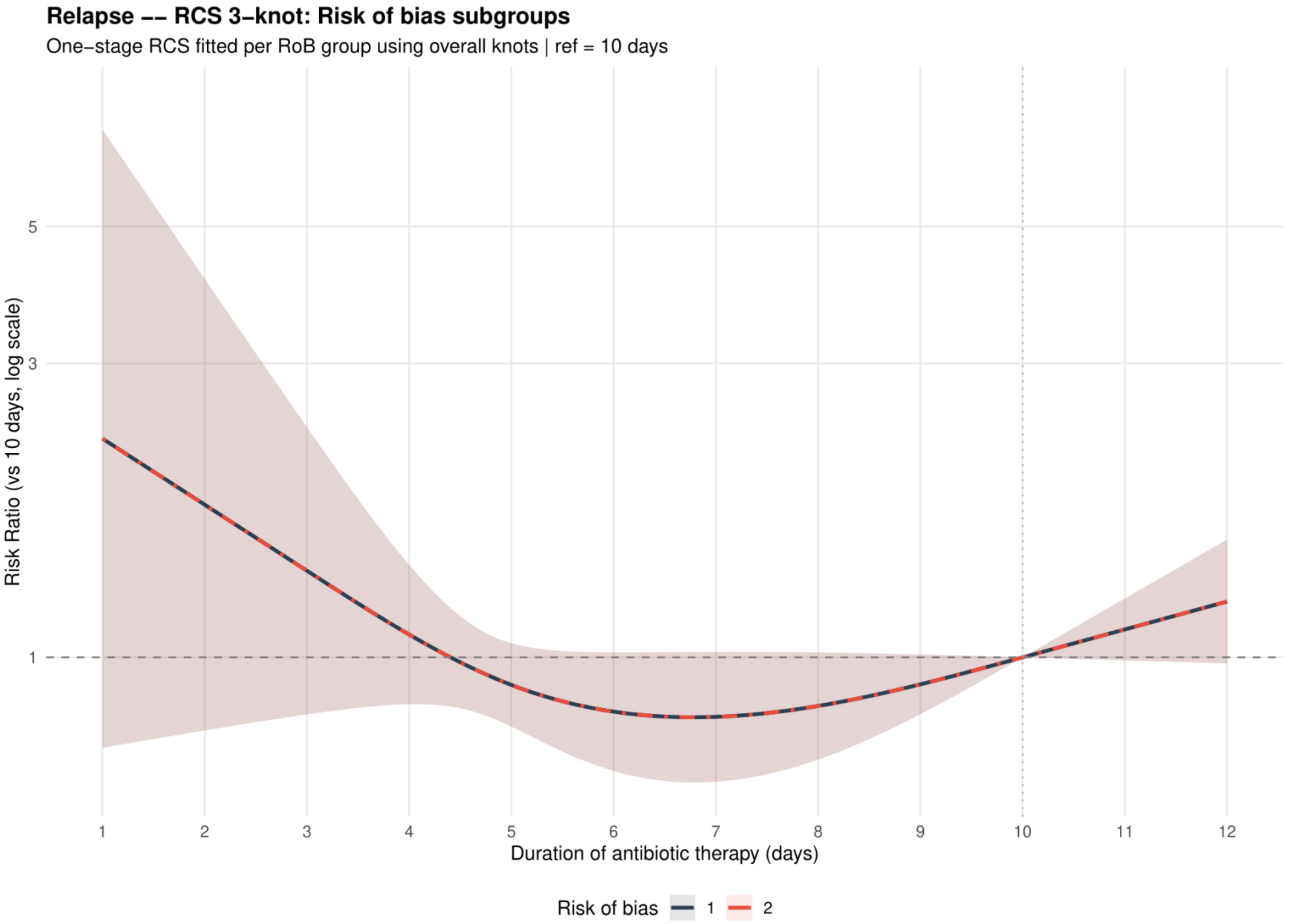


Curves show risk ratios relative to 10 days with 95% confidence intervals. Risk of bias codes: 1 = overall; 2 = low or probably low; 3 = probably high or high.

**Supplement 45. Clinical relapse: funnel plot for the primary analysis**

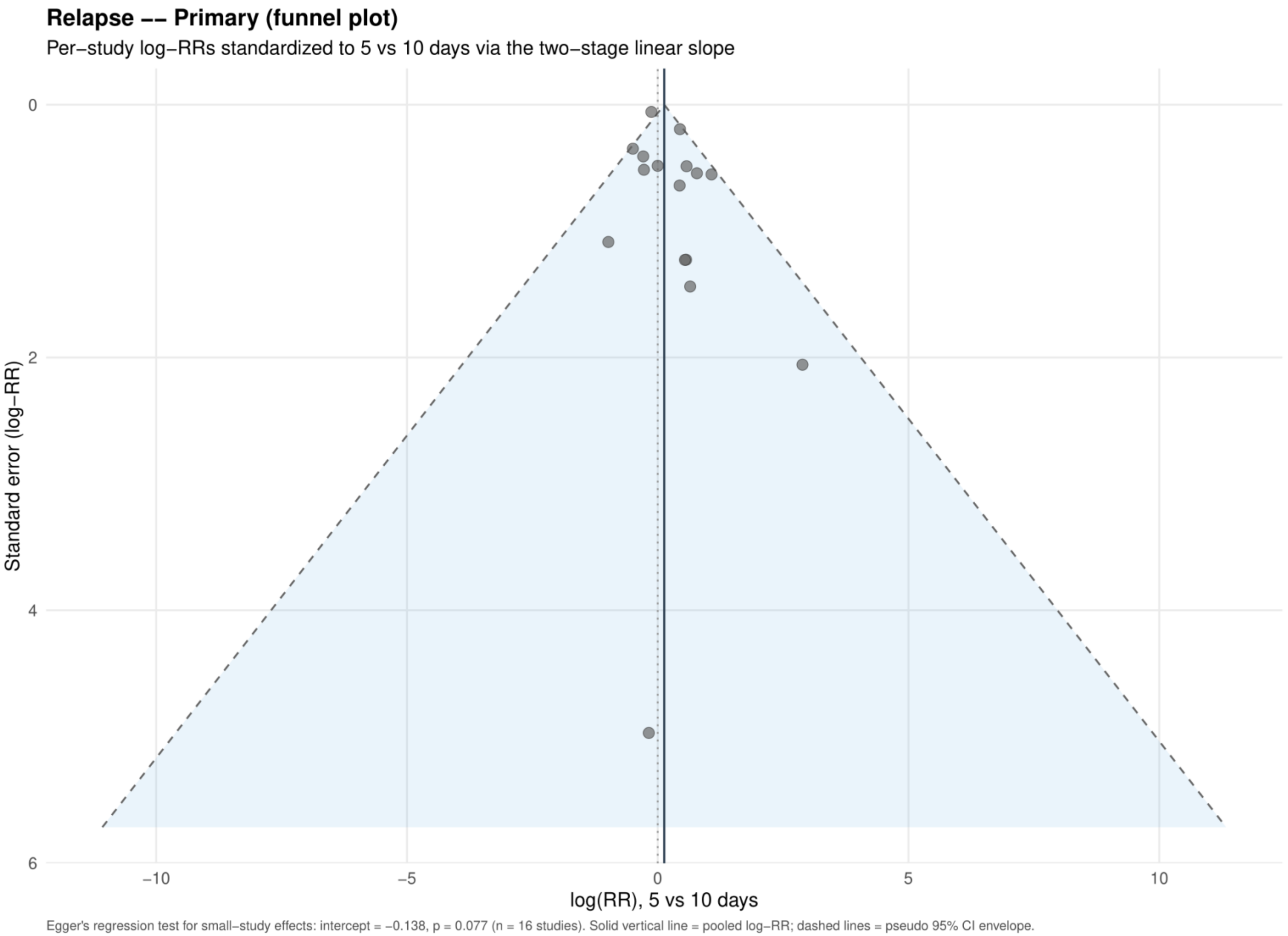


Study specific log risk ratios were standardized to 5 versus 10 days using the two stage linear slope. The solid vertical line is the pooled log risk ratio and dashed lines are pseudo 95% confidence limits; Egger's test is reported below the plot.

**Supplement 46. Clinical relapse: dose-response analysis restricted to the same antibiotic**

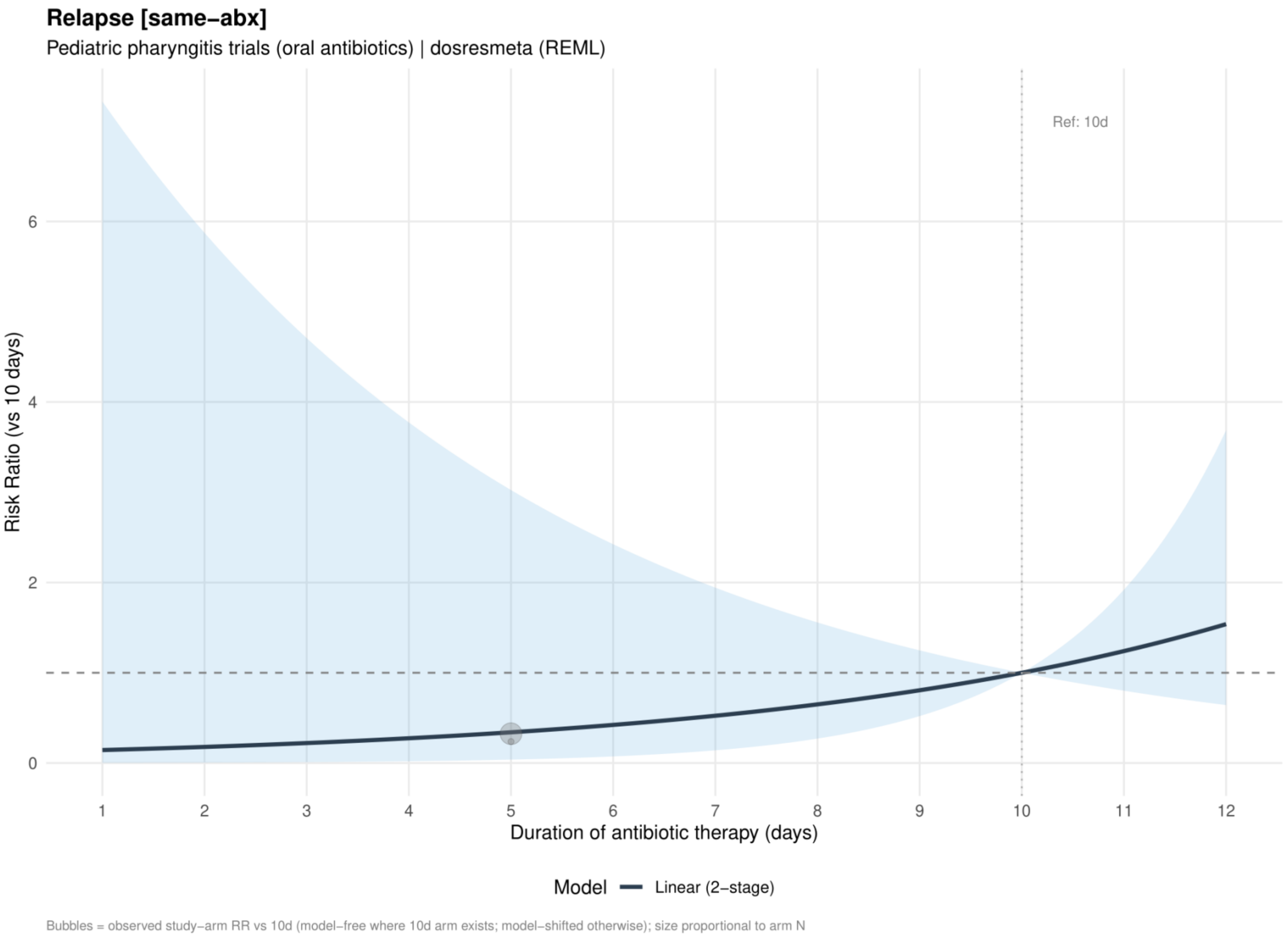


Curves show risk ratios relative to 10 days. The solid slate curve is the two stage linear model and, when present, the dashed red curve is the one stage restricted cubic spline with three knots; shaded bands show 95% confidence intervals. Bubbles show observed study arm effects and are scaled by arm sample size.

**Supplement 47. Clinical relapse: dose-response analysis restricted to the same antibiotic class**

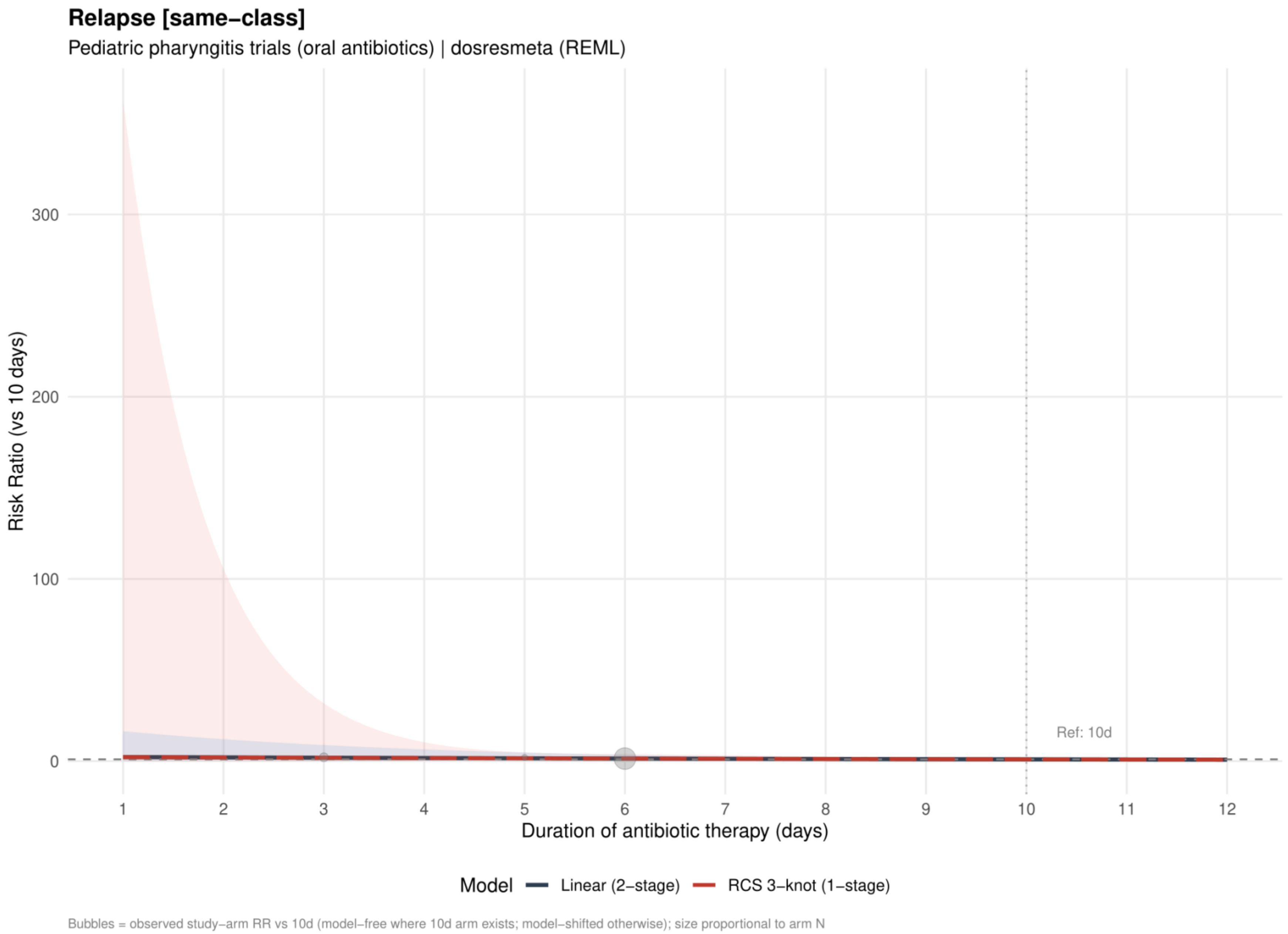


Curves show risk ratios relative to 10 days. The solid slate curve is the two stage linear model and, when present, the dashed red curve is the one stage restricted cubic spline with three knots; shaded bands show 95% confidence intervals. Bubbles show observed study arm effects and are scaled by arm sample size.

**Supplement 48. Clinical relapse: linear estimates by risk of bias restricted to the same antibiotic class**

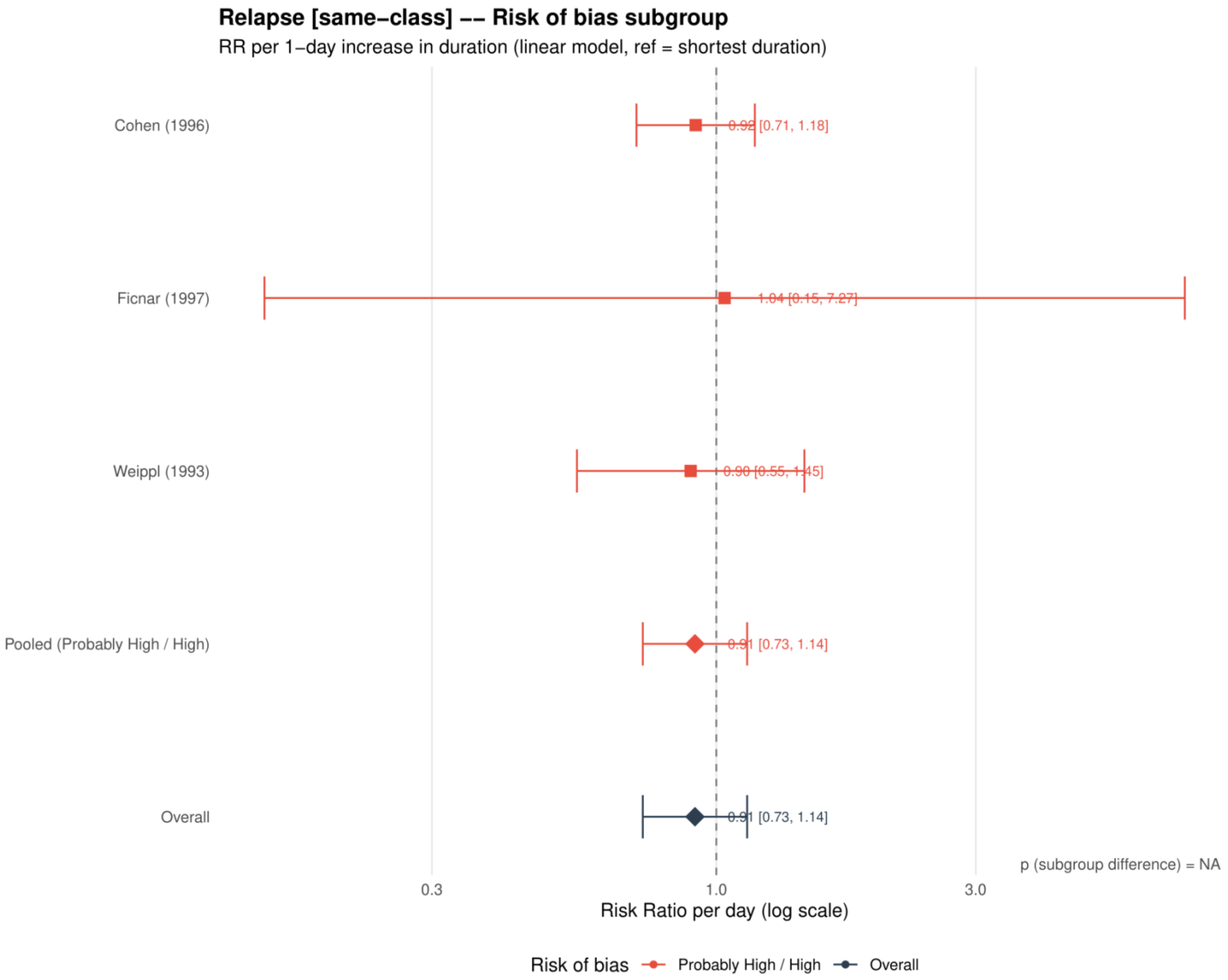


Study specific and pooled estimates are risk ratios per 1 day increase in antibiotic duration, with 95% confidence intervals.
*Only one collapsed risk of bias category contributed; the subgroup difference was not estimable.*

**Supplement 49. Clinical relapse: restricted cubic spline estimates by risk of bias restricted to the same antibiotic class**

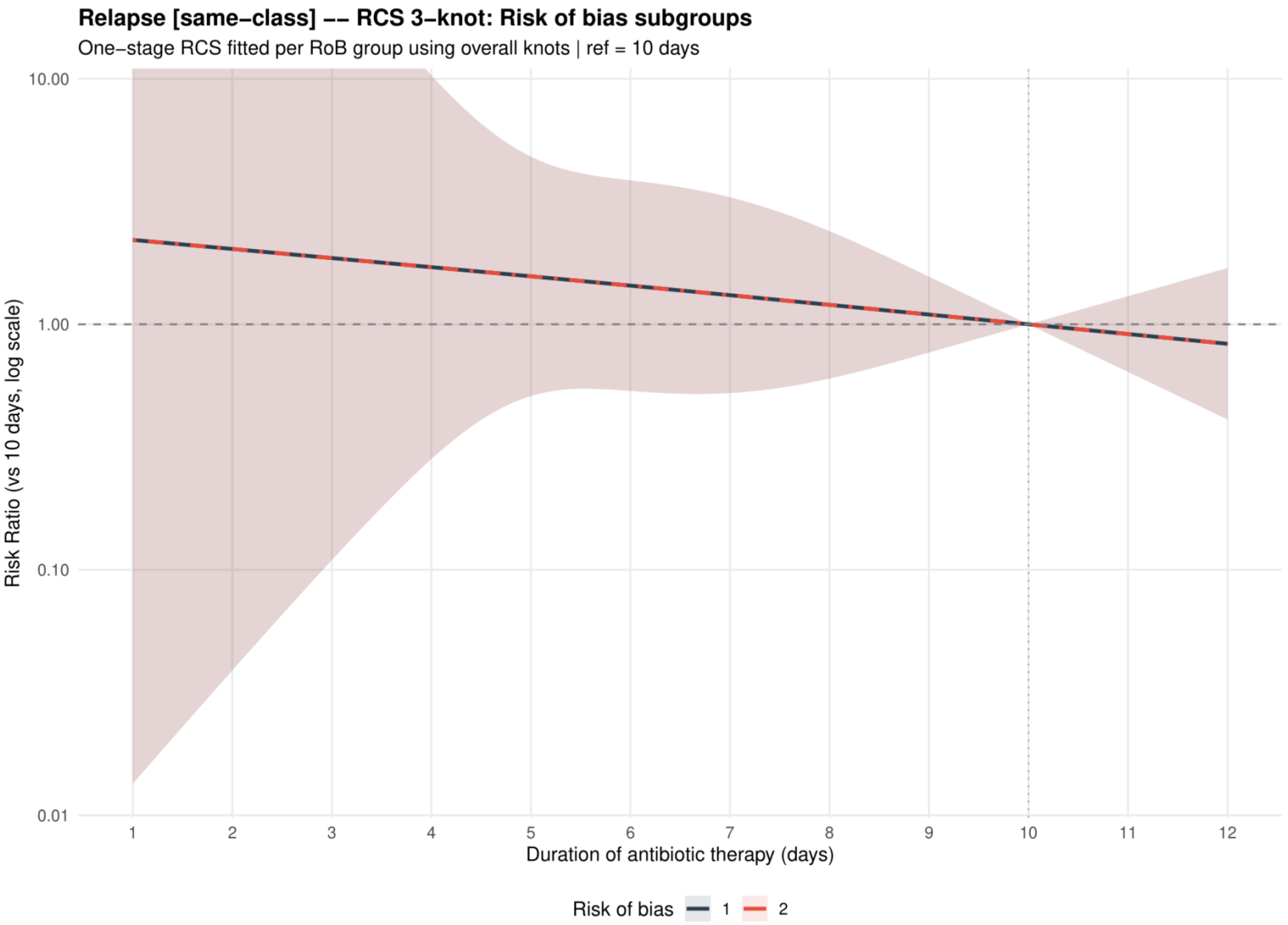


Curves show risk ratios relative to 10 days with 95% confidence intervals. Risk of bias codes: 1 = overall; 2 = low or probably low; 3 = probably high or high.

**Supplement 50. Clinical relapse: dose-response analysis excluding azithromycin**

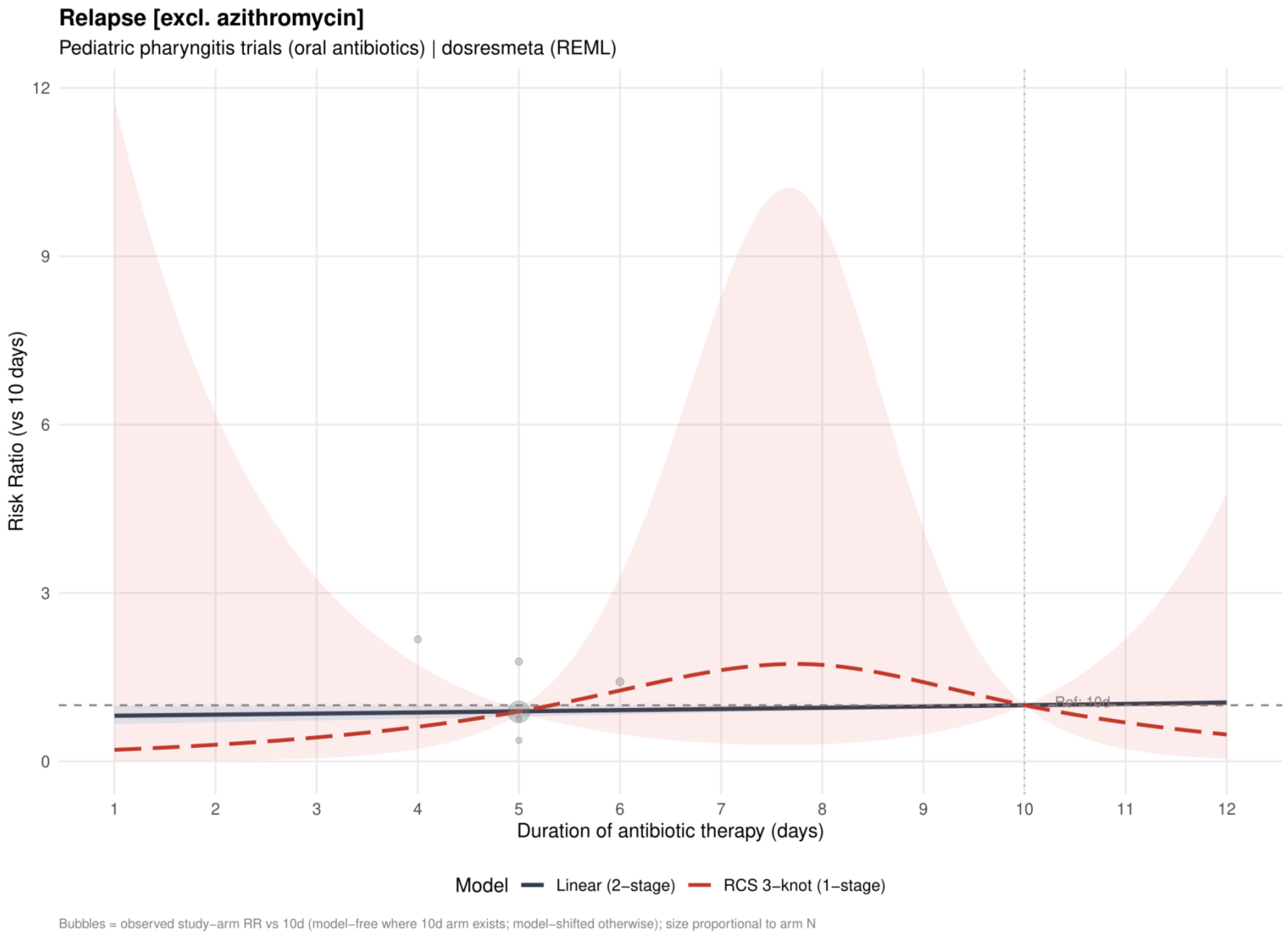


Curves show risk ratios relative to 10 days. The solid slate curve is the two stage linear model and, when present, the dashed red curve is the one stage restricted cubic spline with three knots; shaded bands show 95% confidence intervals. Bubbles show observed study arm effects and are scaled by arm sample size.

**Supplement 51. Clinical relapse: linear estimates by risk of bias excluding azithromycin**

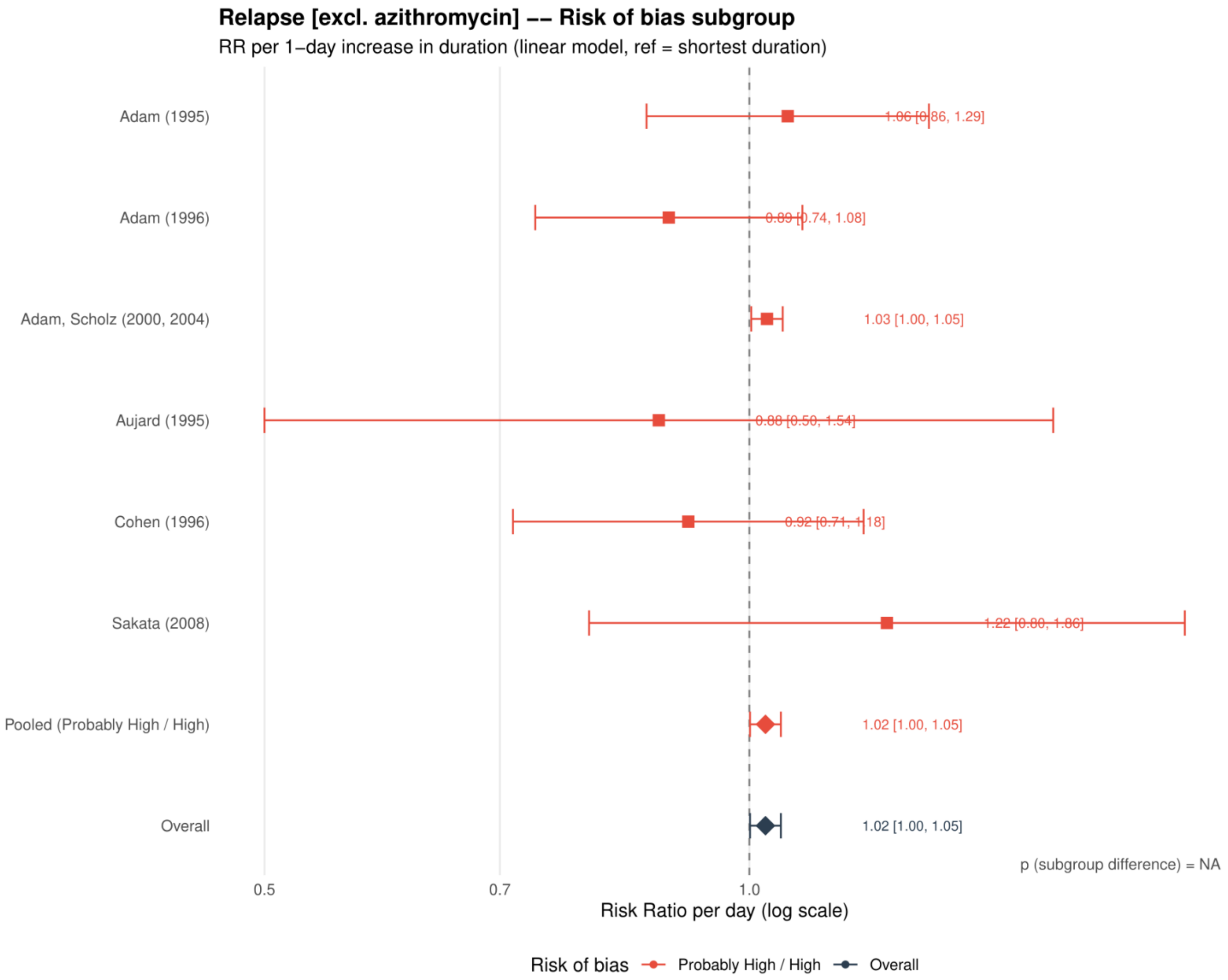


Study specific and pooled estimates are risk ratios per 1 day increase in antibiotic duration, with 95% confidence intervals.
*Only one collapsed risk of bias category contributed; the subgroup difference was not estimable.*

**Supplement 52. Clinical relapse: restricted cubic spline estimates by risk of bias excluding azithromycin**

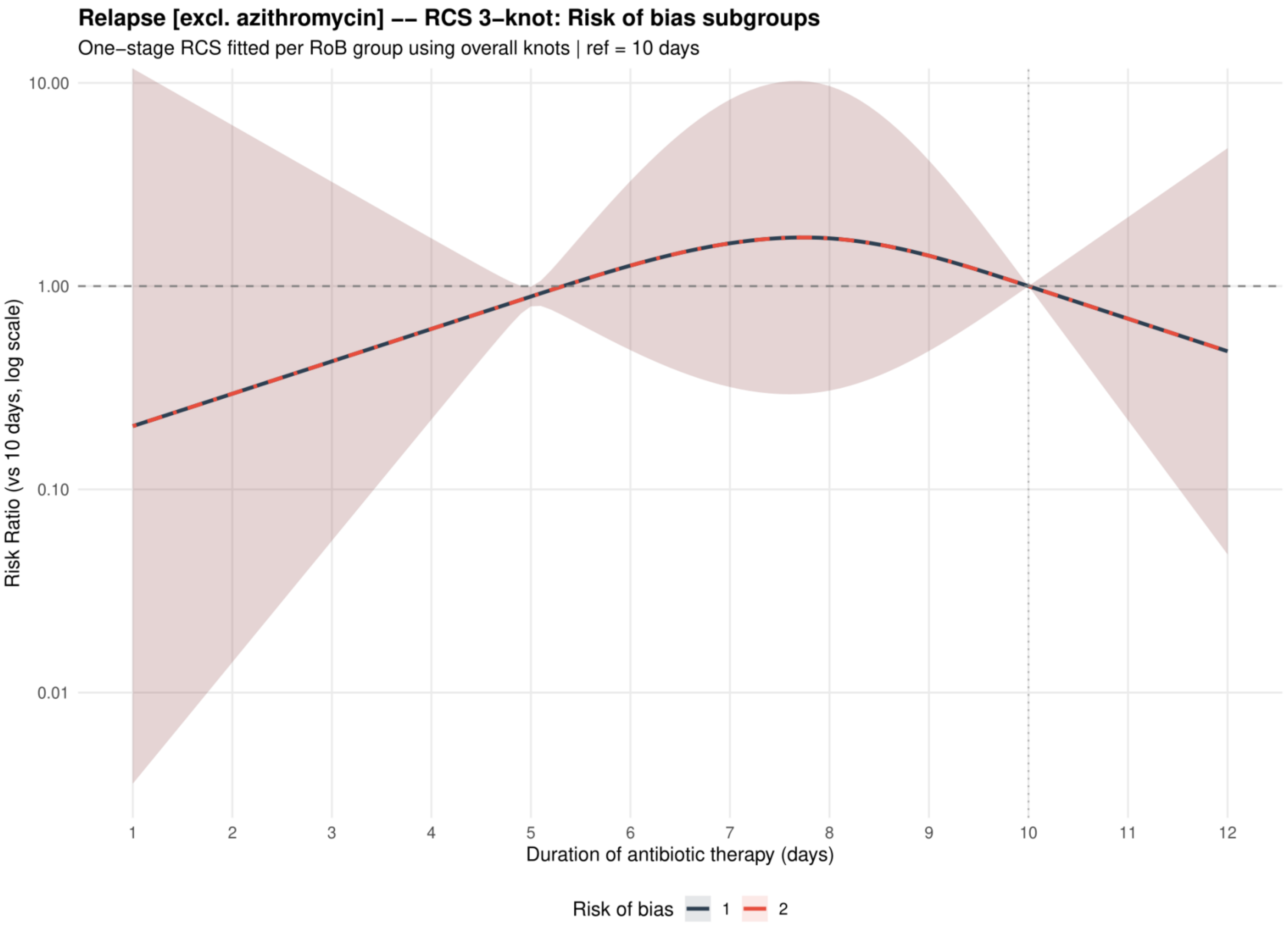


Curves show risk ratios relative to 10 days with 95% confidence intervals. Risk of bias codes: 1 = overall; 2 = low or probably low; 3 = probably high or high.

**Supplement 53. Clinical relapse: dose-response analysis excluding macrolides**

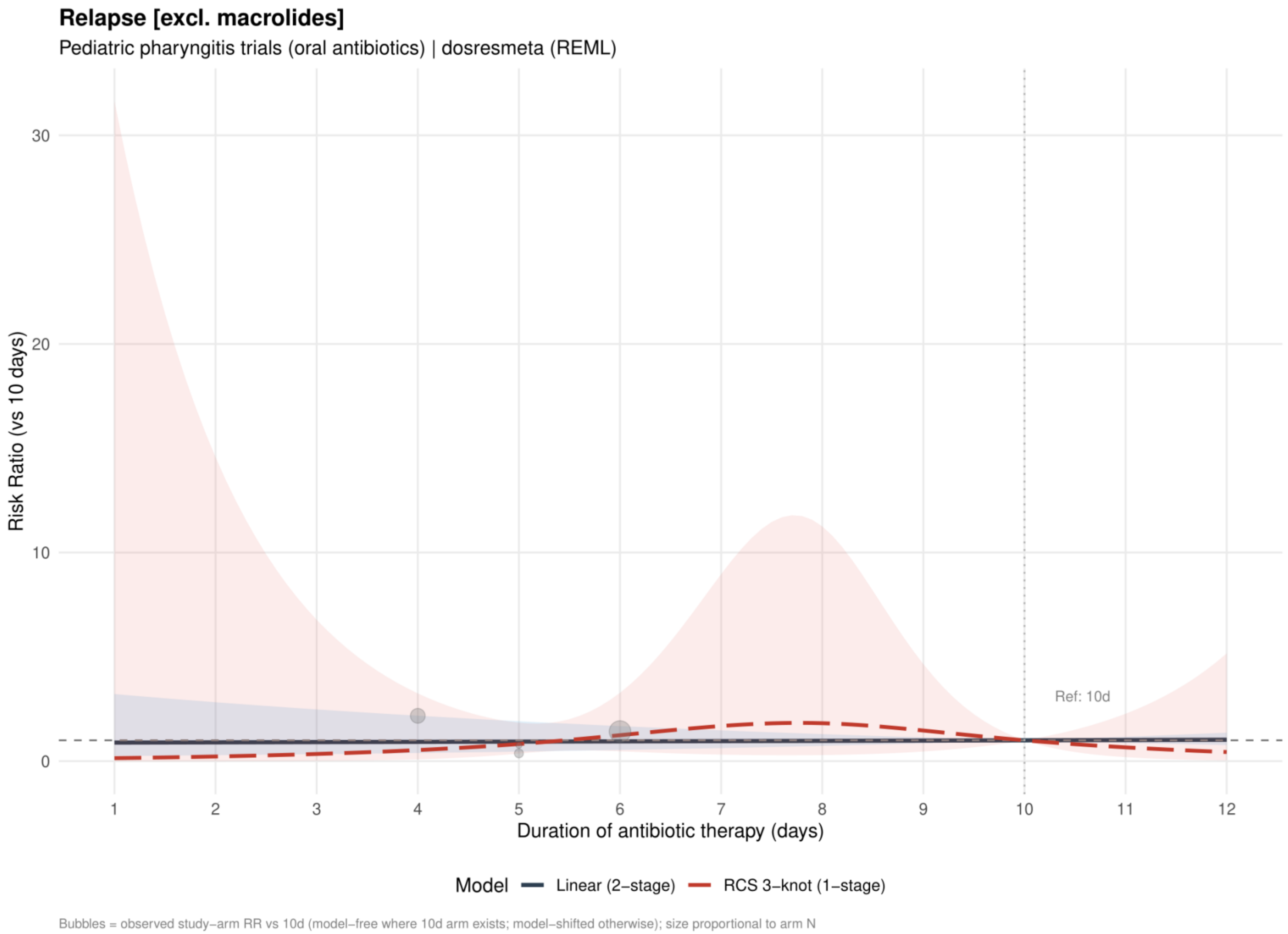


Curves show risk ratios relative to 10 days. The solid slate curve is the two stage linear model and, when present, the dashed red curve is the one stage restricted cubic spline with three knots; shaded bands show 95% confidence intervals. Bubbles show observed study arm effects and are scaled by arm sample size.

**Supplement 54. Clinical relapse: linear estimates by risk of bias excluding macrolides**

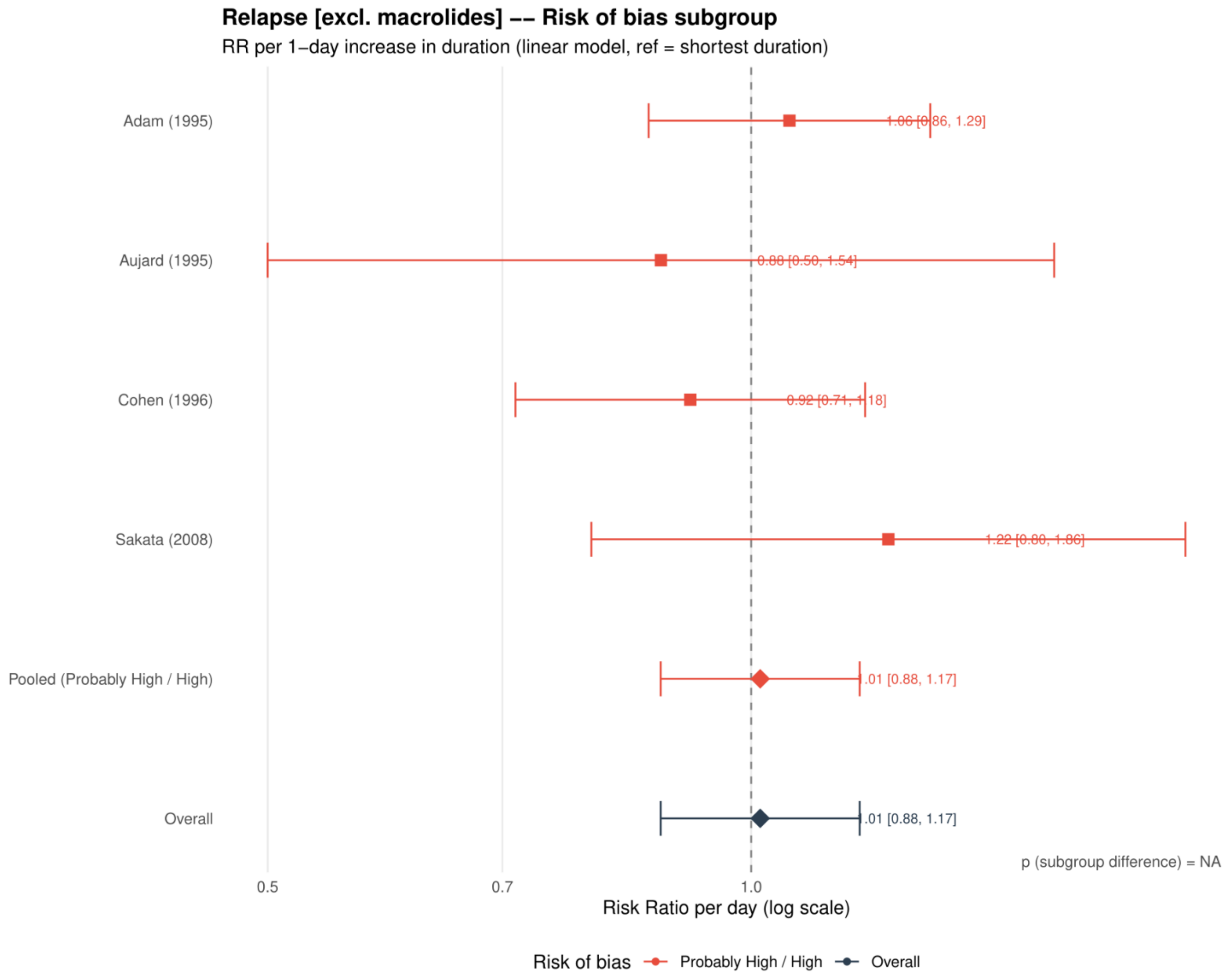


Study specific and pooled estimates are risk ratios per 1 day increase in antibiotic duration, with 95% confidence intervals.
*Only one collapsed risk of bias category contributed; the subgroup difference was not estimable.*

**Supplement 55. Clinical relapse: restricted cubic spline estimates by risk of bias excluding macrolides**

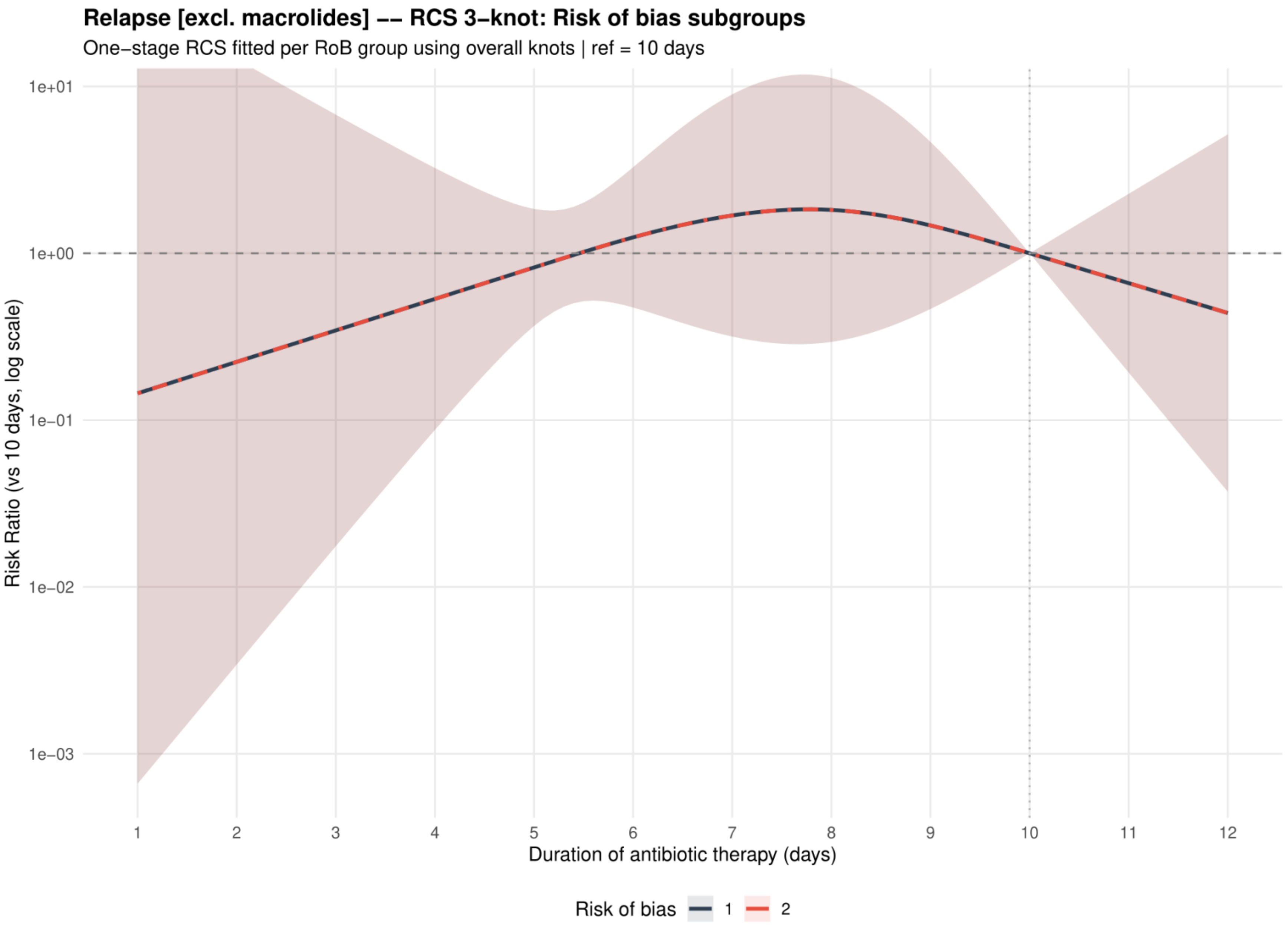


Curves show risk ratios relative to 10 days with 95% confidence intervals. Risk of bias codes: 1 = overall; 2 = low or probably low; 3 = probably high or high.

**Supplement 56. Clinical relapse: dose-response analysis including placebo or no treatment as 0 days**

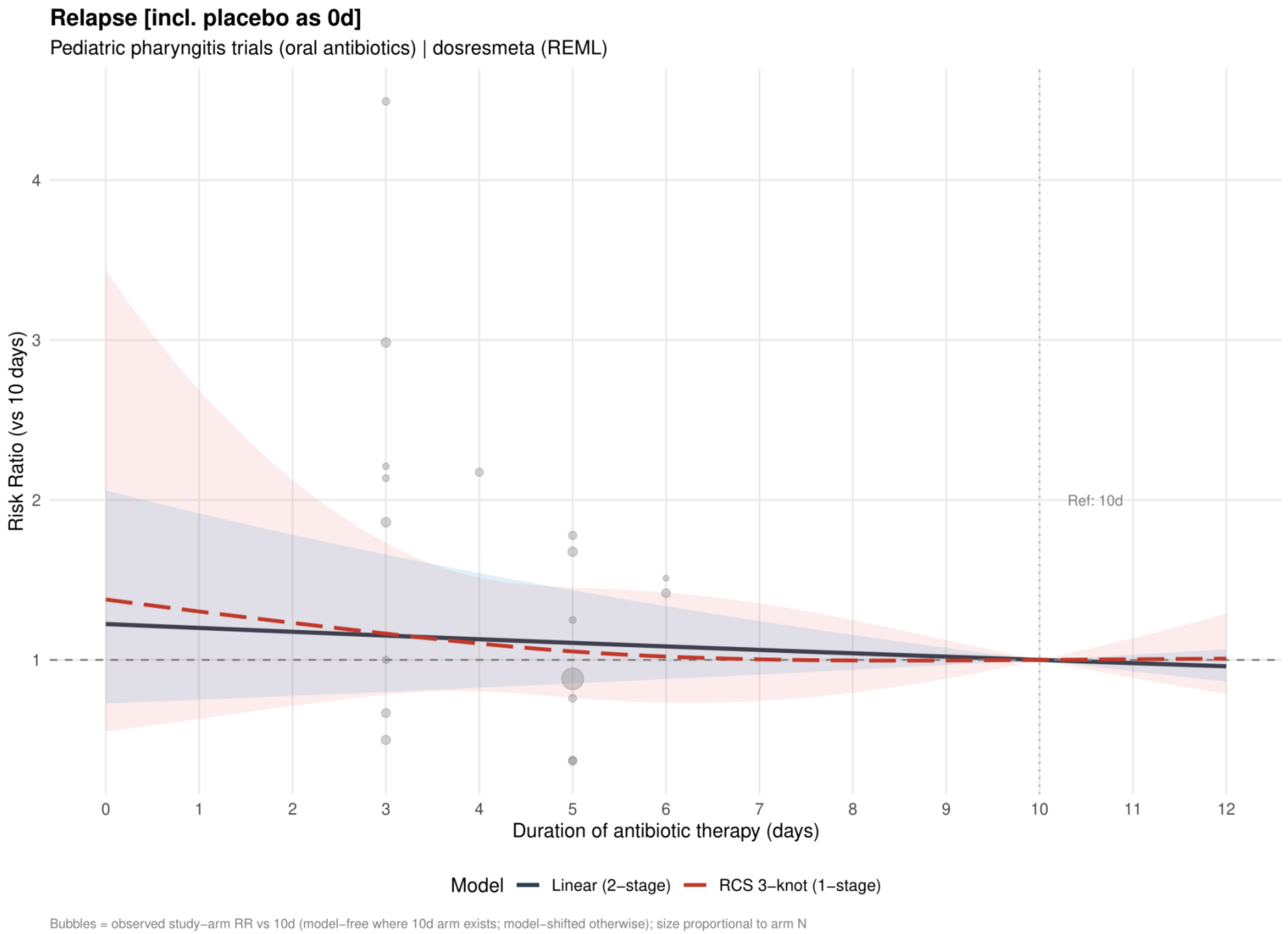


Curves show risk ratios relative to 10 days. The solid slate curve is the two stage linear model and, when present, the dashed red curve is the one stage restricted cubic spline with three knots; shaded bands show 95% confidence intervals. Bubbles show observed study arm effects and are scaled by arm sample size.

**Supplement 57. Clinical relapse: linear estimates by risk of bias including placebo or no treatment as 0 days**

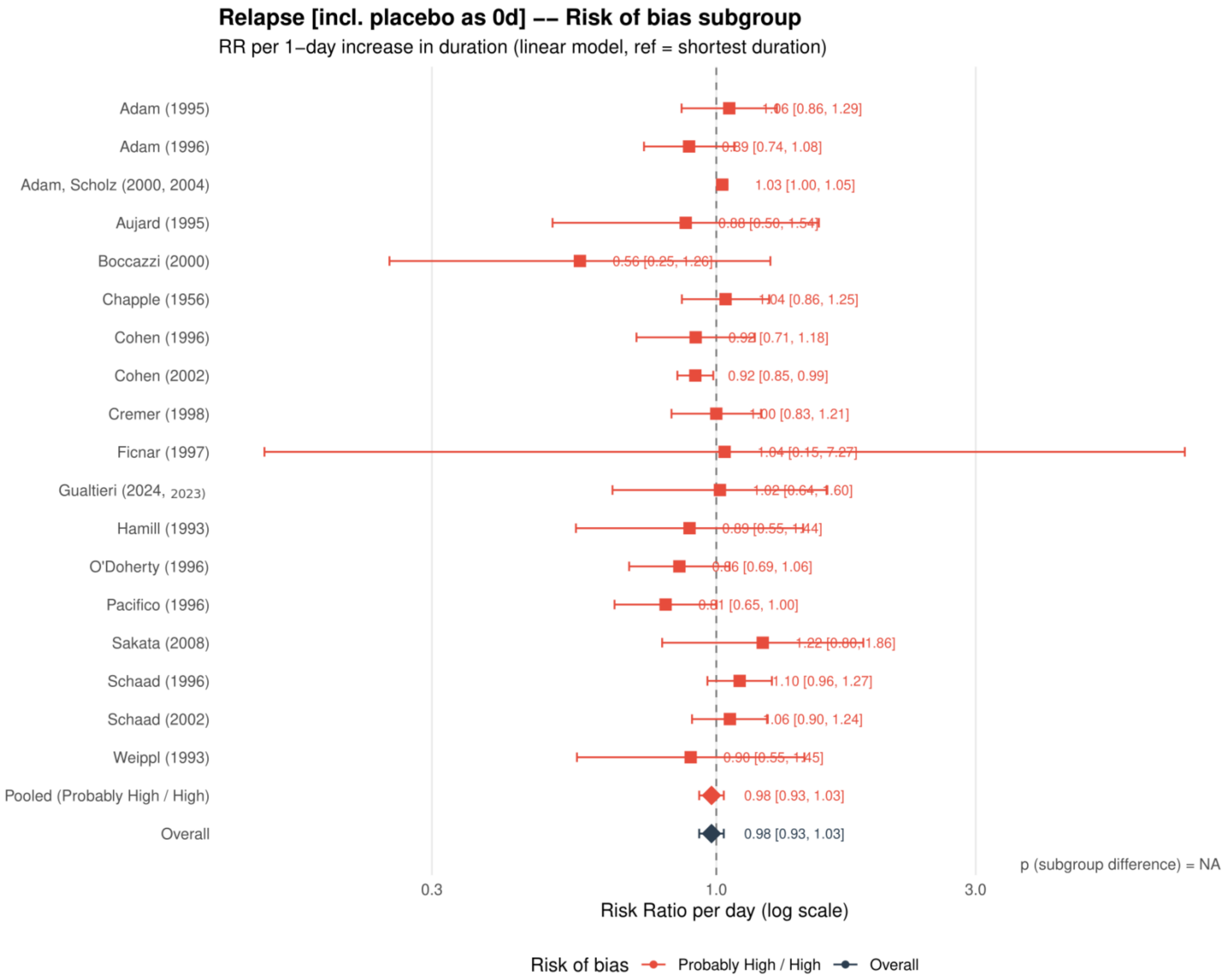


Study specific and pooled estimates are risk ratios per 1 day increase in antibiotic duration, with 95% confidence intervals.
*Only one collapsed risk of bias category contributed; the subgroup difference was not estimable.*

**Supplement 58. Clinical relapse: restricted cubic spline estimates by risk of bias including placebo or no treatment as 0 days**

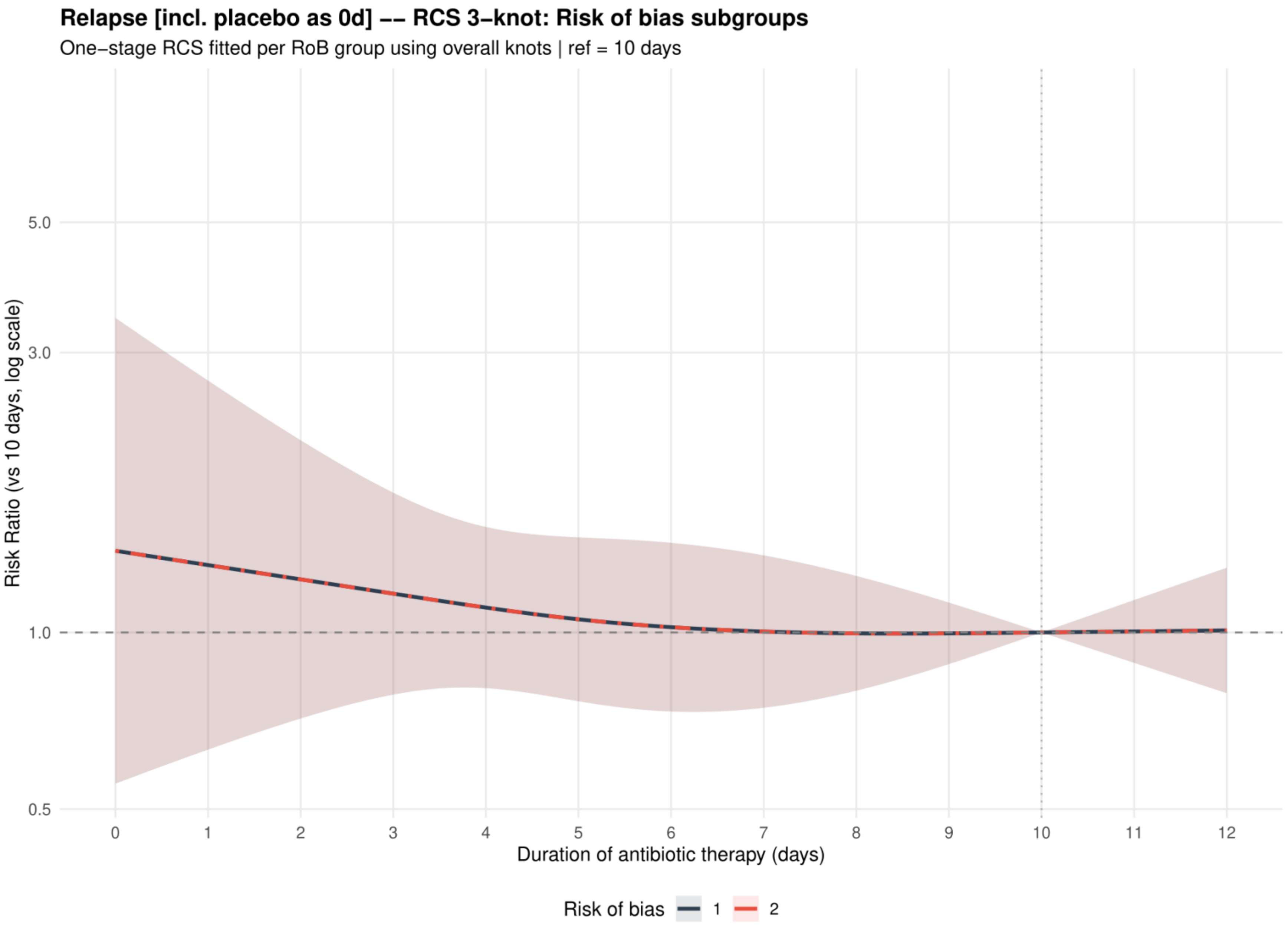


Curves show risk ratios relative to 10 days with 95% confidence intervals. Risk of bias codes: 1 = overall; 2 = low or probably low; 3 = probably high or high.

**Supplement 59. Clinical relapse: funnel plot including placebo or no treatment as 0 days**

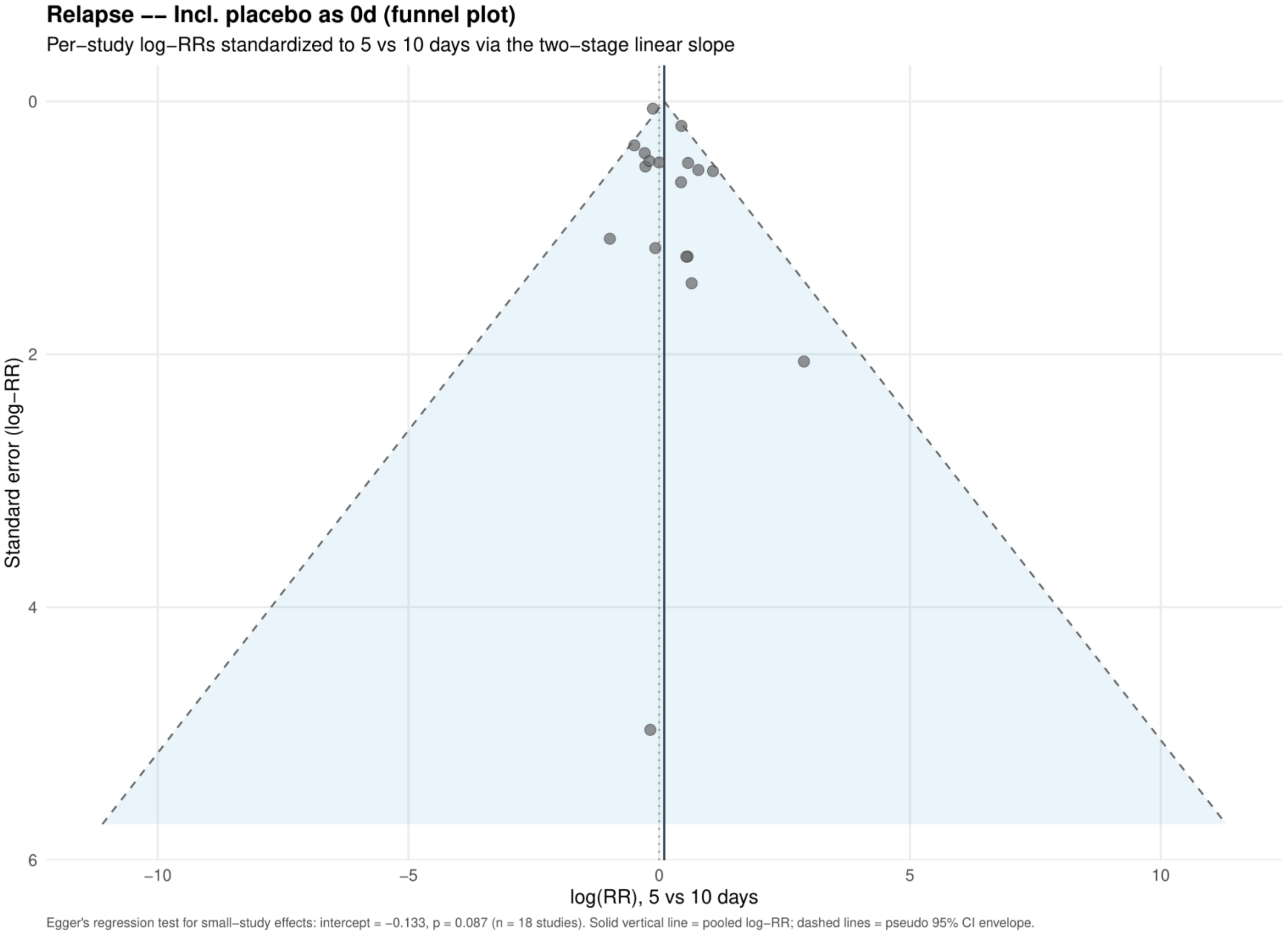


Study specific log risk ratios were standardized to 5 versus 10 days using the two stage linear slope. The solid vertical line is the pooled log risk ratio and dashed lines are pseudo 95% confidence limits; Egger's test is reported below the plot.

**Supplement 60. Adverse events: dose-response analysis for the primary analysis**

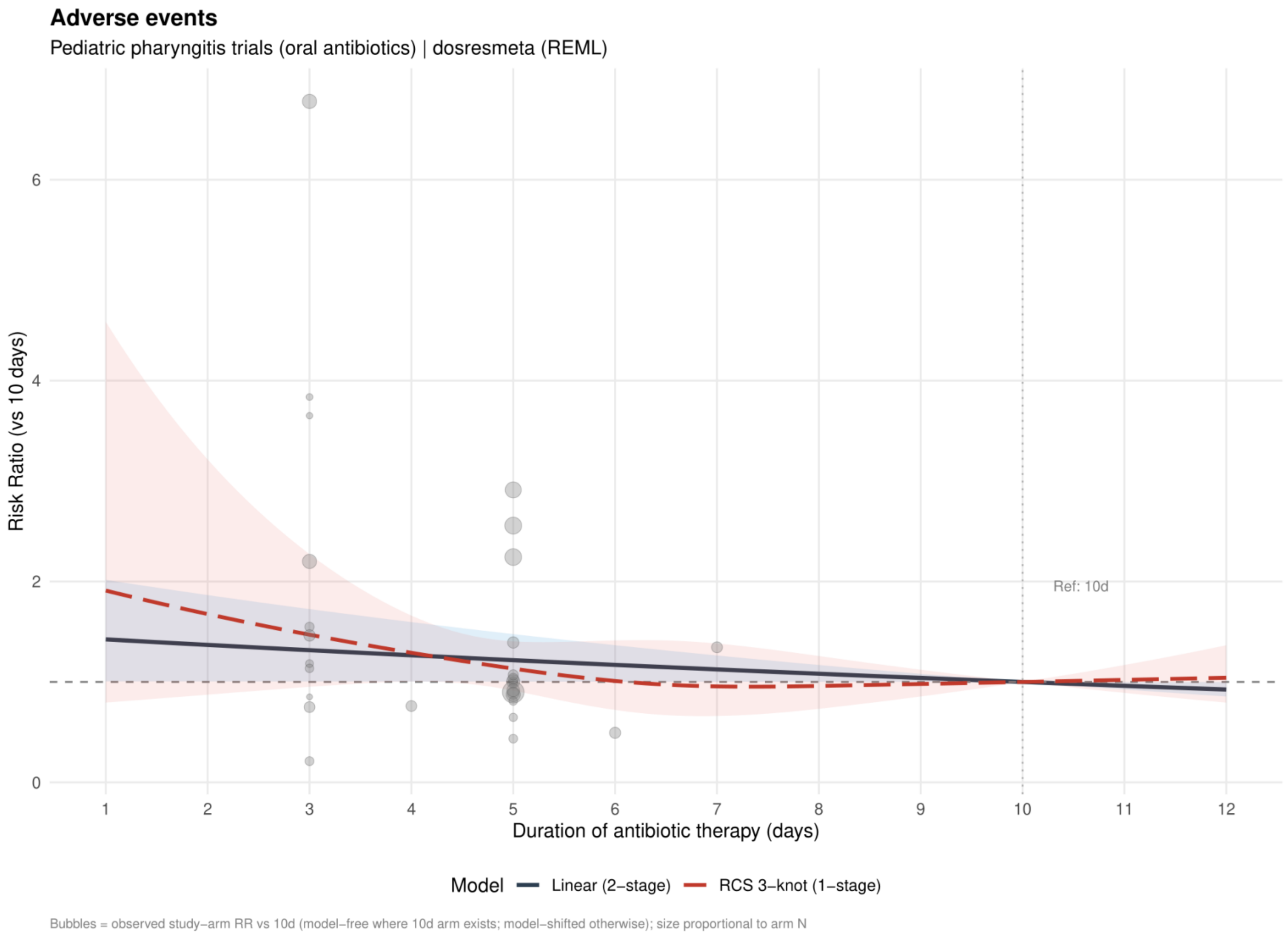


Curves show risk ratios relative to 10 days. The solid slate curve is the two stage linear model and, when present, the dashed red curve is the one stage restricted cubic spline with three knots; shaded bands show 95% confidence intervals. Bubbles show observed study arm effects and are scaled by arm sample size.

**Supplement 61. Adverse events: linear estimates by risk of bias for the primary analysis**

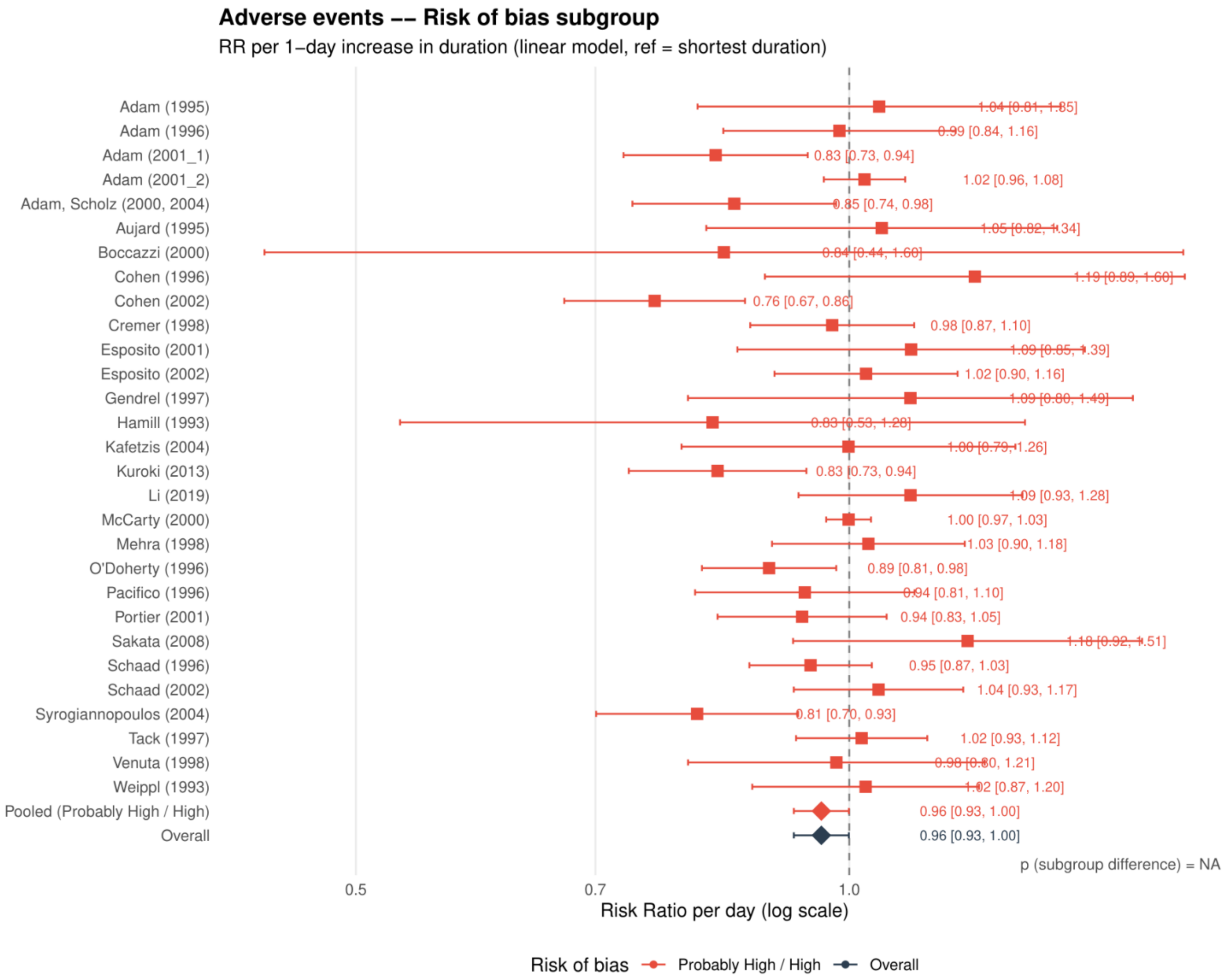


Study specific and pooled estimates are risk ratios per 1 day increase in antibiotic duration, with 95% confidence intervals.
*Only one collapsed risk of bias category contributed; the subgroup difference was not estimable.*

**Supplement 62. Adverse events: restricted cubic spline estimates by risk of bias for the primary analysis**

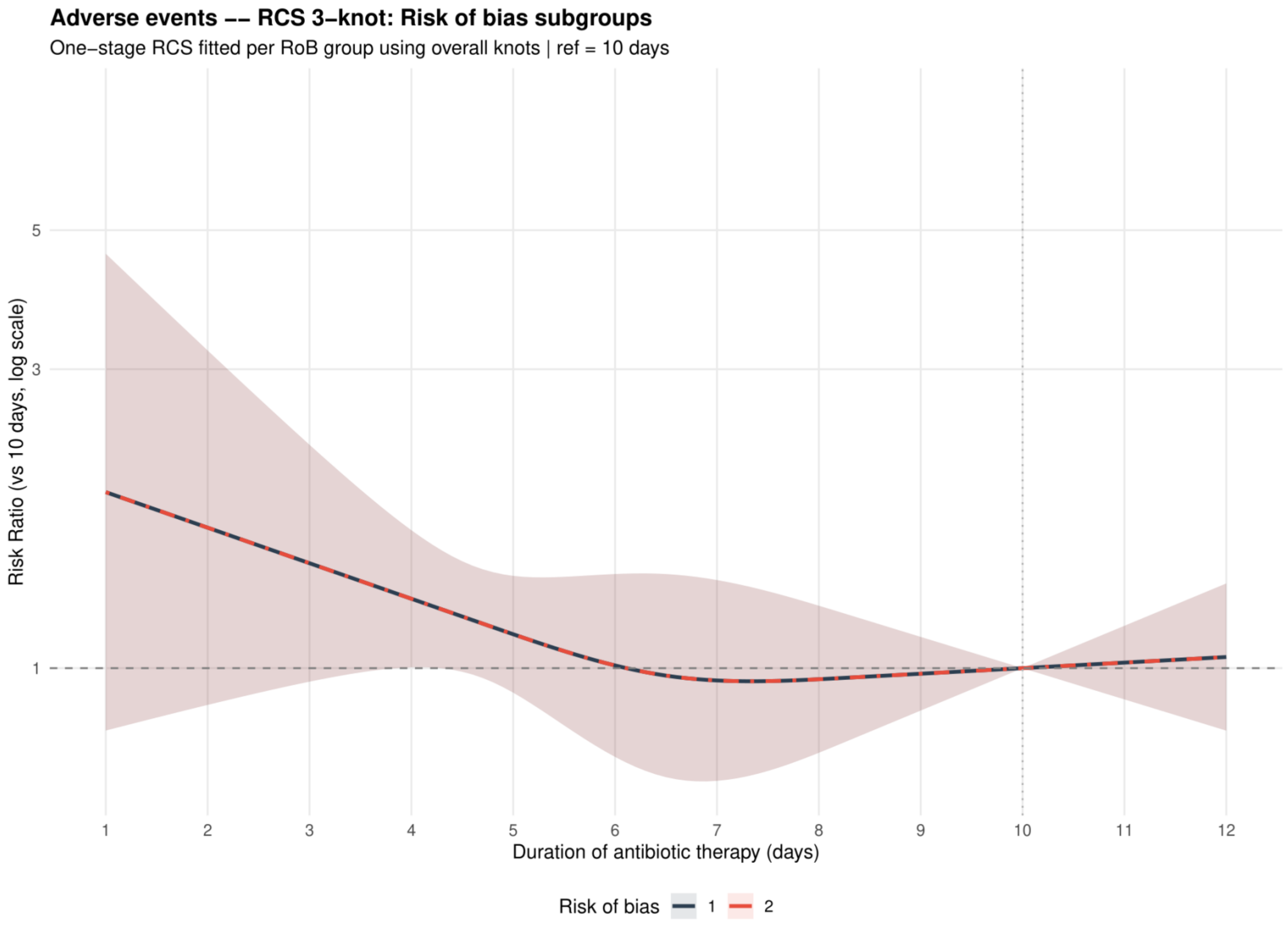


Curves show risk ratios relative to 10 days with 95% confidence intervals. Risk of bias codes: 1 = overall; 2 = low or probably low; 3 = probably high or high.

**Supplement 63. Adverse events: funnel plot for the primary analysis**

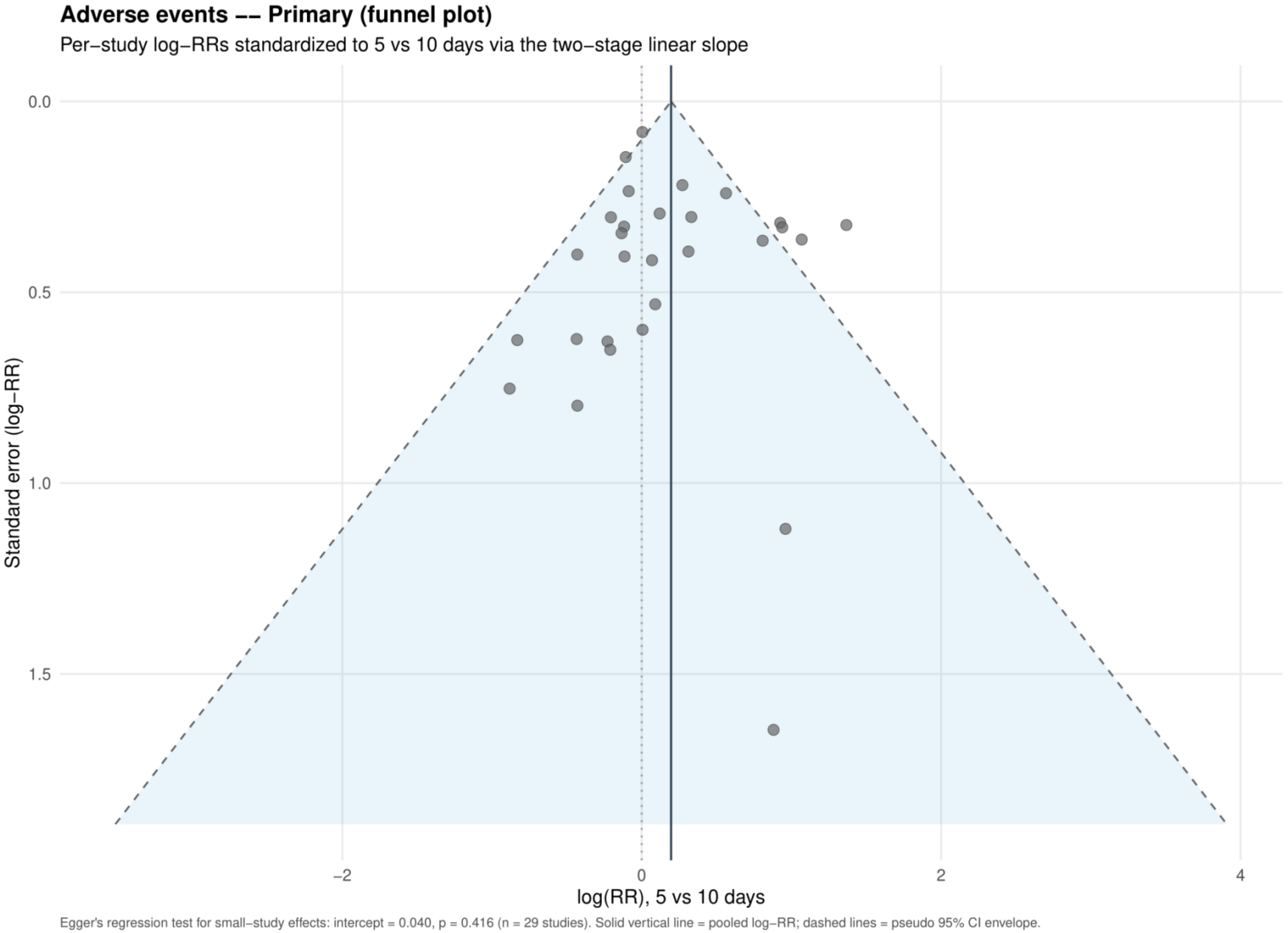


Study specific log risk ratios were standardized to 5 versus 10 days using the two stage linear slope. The solid vertical line is the pooled log risk ratio and dashed lines are pseudo 95% confidence limits; Egger's test is reported below the plot.

**Supplement 64. Adverse events: dose-response analysis restricted to the same antibiotic and dose**

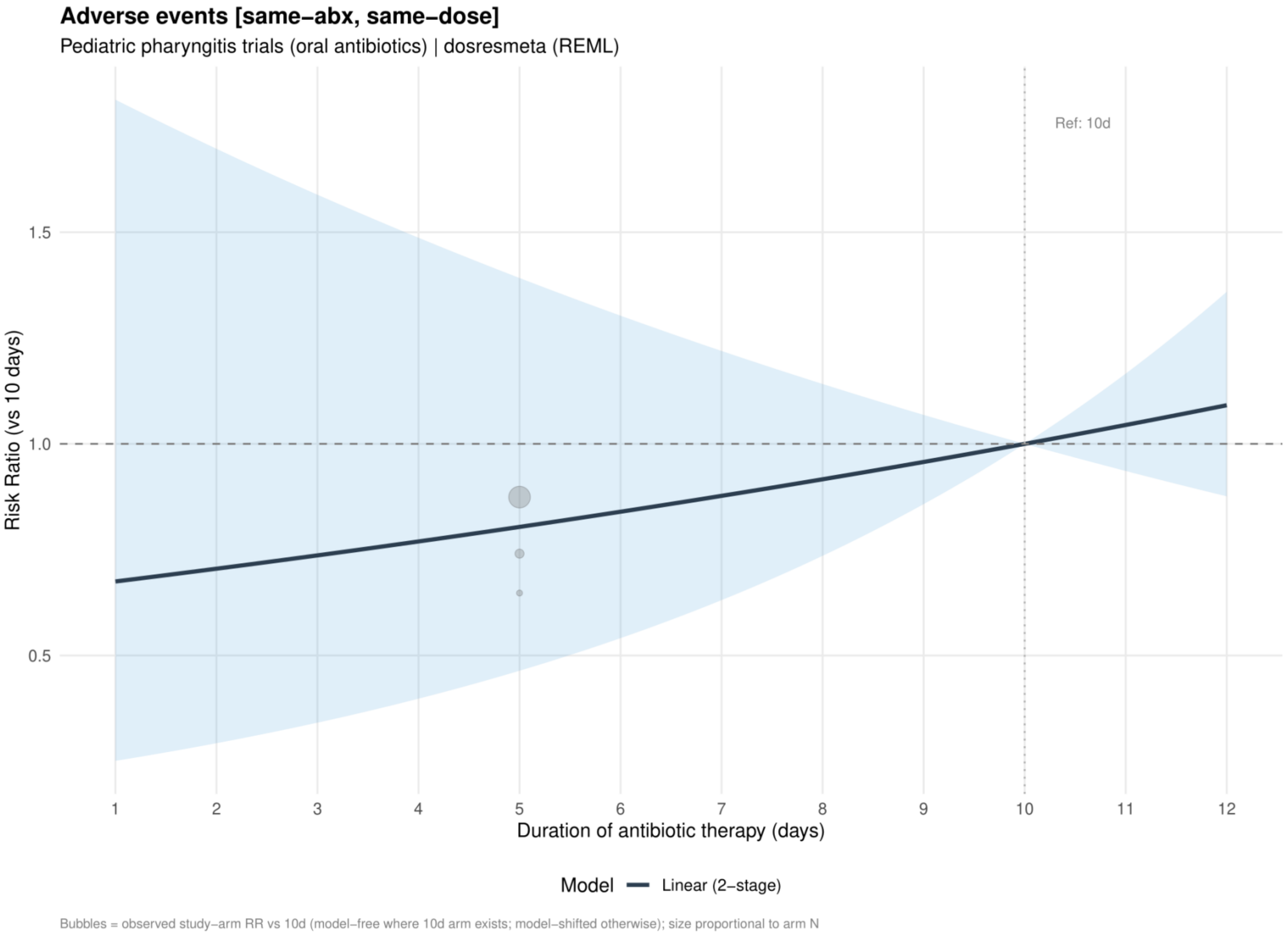


Curves show risk ratios relative to 10 days. The solid slate curve is the two stage linear model and, when present, the dashed red curve is the one stage restricted cubic spline with three knots; shaded bands show 95% confidence intervals. Bubbles show observed study arm effects and are scaled by arm sample size.

*All same-antibiotic comparisons reporting adverse events used the same daily dose; this analysis is identical to Supplement 65.*

**Supplement 65. Adverse events: dose-response analysis restricted to the same antibiotic**

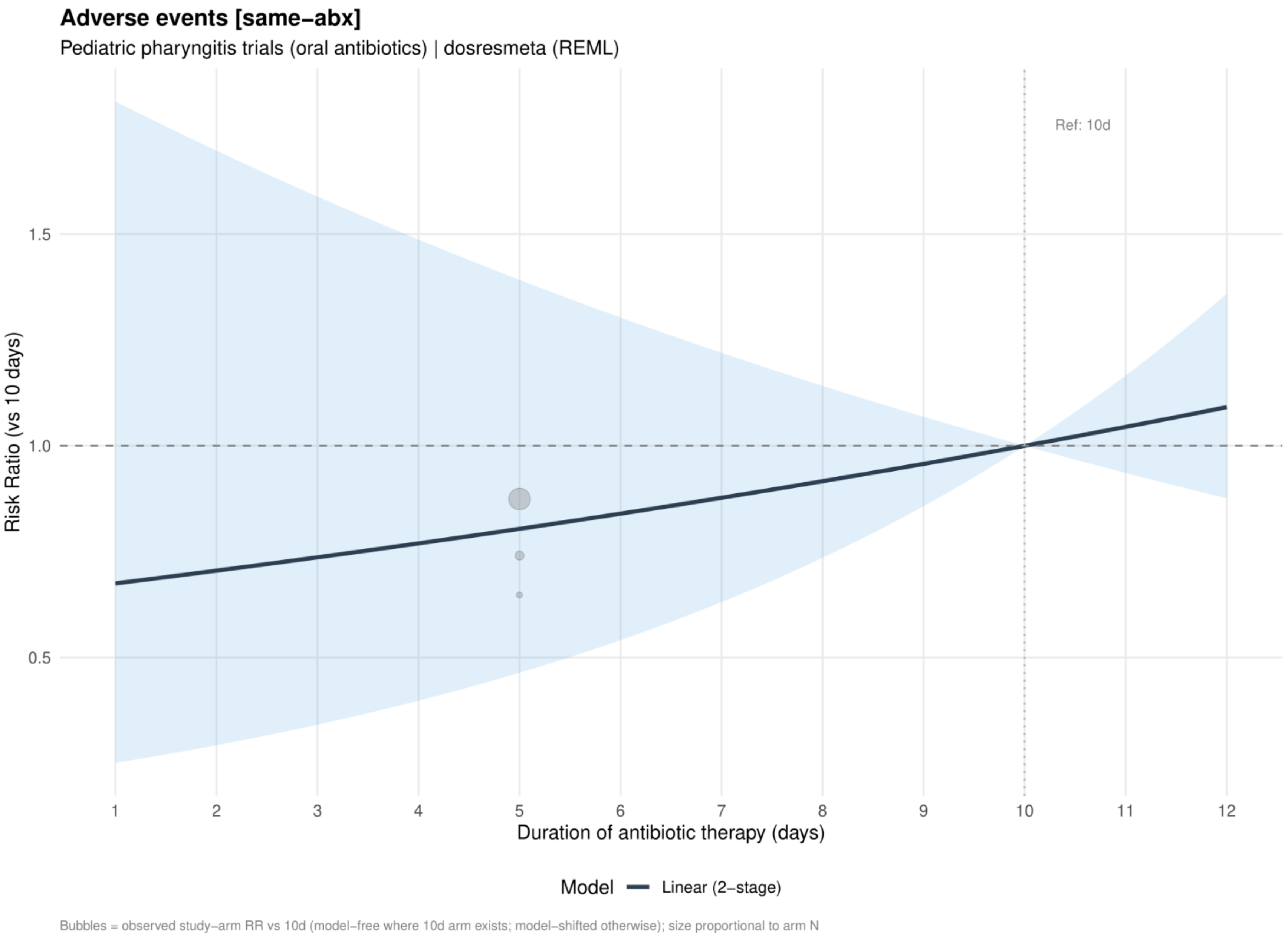


Curves show risk ratios relative to 10 days. The solid slate curve is the two stage linear model and, when present, the dashed red curve is the one stage restricted cubic spline with three knots; shaded bands show 95% confidence intervals. Bubbles show observed study arm effects and are scaled by arm sample size.

**Supplement 66. Adverse events: dose-response analysis restricted to the same antibiotic class**

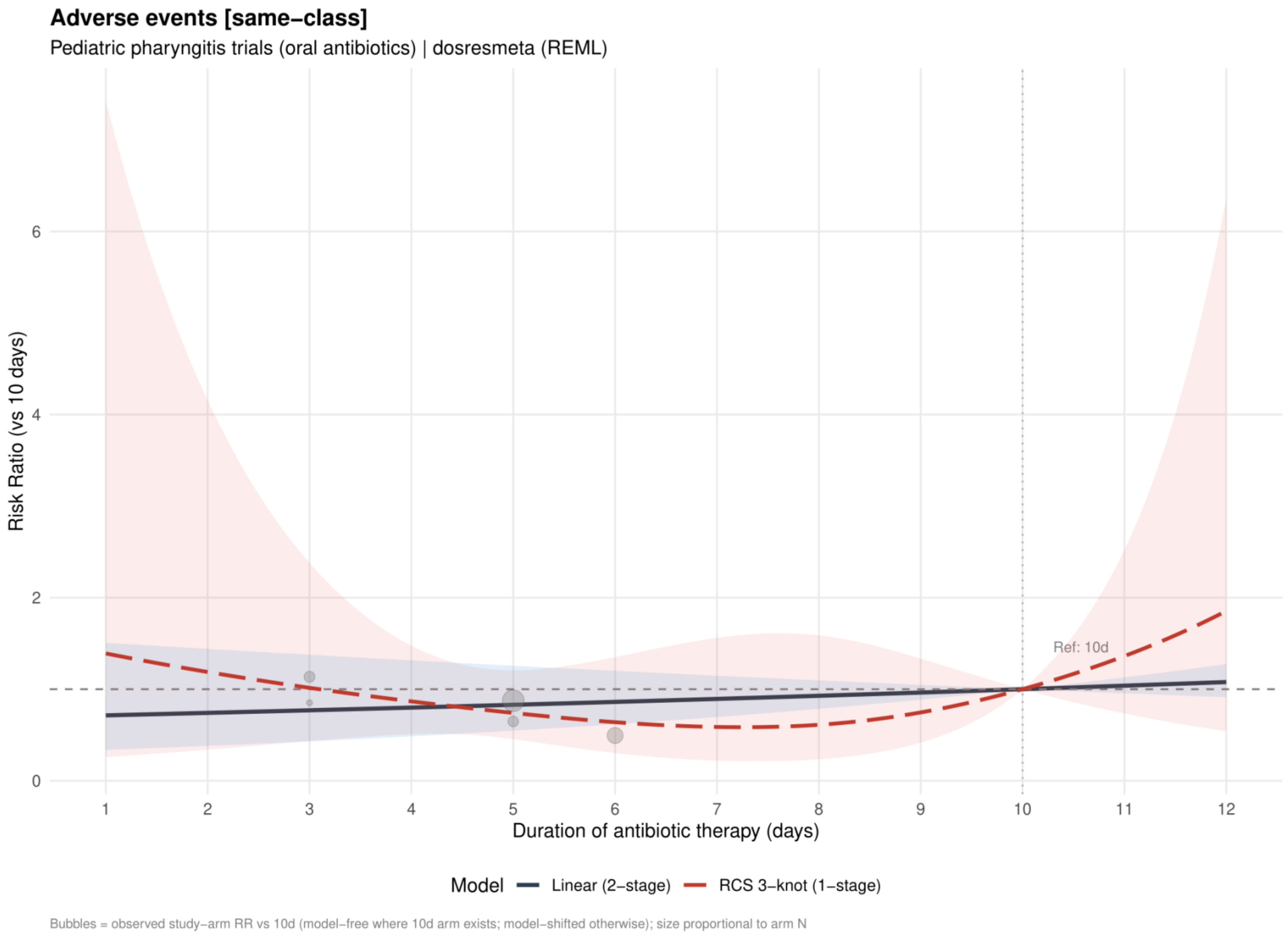


Curves show risk ratios relative to 10 days. The solid slate curve is the two stage linear model and, when present, the dashed red curve is the one stage restricted cubic spline with three knots; shaded bands show 95% confidence intervals. Bubbles show observed study arm effects and are scaled by arm sample size.

**Supplement 67. Adverse events: linear estimates by risk of bias restricted to the same antibiotic class**

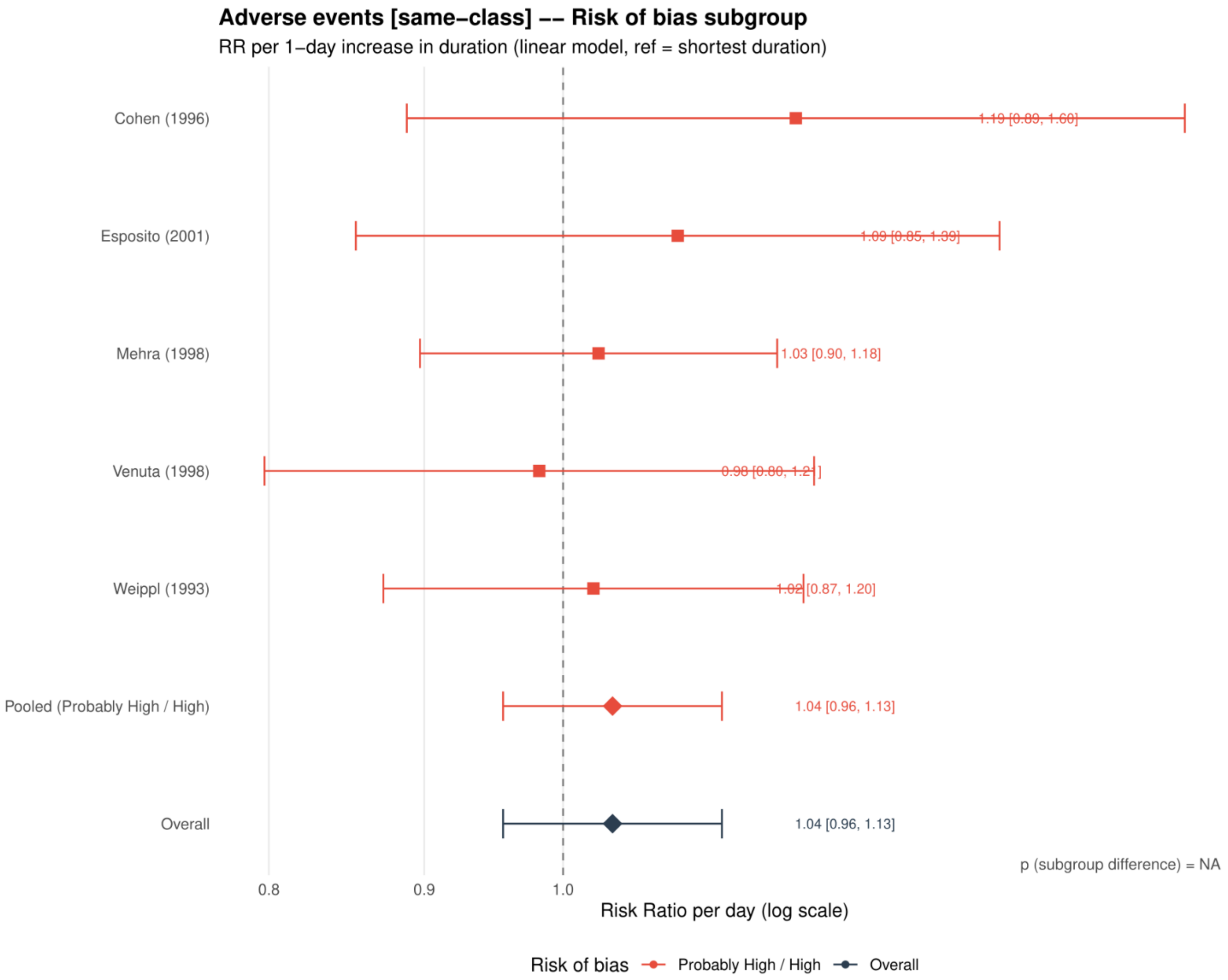


Study specific and pooled estimates are risk ratios per 1 day increase in antibiotic duration, with 95% confidence intervals.
*Only one collapsed risk of bias category contributed; the subgroup difference was not estimable.*

**Supplement 68. Adverse events: restricted cubic spline estimates by risk of bias restricted to the same antibiotic class**

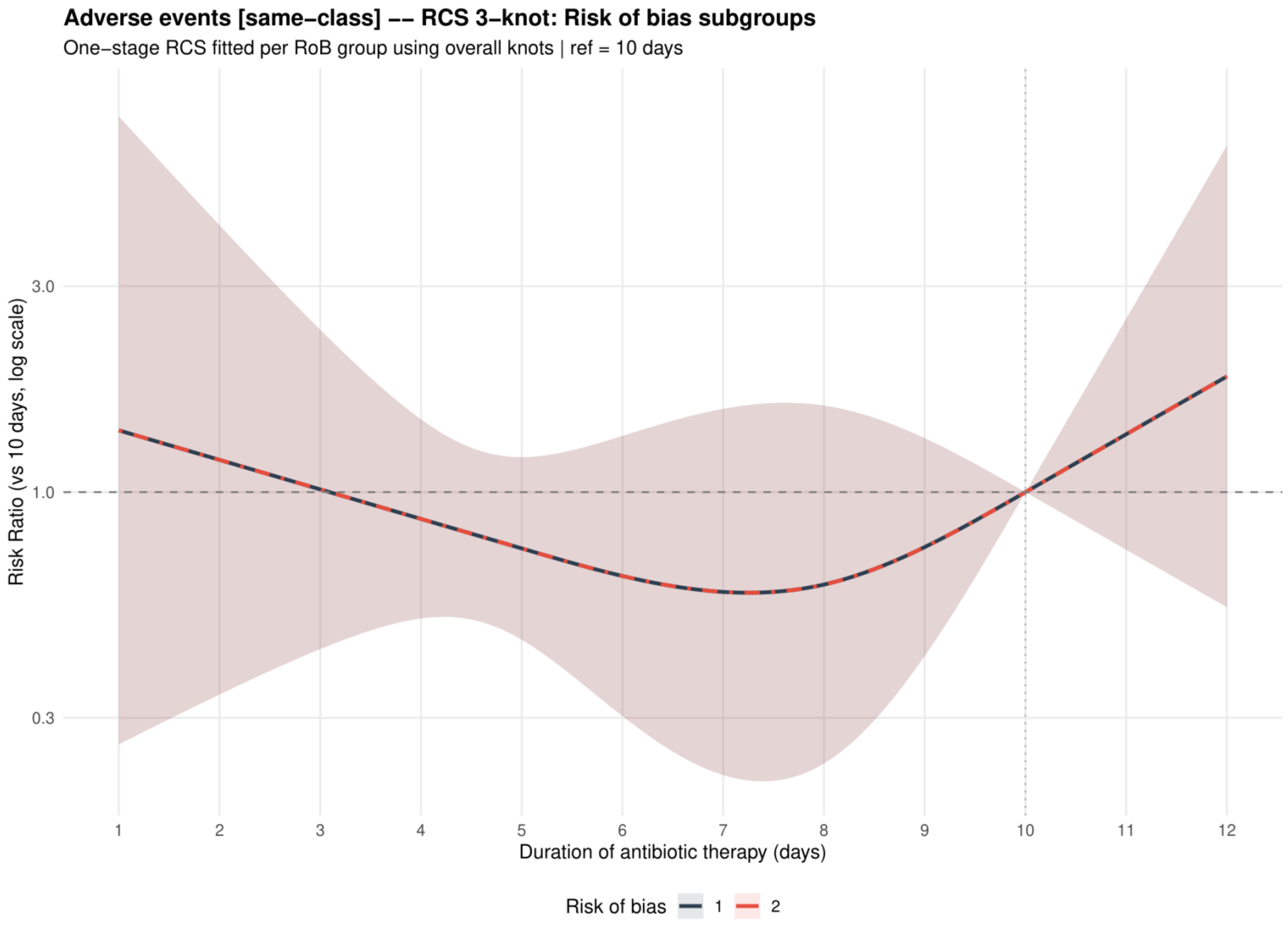


Curves show risk ratios relative to 10 days with 95% confidence intervals. Risk of bias codes: 1 = overall; 2 = low or probably low; 3 = probably high or high.

**Supplement 69. Adverse events: dose-response analysis excluding azithromycin**

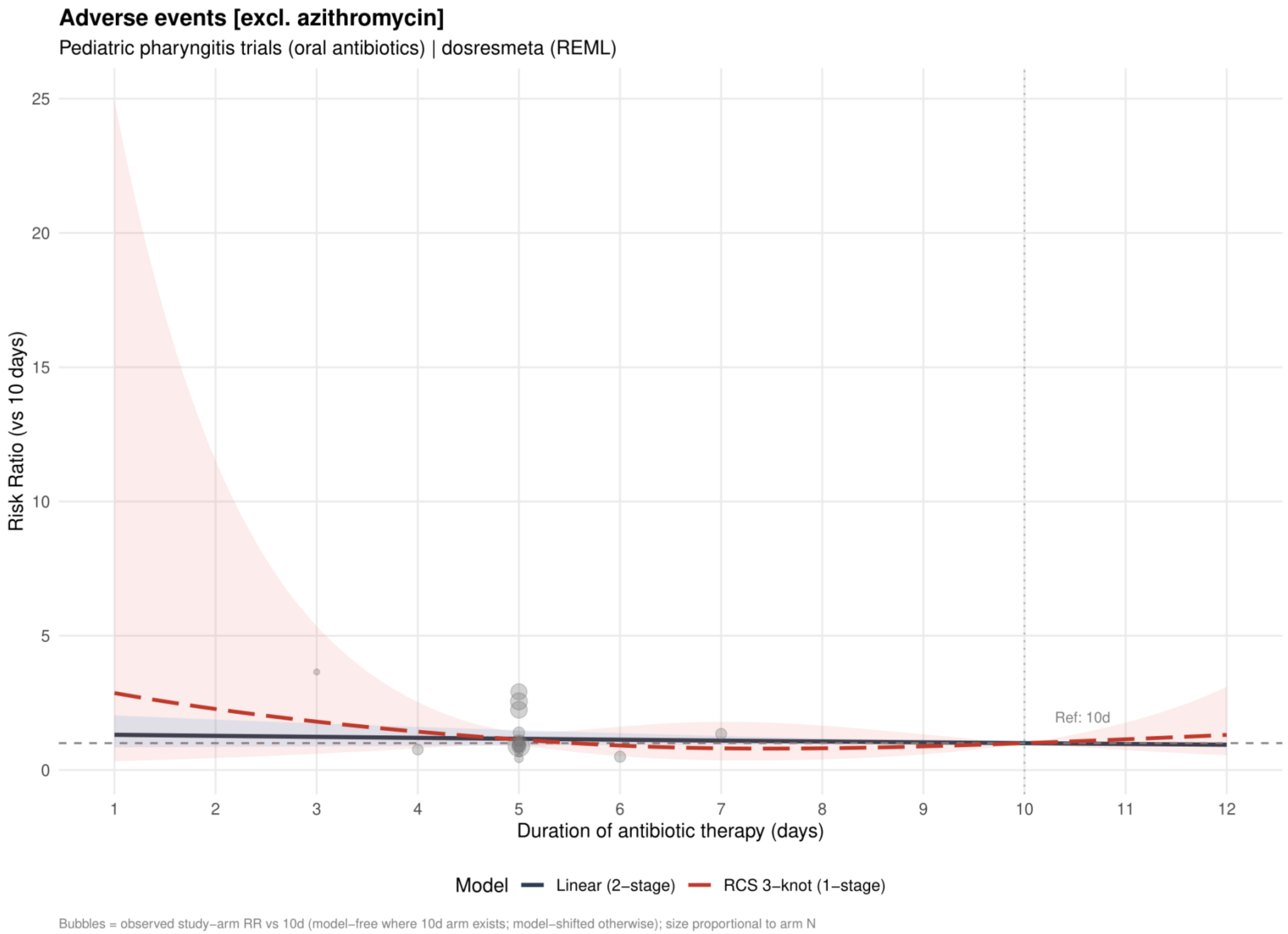


Curves show risk ratios relative to 10 days. The solid slate curve is the two stage linear model and, when present, the dashed red curve is the one stage restricted cubic spline with three knots; shaded bands show 95% confidence intervals. Bubbles show observed study arm effects and are scaled by arm sample size.

**Supplement 70. Adverse events: linear estimates by risk of bias excluding azithromycin**

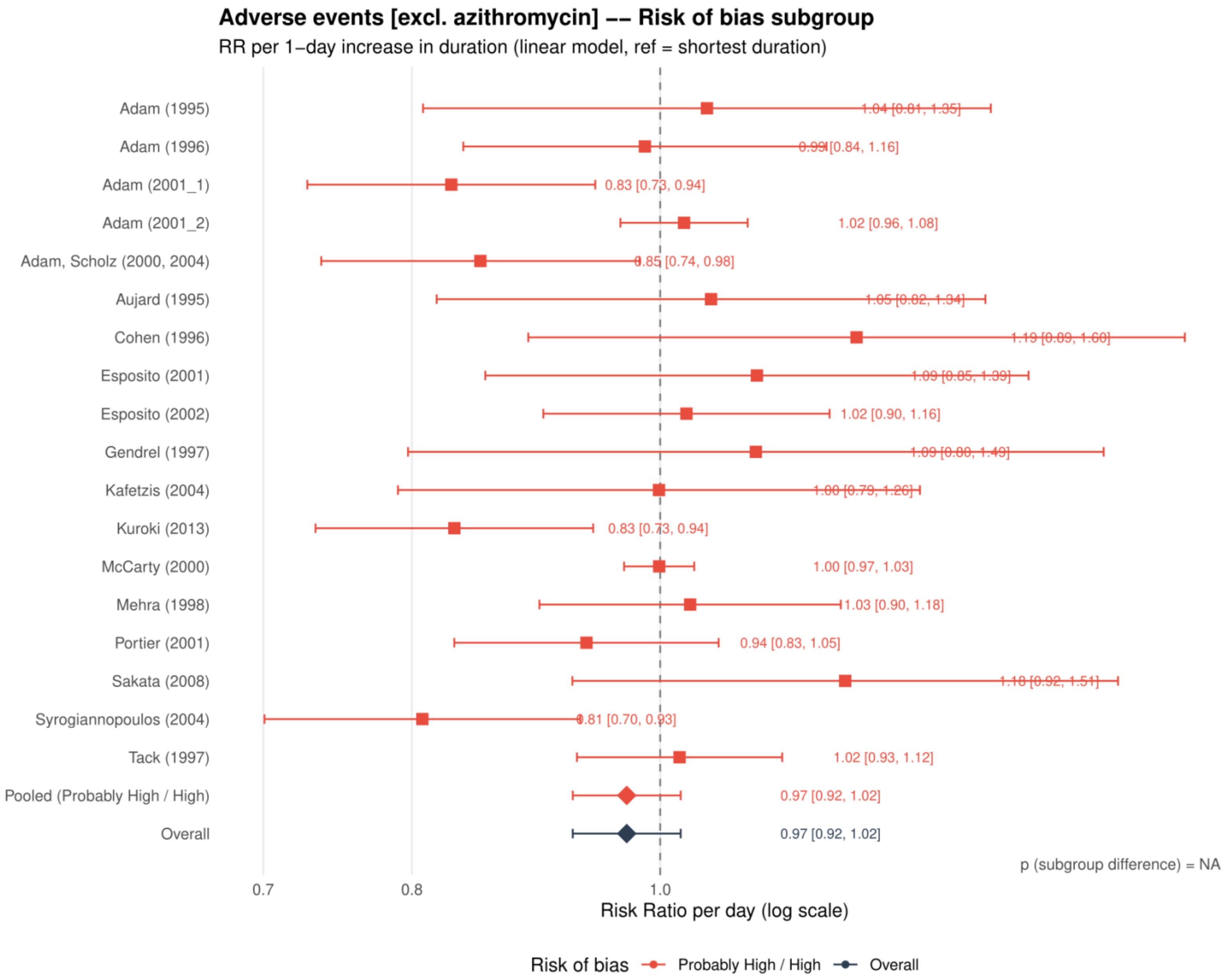


Study specific and pooled estimates are risk ratios per 1 day increase in antibiotic duration, with 95% confidence intervals.
*Only one collapsed risk of bias category contributed; the subgroup difference was not estimable.*

**Supplement 71. Adverse events: restricted cubic spline estimates by risk of bias excluding azithromycin**

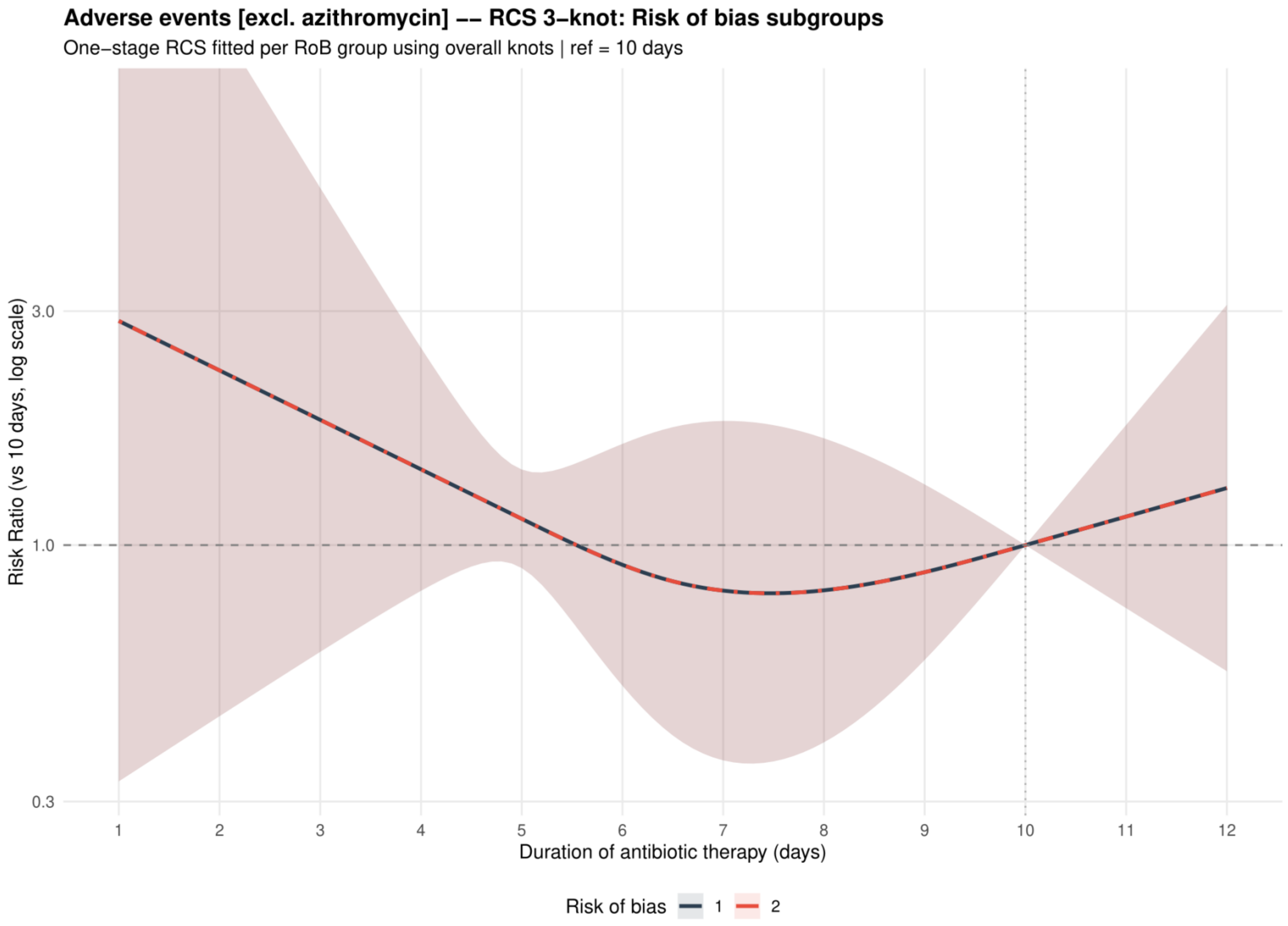


Curves show risk ratios relative to 10 days with 95% confidence intervals. Risk of bias codes: 1 = overall; 2 = low or probably low; 3 = probably high or high.

**Supplement 72. Adverse events: dose-response analysis excluding macrolides**

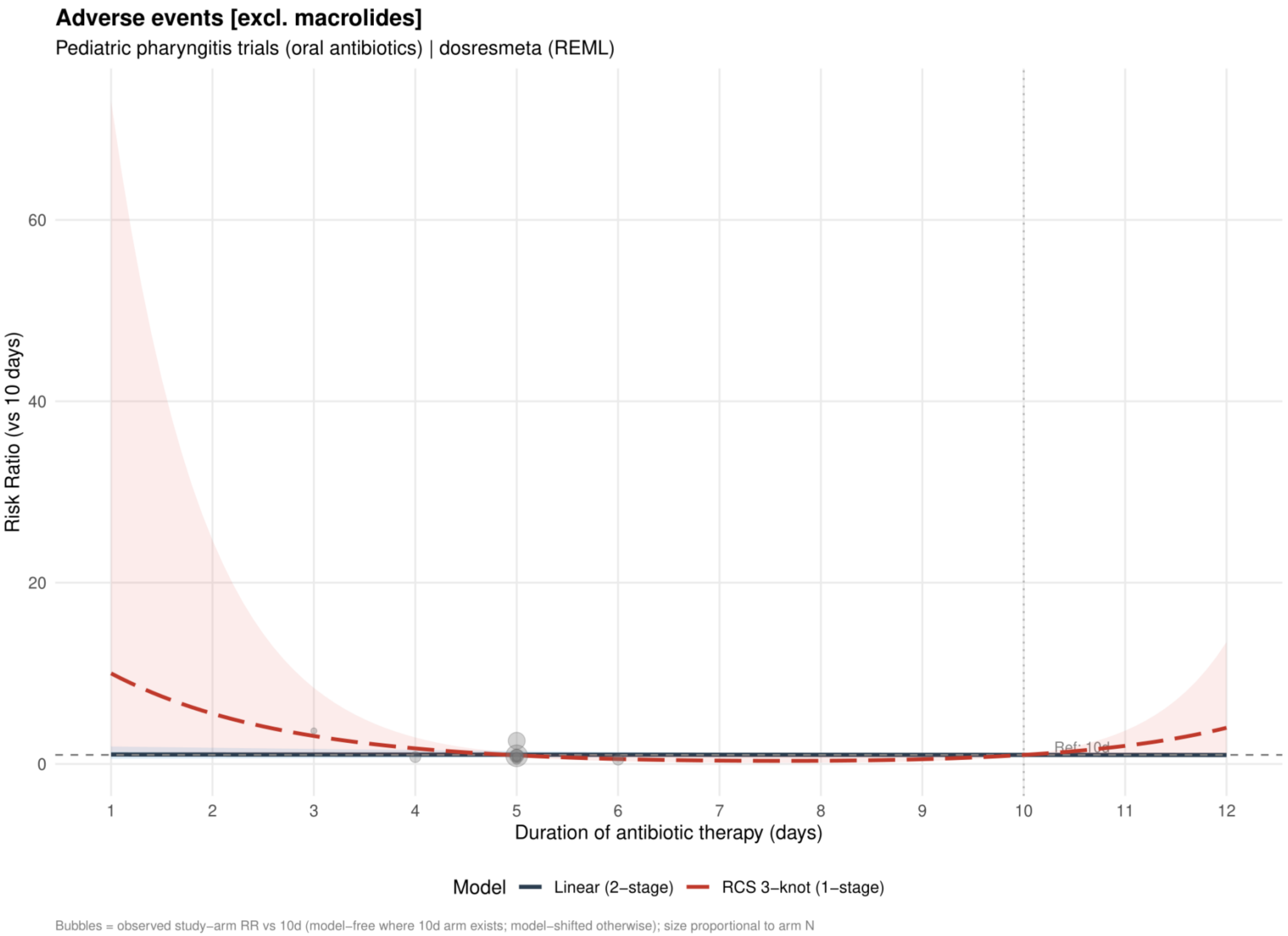


Curves show risk ratios relative to 10 days. The solid slate curve is the two stage linear model and, when present, the dashed red curve is the one stage restricted cubic spline with three knots; shaded bands show 95% confidence intervals. Bubbles show observed study arm effects and are scaled by arm sample size.

**Supplement 73. Adverse events: linear estimates by risk of bias excluding macrolides**

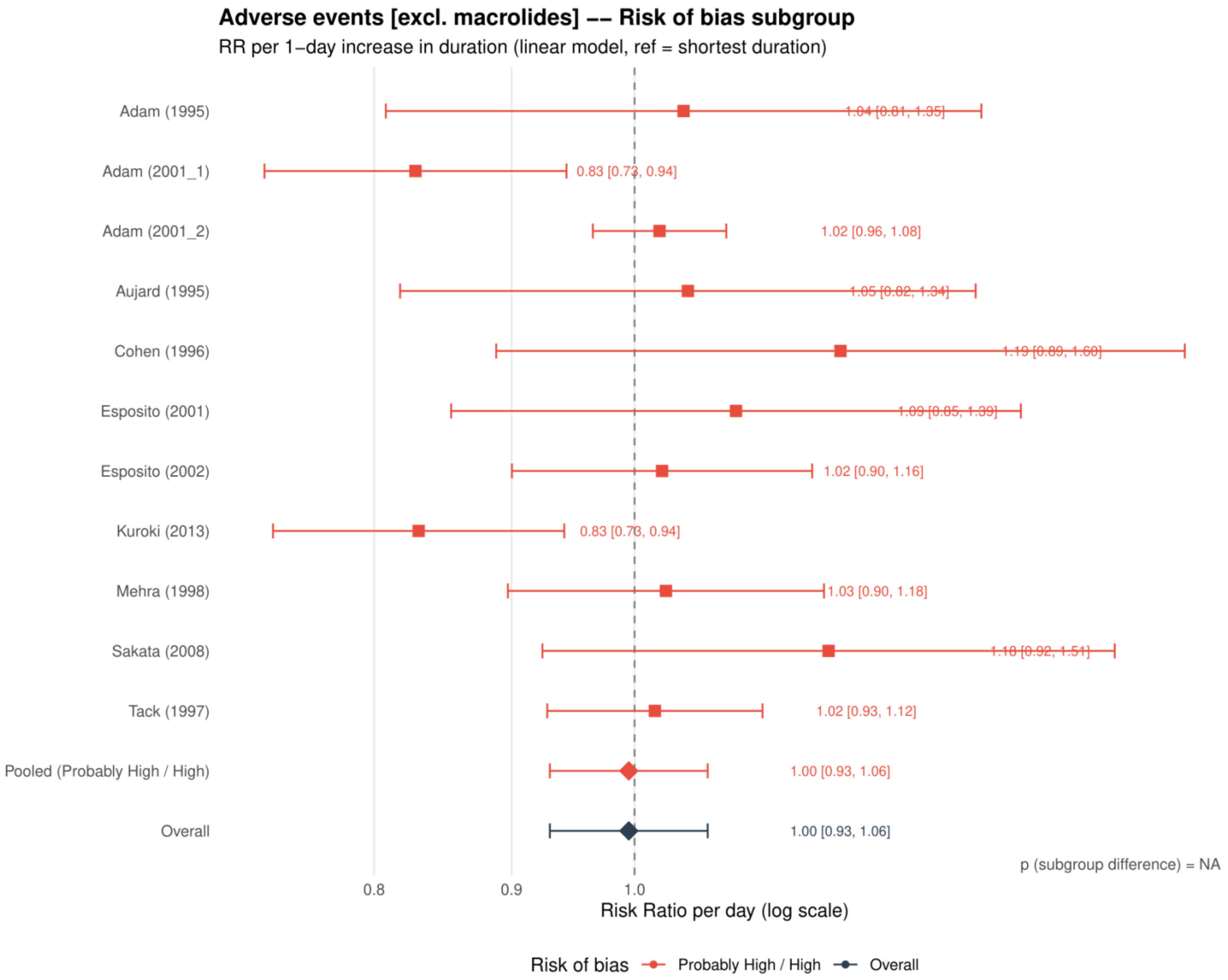


Study specific and pooled estimates are risk ratios per 1 day increase in antibiotic duration, with 95% confidence intervals.
*Only one collapsed risk of bias category contributed; the subgroup difference was not estimable.*

**Supplement 74. Adverse events: restricted cubic spline estimates by risk of bias excluding macrolides**

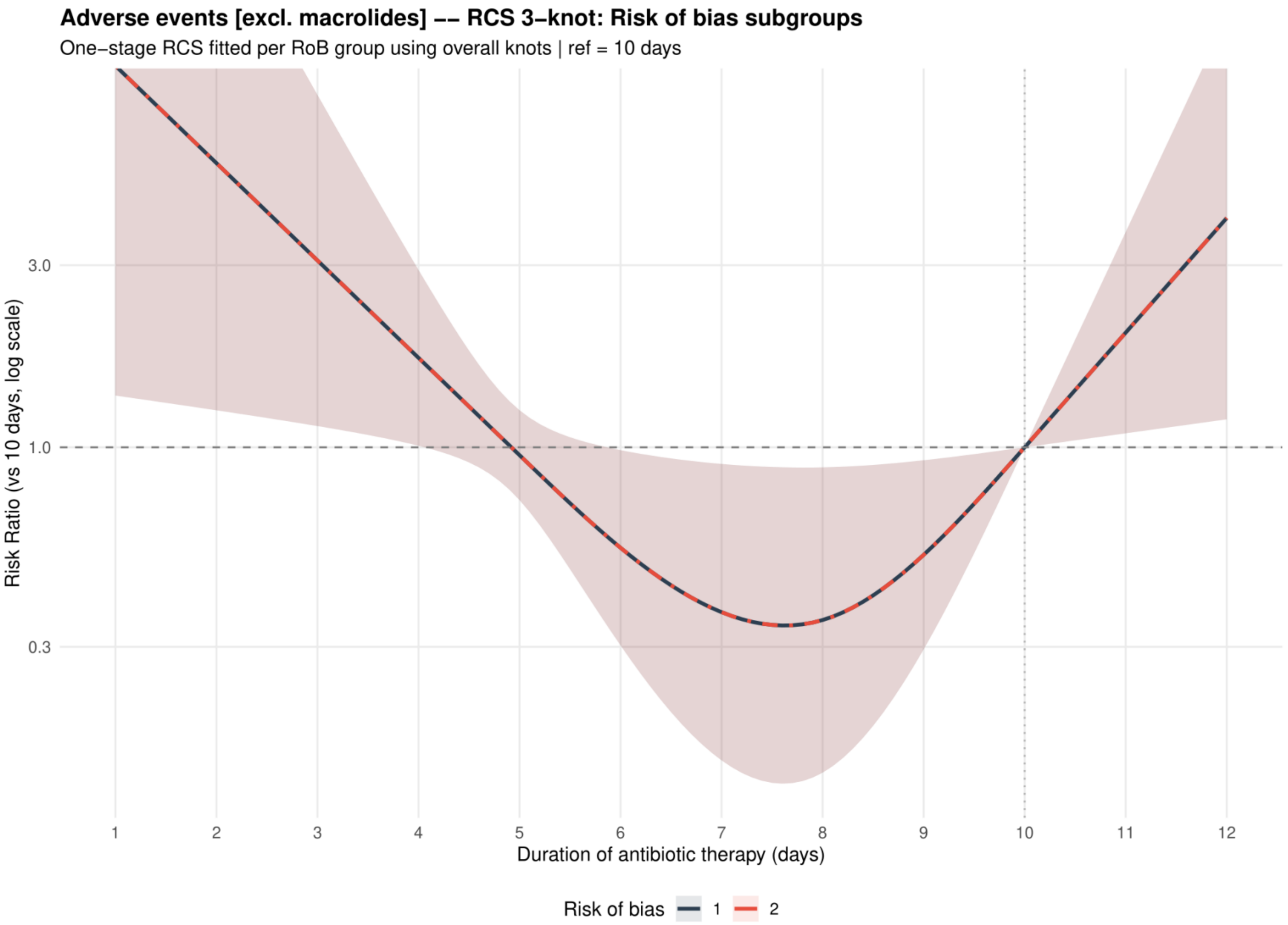


Curves show risk ratios relative to 10 days with 95% confidence intervals. Risk of bias codes: 1 = overall; 2 = low or probably low; 3 = probably high or high.

**Supplement 75. Adverse events: dose-response analysis including placebo or no treatment as 0 days**

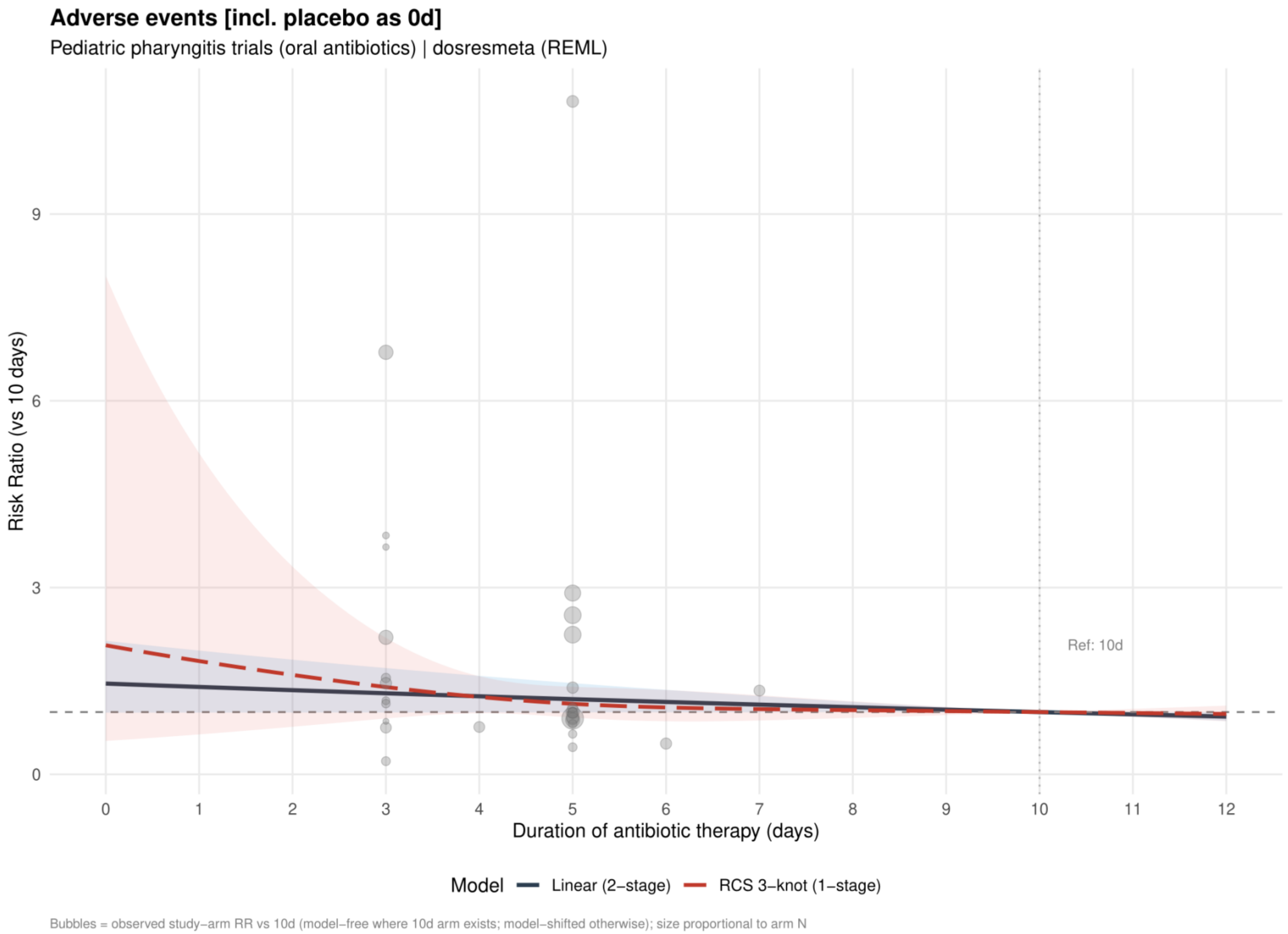


Curves show risk ratios relative to 10 days. The solid slate curve is the two stage linear model and, when present, the dashed red curve is the one stage restricted cubic spline with three knots; shaded bands show 95% confidence intervals. Bubbles show observed study arm effects and are scaled by arm sample size.

**Supplement 76. Adverse events: linear estimates by risk of bias including placebo or no treatment as 0 days**

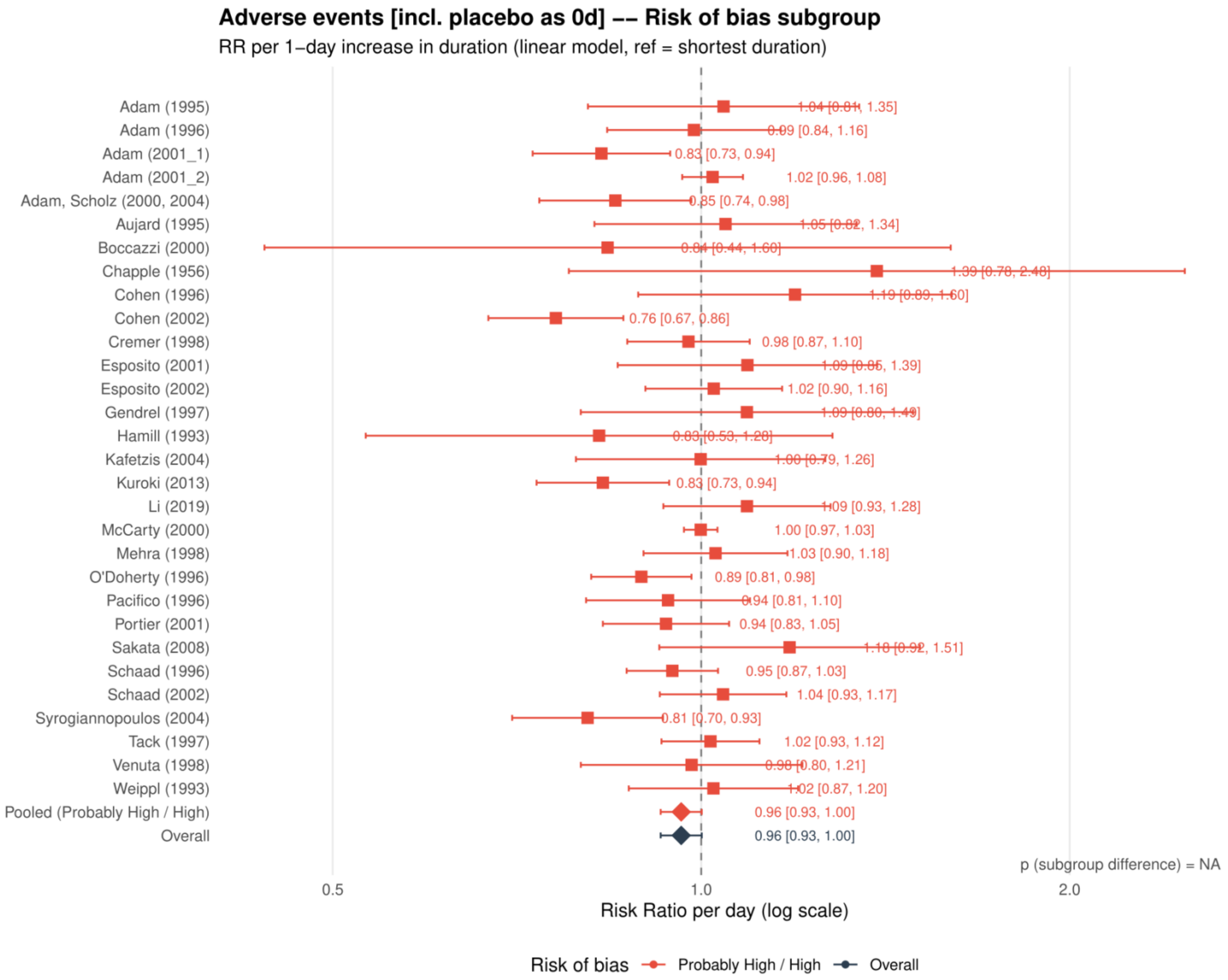


Study specific and pooled estimates are risk ratios per 1 day increase in antibiotic duration, with 95% confidence intervals.

*Only one collapsed risk of bias category contributed; the subgroup difference was not estimable.*

**Supplement 77. Adverse events: restricted cubic spline estimates by risk of bias including placebo or no treatment as 0 days**

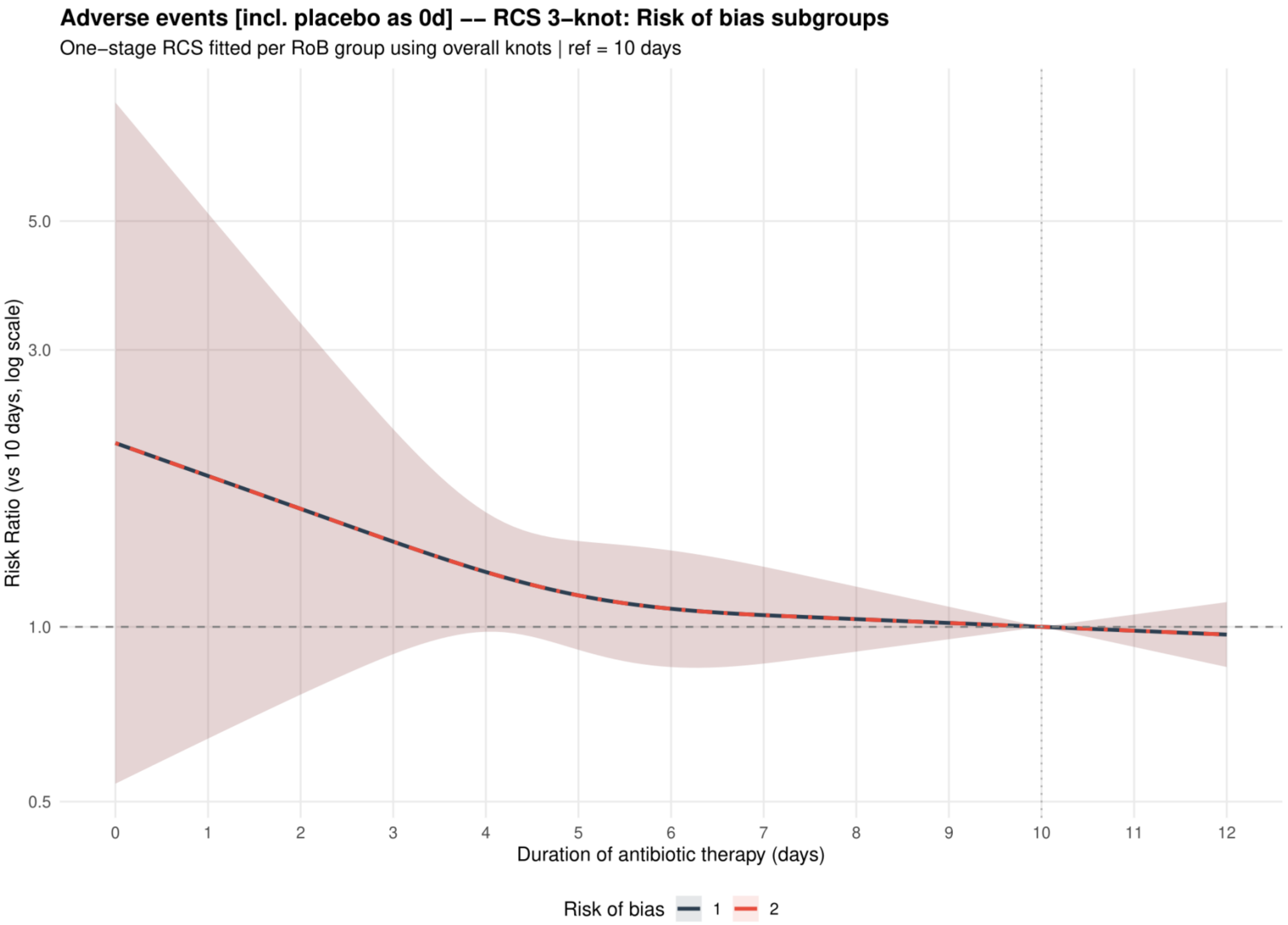


Curves show risk ratios relative to 10 days with 95% confidence intervals. Risk of bias codes: 1 = overall; 2 = low or probably low; 3 = probably high or high.

**Supplement 78. Adverse events: funnel plot including placebo or no treatment as 0 days**

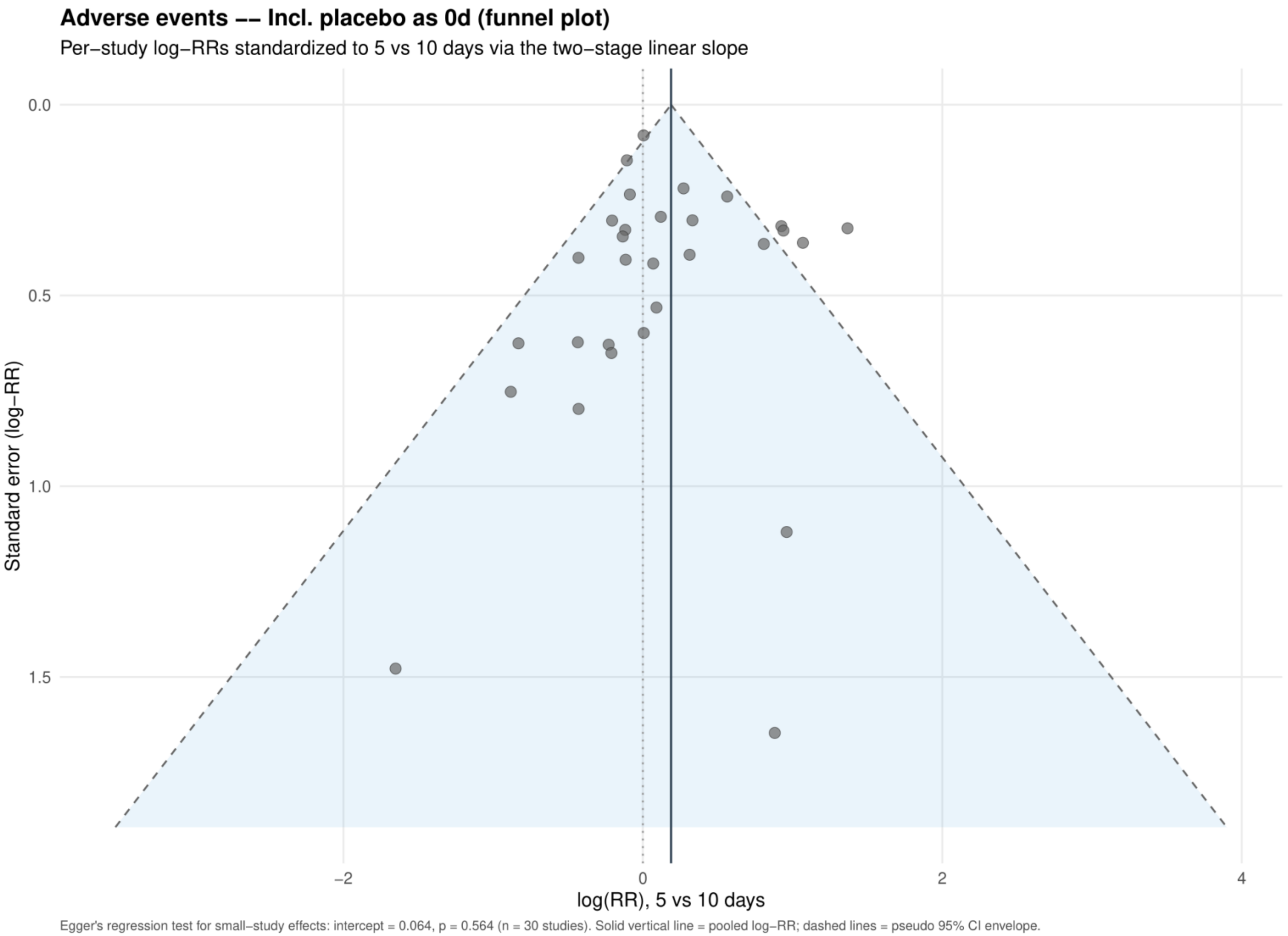


Study specific log risk ratios were standardized to 5 versus 10 days using the two stage linear slope. The solid vertical line is the pooled log risk ratio and dashed lines are pseudo 95% confidence limits; Egger's test is reported below the plot.

**Supplement 79. Mortality: dose-response analysis for the primary analysis**

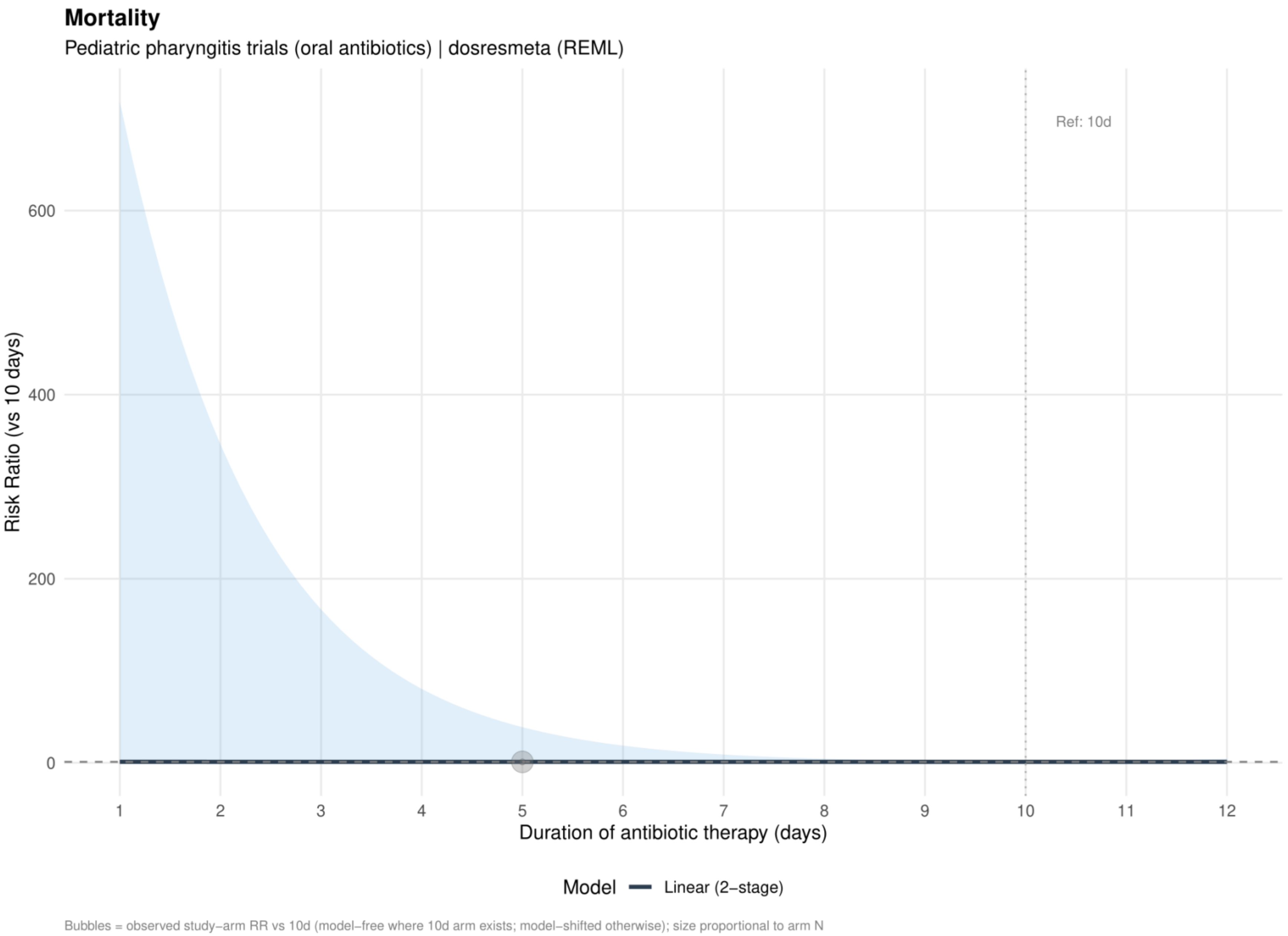


Curves show risk ratios relative to 10 days. The solid slate curve is the two stage linear model and, when present, the dashed red curve is the one stage restricted cubic spline with three knots; shaded bands show 95% confidence intervals. Bubbles show observed study arm effects and are scaled by arm sample size.

**Supplement 80. Mortality: linear estimates by risk of bias for the primary analysis**

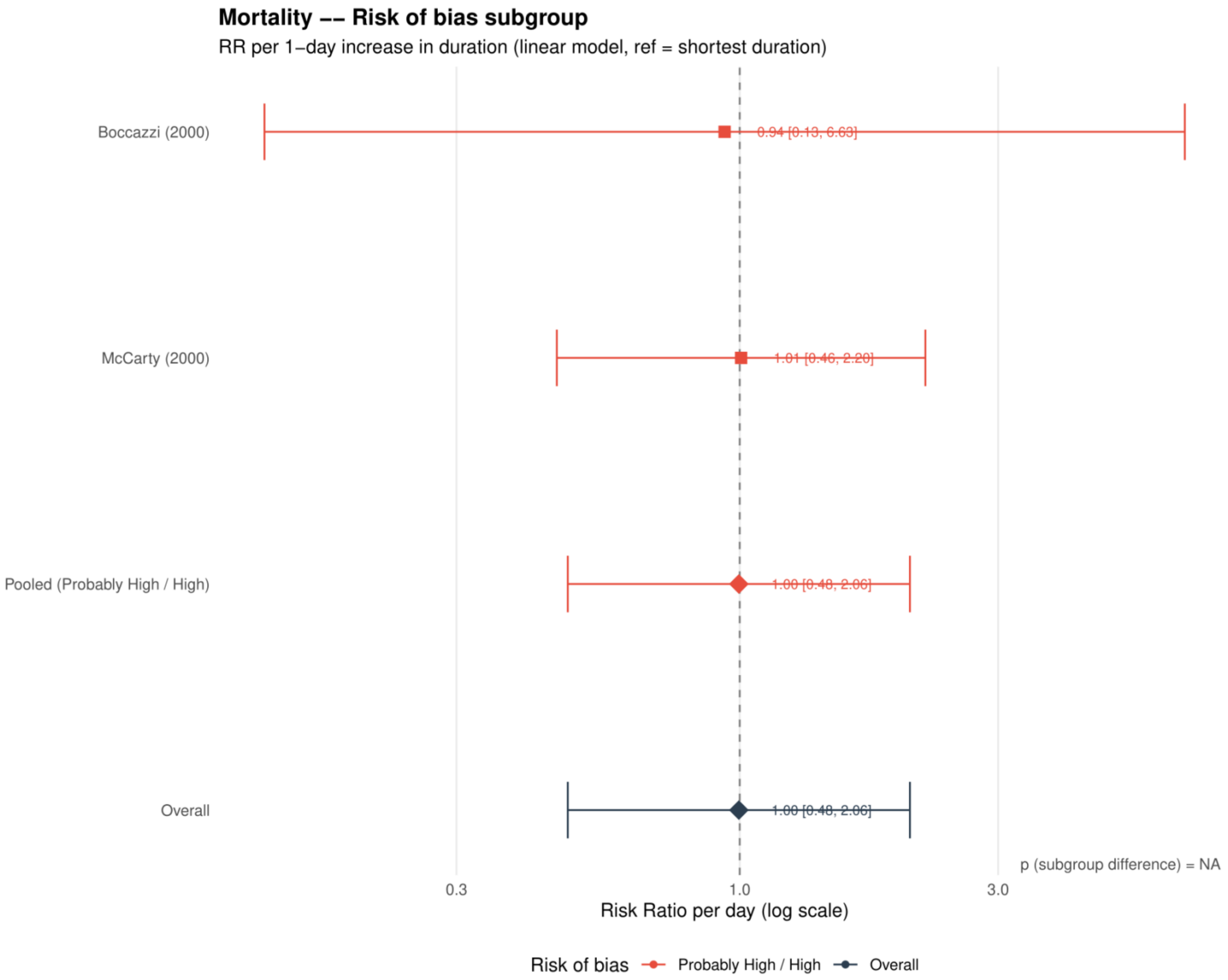


Study specific and pooled estimates are risk ratios per 1 day increase in antibiotic duration, with 95% confidence intervals.
*Only one collapsed risk of bias category contributed; the subgroup difference was not estimable.*

**Supplement 81. Mortality: dose-response analysis including placebo or no treatment as 0 days**

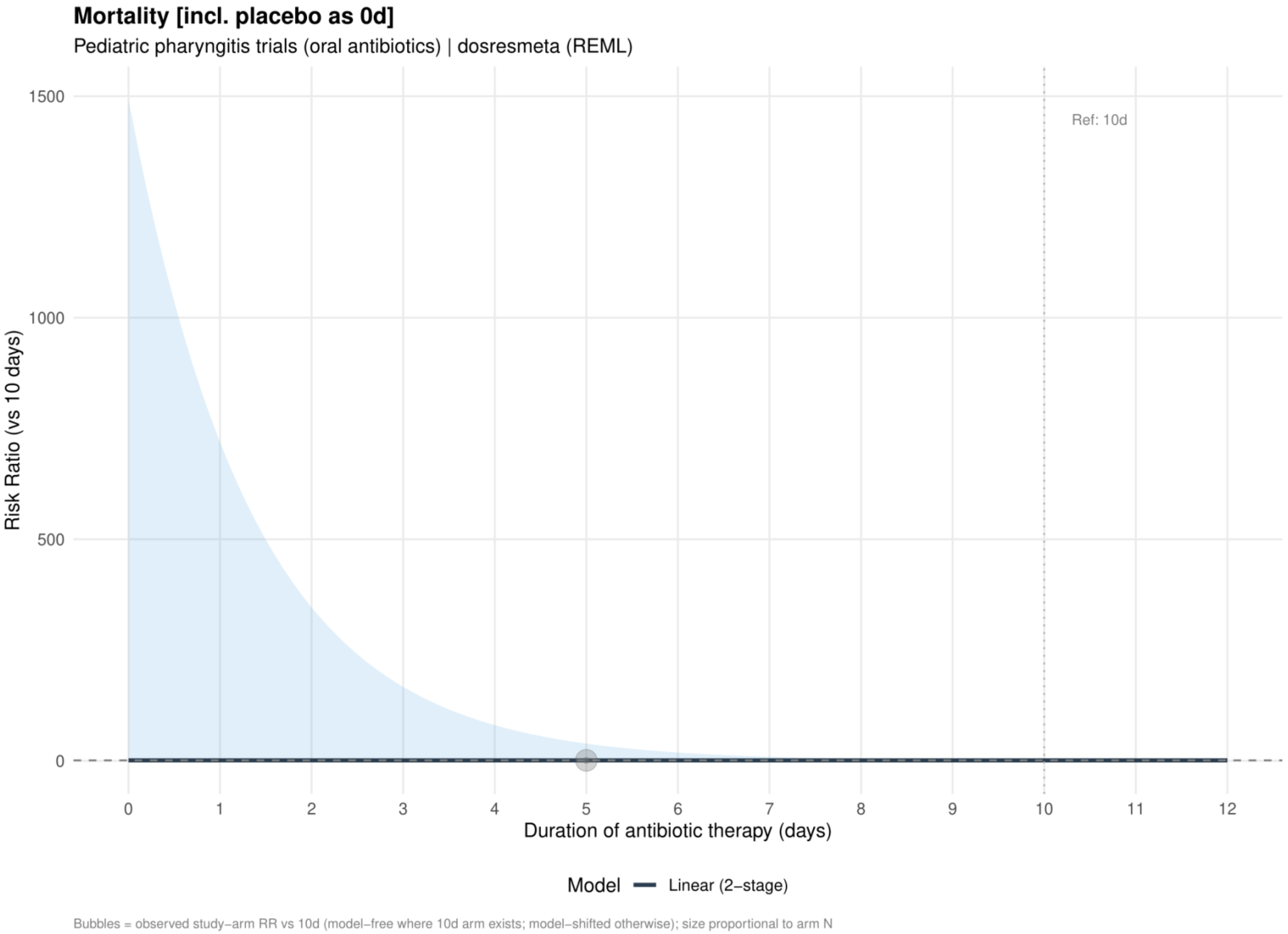


Curves show risk ratios relative to 10 days. The solid slate curve is the two stage linear model and, when present, the dashed red curve is the one stage restricted cubic spline with three knots; shaded bands show 95% confidence intervals. Bubbles show observed study arm effects and are scaled by arm sample size.

*No placebo or no treatment mortality data were available; this analysis is identical to Supplement 79.*

**Supplement 82. Mortality: linear estimates by risk of bias including placebo or no treatment as 0 days**

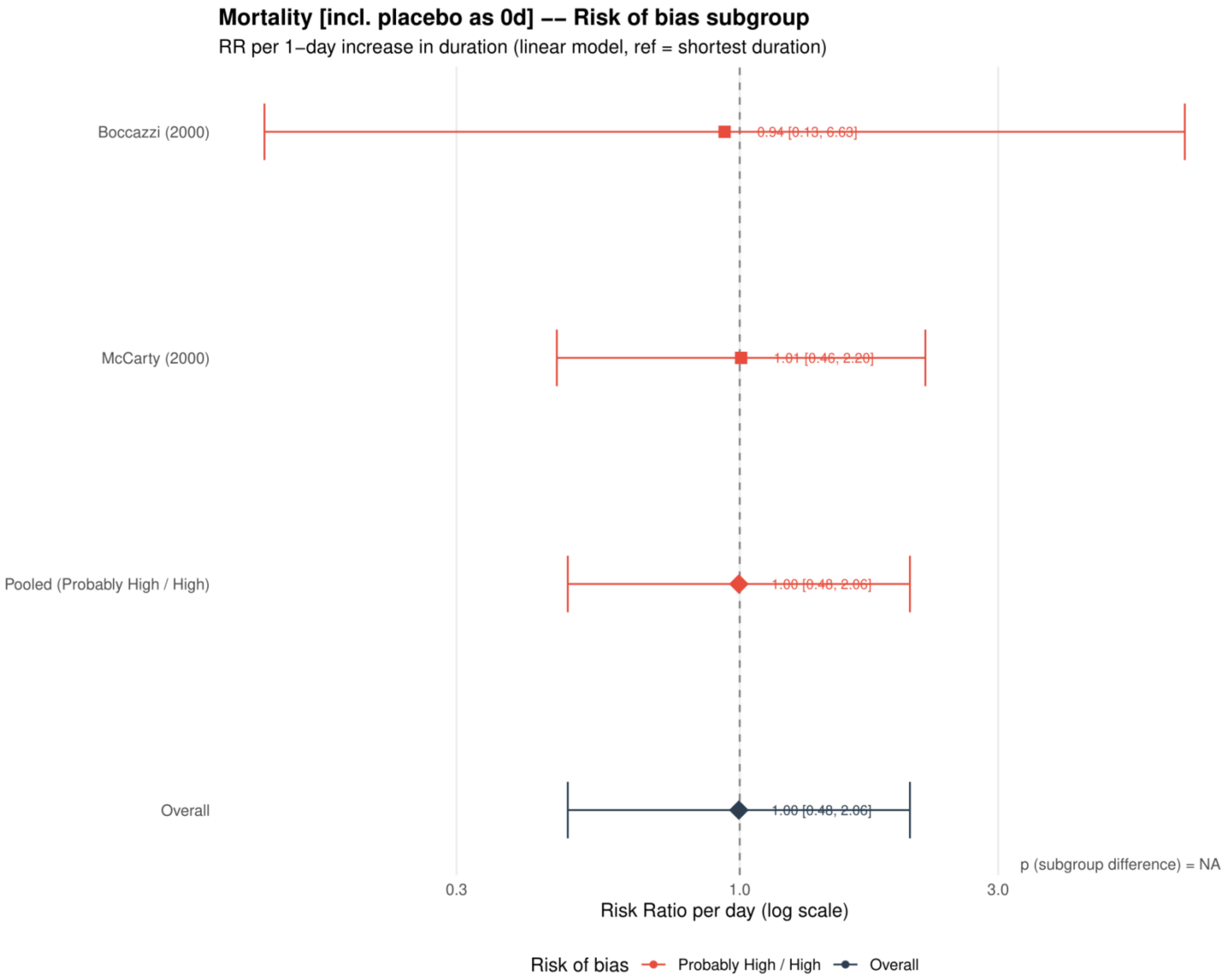


Study specific and pooled estimates are risk ratios per 1 day increase in antibiotic duration, with 95% confidence intervals.
*Only one collapsed risk of bias category contributed; the subgroup difference was not estimable.*
*No placebo or no treatment mortality data were available; this analysis is identical to Supplement 80.*

**Supplement 83. Serious adverse events: dose-response analysis for the primary analysis**

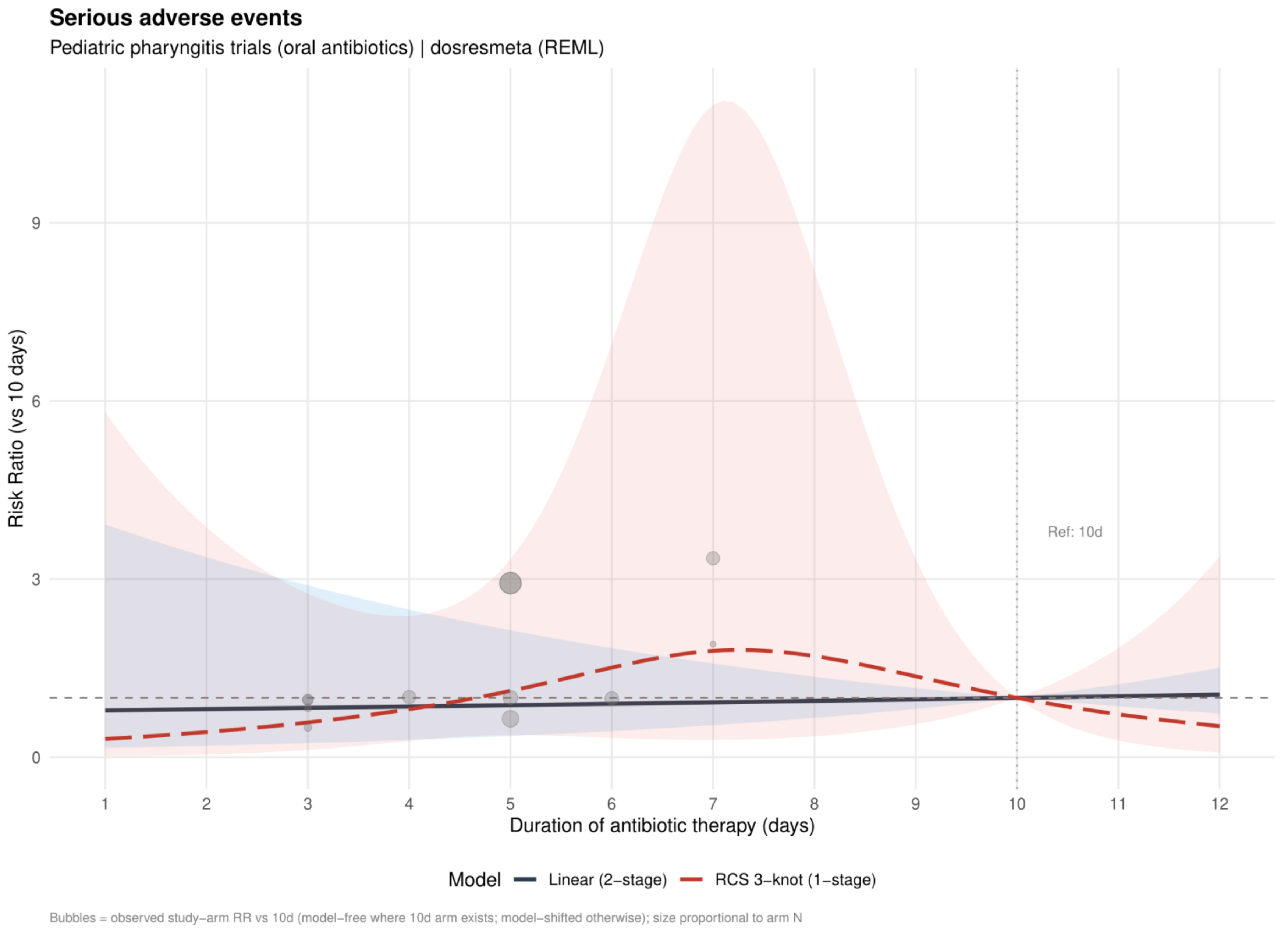


Curves show risk ratios relative to 10 days. The solid slate curve is the two stage linear model and, when present, the dashed red curve is the one stage restricted cubic spline with three knots; shaded bands show 95% confidence intervals. Bubbles show observed study arm effects and are scaled by arm sample size.

**Supplement 84. Serious adverse events: linear estimates by risk of bias for the primary analysis**

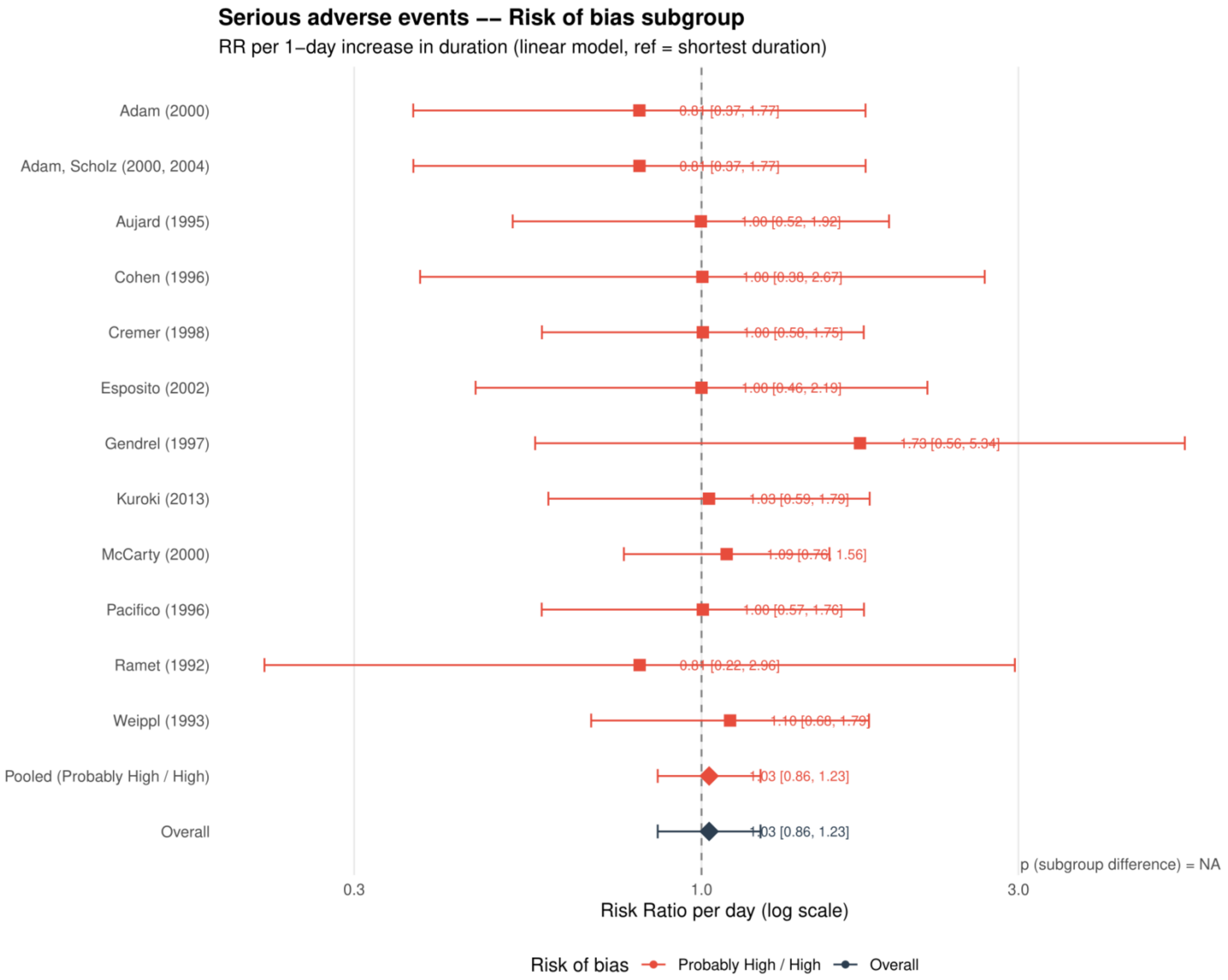


Study specific and pooled estimates are risk ratios per 1 day increase in antibiotic duration, with 95% confidence intervals.
*Only one collapsed risk of bias category contributed; the subgroup difference was not estimable.*

**Supplement 85. Serious adverse events: restricted cubic spline estimates by risk of bias for the primary analysis**

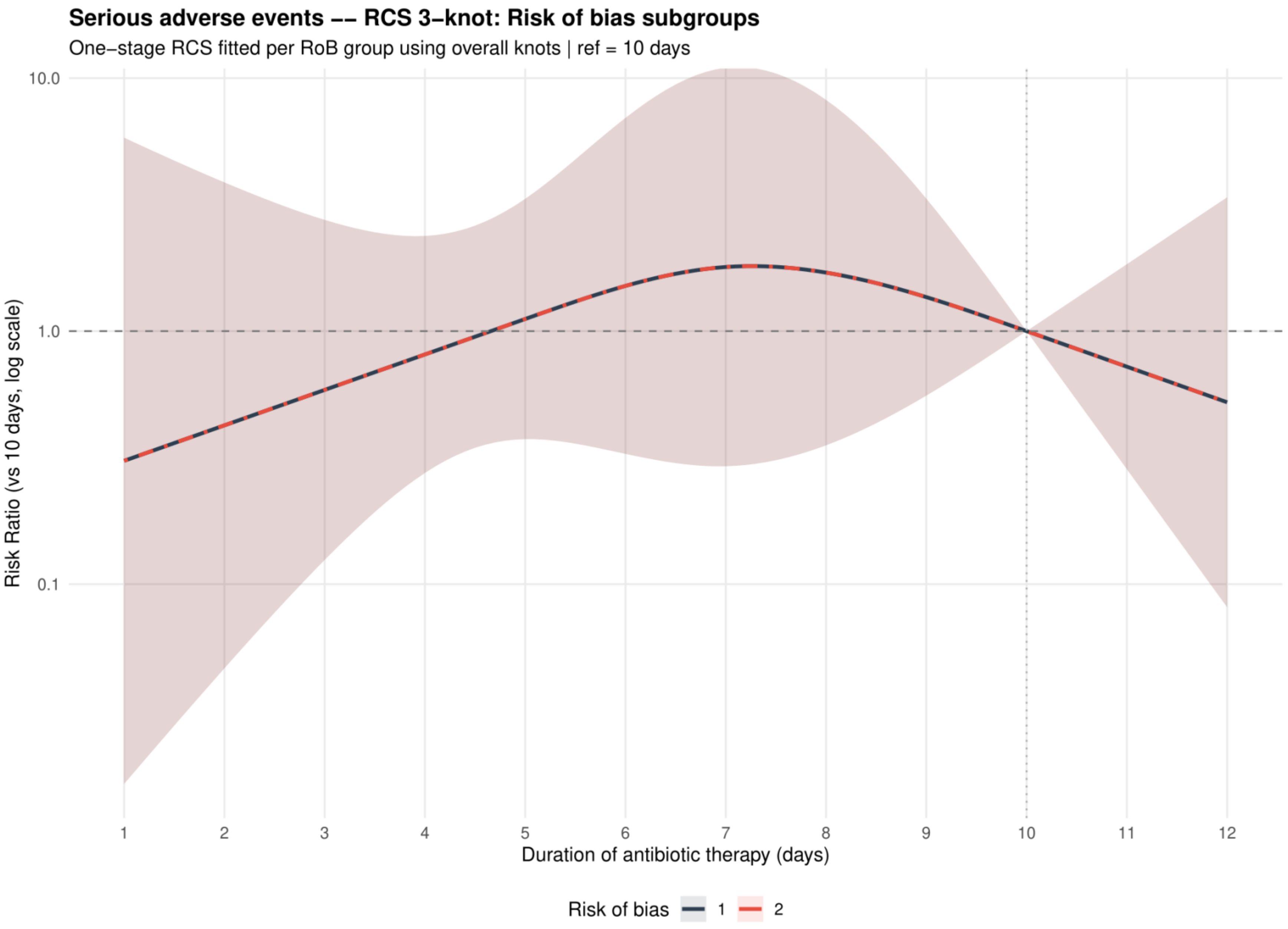


Curves show risk ratios relative to 10 days with 95% confidence intervals. Risk of bias codes: 1 = overall; 2 = low or probably low; 3 = probably high or high.

**Supplement 86. Serious adverse events: funnel plot for the primary analysis**

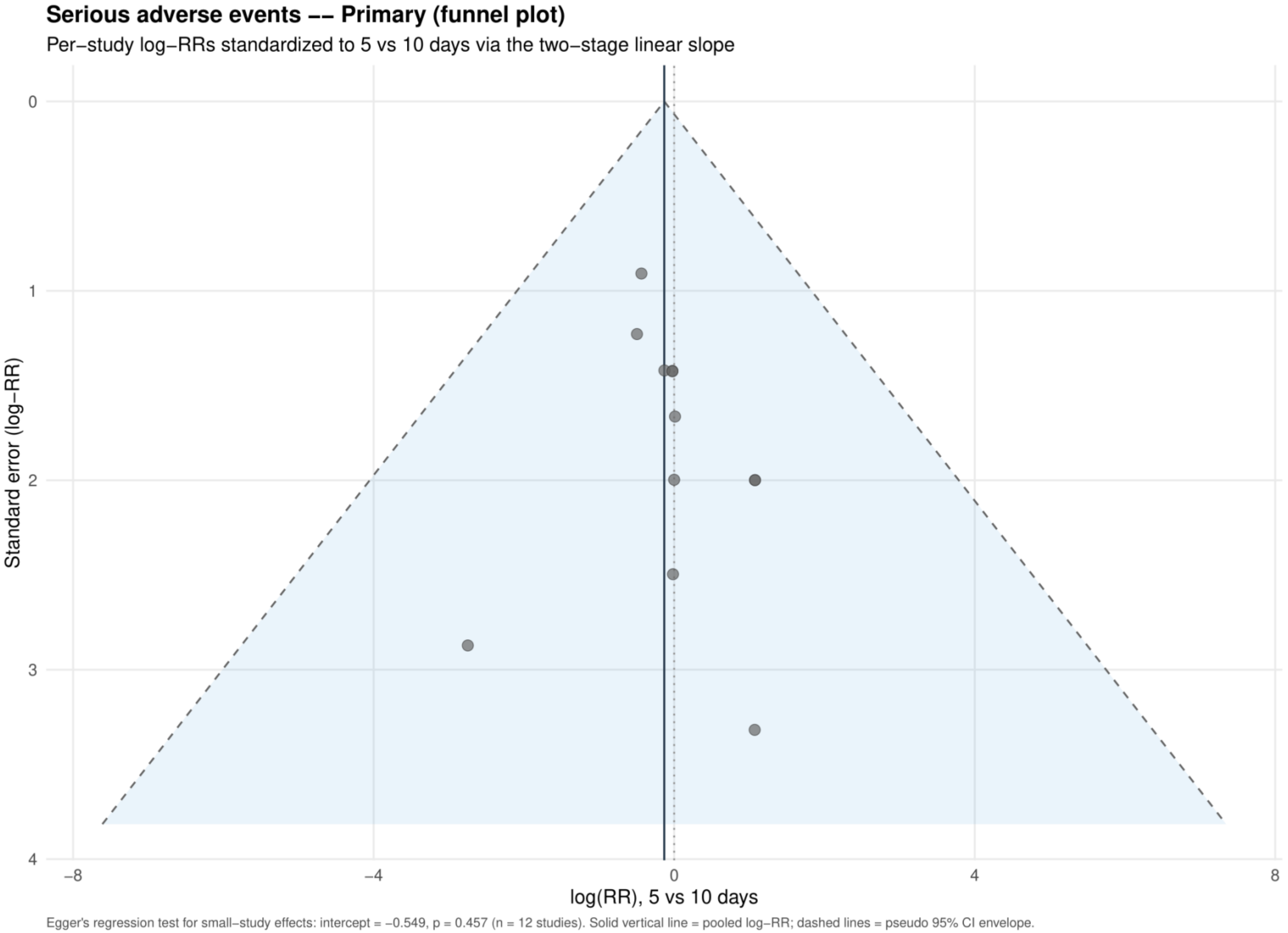


Study specific log risk ratios were standardized to 5 versus 10 days using the two stage linear slope. The solid vertical line is the pooled log risk ratio and dashed lines are pseudo 95% confidence limits; Egger's test is reported below the plot.

**Supplement 87. Serious adverse events: dose-response analysis restricted to the same antibiotic class**

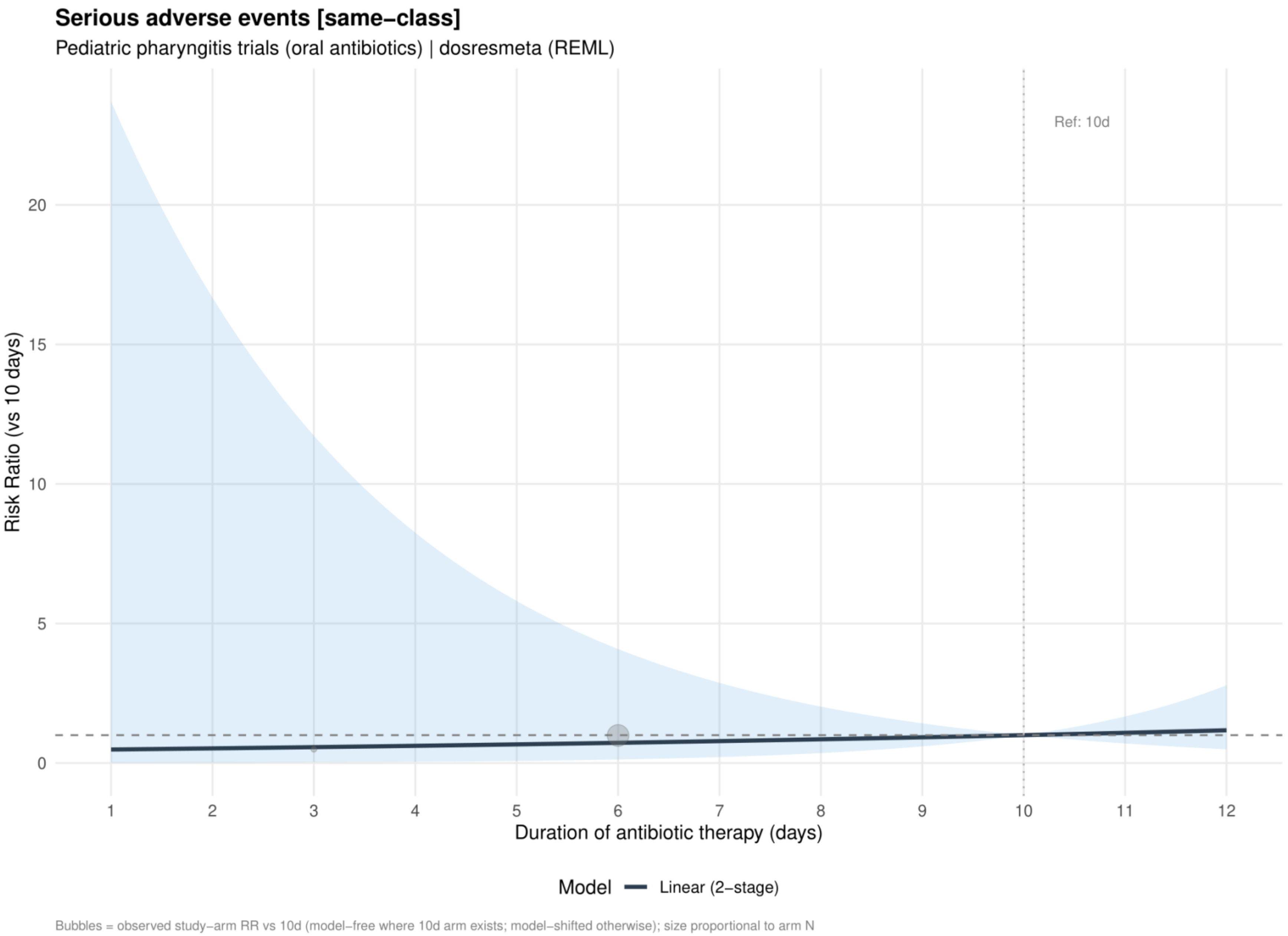


Curves show risk ratios relative to 10 days. The solid slate curve is the two stage linear model and, when present, the dashed red curve is the one stage restricted cubic spline with three knots; shaded bands show 95% confidence intervals. Bubbles show observed study arm effects and are scaled by arm sample size.

**Supplement 88. Serious adverse events: linear estimates by risk of bias restricted to the same antibiotic class**

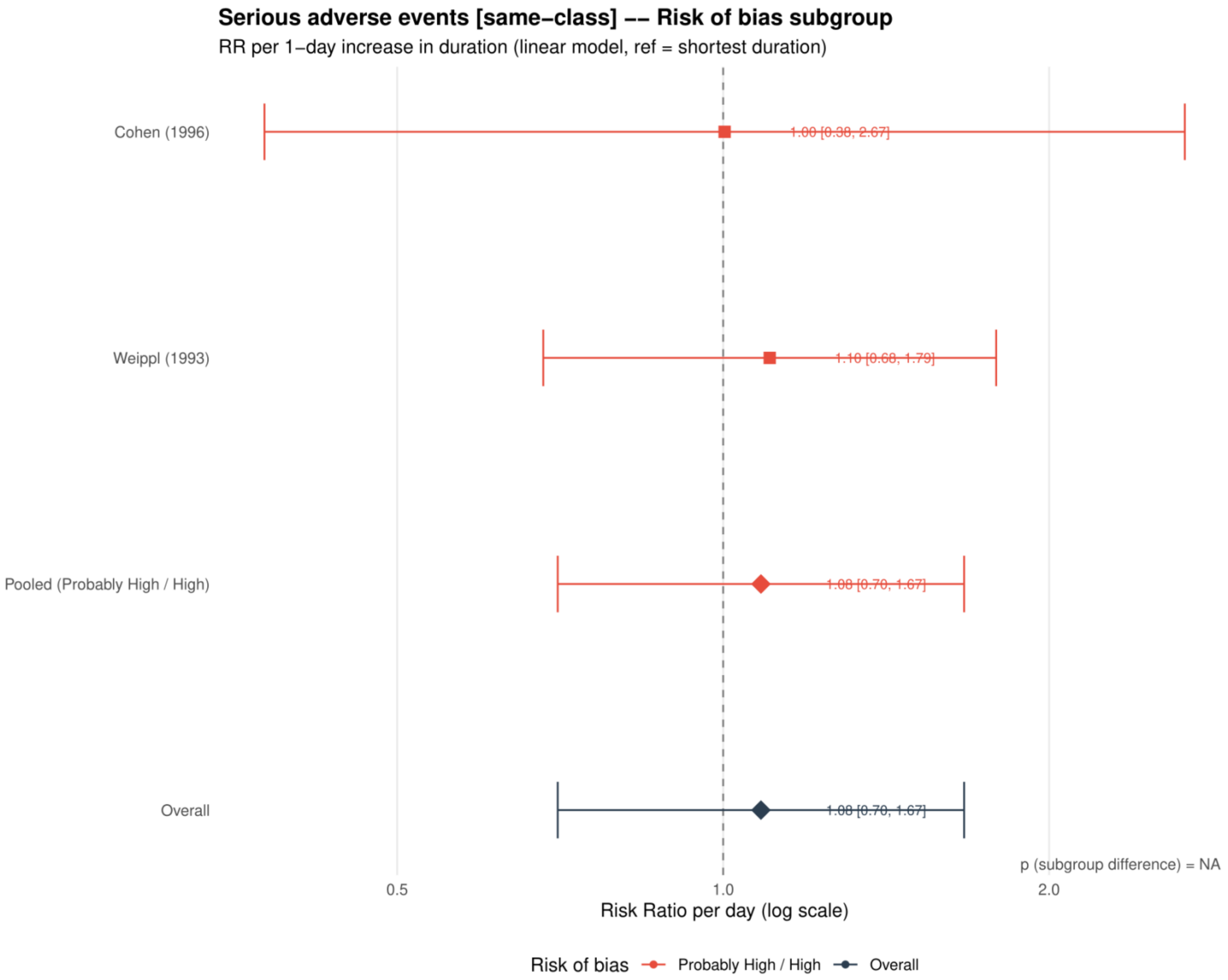


Study specific and pooled estimates are risk ratios per 1 day increase in antibiotic duration, with 95% confidence intervals.
*Only one collapsed risk of bias category contributed; the subgroup difference was not estimable.*

**Supplement 89. Serious adverse events: dose-response analysis excluding azithromycin**

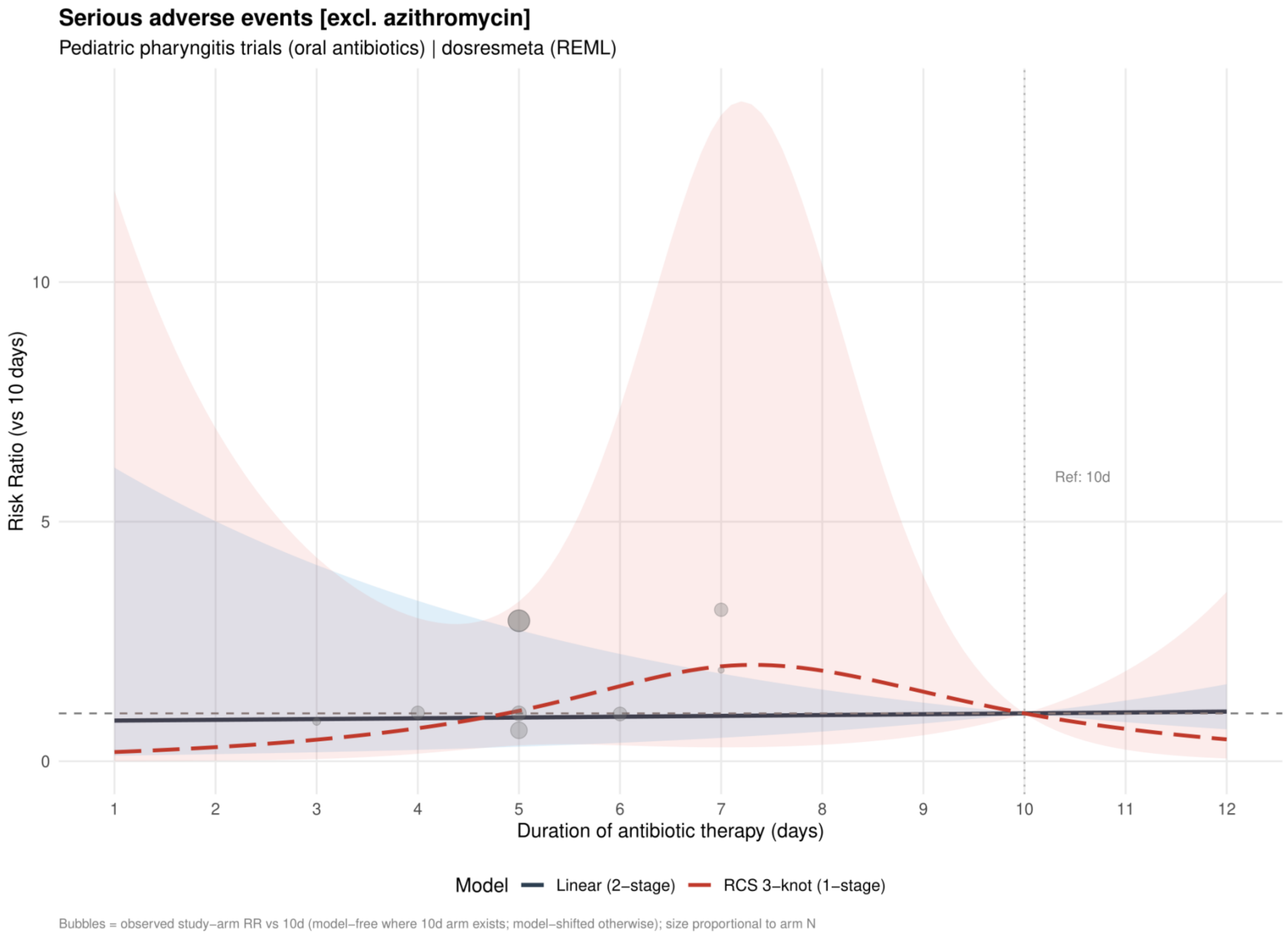


Curves show risk ratios relative to 10 days. The solid slate curve is the two stage linear model and, when present, the dashed red curve is the one stage restricted cubic spline with three knots; shaded bands show 95% confidence intervals. Bubbles show observed study arm effects and are scaled by arm sample size.

**Supplement 90. Serious adverse events: linear estimates by risk of bias excluding azithromycin**

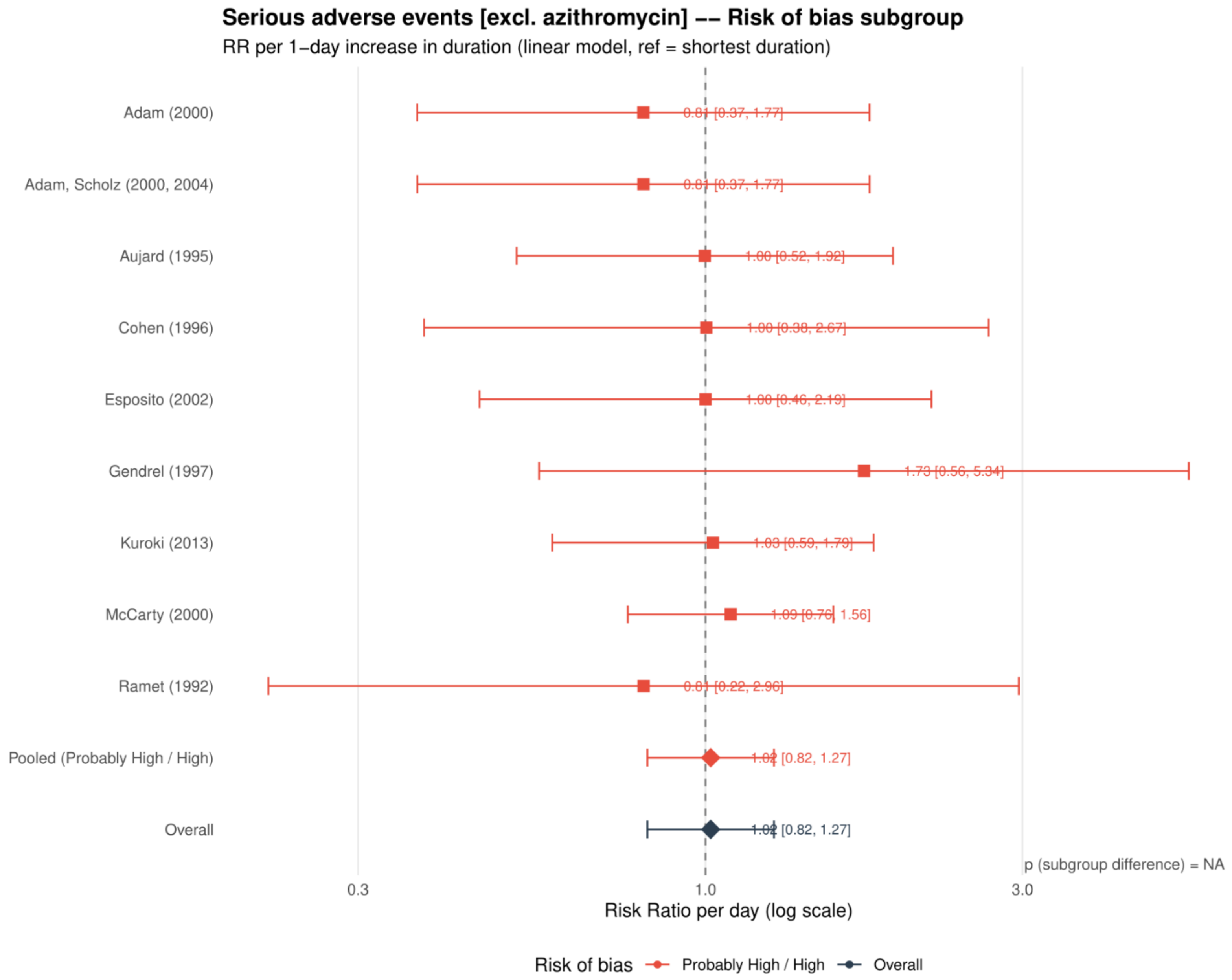


Study specific and pooled estimates are risk ratios per 1 day increase in antibiotic duration, with 95% confidence intervals.
*Only one collapsed risk of bias category contributed; the subgroup difference was not estimable.*

**Supplement 91. Serious adverse events: restricted cubic spline estimates by risk of bias excluding azithromycin**

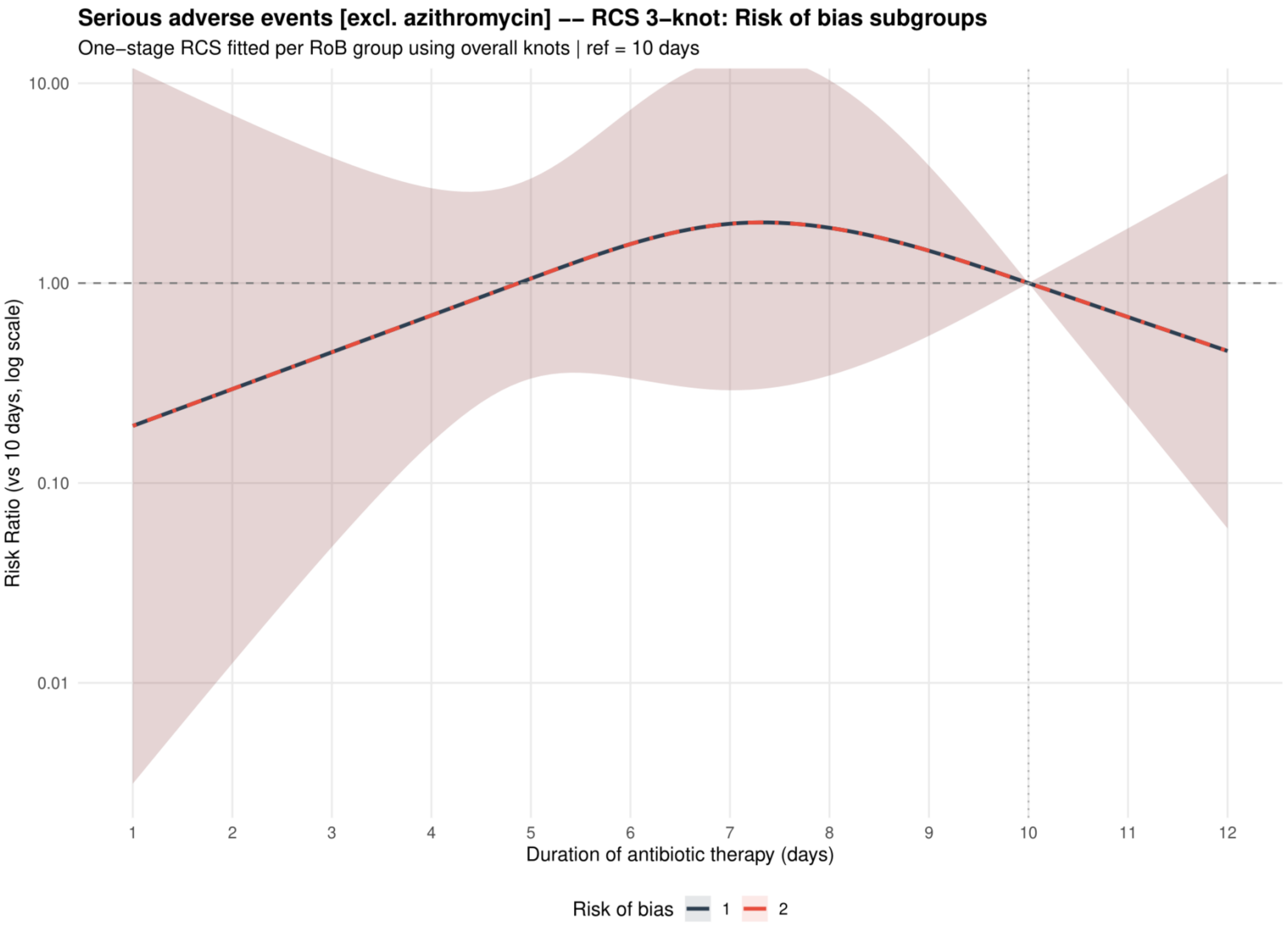


Curves show risk ratios relative to 10 days with 95% confidence intervals. Risk of bias codes: 1 = overall; 2 = low or probably low; 3 = probably high or high.

**Supplement 92. Serious adverse events: dose-response analysis excluding macrolides**

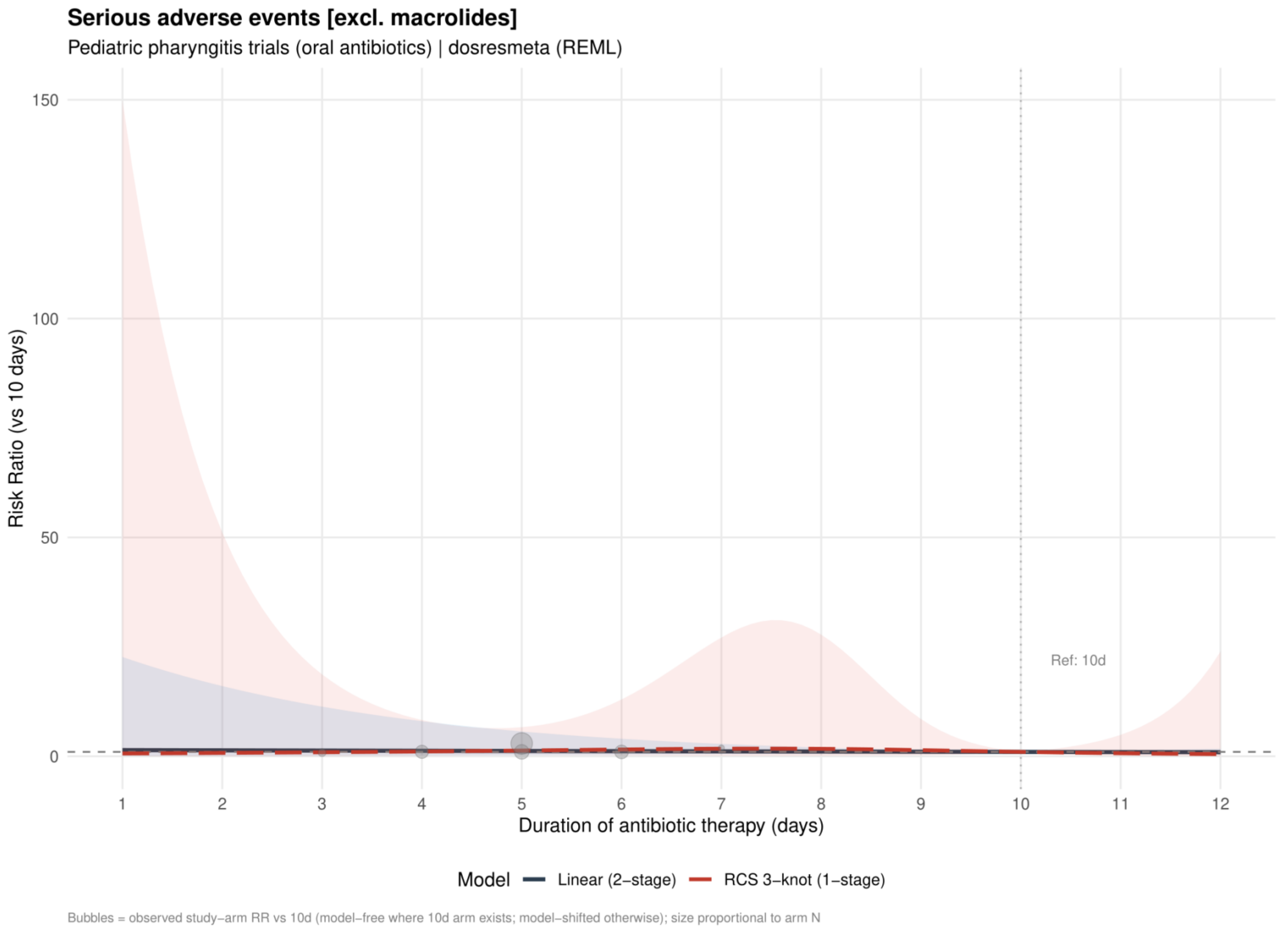


Curves show risk ratios relative to 10 days. The solid slate curve is the two stage linear model and, when present, the dashed red curve is the one stage restricted cubic spline with three knots; shaded bands show 95% confidence intervals. Bubbles show observed study arm effects and are scaled by arm sample size.

**Supplement 93. Serious adverse events: linear estimates by risk of bias excluding macrolides**

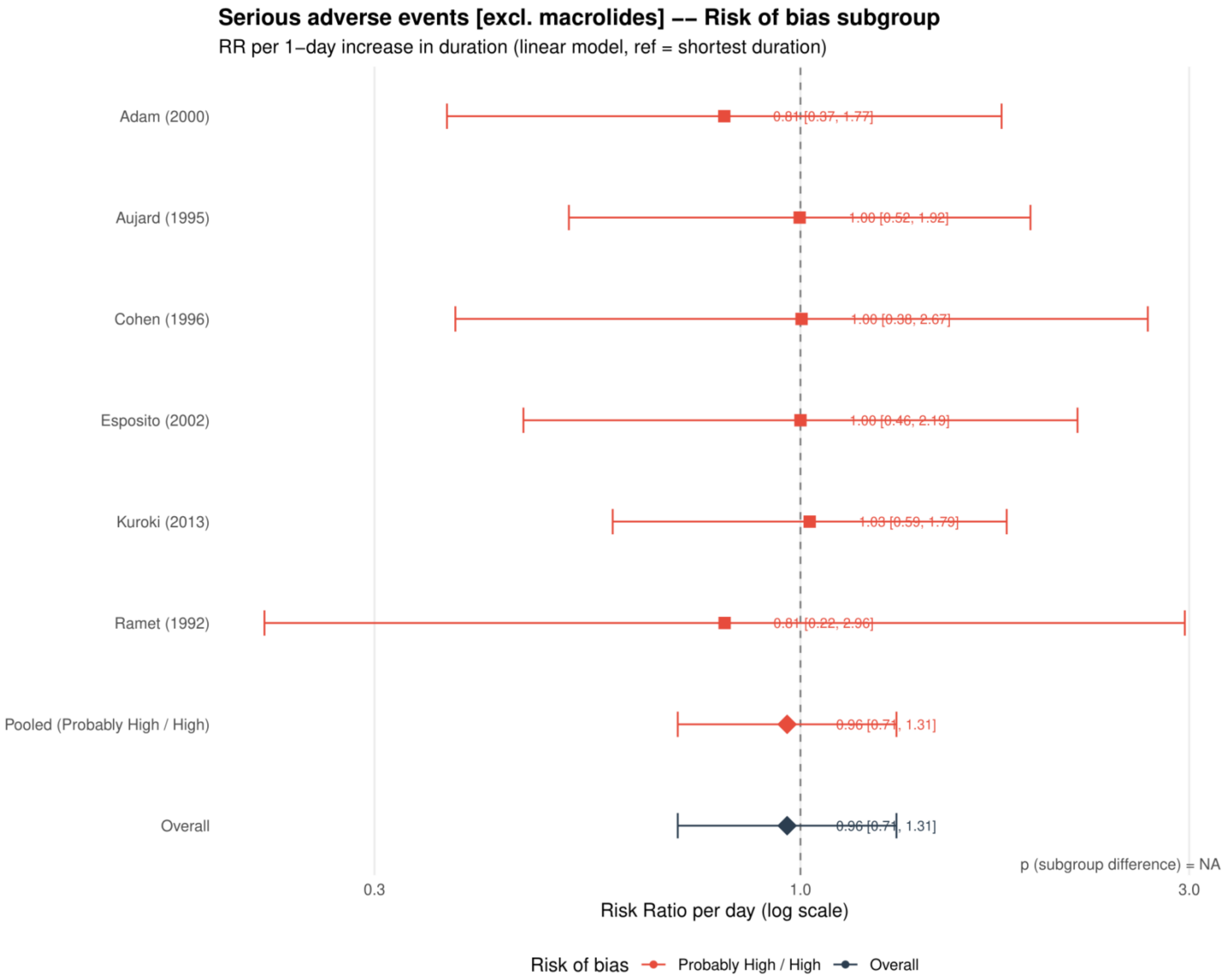


Study specific and pooled estimates are risk ratios per 1 day increase in antibiotic duration, with 95% confidence intervals.
*Only one collapsed risk of bias category contributed; the subgroup difference was not estimable.*

**Supplement 94. Serious adverse events: restricted cubic spline estimates by risk of bias excluding macrolides**

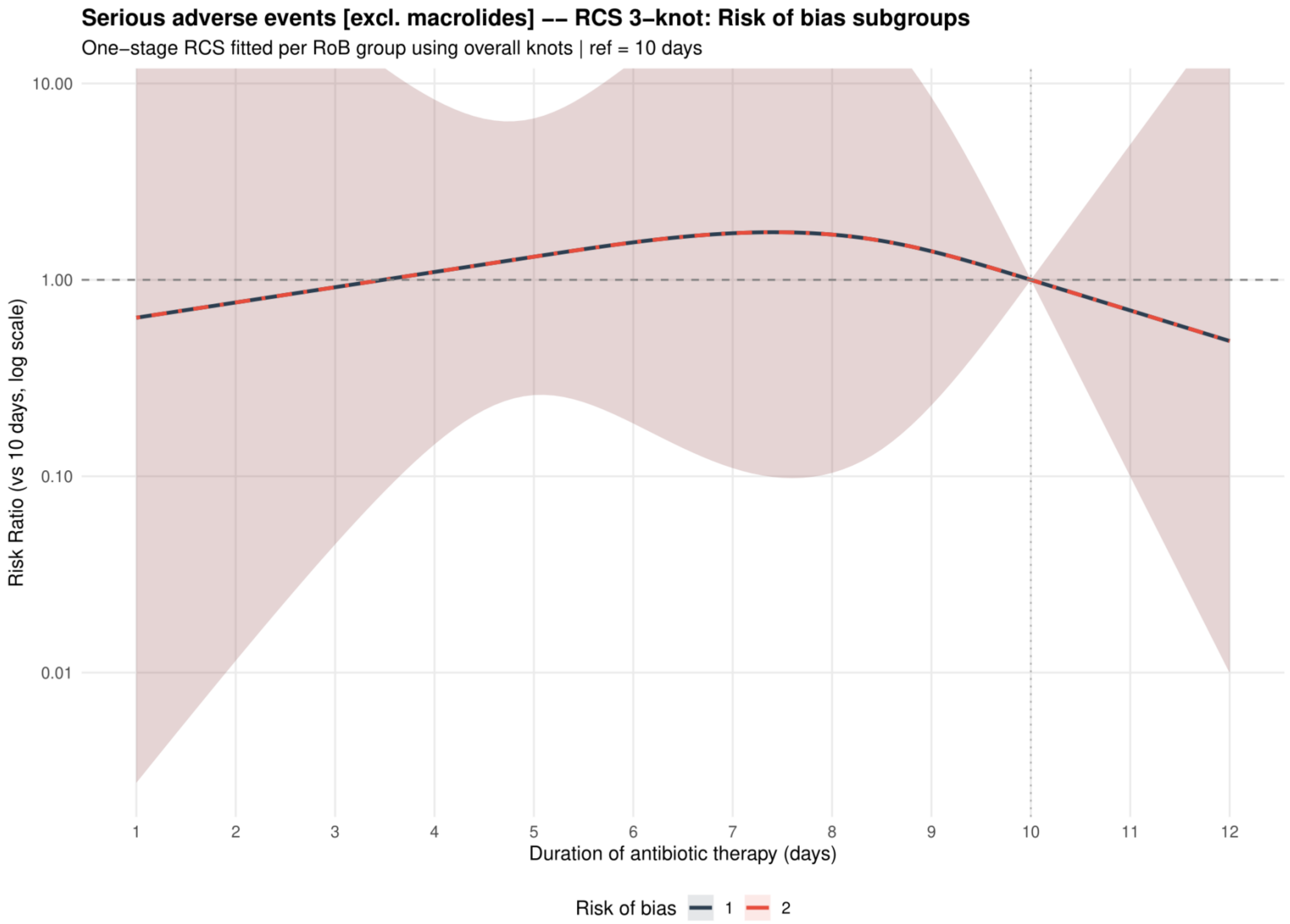


Curves show risk ratios relative to 10 days with 95% confidence intervals. Risk of bias codes: 1 = overall; 2 = low or probably low; 3 = probably high or high.

**Supplement 95. Serious adverse events: dose-response analysis including placebo or no treatment as 0 days**

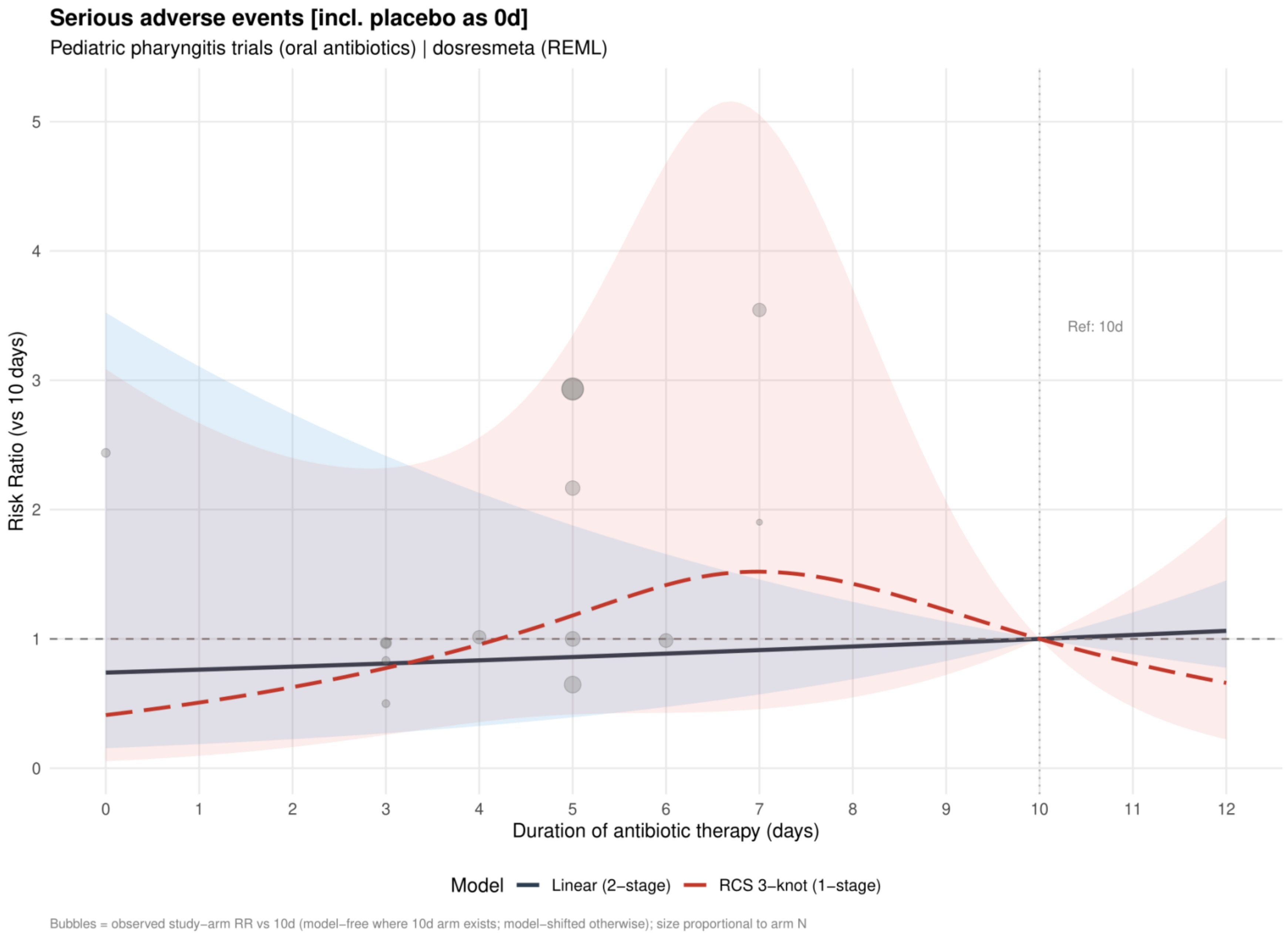


Curves show risk ratios relative to 10 days. The solid slate curve is the two stage linear model and, when present, the dashed red curve is the one stage restricted cubic spline with three knots; shaded bands show 95% confidence intervals. Bubbles show observed study arm effects and are scaled by arm sample size.

**Supplement 96. Serious adverse events: linear estimates by risk of bias including placebo or no treatment as 0 days**

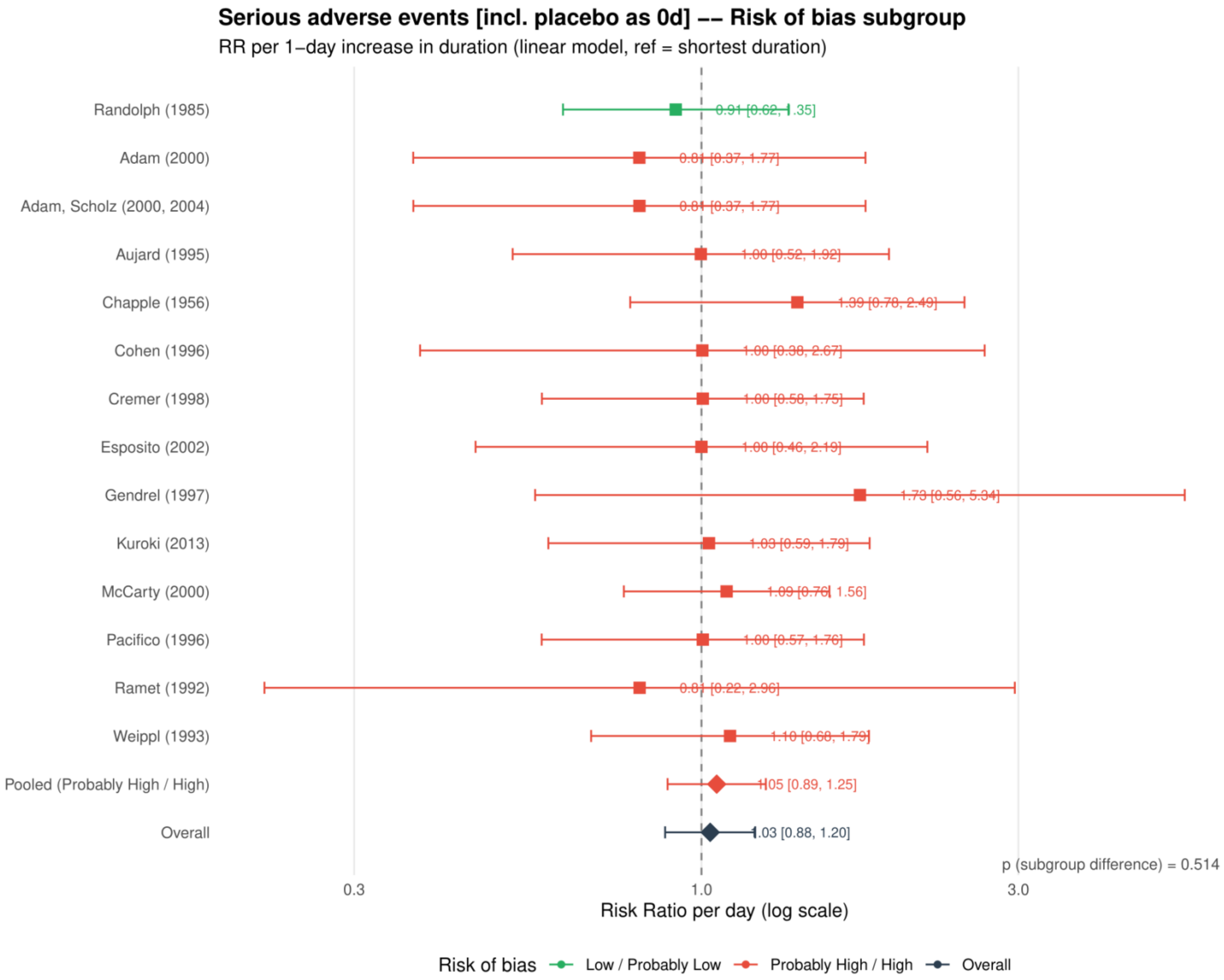


Study specific and pooled estimates are risk ratios per 1 day increase in antibiotic duration, with 95% confidence intervals.

**Supplement 97. Serious adverse events: restricted cubic spline estimates by risk of bias including placebo or no treatment as 0 days**

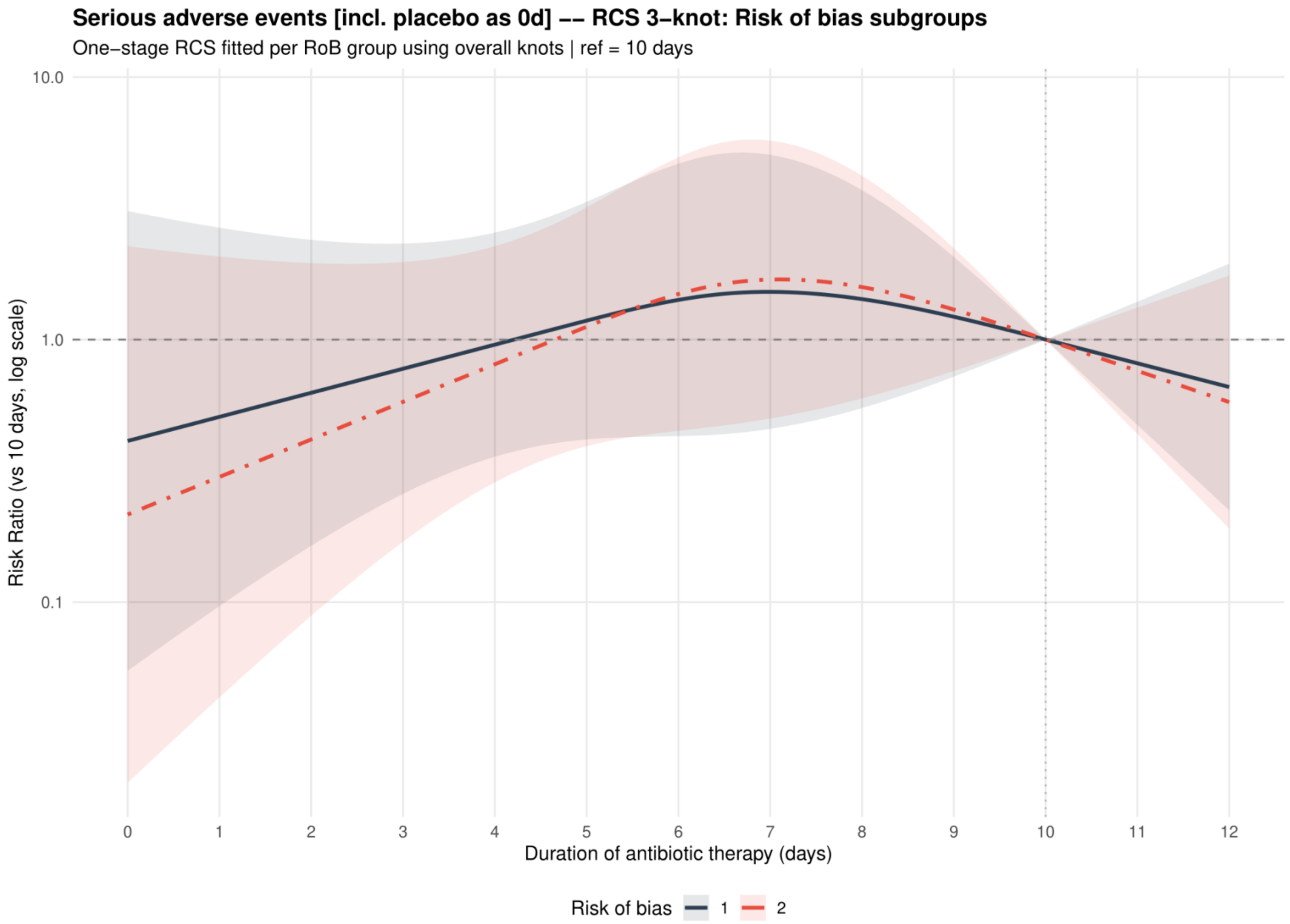


Curves show risk ratios relative to 10 days with 95% confidence intervals. Risk of bias codes: 1 = overall; 2 = low or probably low; 3 = probably high or high.

**Supplement 98. Serious adverse events: funnel plot including placebo or no treatment as 0 days**

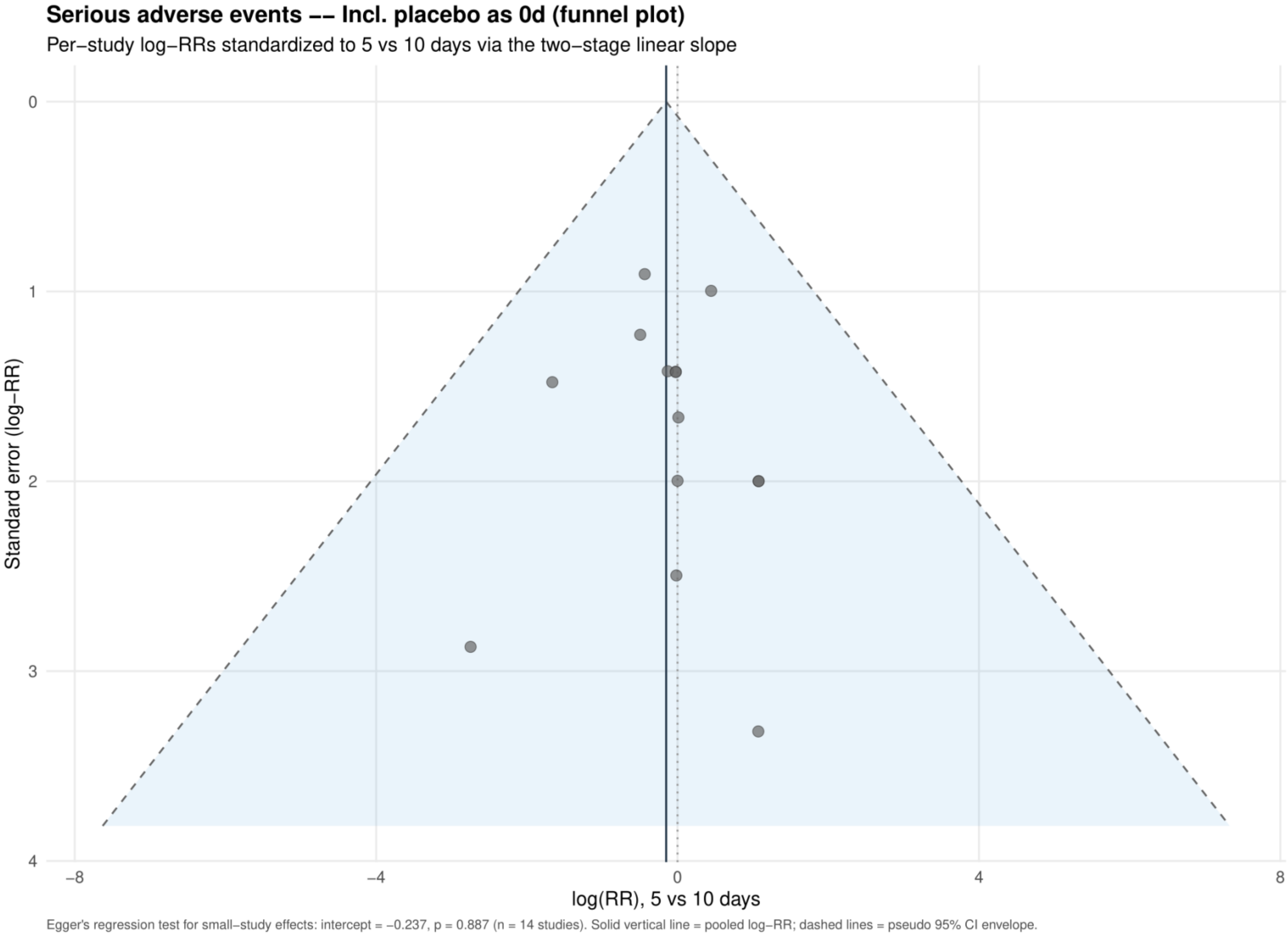


Study specific log risk ratios were standardized to 5 versus 10 days using the two stage linear slope. The solid vertical line is the pooled log risk ratio and dashed lines are pseudo 95% confidence limits; Egger's test is reported below the plot.

**Supplement 99. Acute rheumatic fever: dose-response analysis for the primary analysis**

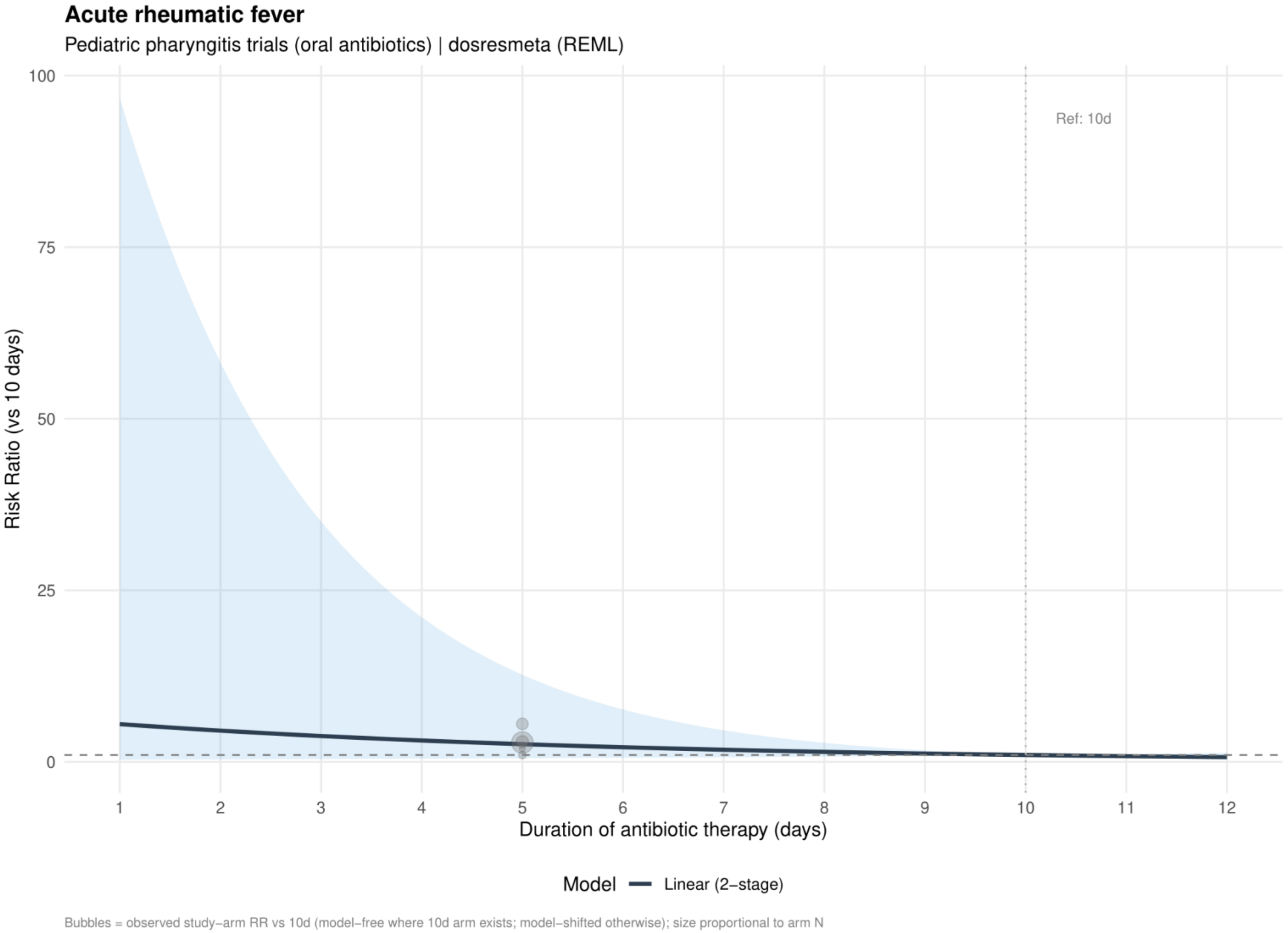


Curves show risk ratios relative to 10 days. The solid slate curve is the two stage linear model and, when present, the dashed red curve is the one stage restricted cubic spline with three knots; shaded bands show 95% confidence intervals. Bubbles show observed study arm effects and are scaled by arm sample size.

**Supplement 100. Acute rheumatic fever: linear estimates by risk of bias for the primary analysis**

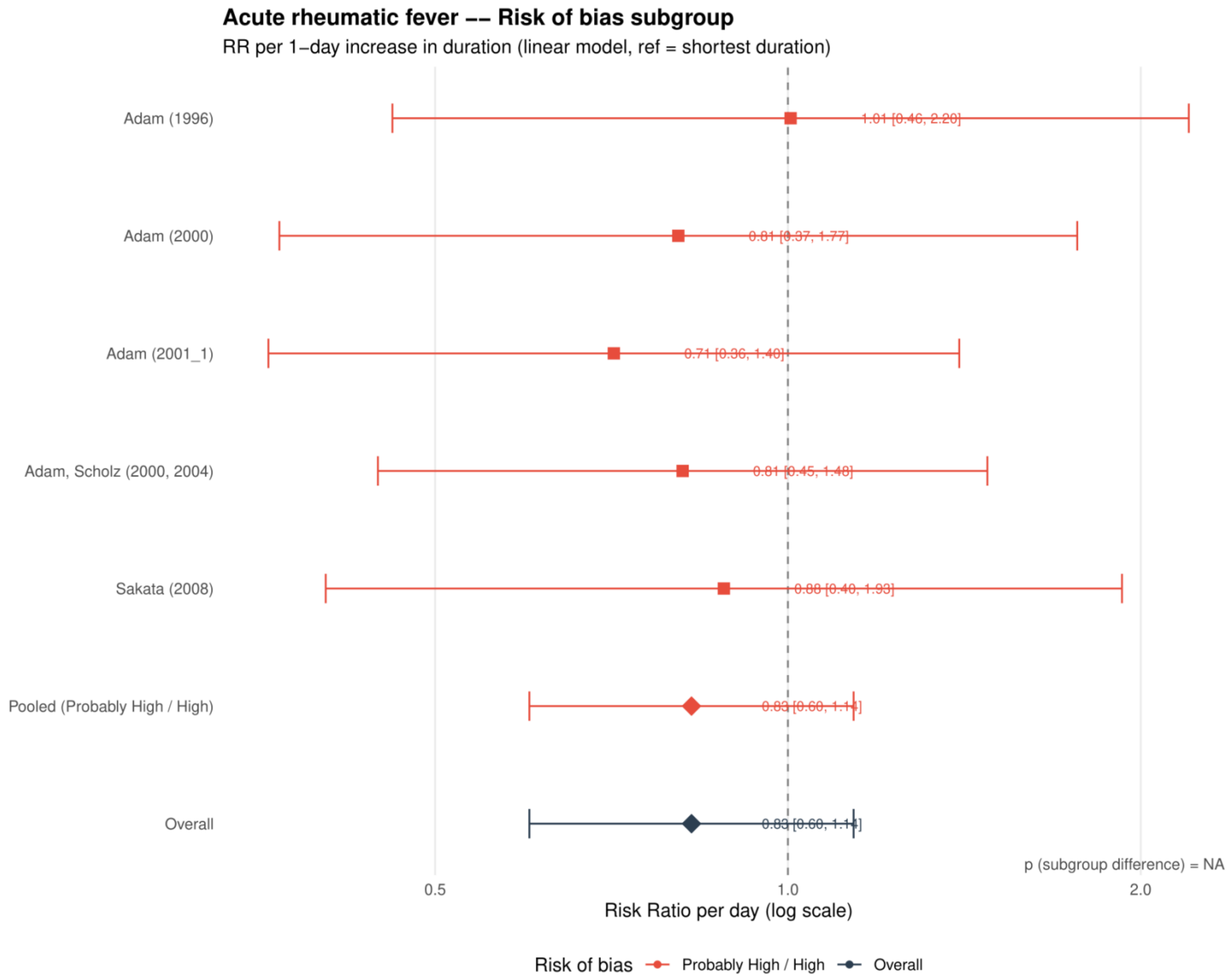


Study specific and pooled estimates are risk ratios per 1 day increase in antibiotic duration, with 95% confidence intervals.
*Only one collapsed risk of bias category contributed; the subgroup difference was not estimable.*

**Supplement 101. Acute rheumatic fever: dose-response analysis excluding azithromycin**

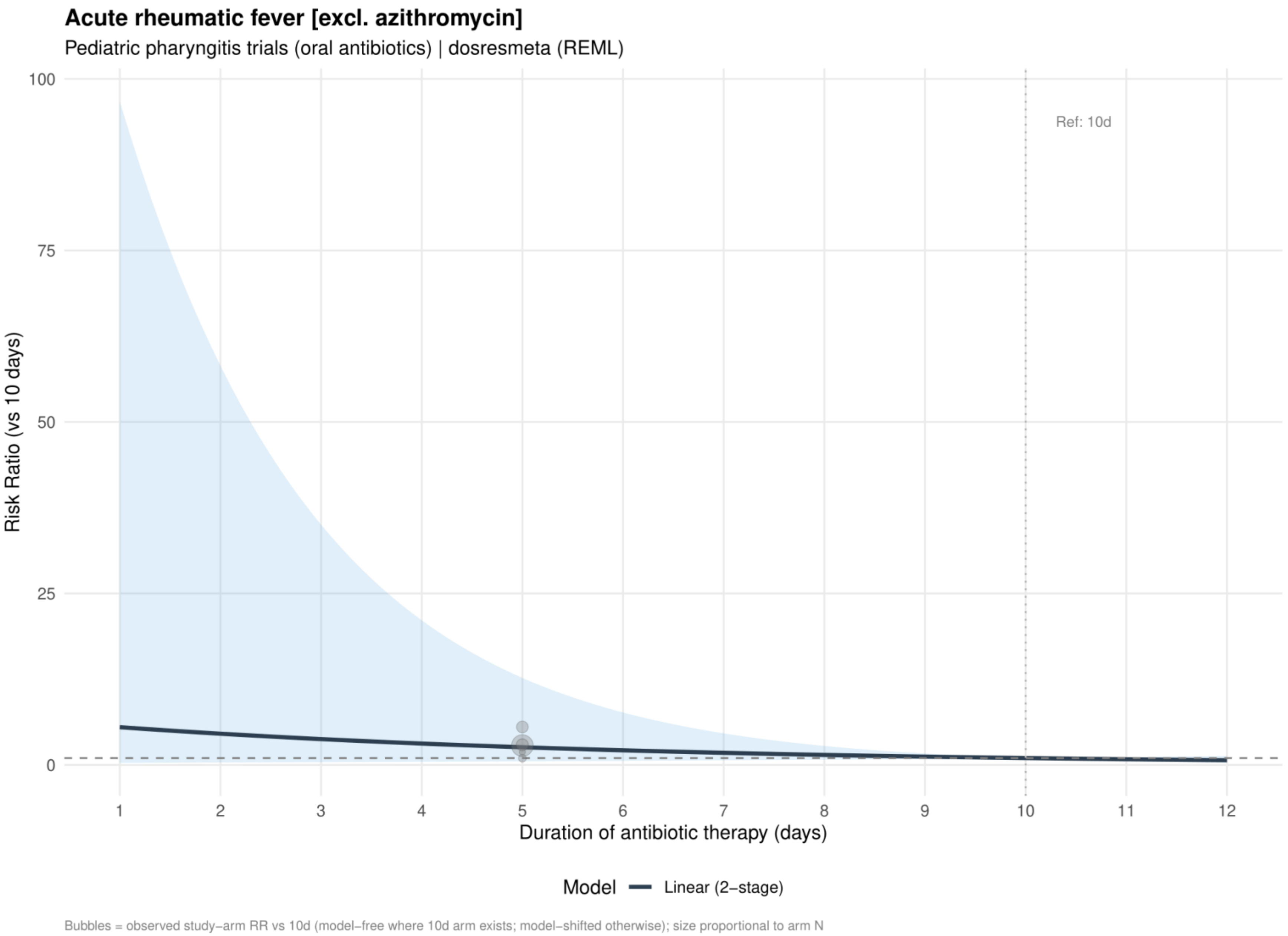


Curves show risk ratios relative to 10 days. The solid slate curve is the two stage linear model and, when present, the dashed red curve is the one stage restricted cubic spline with three knots; shaded bands show 95% confidence intervals. Bubbles show observed study arm effects and are scaled by arm sample size.
*No azithromycin trials contributed to acute rheumatic fever; this analysis is identical to Supplement 99.*

**Supplement 102. Acute rheumatic fever: linear estimates by risk of bias excluding azithromycin**

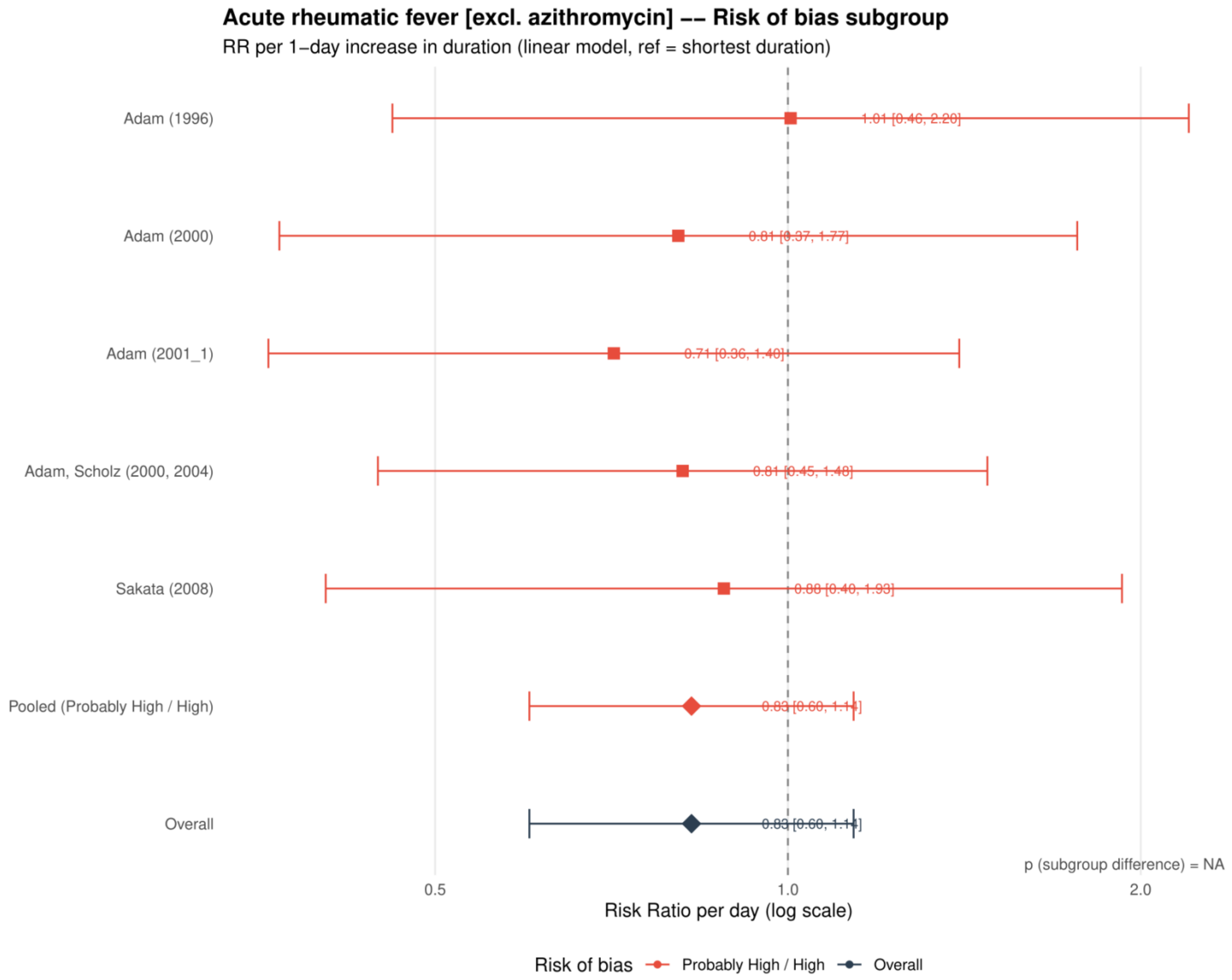


Study specific and pooled estimates are risk ratios per 1 day increase in antibiotic duration, with 95% confidence intervals.
*Only one collapsed risk of bias category contributed; the subgroup difference was not estimable.*
*No azithromycin trials contributed to acute rheumatic fever; this analysis is identical to Supplement 100.*

**Supplement 103. Acute rheumatic fever: dose-response analysis excluding macrolides**

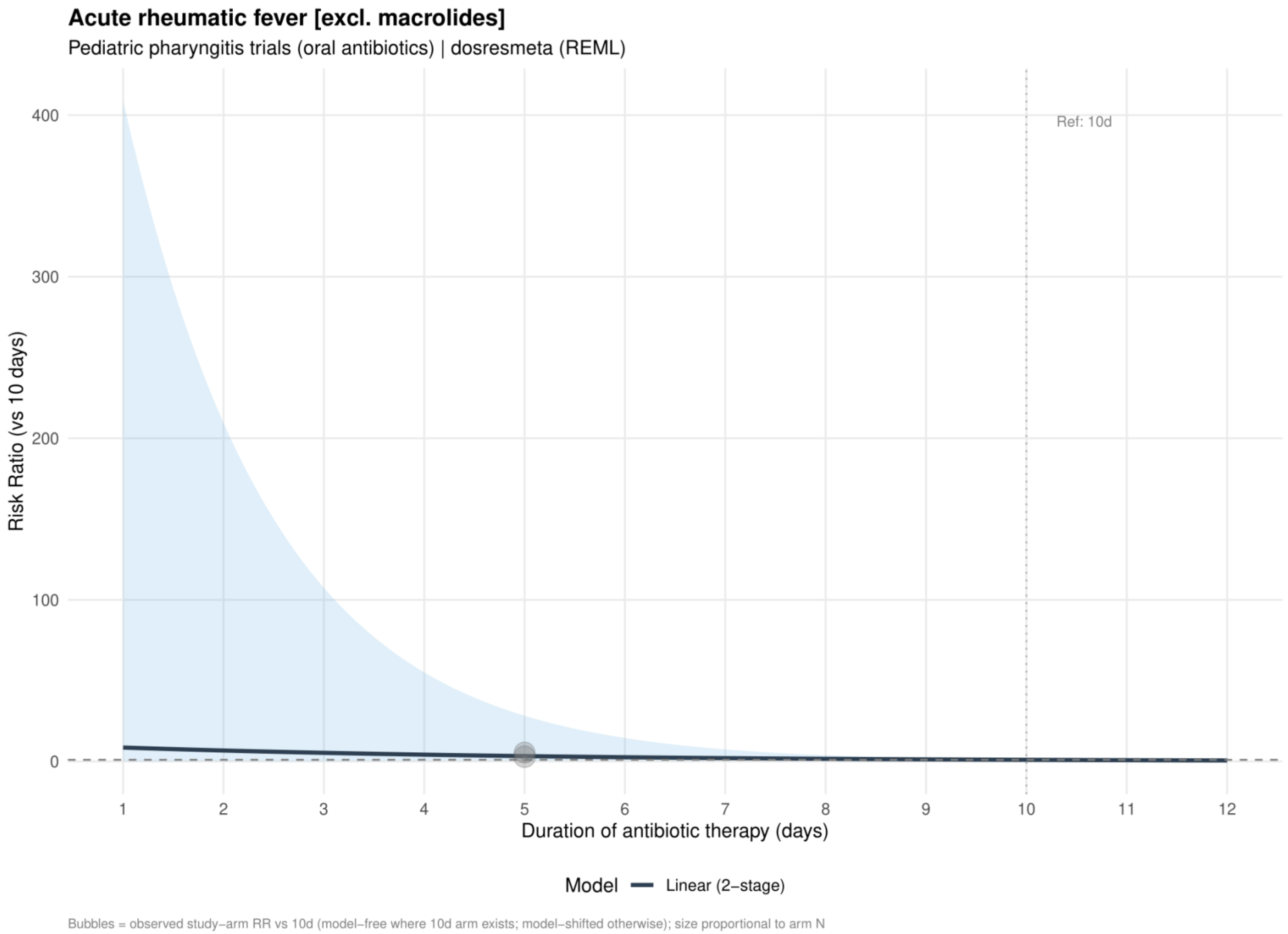


Curves show risk ratios relative to 10 days. The solid slate curve is the two stage linear model and, when present, the dashed red curve is the one stage restricted cubic spline with three knots; shaded bands show 95% confidence intervals. Bubbles show observed study arm effects and are scaled by arm sample size.

**Supplement 104. Acute rheumatic fever: linear estimates by risk of bias excluding macrolides**

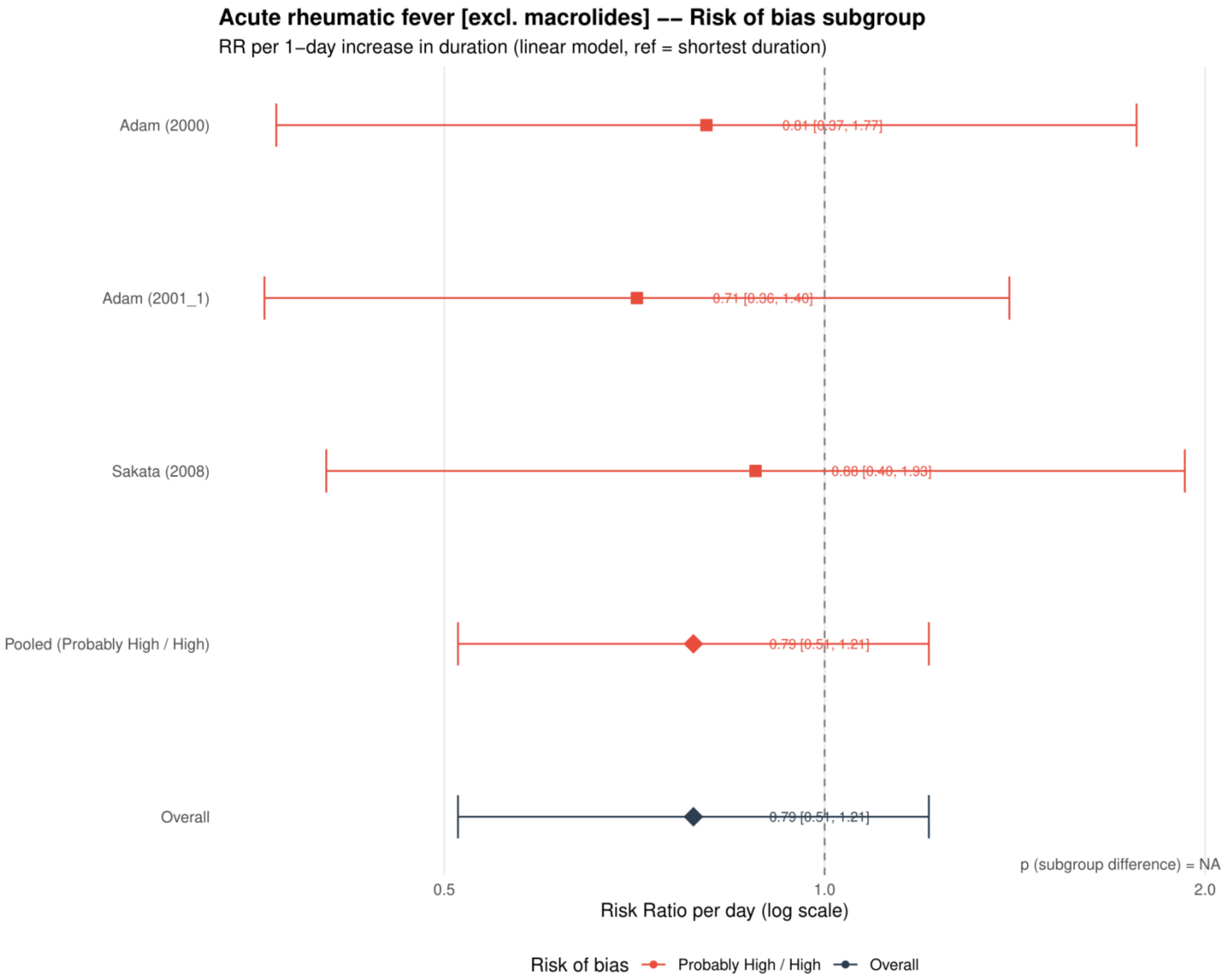


Study specific and pooled estimates are risk ratios per 1 day increase in antibiotic duration, with 95% confidence intervals.
*Only one collapsed risk of bias category contributed; the subgroup difference was not estimable.*

**Supplement 105. Acute rheumatic fever: dose-response analysis including placebo or no treatment as 0 days**

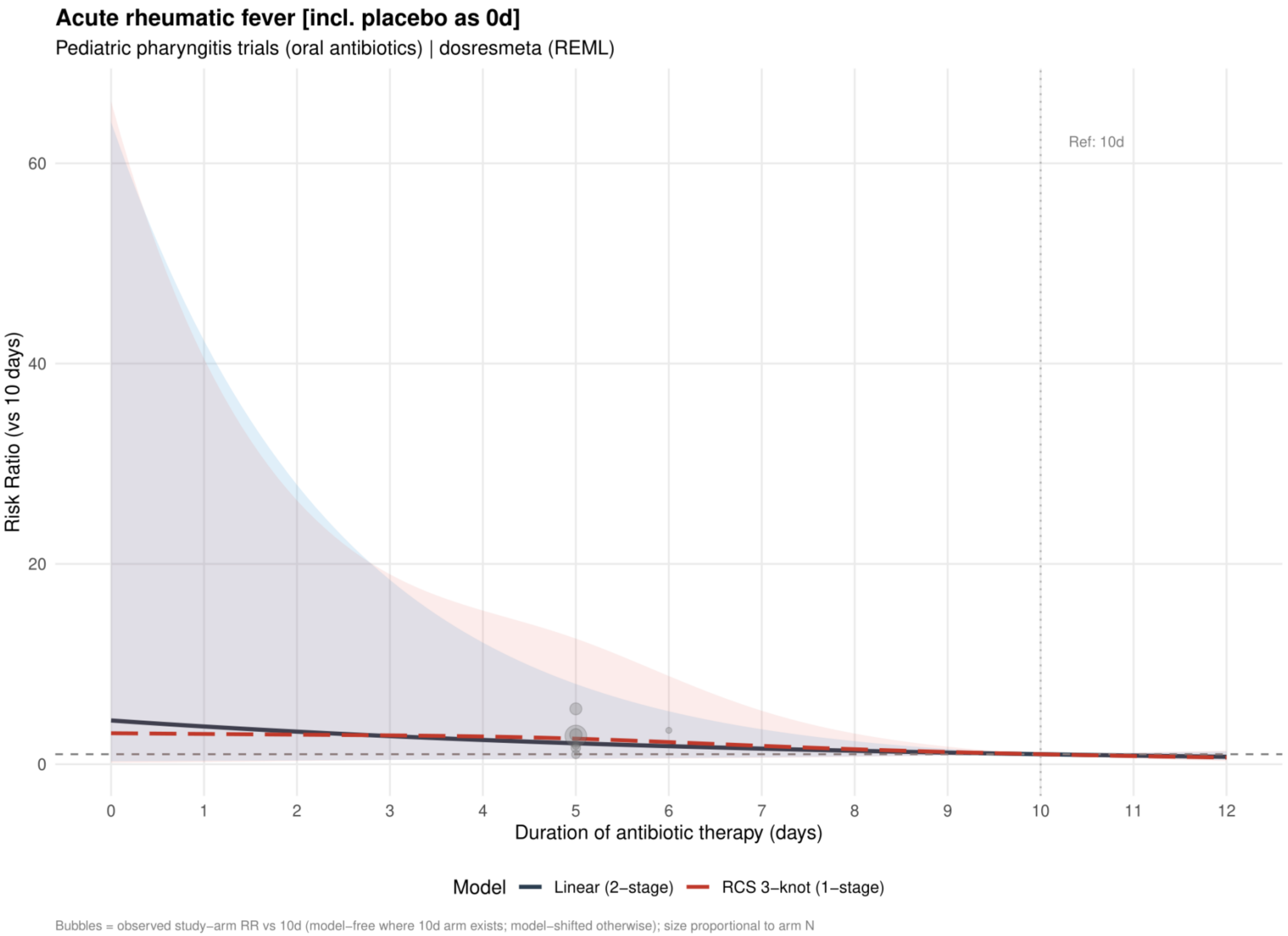


Curves show risk ratios relative to 10 days. The solid slate curve is the two stage linear model and, when present, the dashed red curve is the one stage restricted cubic spline with three knots; shaded bands show 95% confidence intervals. Bubbles show observed study arm effects and are scaled by arm sample size.

**Supplement 106. Acute rheumatic fever: linear estimates by risk of bias including placebo or no treatment as 0 days**

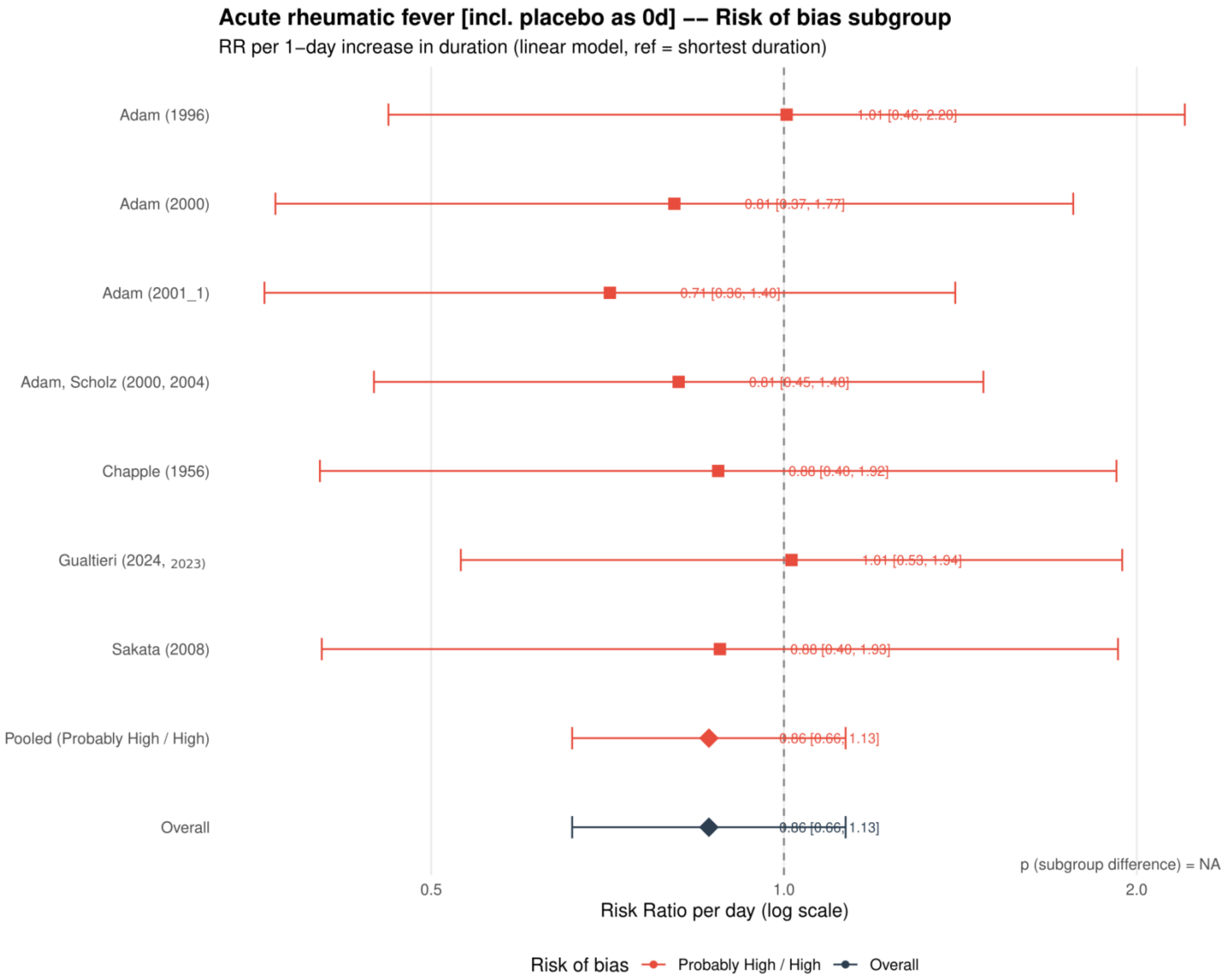


Study specific and pooled estimates are risk ratios per 1 day increase in antibiotic duration, with 95% confidence intervals.
*Only one collapsed risk of bias category contributed; the subgroup difference was not estimable.*

**Supplement 107. Acute rheumatic fever: restricted cubic spline estimates by risk of bias including placebo or no treatment as 0 days**

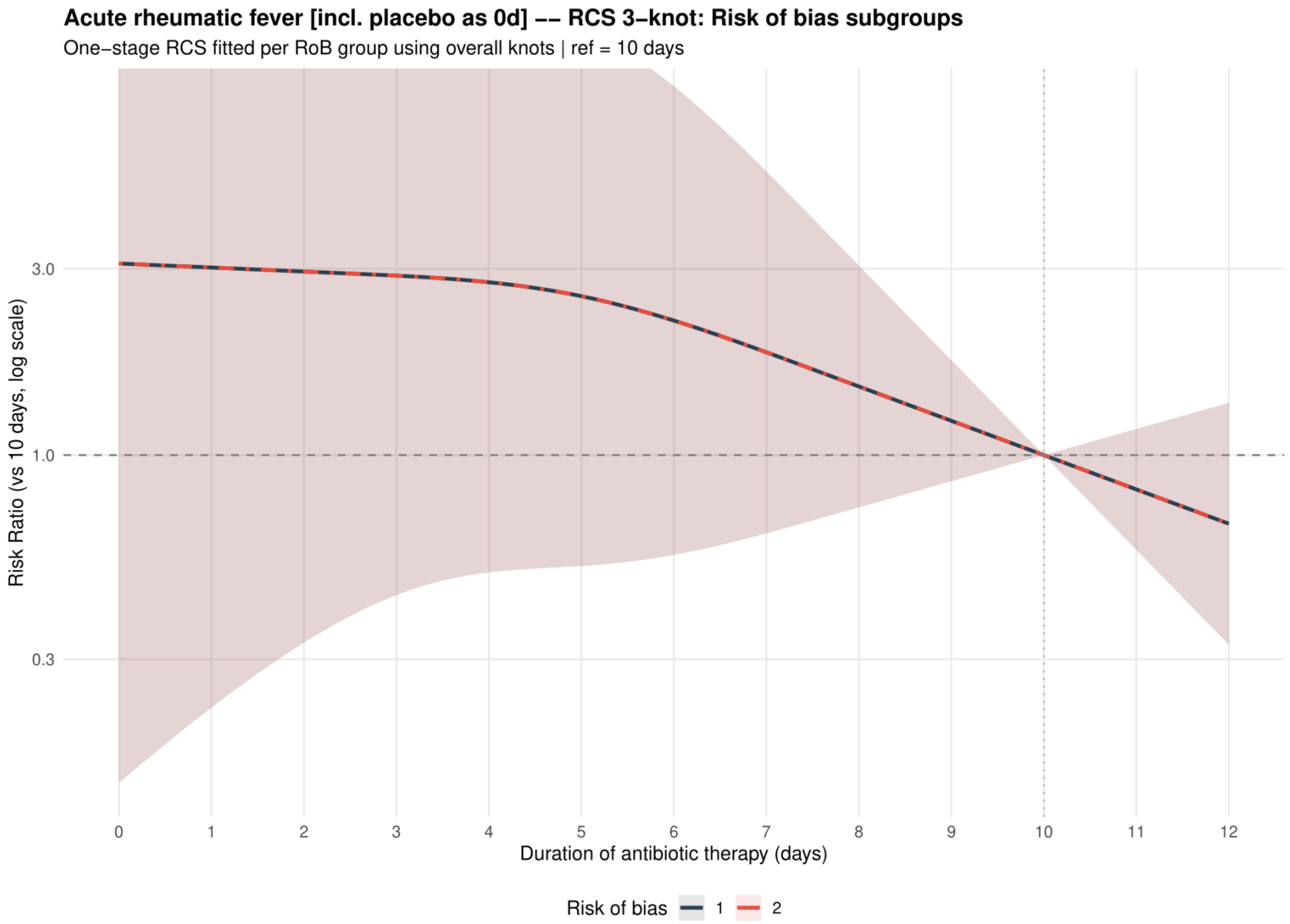


Curves show risk ratios relative to 10 days with 95% confidence intervals. Risk of bias codes: 1 = overall; 2 = low or probably low; 3 = probably high or high.

**Supplement 108. Purulent complications: dose-response analysis for the primary analysis**

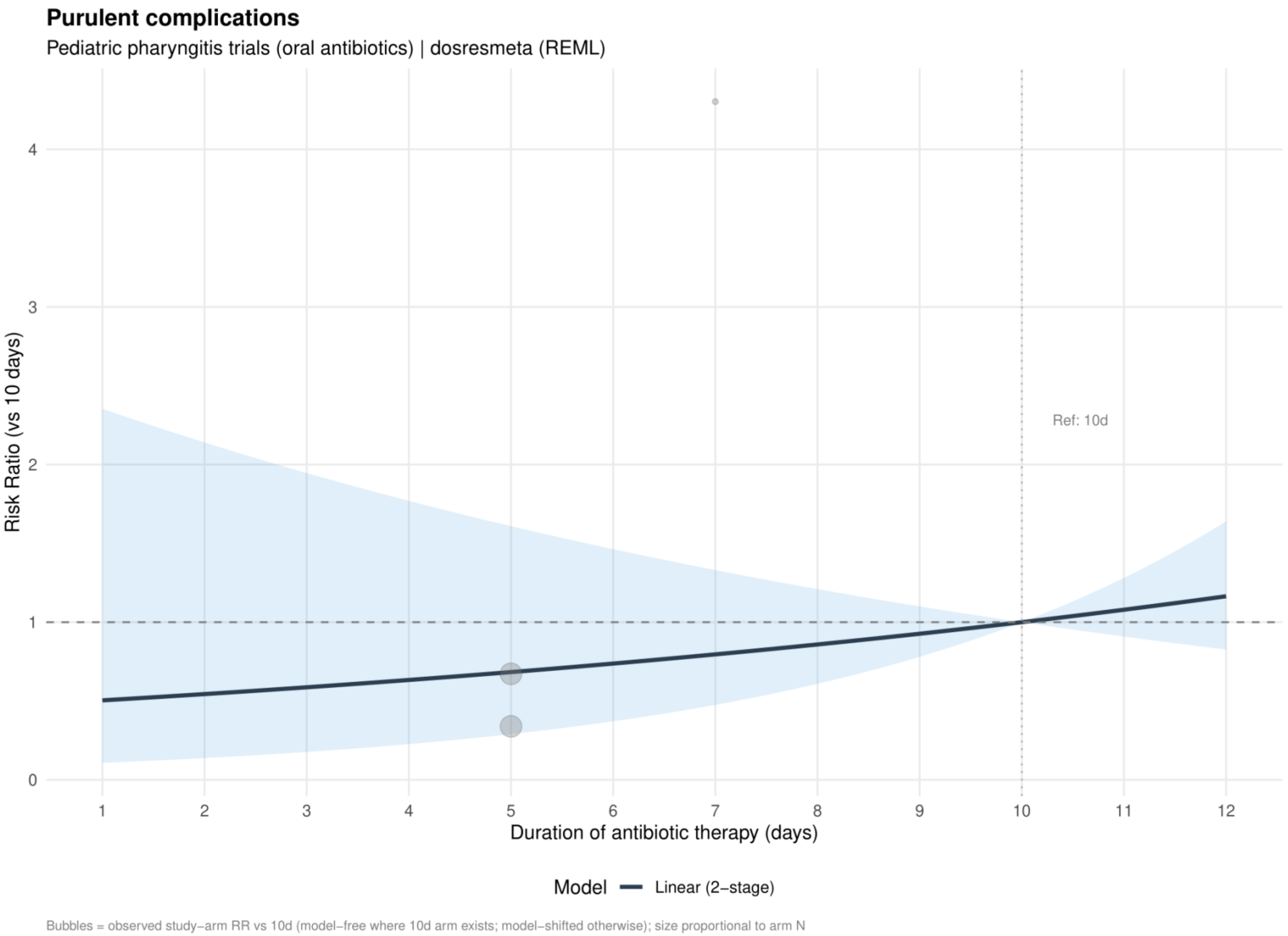


Curves show risk ratios relative to 10 days. The solid slate curve is the two stage linear model and, when present, the dashed red curve is the one stage restricted cubic spline with three knots; shaded bands show 95% confidence intervals. Bubbles show observed study arm effects and are scaled by arm sample size.

**Supplement 109. Purulent complications: linear estimates by risk of bias for the primary analysis**

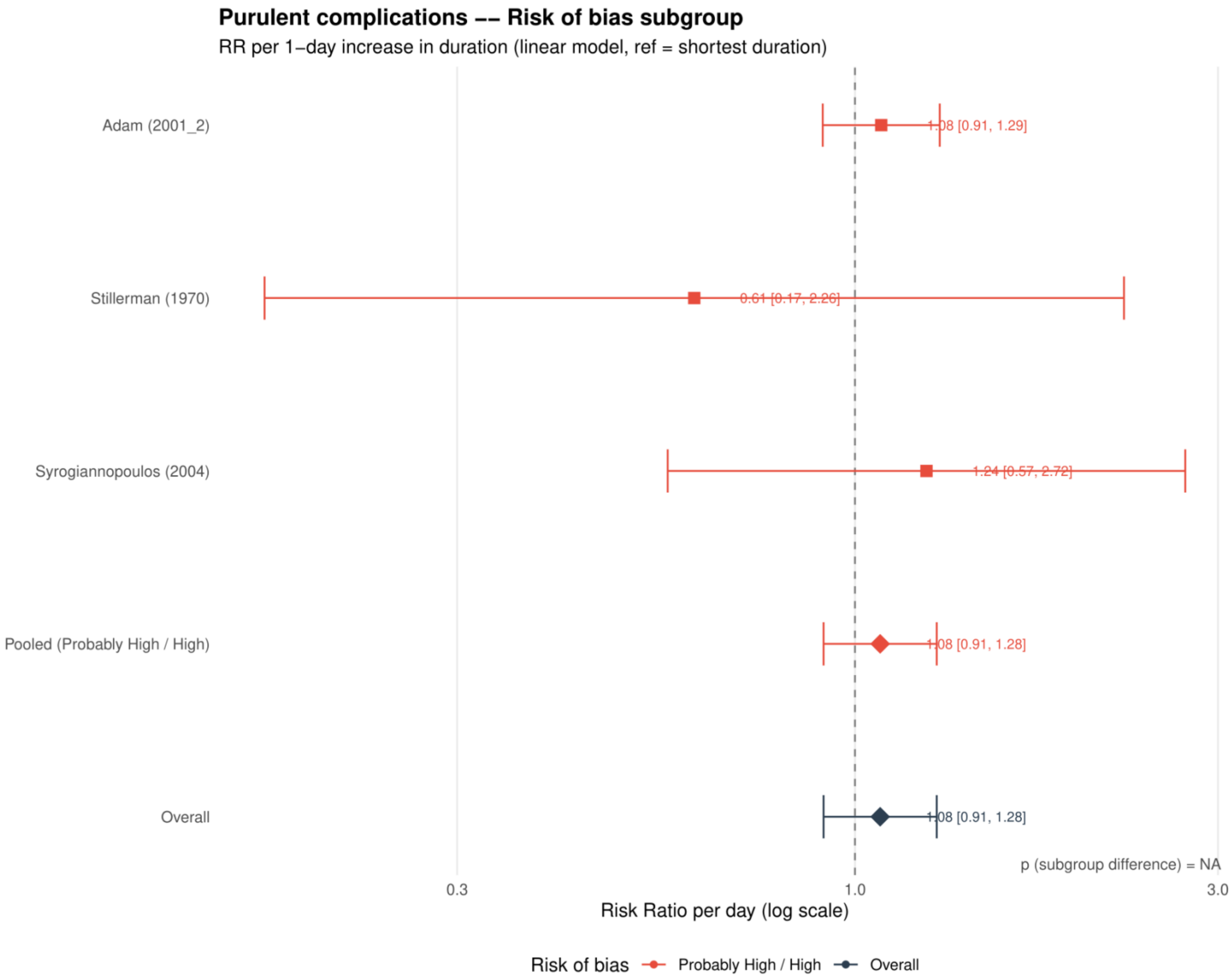


Study specific and pooled estimates are risk ratios per 1 day increase in antibiotic duration, with 95% confidence intervals.
*Only one collapsed risk of bias category contributed; the subgroup difference was not estimable.*

**Supplement 110. Purulent complications: dose-response analysis excluding azithromycin**

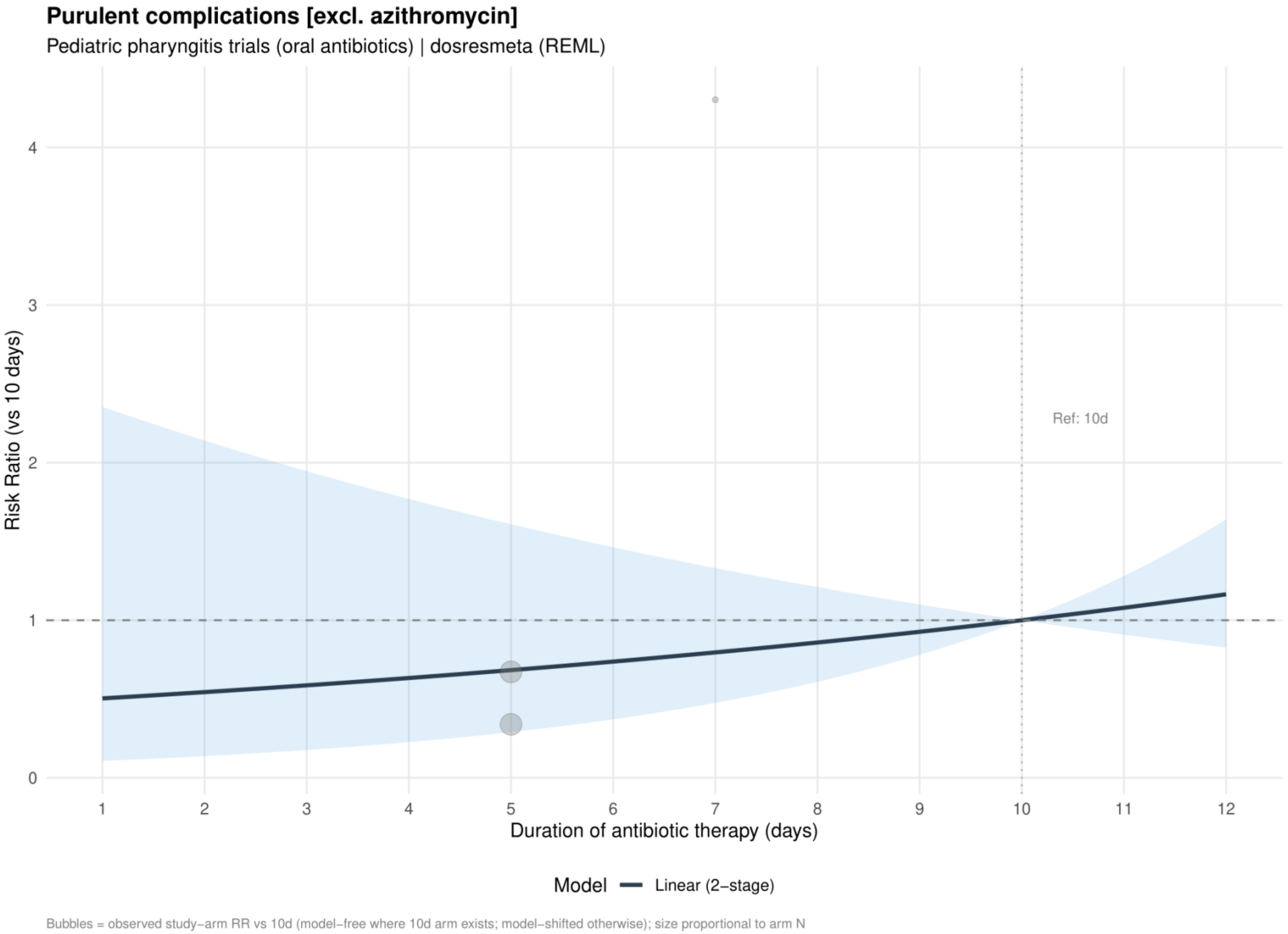


Curves show risk ratios relative to 10 days. The solid slate curve is the two stage linear model and, when present, the dashed red curve is the one stage restricted cubic spline with three knots; shaded bands show 95% confidence intervals. Bubbles show observed study arm effects and are scaled by arm sample size.
*No azithromycin trials contributed to purulent complications; this analysis is identical to Supplement 108.*

**Supplement 111. Purulent complications: linear estimates by risk of bias excluding azithromycin**

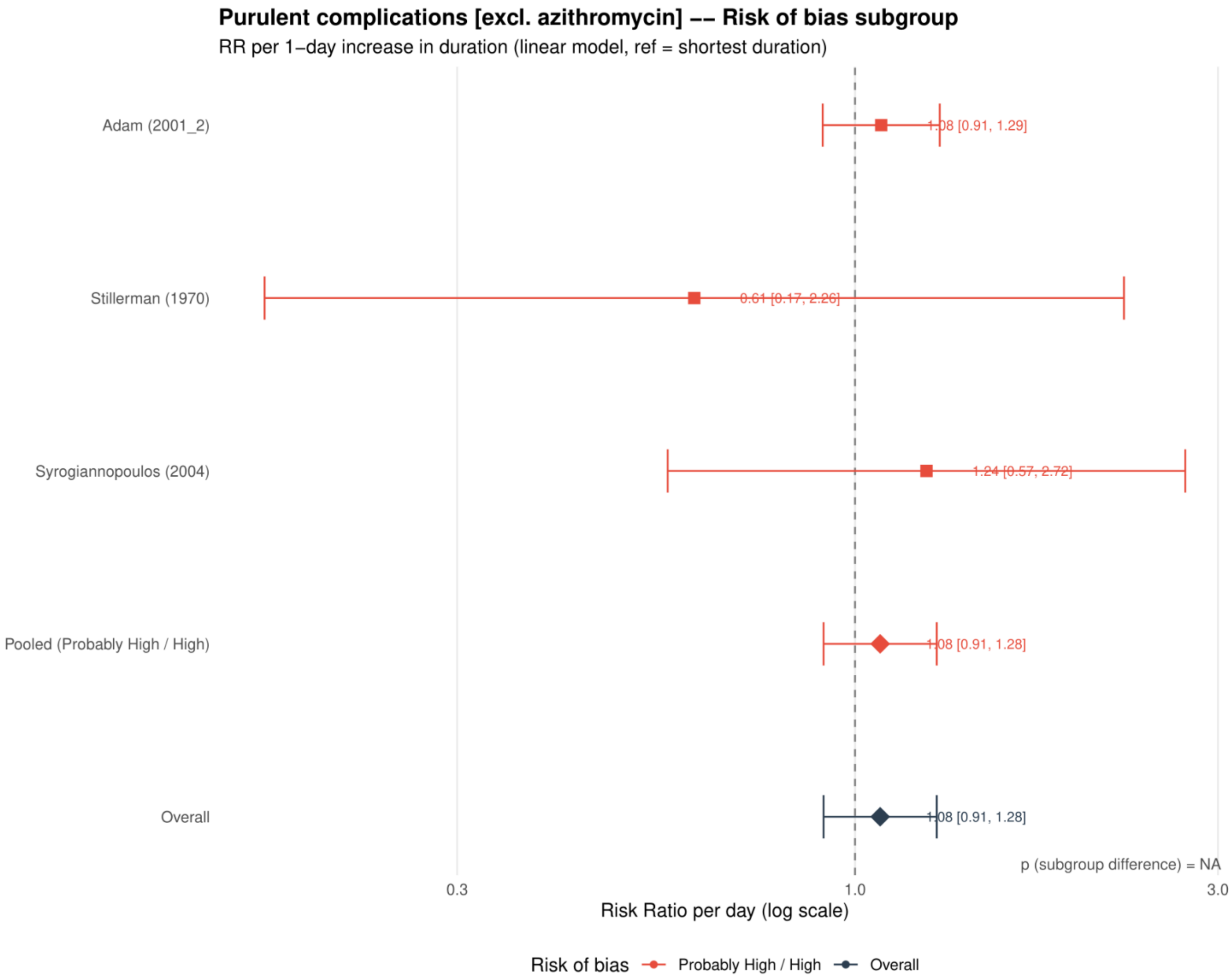


Study specific and pooled estimates are risk ratios per 1 day increase in antibiotic duration, with 95% confidence intervals.
*Only one collapsed risk of bias category contributed; the subgroup difference was not estimable.*
*No azithromycin trials contributed to purulent complications; this analysis is identical to Supplement 109.*

**Supplement 112. Purulent complications: dose-response analysis excluding macrolides**

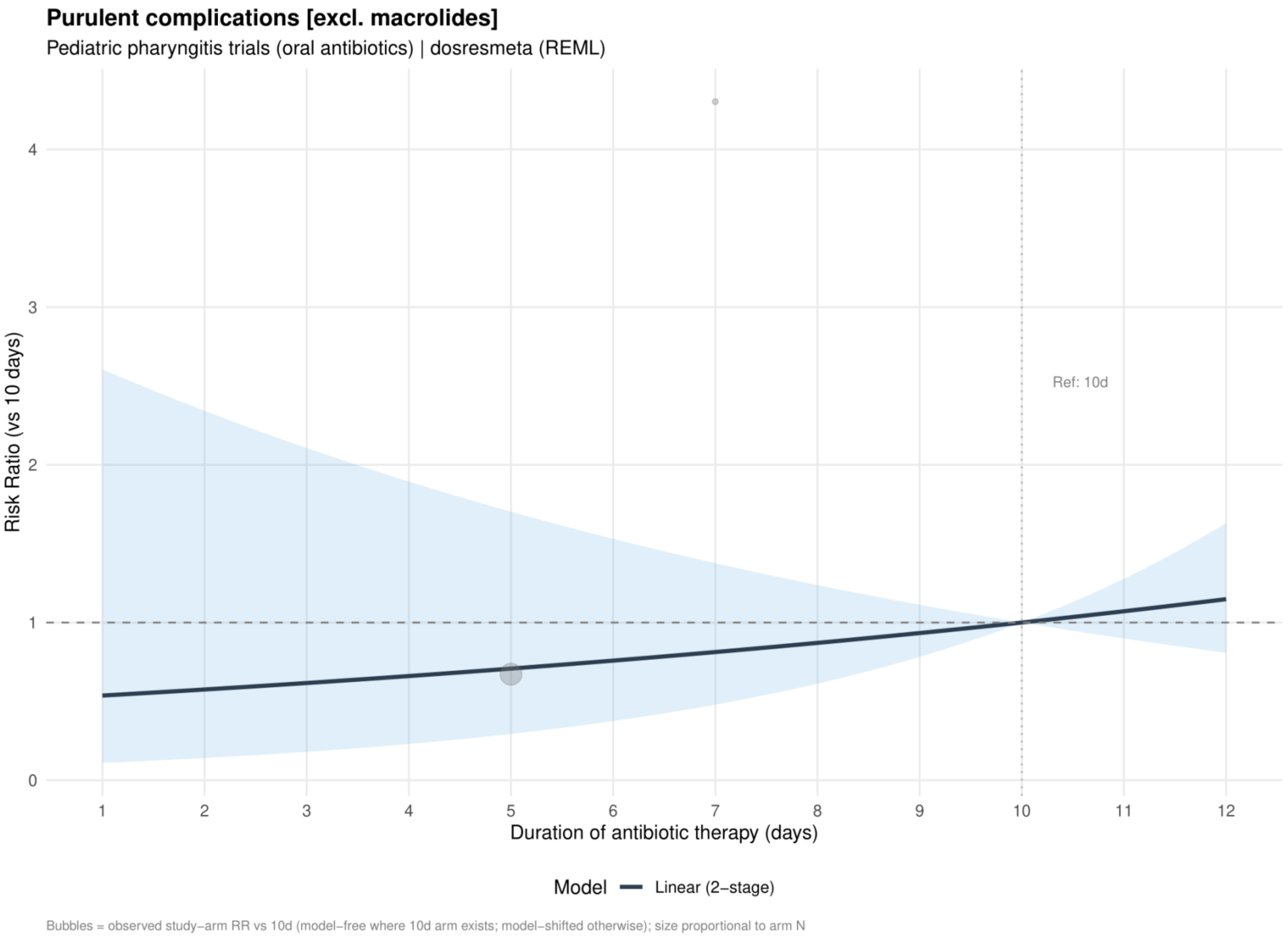


Curves show risk ratios relative to 10 days. The solid slate curve is the two stage linear model and, when present, the dashed red curve is the one stage restricted cubic spline with three knots; shaded bands show 95% confidence intervals. Bubbles show observed study arm effects and are scaled by arm sample size.

**Supplement 113. Purulent complications: linear estimates by risk of bias excluding macrolides**

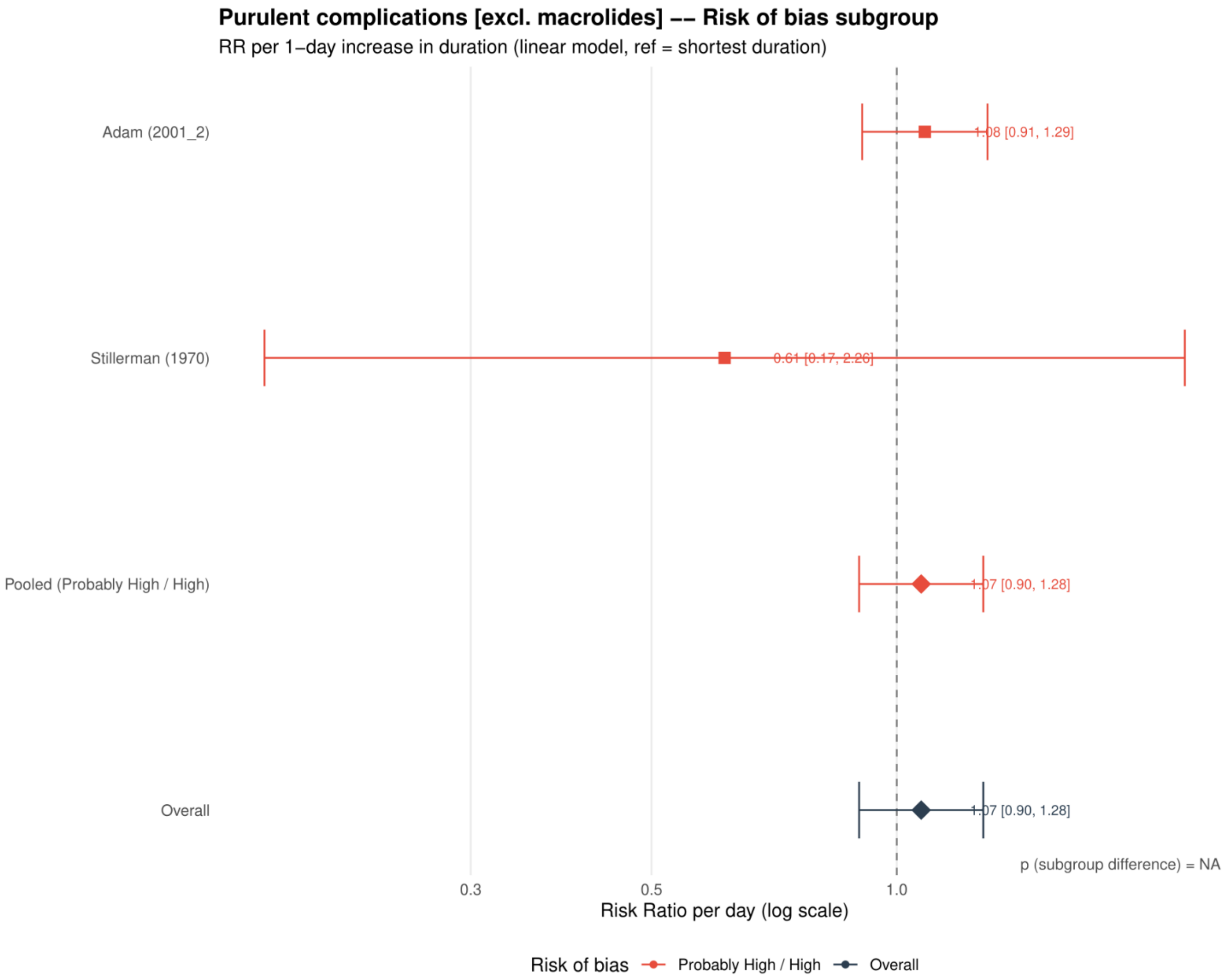


Study specific and pooled estimates are risk ratios per 1 day increase in antibiotic duration, with 95% confidence intervals.
*Only one collapsed risk of bias category contributed; the subgroup difference was not estimable.*

**Supplement 114. Purulent complications: dose-response analysis including placebo or no treatment as 0 days**

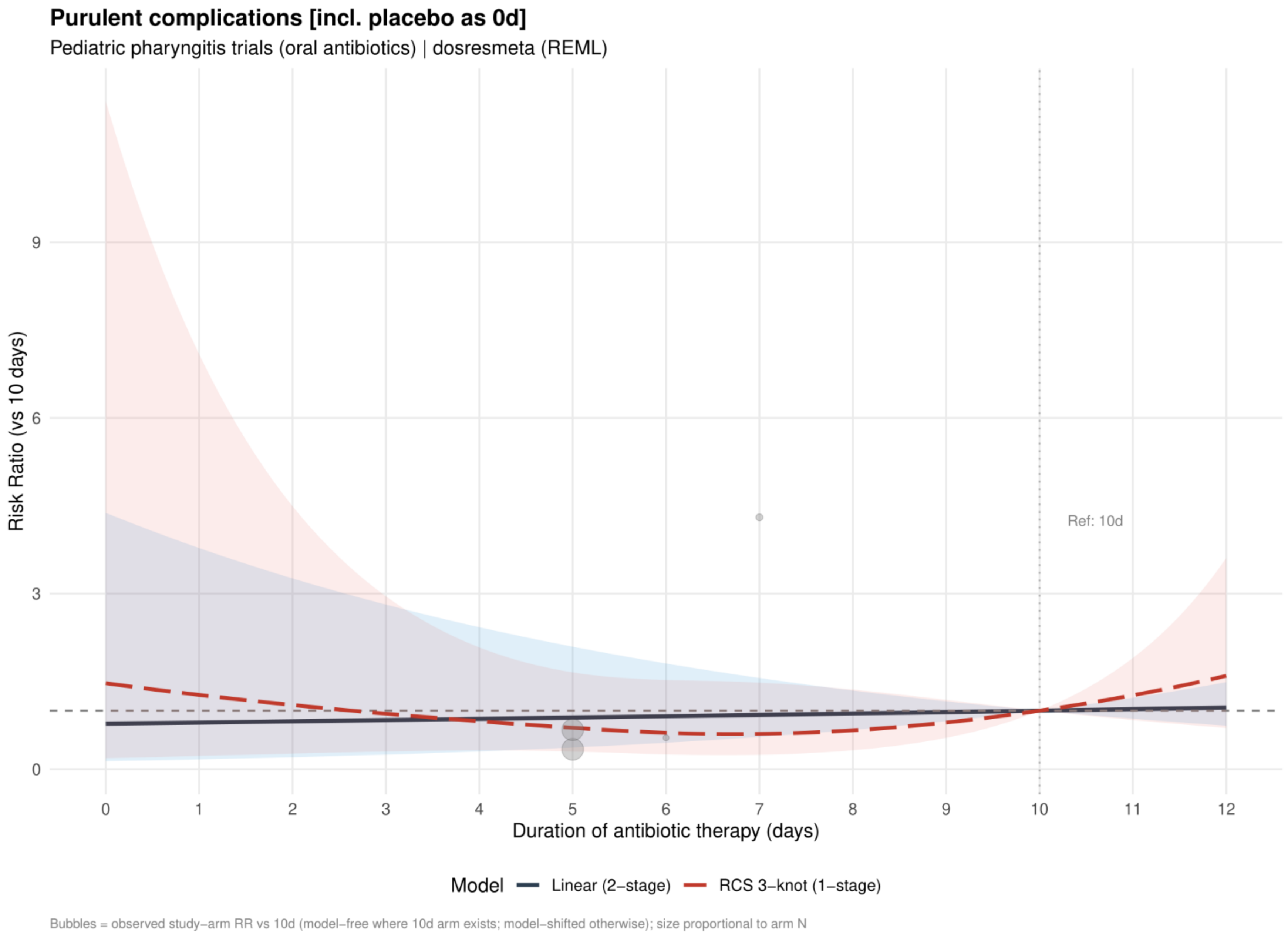


Curves show risk ratios relative to 10 days. The solid slate curve is the two stage linear model and, when present, the dashed red curve is the one stage restricted cubic spline with three knots; shaded bands show 95% confidence intervals. Bubbles show observed study arm effects and are scaled by arm sample size.

**Supplement 115. Purulent complications: linear estimates by risk of bias including placebo or no treatment as 0 days**

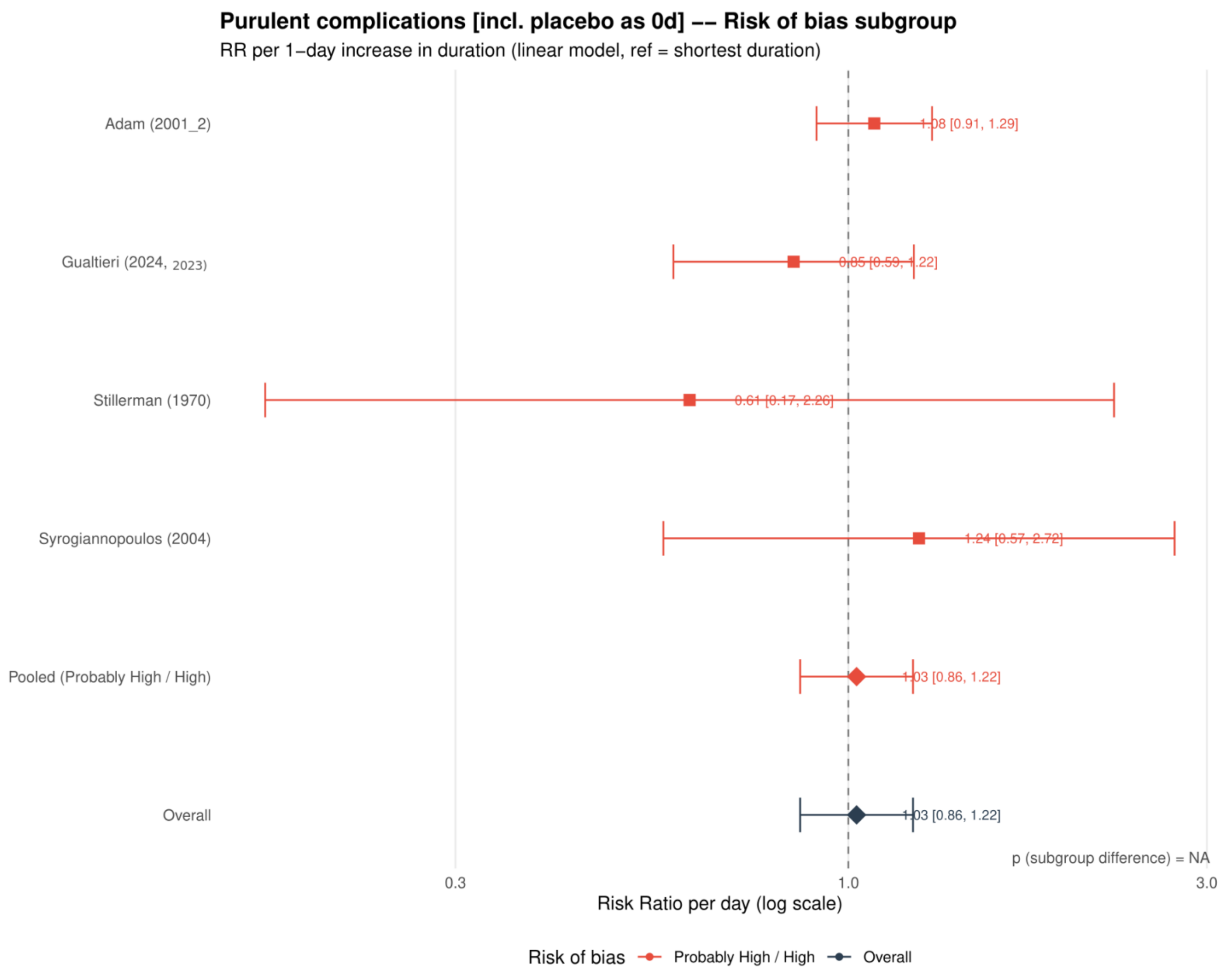


Study specific and pooled estimates are risk ratios per 1 day increase in antibiotic duration, with 95% confidence intervals.
*Only one collapsed risk of bias category contributed; the subgroup difference was not estimable.*

**Supplement 116. Purulent complications: restricted cubic spline estimates by risk of bias including placebo or no treatment as 0 days**

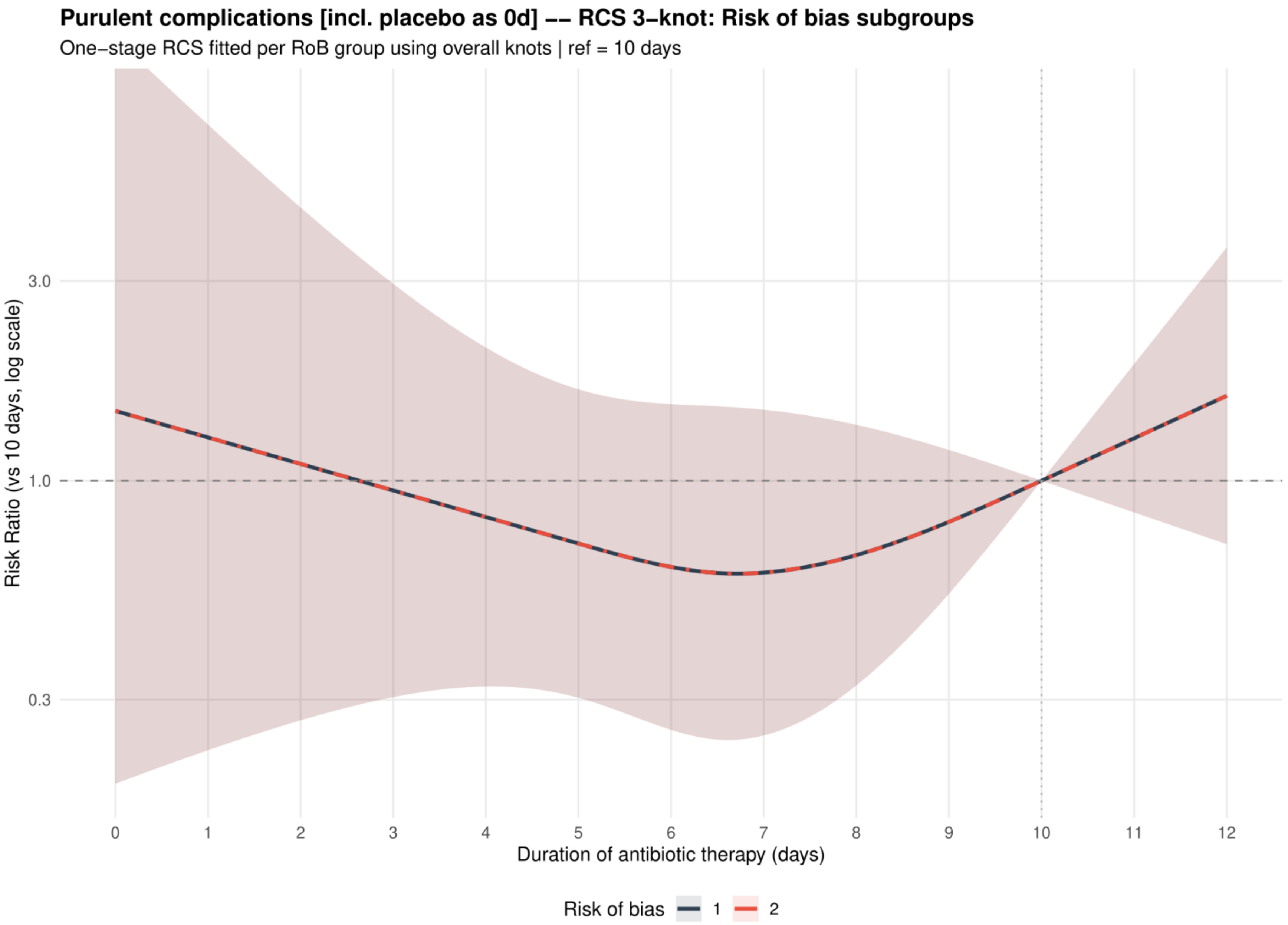


Curves show risk ratios relative to 10 days with 95% confidence intervals. Risk of bias codes: 1 = overall; 2 = low or probably low; 3 = probably high or high.

**Supplement 117. Poststreptococcal glomerulonephritis: dose-response analysis for the primary analysis**

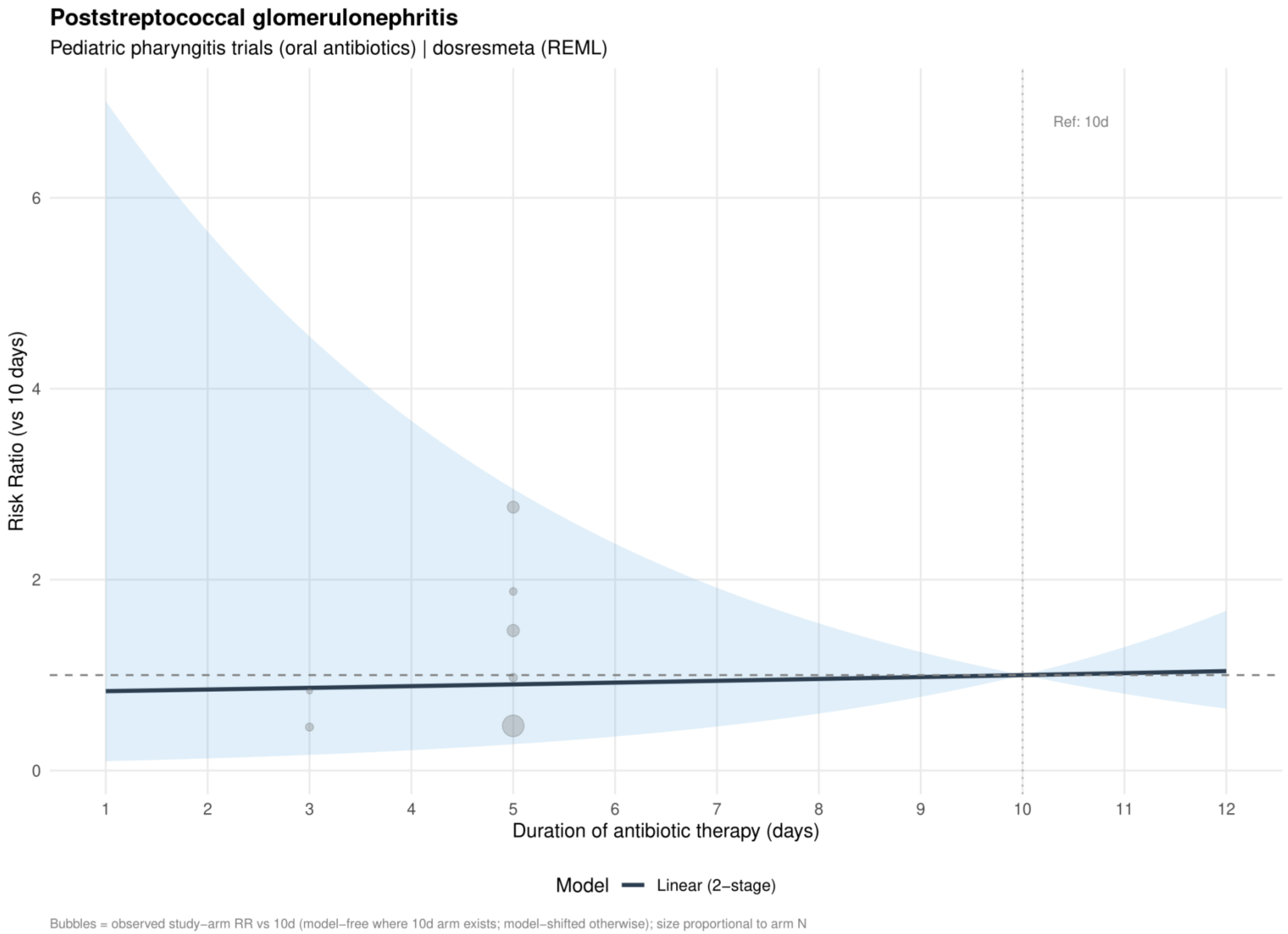


Curves show risk ratios relative to 10 days. The solid slate curve is the two stage linear model and, when present, the dashed red curve is the one stage restricted cubic spline with three knots; shaded bands show 95% confidence intervals. Bubbles show observed study arm effects and are scaled by arm sample size.

**Supplement 118. Poststreptococcal glomerulonephritis: linear estimates by risk of bias for the primary analysis**

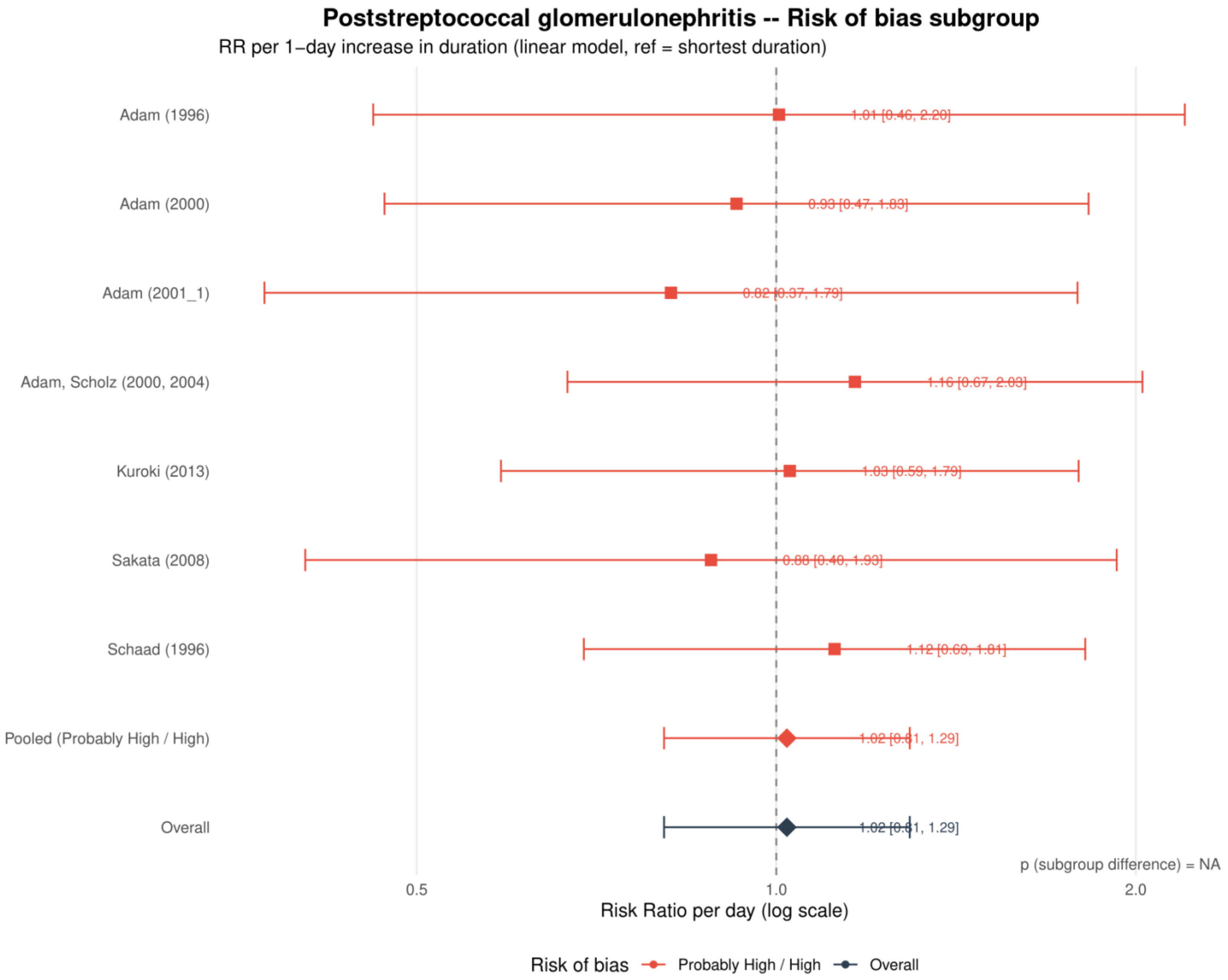


Study specific and pooled estimates are risk ratios per 1 day increase in antibiotic duration, with 95% confidence intervals.
*Only one collapsed risk of bias category contributed; the subgroup difference was not estimable.*

**Supplement 119. Poststreptococcal glomerulonephritis: dose-response analysis excluding azithromycin**

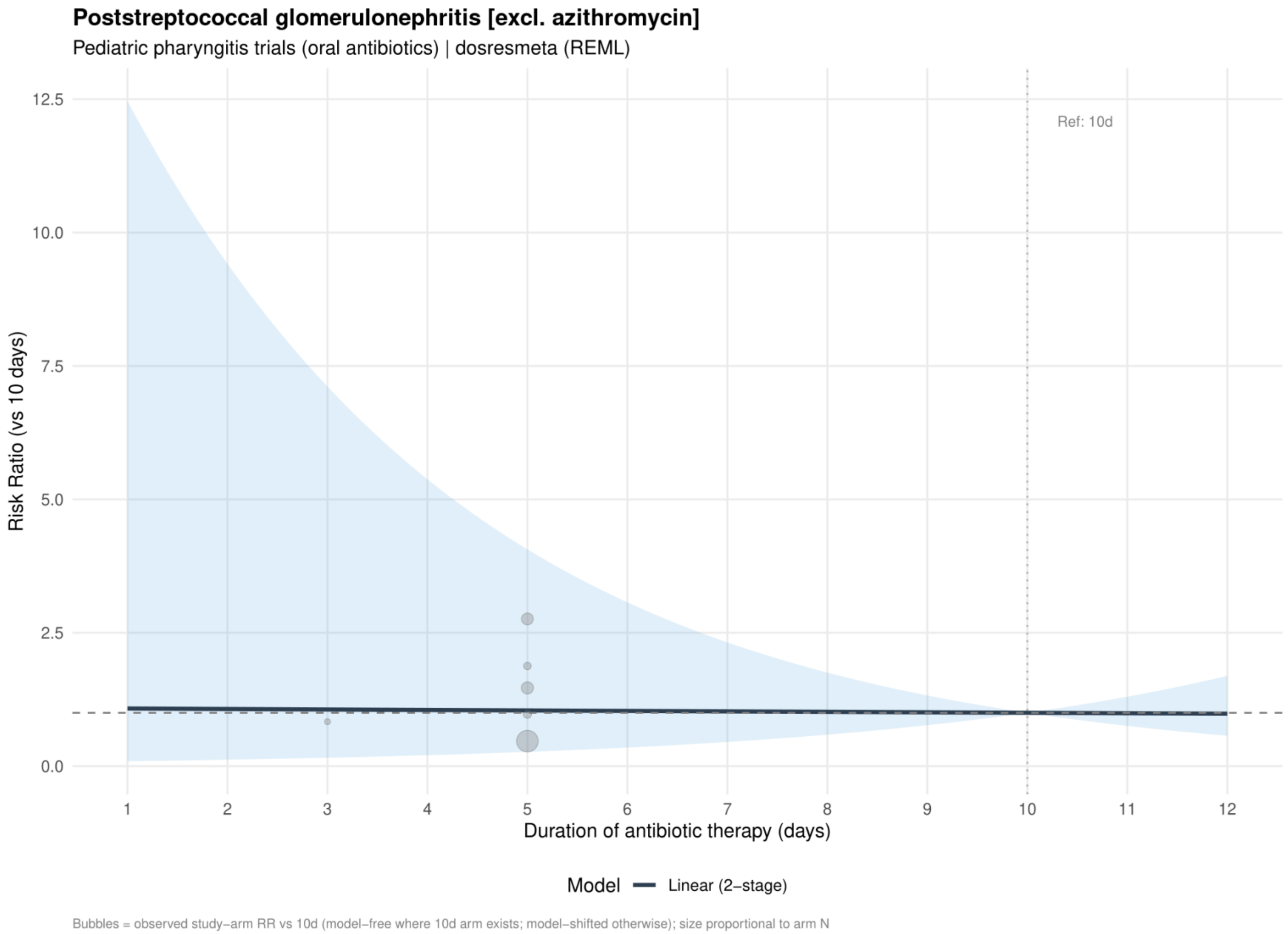


Curves show risk ratios relative to 10 days. The solid slate curve is the two stage linear model and, when present, the dashed red curve is the one stage restricted cubic spline with three knots; shaded bands show 95% confidence intervals. Bubbles show observed study arm effects and are scaled by arm sample size.

**Supplement 120. Poststreptococcal glomerulonephritis: linear estimates by risk of bias excluding azithromycin**

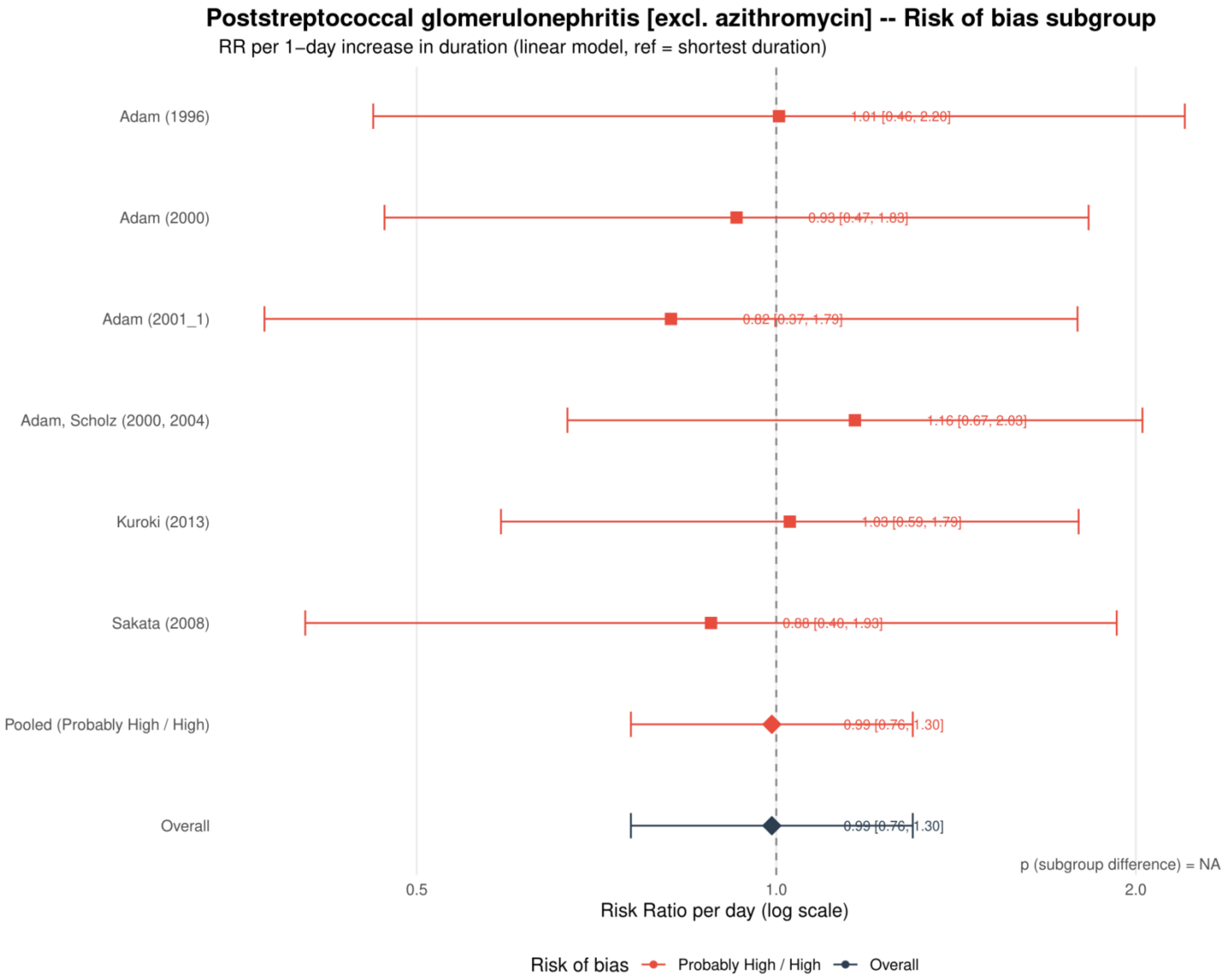


Study specific and pooled estimates are risk ratios per 1 day increase in antibiotic duration, with 95% confidence intervals.

*Only one collapsed risk of bias category contributed; the subgroup difference was not estimable.*

**Supplement 121. Poststreptococcal glomerulonephritis: dose-response analysis excluding macrolides**

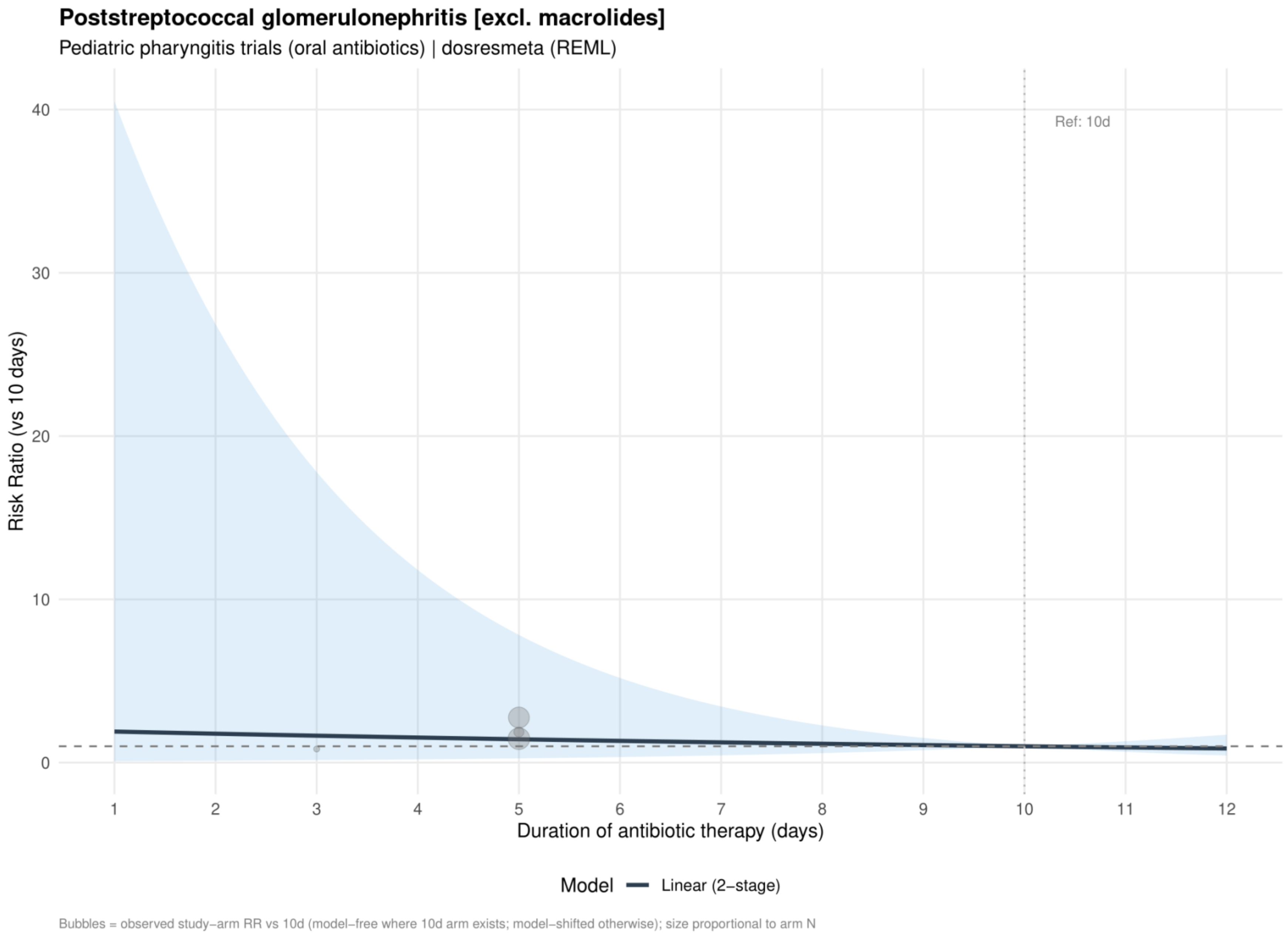


Curves show risk ratios relative to 10 days. The solid slate curve is the two stage linear model and, when present, the dashed red curve is the one stage restricted cubic spline with three knots; shaded bands show 95% confidence intervals. Bubbles show observed study arm effects and are scaled by arm sample size.

**Supplement 122. Poststreptococcal glomerulonephritis: linear estimates by risk of bias excluding macrolides**

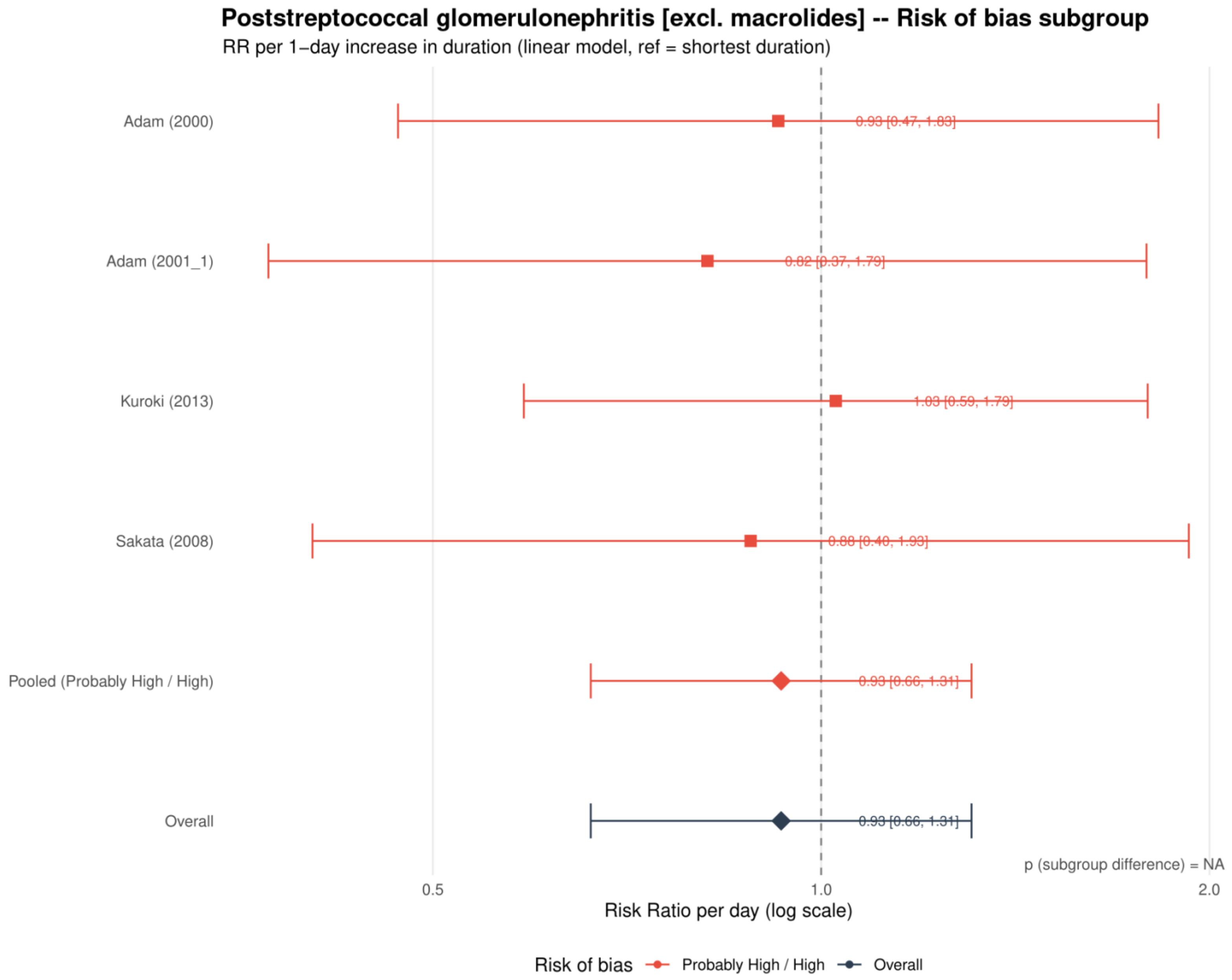


Study specific and pooled estimates are risk ratios per 1 day increase in antibiotic duration, with 95% confidence intervals.
*Only one collapsed risk of bias category contributed; the subgroup difference was not estimable.*

**Supplement 123. Poststreptococcal glomerulonephritis: dose-response analysis including placebo or no treatment as 0 days**

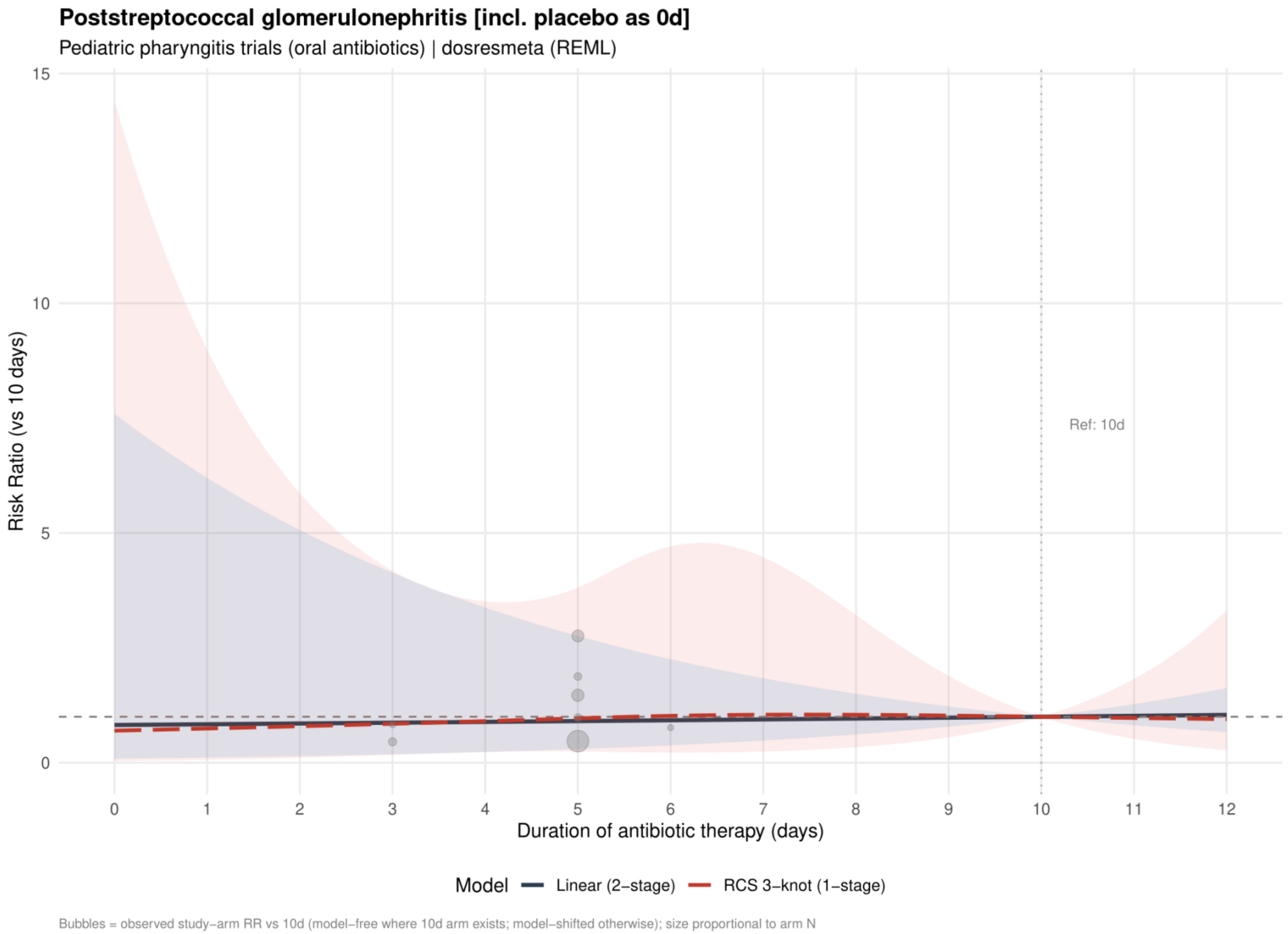


Curves show risk ratios relative to 10 days. The solid slate curve is the two stage linear model and, when present, the dashed red curve is the one stage restricted cubic spline with three knots; shaded bands show 95% confidence intervals. Bubbles show observed study arm effects and are scaled by arm sample size.

**Supplement 124. Poststreptococcal glomerulonephritis: linear estimates by risk of bias including placebo or no treatment as 0 days**

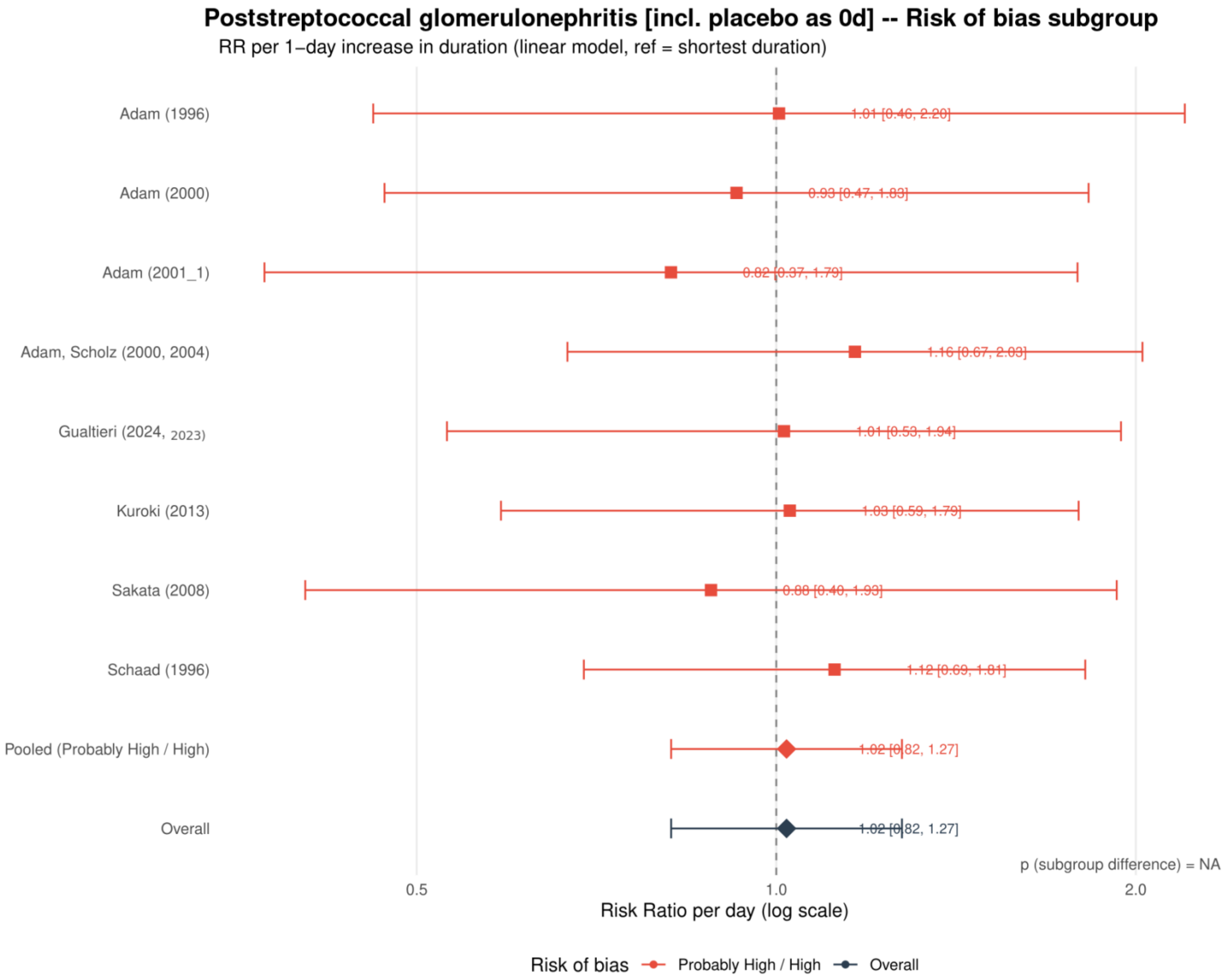


Study specific and pooled estimates are risk ratios per 1 day increase in antibiotic duration, with 95% confidence intervals.

*Only one collapsed risk of bias category contributed; the subgroup difference was not estimable.*

**Supplement 125. Poststreptococcal glomerulonephritis: restricted cubic spline estimates by risk of bias including placebo or no treatment as 0 days**

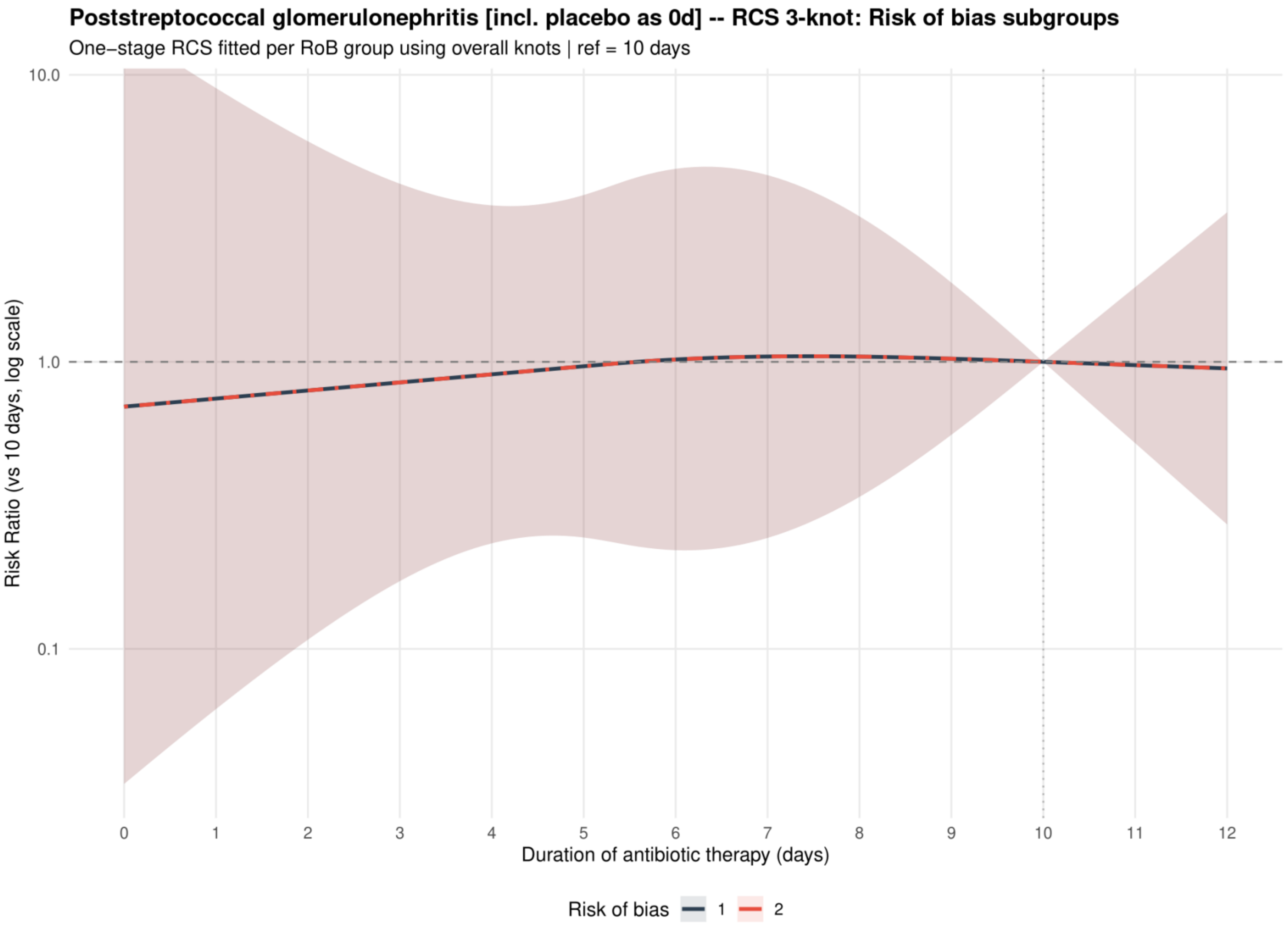


Curves show risk ratios relative to 10 days with 95% confidence intervals. Risk of bias codes: 1 = overall; 2 = low or probably low; 3 = probably high or high.
